# A time-sequenced approach to machine learning prognostic modelling with implementation on running-related injury prediction

**DOI:** 10.1101/2025.05.07.25327162

**Authors:** Han Wu, Katherine Brooke-Wavell, Michael R. Barnes, Zainab Awan, Sarabjit Mastana, Sam Allen, Richard C. Blagrove

**Author notes:** Correspondence to: Mr. Han Wu, National Centre for Sport and Exercise Medicine, School of Sport, Exercise and Health Sciences, Loughborough University, Loughborough, UK.

## Abstract

**Background:** The use of machine learning (ML) methods in medical prognostic modelling is gaining popularity, yet all currently available source models were designed from a general mathematics perspective. These models encounter limitations when embedding discipline-specific information, which restricts model practical interpretability. This study aimed to introduce two novel prognostic ML source models designed with an area-specific approach by testing their performance against commonly used ML methods, and explored their interpretability.

**Methods:** Measurements associated with multidisciplinary risk factors (genetics, history, neuromuscular capacity, biomechanics, body composition, nutrition, training) were collected on competitive endurance runners, who were subsequently monitored weekly over 12 months for running-related injuries (RRIs). Data was fitted with commonly used ML methods and the novel models using a stratified 10-fold cross validation framework for performance comparisons. Interpretable feature interactions were tested for statistical significance, and extracted feature importance scores were tested for correlation with Shapley Additive Explanation (SHAP) values.

**Results:** 6,181 valid weekly samples were collected from 142 competitive endurance runners. The novel methods’ performances (AUC 0·736-0·753, Accuracy 0·822-0·849, Sensitivity 0·376-0·455, Specificity 0·859-0·896) matched those of commonly used ML methods (AUC 0·649-0·784, Accuracy 0·662-0·857, Sensitivity 0·337-0·568, Specificity 0·671-0·904). Pairwise feature interactions revealed stable patterns (p<0·001). Method-specific computationally efficient feature importance scores moderately correlated with SHAP values (r=0·12-0·72), showing increases as model parameterization increased.

**Conclusion:** The novel methods showed comparable performance and better interpretability against common ML methods. Interpretability improved with increasing parameterization, suggesting performance may further improve with larger datasets and more features. Future research should perform higher quality validations using larger datasets before these methods can be widely adopted in prognostic modelling research.

**Funding:** This research is supported by the Alan Turing Institute Enrichment Scheme and the China Scholarship Council.

## 1. Introduction

Prognostic models can predict future health outcomes utilizing multiple predictor variables that guide medical decisions across various specialties.^1^ The recent advancements in machine learning (ML) offer promising opportunities to enhance the performance of prognostic models through flexible, non-linear, and complex model structures.^2^ Consequently, many studies have sought to leverage ML for creating specialized prognostic models across different medical disciplines.^2–5^

Despite their advantages, ML-based prognostic models have been criticized for their lack of interpretability, which becomes particularly problematic in high-stakes medical settings.^6^ Calls for explainable artificial intelligence (XAI) in medicine have consequently gained increasing support.^6^ To date, most attempts to apply ML to prognostic modelling have been based on general-purpose source models developed by mathematicians or computer scientists without the specific goals of prognostic modelling in mind, thus may not align with the rationale of medical decision-making. For instance, Bayesian Networks (BNs) can incorporate prior knowledge and preserve logic,^7^ yet they lack flexibility by assuming conditional independence (variables that share the same cause are treated as unrelated),^7,8^ which is often not suitable for prognostic risk factors.^9^

The cost of sport injuries represents a substantial financial burden to medical services and sport organizations.^10^ Sports injury prediction is a specialized area within prognostic modelling and has also drawn research interests utilizing ML.^11–13^ Proposals have been made to establish domain-specific algorithms to emulate complex interactions between injury risk factors in specific sports,^14^ but no models have been developed and validated on real-world datasets. Endurance running is one of the most popular sports worldwide yet carries a significant risk of injury,^15^ warranting research to develop targeted prognostic models.

This study aimed to establish two ML prognostic source models by embedding area specific assumptions (i.e., a time-sequenced approach) at the algorithmic level to improve interpretability. A time-sequenced approach assumes that an individual is born with a predisposed risk for a certain disease or injury, which is modified through time as life events unfold.^16^ These models were evaluated against existing ML models using multidisciplinary data from competitive endurance runners for their ability to predict running-related injuries (RRIs) on a weekly basis. Interpretable information from these models was compared to Shapley Additive Explanations (SHAP)^17^ to highlight their unique aspects. It was hypothesized that the novel models match the AUC, accuracy, specificity, and sensitivity of state-of-the-art ML models, can provide feature influence metrics that correlate with SHAP values (Pearson’s r >0·7) while requiring less computation, and can reveal pair-wise feature interactions that do not occur by chance (p<0·05).

## 2. Methods

### 2.1. Ethics and Participant Recruitment

This study used a prospective design examining risk factors associated with endurance running-related injuries over a 1-year period in 149 participants. The study received approval from the National Health Service (NHS) Research Ethics Committee (South West - Central Bristol) and the Loughborough University Ethics Sub-Committee. Participants were recruited between November 2022 and July 2023. Inclusion criteria were: (i) age 14-50 years, (ii) participate in competitive endurance (≥ 5 km) running for at least three years, (iii) minimum four hours running per week, and (iv) absence of injuries at initiation. Exclusion criteria were: (i) use of medications or presence of medical conditions significantly affecting bone health, (ii) pins or plates in limbs subjected to bone scans, (iii) pregnant or breastfeeding within the previous six months, and (iv) current use of vaping or smoking. Recruitment was conducted via social media and direct contact with running clubs and coaches across the UK. The study was explained to each participant, and signed informed consent was obtained. For participants under 18 years old, assent from the participant and consent from their parent/carer were obtained.

Competitive endurance runners are a population for which sample sizes are inherently limited. The current study’s sample size reflects the availability of participants meeting inclusion criteria, which is consistent with previous large-scale prospective studies.^18^

### 2.2. Baseline Measurements

Participants completed an online questionnaire (Onlinesurveys, Jisc, Bristol, UK) encompassing 12-month injury history, the Bone-specific Physical Activity Questionnaire (BPAQ),^19^ six-month performance records, 12-month training history, the Eating Disorder Examination Questionnaire (EDE-Q),^20^ and the Low Energy Availability in Females Questionnaire (LEAF-Q).^21^

On arrival at the laboratory, participants had their height and body mass taken (Seca 274 stadiometer, Birmingham, UK). Dual-energy X-ray Absorptiometry (DXA) scan (GE Lunar iDXA, GE HealthCare Technologies, Chicago, US) for the whole body, lumbar spine, hips, and non-dominant forearm were used to measure bone mineral density and body composition. A Peripheral Quantitative Computed Tomography (pQCT) scan (XCT 2000L, Novotec Medical GmbH and Stratec Medizintechnik GmbH, Pforzheim, Germany) was also conducted for the non-dominant tibia at 4%, 38%, and 66% of tibial lengths to measure bone and tissue structures. Q angle was measured using a goniometer, and sit-to-stand navicular drop was assessed using an established protocol.^22^ Running ground reaction forces and stride frequency were recorded on an instrumented treadmill (Treadmetrix, Utah, US) at two speeds (10 km/h and 12 km/h), for one minute at 1000 Hz sampling frequency. Hip abduction/adduction and knee flexion/extension torque were measured using an isokinetic dynamometer (Isomed 2000, D&R Ferstl GmbH, Hemau, Germany) at 60 degrees/sec, over five sets of four repetitions as indicators of muscular strength.

### 2.3. Prospective Tracking Period

Following baseline testing, participants completed a weekly online questionnaire (Qualtrics XM, Seattle, US) for 52 weeks, reporting running volume, other physical training, and injury information using the Oslo Sports Trauma Research Centre Overuse Injury Questionnaire.^23^ Every four months, participants revisited the laboratory for DXA and pQCT scans, replicating baseline procedures. Between visits, participants maintained a three-day food diary using the Libro App (Nutritics, Dublin, Ireland) with standardized digital food weighing scales (Duratool, Farnell; Leeds, UK) provided.

Participants provided 2 ml of saliva (Isohelix GeneFix™ Saliva-Prep 2 DNA Kit, Cell Projects, Harrietsham, UK) during a visit for DNA extraction, which was subsequently analyzed by CD Genomics (New York, US) for selected single-nucleotide polymorphisms (SNPs) previously correlated with RRIs.

Supplementary Material (SM) S1-3 contains detailed information regarding experimental procedures and rationale for risk factor inclusions.

### 2.4. Data Preprocessing

Each participant-week constituted a sample, with the objective of utilizing all available information preceding each week to predict injury occurrence within a given week. Injury was defined as an increase in Oslo score for any bodily region compared to the prior week plus the participant’s subjective identification of the injury being running-related.^23^ Samples lacking prior week data were excluded and identified injury cases were individually inspected to reduce errors.

Features were potential risk factors categorized into three classes based on the quality of associated evidence (SM S3). Analyses were performed separately for Class 1 (highest evidence level; n=39) and all features (Class 1-3; n=257). Data imputation was conducted using 0, median, or mean depending on feature properties, and all data was normalized to 0-1 for ML training (SM S4).

### 2.5. Model Design

Features were divided into four categories:

1. **Genetics**: Genotypic information.
2. **History**: Experiences from birth to baseline testing.
3. **Phenotype**: Current physical status.
4. **Behavior**: Actions during prospective tracking (considered as ‘triggering events’).

Information flows through these categories sequentially, reflecting temporal risk modification.

#### 2.5.1. Time-Sequenced Neural Network (TSNN)

TSNN resembles a conventional neural network (NN) but integrates history, phenotype, and behavior features sequentially into the hidden layers, granting information to naïve nodes. Each hidden layer includes an additional non-feature-attached node to account for unmeasured information.

A NN node calculation before activation is:

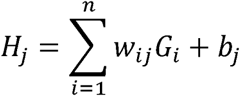

Where *H_j_* is the output value at the *j*-th second layer (history) node, *w_ij_* is the weight from the *i*-th input (genetic) feature, and *b_j_* is the bias term.

In contrast, a TSNN node calculation before activation is:

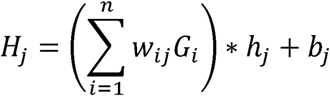

Where *h_j_* represents the history feature values at the *j*-th history node.

This formulation allows the node to reflect the extent a historical risk factor is influenced by genetic predispositions. A ReLU activation function ensures that historical risk factors below a triggering threshold remain inactive, thereby not impacting subsequent phenotype nodes. Fig. 1 illustrates the basic structure of TSNN.

**Figure 1:**
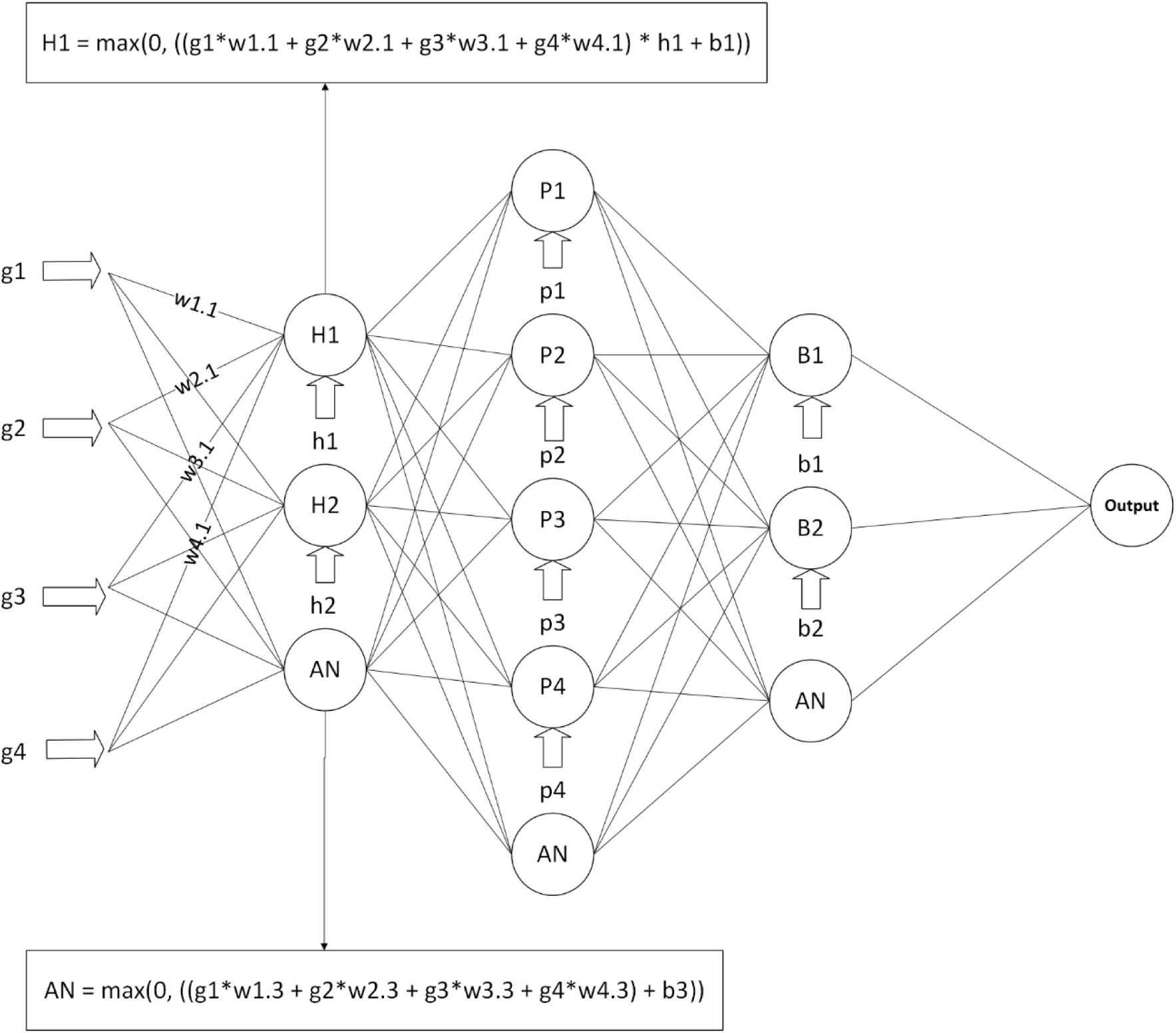
Illustration of TSNN’s basic structure. g: genotype input features; h: history input features; p: phenotype input features; b: behavior input features; H: history nodes; P: phenotype nodes; B: behavior nodes; w: weights; b: bias terms; AN: additional node.

#### 2.5.2. Time-Sequenced Graph Neural Network (TSGNN)

TSGNN resembles a one-layer Graph Neural Network but differs in that self-feature values are timed rather than added. TSGNN’s edge assignment logic is:

- **Genetic Nodes**: Send edges to all other nodes.
- **History Nodes**: Send edges to all other non-genetic nodes.
- **Phenotype Nodes**: Send edges to all other non-genetic/history nodes.
- **Behavior Nodes**: Send edges to all other behavior nodes.

After exchanging information among nodes, each node directs an edge toward the output. Fig. 2 depicts the basic structure of TSGNN.

**Figure 2:**
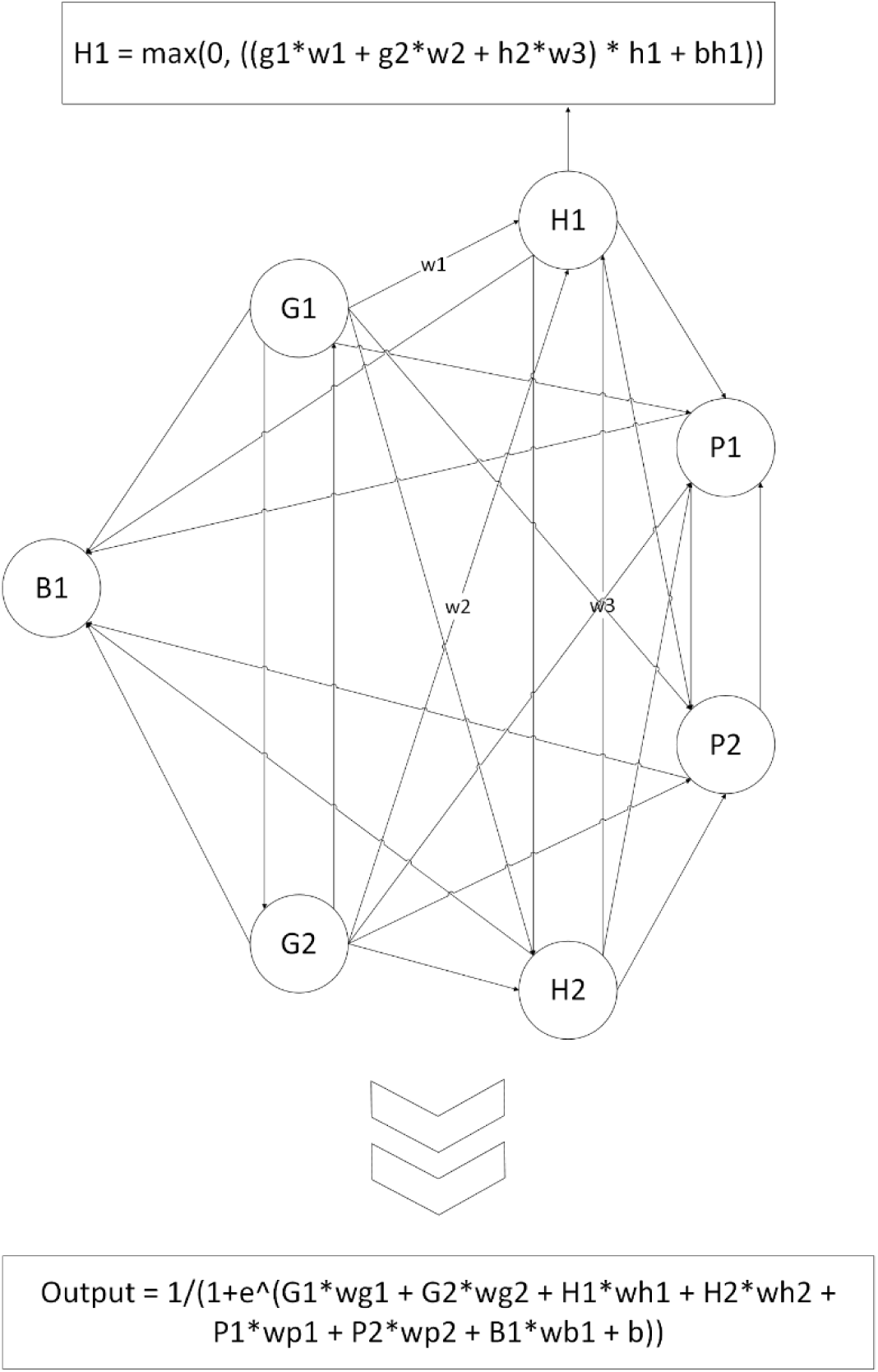
Illustration of TSGNN’s basic structure. G: genotype nodes; H: history nodes; P: phenotype nodes; B: behavior node; g: genotype feature values; h1: history feature values at H1; w: weights; b: bias terms. G1 sends to every node except itself; receives only from G2. H1 sends to H2, P1, P2, B1; receives from G1, G2, H2. P1 sends to P2, B1; receives from G1, G2, H1, H2, P2. B1 receives from every other node (only 1 behavior node for simplicity). After information transfer, output is calculated using updated node values.

TSGNN tackles two of TSNN’s inherent weaknesses. Firstly, TSNN operates under the assumption that features within the same timeframe interact additively, while interactions across different timeframes are multiplicative; TSGNN breaks this assumption. Secondly, influences from proximal features in TSNN might diminish during propagation;^24^ TSGNN adopts a shallower structure to ensure each feature can directly impact the outcome.

### 2.6. Model Selection

Decision tree, BN, and Logistic regression were selected due to their simple transparent structures, which provide high interpretability. Among more complex algorithms, Tree-based ensemble methods (Random Forest, Adaboost, Gradient Boosting) were selected for their ability to handle class imbalance through techniques like weighted splitting and iterative error correction. K Nearest Neighbor (KNN) was selected for similar-case matching. Support Vector Machine (SVM) was selected for its effectiveness in high-dimensional spaces. Multi-layer Perceptron (MLP) risks overfitting for small datasets but was selected as benchmark because of its structural similarity to the novel models (alpha value of L2 regularization was included in hyperparameter tuning to reduce risks of overfitting). Naïve Bayes was selected due to its probabilistic predictions which align with clinical risk communications. These models are common in previous prognostic ML research.^25,26^

### 2.7. Model Evaluation

Area Under the Curve (AUC) for the Receiver Operating Characteristic within a stratified 10-fold cross-validation framework was employed as the evaluation metric to prevent overfitting and for comparison against previous ML RRI studies.^12,13,27^ Due to high candidate feature-to-sample ratio, Relief and logistic Least Absolute Shrinkage and Selection Operator (LASSO) were employed to reduce dimensionality.

- **Relief**: Features were ranked and tested with scikit-learn default hyperparameters iteratively (test top feature, then top 1+2, etc.). The subset with the highest average AUC was selected.
- **LASSO**: Employed 40 equally spaced alpha values (0·1-0·00001) to rank features separately, followed by iterative testing. If the best-performing subsets from Relief and LASSO were close (AUC difference < 0·01), both were subjected to hyperparameter tuning; otherwise, only the superior subset was tuned.

Due to the small sample size, which reduces computation load, grid search was employed for all commonly used hyperparameters (SM S7) to enhance robustness. Oversampling strategies and sampling rates were concurrently tuned to address class imbalance. If optimal hyperparameters reached the boundaries of the specified ranges during the initial run, the ranges were expanded, and a narrowed search was conducted until all hyperparameters resided within boundaries.

TSNN and TSGNN features in different categories were treated differently, rendering global ranking and iterative testing inappropriate. Consequently, the number of features included in each category were treated as separate hyperparameters, necessitating concurrent feature selection and hyperparameter tuning. Given the expanded feature space, targeted tuning was performed using Optuna (200 combinations for Class 1 and 500 combinations for all features per ranking method). Hyperparameter ranges were based on the best hyperparameters for MLP. During implementation, no hyperparameters reached the specified boundaries, consequently no re-runs were conducted. Feature ranking was performed using either Relief or LASSO (fixed alpha=0·04), applied in two ways each (totaling four ranking methods): (1) ranking all features first, then separating by category, or (2) separating features by category first, then ranking within category.

Following hyperparameter tuning, models were retrained using the optimal hyperparameters to determine a threshold that maximizes the f1 score. Accuracy, sensitivity, and specificity were reported under this threshold.

### 2.8. Model Interpretation

Due to the inherent stochasticity of neural networks,^28^ different initializations led to convergence at distinct local minima, resulting in variations in learned parameters. To extract consistent trends, each model was trained 20 times with random initializations.

#### 2.8.1. Feature Interactions

TSNN feature pairs exhibit clear, mathematically defined unidirectional interactions due to the model’s inherent structure: consistently positive or negative weights can be directly interpreted as synergistic or antagonistic relationships, respectively. Each TSGNN feature-attached node transmits information to other nodes before receiving input from other features. Consequently, consistent weights extracted from TSGNN reflect partial synergistic or antagonistic relationships. Probabilities for numbers of consistent weights were estimated using binomial distributions, distinguishing systematic patterns from random fluctuations.

#### 2.8.2. Feature Contributions

The structures of TSNN and TSGNN enable the estimation of each feature’s contribution to a given sample by decomposing the model into contribution pathways and summing the routes that pass through the corresponding feature node. This approach requires significantly less computation than SHAP.

Feature contribution values were averaged across 20 independent runs and compared to similarly averaged SHAP values. Pearson’s correlation and Spearman’s rank correlation coefficients were calculated for 100 randomly selected samples. These analyses were performed for all features and for the five largest positive and negative features based on feature contributions (SM S9).

## 3. Results

A total of 142 participants contributed 6,181 valid samples, of which 564 included identifications of injuries. Fig. 3 illustrates participant recruitment and adherence.

**Figure 3:**
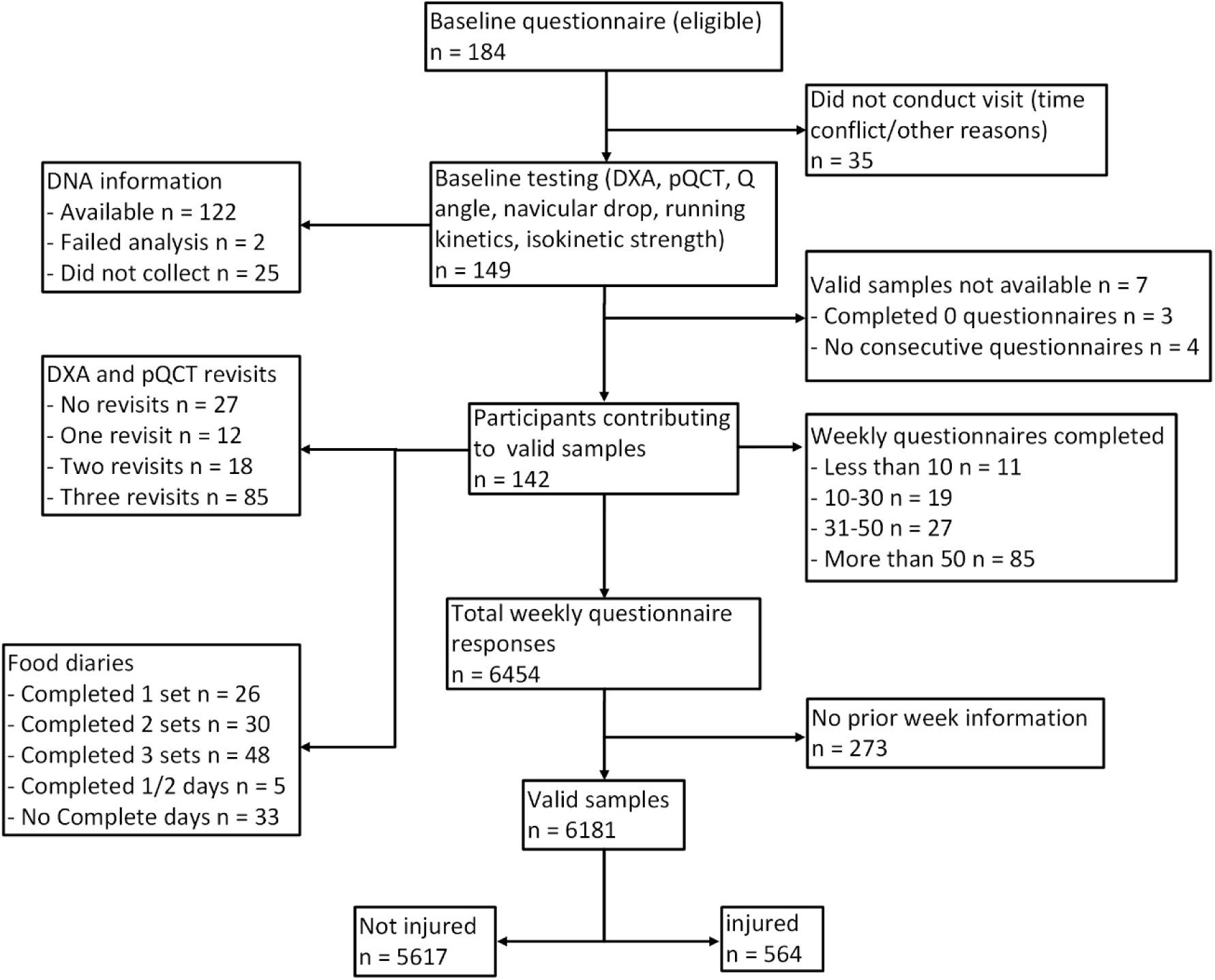
Flowchart illustrating participant recruitment, drop-out, and adherence during the study.

### 3.1. Cohort Descriptives

Among the 142 participants, 78 were male and 64 female. The World Athletics points score^29^ derived from six-month performance records averaged 811 ± 181 for females and 584 ± 245 for males. Participants reported an average of 47·7 ± 91·4 days affected by lower limb injuries in the 12 months prior to the study. Thirty-four participants were uninjured in the 12 months preceding the study. Table 1 presents basic descriptive statistics (more see SM S4.1).

**Table 1:** Descriptive statistics at baseline.

| | Male (±SD) | Female (±SD) | Total (±SD) |
| --- | --- | --- | --- |
| Participant count | 78 | 64 | 142 |
| Age (years) | 32.0 ± 9.7 | 30.9 ± 9.9 | 31.5 ± 9.8 |
| Body mass (kg) | 72.0 ± 8.6 | 55.8 ± 6.2 | 64.7 ± 11.1 |
| Performance score<sup>29</sup> | 584 ± 245 | 811 ± 181 | 686 ± 245 |
| Past year running frequency<br>(runs per week) | 6.24 ± 2.00 | 5.42 ± 1.39 | 5.88 ± 1.80 |
| BMI (kg.m<sup>-2</sup>) | 22.3 ± 2.2 | 20.4 ± 1.9 | 21.4 ± 2.2 |
| Past year injury days | 42.4 ± 91.4 | 54.1 ± 91.7 | 47.7 ± 91.4 |
| Participants injured (uninjured)<br>past year | 55 (23) | 53 (11) | 108 (34) |

### 3.2. Model Performance

Table 2 displays model performance. Most models achieved average AUCs around 0·75, with small or no improvement when utilizing all features compared to only Class 1. Naïve Bayes and BN underperformed, with AUCs below 0·7 for both feature sets. Tree-based ensemble models (Random Forest, Gradient Boosting, AdaBoost) demonstrated superior overall performance across both feature pools. TSNN and TSGNN exhibited slightly lower performance with Class 1 features (0·736 ± 0·040 and 0·746 ± 0·031) compared to most models, but reached near-average performance when all features were used (0·753 ± 0·033 and 0·753 ± 0·022). Multiple logistic regression performed worse than most ML models with Class 1 features (0·674 ± 0·047) but showed significant improvement with the full feature set (0·762 ± 0·027).

**Table 2:**
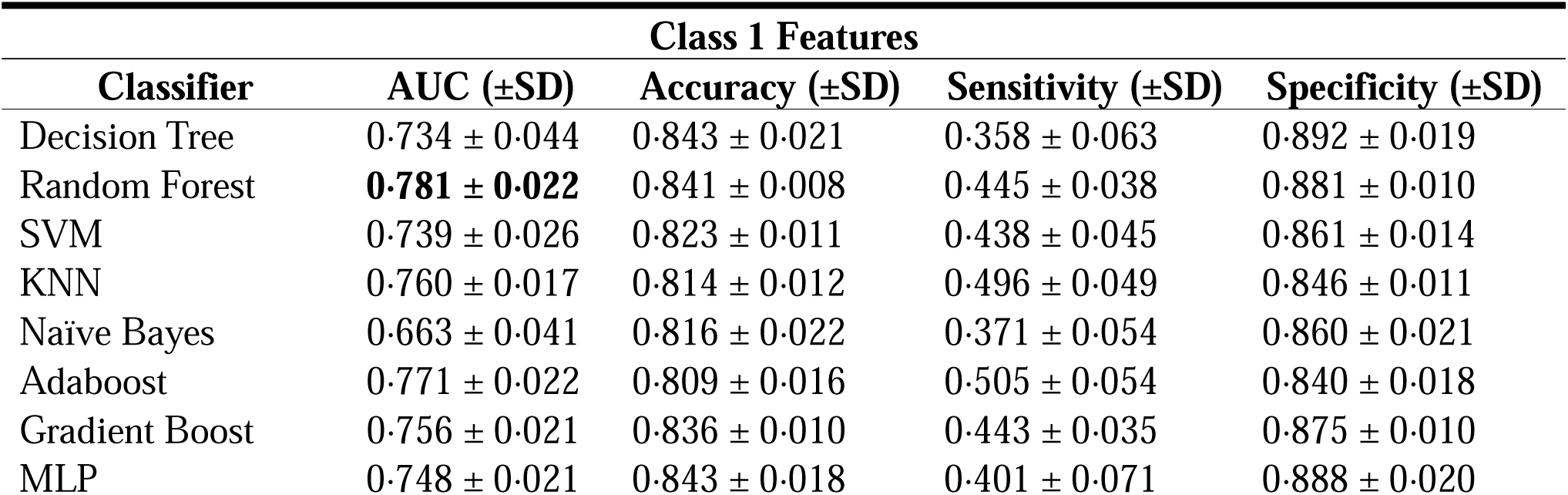
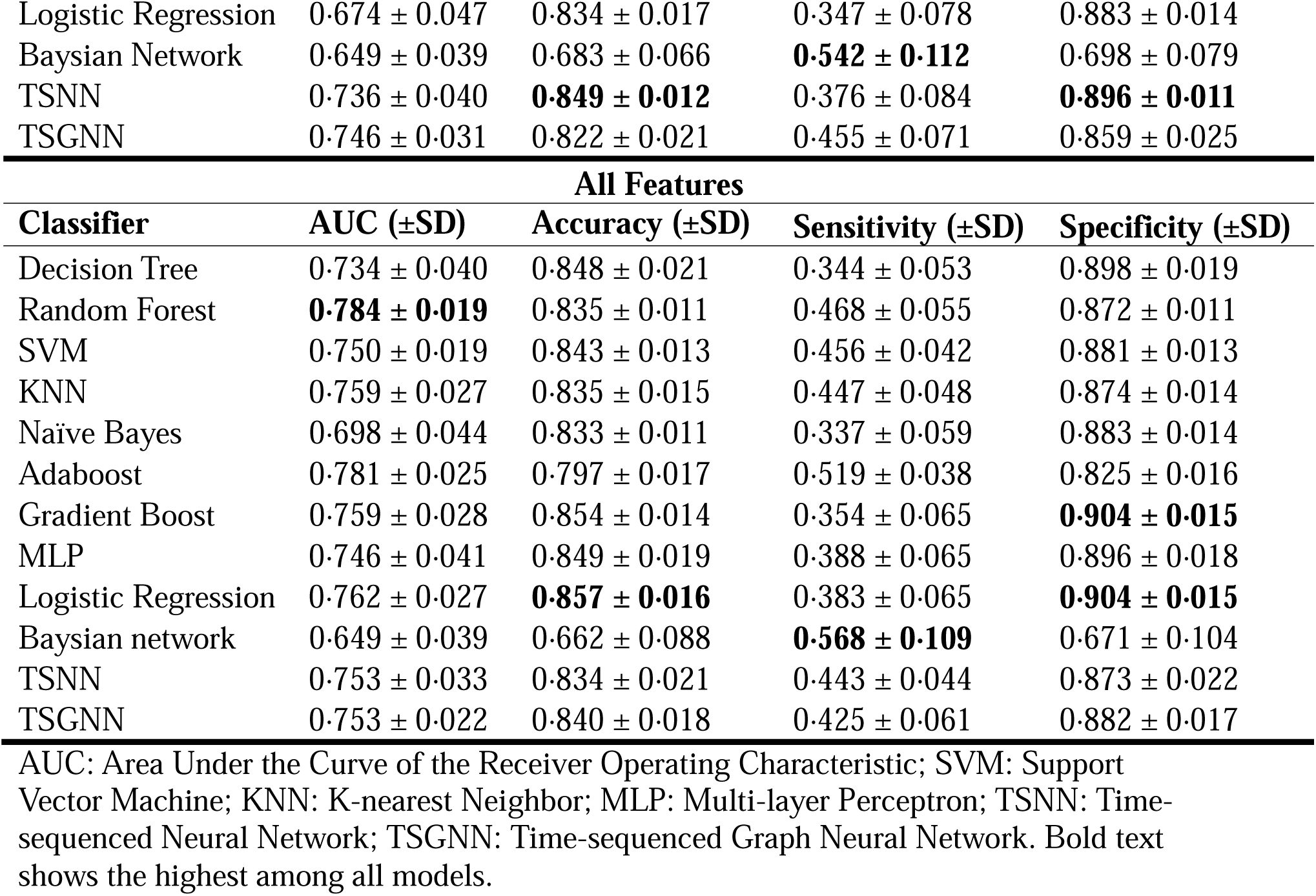
Model performance when using class 1 features or all features as the feature pool.

| Classifier | Class 1 Features |  |  |  |
| --- | --- | --- | --- | --- |
| | AUC (±SD) | Accuracy (±SD) | Sensitivity (±SD) | Specificity (±SD) |
| Decision Tree | 0.734 ± 0.044 | 0.843 ± 0.021 | 0.358 ± 0.063 | 0.892 ± 0.019 |
| Random Forest | **0.781 ± 0.022** | 0.841 ± 0.008 | 0.445 ± 0.038 | 0.881 ± 0.010 |
| SVM | 0.739 ± 0.026 | 0.823 ± 0.011 | 0.438 ± 0.045 | 0.861 ± 0.014 |
| KNN | 0.760 ± 0.017 | 0.814 ± 0.012 | 0.496 ± 0.049 | 0.846 ± 0.011 |
| Naïve Bayes | 0.663 ± 0.041 | 0.816 ± 0.022 | 0.371 ± 0.054 | 0.860 ± 0.021 |
| Adaboost | 0.771 ± 0.022 | 0.809 ± 0.016 | 0.505 ± 0.054 | 0.840 ± 0.018 |
| Gradient Boost | 0.756 ± 0.021 | 0.836 ± 0.010 | 0.443 ± 0.035 | 0.875 ± 0.010 |
| MLP | 0.748 ± 0.021 | 0.843 ± 0.018 | 0.401 ± 0.071 | 0.888 ± 0.020 |

| Logistic Regression | 0.674 ± 0.047 | 0.834 ± 0.017 | 0.347 ± 0.078 | 0.883 ± 0.014 |
| --- | --- | --- | --- | --- |
| Baysian Network | 0.649 ± 0.039 | 0.683 ± 0.066 | **0.542 ± 0.112** | 0.698 ± 0.079 |
| TSNN | 0.736 ± 0.040 | **0.849 ± 0.012** | 0.376 ± 0.084 | **0.896 ± 0.011** |
| TSGNN | 0.746 ± 0.031 | 0.822 ± 0.021 | 0.455 ± 0.071 | 0.859 ± 0.025 |
| All Features |  |  |  |  |
| Classifier | AUC (±SD) | Accuracy (±SD) | Sensitivity (±SD) | Specificity (±SD) |
| Decision Tree | 0.734 ± 0.040 | 0.848 ± 0.021 | 0.344 ± 0.053 | 0.898 ± 0.019 |
| Random Forest | **0.784 ± 0.019** | 0.835 ± 0.011 | 0.468 ± 0.055 | 0.872 ± 0.011 |
| SVM | 0.750 ± 0.019 | 0.843 ± 0.013 | 0.456 ± 0.042 | 0.881 ± 0.013 |
| KNN | 0.759 ± 0.027 | 0.835 ± 0.015 | 0.447 ± 0.048 | 0.874 ± 0.014 |
| Naïve Bayes | 0.698 ± 0.044 | 0.833 ± 0.011 | 0.337 ± 0.059 | 0.883 ± 0.014 |
| Adaboost | 0.781 ± 0.025 | 0.797 ± 0.017 | 0.519 ± 0.038 | 0.825 ± 0.016 |
| Gradient Boost | 0.759 ± 0.028 | 0.854 ± 0.014 | 0.354 ± 0.065 | **0.904 ± 0.015** |
| MLP | 0.746 ± 0.041 | 0.849 ± 0.019 | 0.388 ± 0.065 | 0.896 ± 0.018 |
| Logistic Regression | 0.762 ± 0.027 | **0.857 ± 0.016** | 0.383 ± 0.065 | **0.904 ± 0.015** |
| Baysian network | 0.649 ± 0.039 | 0.662 ± 0.088 | **0.568 ± 0.109** | 0.671 ± 0.104 |
| TSNN | 0.753 ± 0.033 | 0.834 ± 0.021 | 0.443 ± 0.044 | 0.873 ± 0.022 |
| TSGNN | 0.753 ± 0.022 | 0.840 ± 0.018 | 0.425 ± 0.061 | 0.882 ± 0.017 |

AUC: Area Under the Curve of the Receiver Operating Characteristic; SVM: Support Vector Machine; KNN: K-nearest Neighbor; MLP: Multi-layer Perceptron; TSNN: Time-sequenced Neural Network; TSGNN: Time-sequenced Graph Neural Network. Bold text shows the highest among all models.

| Class 1 Features |  |  |  |  |
| --- | --- | --- | --- | --- |
| Classifier | AUC (±SD) | Accuracy (±SD) | Sensitivity (±SD) | Specificity (±SD) |
| Decision Tree | 0.734 ± 0.044 | 0.843 ± 0.021 | 0.358 ± 0.063 | 0.892 ± 0.019 |
| Random Forest | **0.781 ± 0.022** | 0.841 ± 0.008 | 0.445 ± 0.038 | 0.881 ± 0.010 |
| SVM | 0.739 ± 0.026 | 0.823 ± 0.011 | 0.438 ± 0.045 | 0.861 ± 0.014 |
| KNN | 0.760 ± 0.017 | 0.814 ± 0.012 | 0.496 ± 0.049 | 0.846 ± 0.011 |
| Naïve Bayes | 0.663 ± 0.041 | 0.816 ± 0.022 | 0.371 ± 0.054 | 0.860 ± 0.021 |
| Adaboost | 0.771 ± 0.022 | 0.809 ± 0.016 | 0.505 ± 0.054 | 0.840 ± 0.018 |
| Gradient Boost | 0.756 ± 0.021 | 0.836 ± 0.010 | 0.443 ± 0.035 | 0.875 ± 0.010 |
| MLP | 0.748 ± 0.021 | 0.843 ± 0.018 | 0.401 ± 0.071 | 0.888 ± 0.020 |
| Logistic Regression | 0.674 ± 0.047 | 0.834 ± 0.017 | 0.347 ± 0.078 | 0.883 ± 0.014 |
| Baysian Network | 0.649 ± 0.039 | 0.683 ± 0.066 | **0.542 ± 0.112** | 0.698 ± 0.079 |
| TSNN | 0.736 ± 0.040 | **0.849 ± 0.012** | 0.376 ± 0.084 | **0.896 ± 0.011** |
| TSGNN | 0.746 ± 0.031 | 0.822 ± 0.021 | 0.455 ± 0.071 | 0.859 ± 0.025 |
| All Features |  |  |  |  |
| Classifier | AUC (±SD) | Accuracy (±SD) | Sensitivity (±SD) | Specificity (±SD) |
| Decision Tree | 0.734 ± 0.040 | 0.848 ± 0.021 | 0.344 ± 0.053 | 0.898 ± 0.019 |
| Random Forest | **0.784 ± 0.019** | 0.835 ± 0.011 | 0.468 ± 0.055 | 0.872 ± 0.011 |
| SVM | 0.750 ± 0.019 | 0.843 ± 0.013 | 0.456 ± 0.042 | 0.881 ± 0.013 |
| KNN | 0.759 ± 0.027 | 0.835 ± 0.015 | 0.447 ± 0.048 | 0.874 ± 0.014 |
| Naïve Bayes | 0.698 ± 0.044 | 0.833 ± 0.011 | 0.337 ± 0.059 | 0.883 ± 0.014 |
| Adaboost | 0.781 ± 0.025 | 0.797 ± 0.017 | 0.519 ± 0.038 | 0.825 ± 0.016 |
| Gradient Boost | 0.759 ± 0.028 | 0.854 ± 0.014 | 0.354 ± 0.065 | **0.904 ± 0.015** |
| MLP | 0.746 ± 0.041 | 0.849 ± 0.019 | 0.388 ± 0.065 | 0.896 ± 0.018 |
| Logistic Regression | 0.762 ± 0.027 | **0.857 ± 0.016** | 0.383 ± 0.065 | **0.904 ± 0.015** |
| Baysian network | 0.649 ± 0.039 | 0.662 ± 0.088 | **0.568 ± 0.109** | 0.671 ± 0.104 |
| TSNN | 0.753 ± 0.033 | 0.834 ± 0.021 | 0.443 ± 0.044 | 0.873 ± 0.022 |
| TSGNN | 0.753 ± 0.022 | 0.840 ± 0.018 | 0.425 ± 0.061 | 0.882 ± 0.017 |

AUC: Area Under the Curve of the Receiver Operating Characteristic; SVM: Support Vector Machine; KNN: K-nearest Neighbor; MLP: Multi-layer Perceptron; TSNN: Time-sequenced Neural Network; TSGNN: Time-sequenced Graph Neural Network. Bold text shows the highest among all models.

### 3.3. Model Interpretation

Table 3 presents the number of consistent weights in TSNN and associated probabilities. While no weight remained consistent across all 20 runs, several weights were stable in 15, 16, or 17 out of 20 runs when models were trained with the full feature set with significantly low probabilities. With TSGNN, 9 out of 633 weights (p<0·001) and 164 out of 27,225 weights (p<0·001) were consistent throughout 20 runs for models trained with class 1 and with all features, respectively.

**Table 3:**
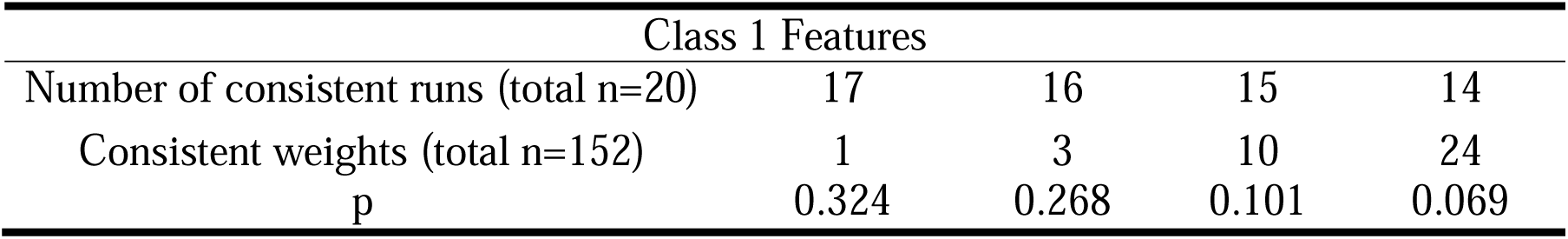
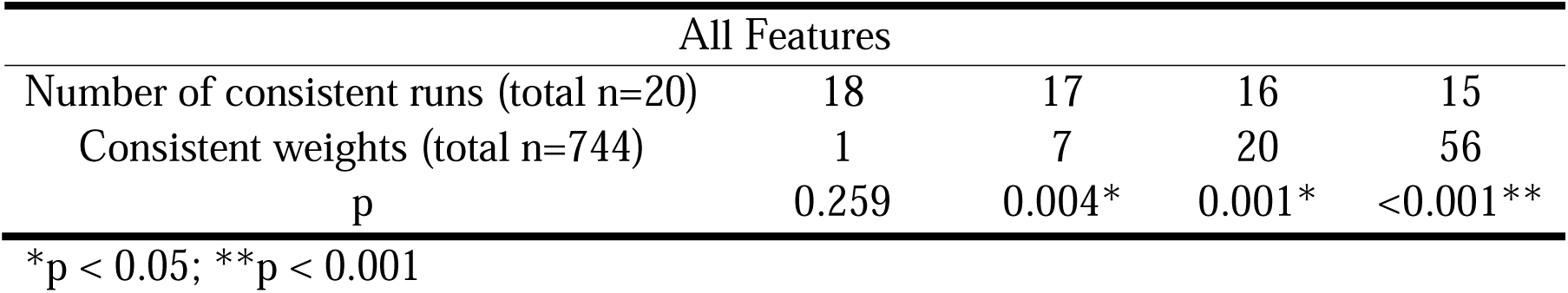
Number of consistent weight values for TSNN.

SM S11.1 shows correlation metrics between calculated feature contribution values and SHAP values. Correlations were higher and had smaller variances when using the entire feature pool compared to only using Class 1. Correlations apart from TSNN Class 1 were mostly moderate, with high correlations achieved from TSGNN all features pool when taking the top 5 positive and negative features (r=0·72 ± 0·26, ρ=0·71 ± 0·27).

## 4. Discussion

This study introduces two prognostic ML algorithms, TSNN and TSGNN, which utilize a time-sequenced approach to integrate area-specific information. These models achieved slightly inferior performance with a smaller feature pool and comparable performance with a larger feature pool relative to other ML models, demonstrating their capacity to capture complex and nonlinear interrelationships among features. Both models exhibited a degree of interpretability, the effectiveness of which increased as model parameterization increased, suggesting greater validity if trained with larger datasets. Future studies should implement TSNN/TSGNN with larger datasets to further assess the models’ efficacy.

The ML models, apart from Bayesian models, showed moderate levels of performance (AUC=0·734-0·784), surpassing previous studies on RRI prediction (AUC=0·58-0·74) despite smaller sample sizes.^12,13,27^ This is likely due to the comprehensive multidisciplinary feature pool which captures more task-relevant information. Additionally, this study is the first to integrate genetic information with physiological and biomechanical data for RRI prediction, bridging a critical gap across disciplines.

TSNN and TSGNN performed comparably to more flexible ML models (tree-based models, MLP, SVM, KNN) rather than more stable models (logistic regression, BN), indicating their potential as surrogates for flexible models with enhanced interpretability when simpler models underperform. Unlike SHAP, which provides feature interaction insights at the sample level, TSNN and TSGNN directly extract pairwise feature interactions. As parameterization increased, the likelihood of consistent weights occurring by chance decreased, likely indicating the revelation of more true feature interactions. Weight consistency was higher for TSGNN than for TSNN, likely due to its shallower structure (two nodes to output versus four), which reduced variation. Another possible explanation for this observation is that TSGNN reveals partial rather than exclusive interactions, increasing the number of observed relationships. In summary, TSNN can identify exclusive unidirectional feature interactions but has less consistent results than TSGNN, with both models showing greater effectiveness at higher parameterization.

The observed randomness in TSNN/TSGNN weights across different initializations may stem from the delicate balance each feature must achieve for individualized predictions, where adjusting a single weight to suit one prediction may adversely affect others. Features may exert both positive and negative influences, which is orchestrated such that they only become influential under certain contexts. Such intricate interplay leads to complex interactions that obscure interpretability.^28^ Different model initializations may converge to different stable states (local minima) that achieve similar performance through varied information processing pathways. Weights that tend to remain consistent under different local minima are more likely to represent true feature inter-relationships rather than balancing attempts to establish context.

Feature contributions derived from TSNN and TSGNN showed moderate correlations with SHAP values, with higher correlations observed with TSGNN and at higher parameterization. It is noteworthy that this route-based additive approach is fundamentally different than SHAP, which estimates feature influence by perturbing model predictions using various feature subsets.^17^ As previously explained, each feature conveys both positive and negative influences on the output, with many of these effects balancing each other out. Summing the contribution pathways passing through a given feature helps capture this process, providing a representation of the feature’s overall impact. However, each contribution route also contains other features, introducing noise. Increasing the number of contribution routes may ‘smooth out’ noise signals and reveal the true influence of a given feature, providing a plausible explanation for why correlation with SHAP values improved as parameterization increased. Compared to TSGNN, each TSNN route contains more noise signals (4 features versus 2 per route), thus TSGNN correlated better with SHAP at similar levels of parameterization. It can be postulated that with more complex models incorporating larger sample and feature numbers, this route-based calculation method will show greater validity at estimating feature influence, offering a computationally efficient alternative to SHAP values.

Interestingly, multiple logistic regression underperformed compared to most ML models when using only Class 1 (AUC=0·674 ± 0·047), but significantly improved with the full feature pool (AUC=0·762 ± 0·027). This finding is somewhat counterintuitive, as increased feature interrelationships typically favor ML models over simpler statistical models.^30^ A plausible explanation is that the Class 1 pool lacked sufficient direct links to RRI, limiting logistic regression’s effectiveness, whereas more flexible ML models could extract indirect information. When the feature pool was expanded, numerous direct links emerged, enhancing logistic regression’s performance, while ML models’ performances were constrained by the limited number of training samples. Notably, only thirteen features overlapped between the Class 1 (21 features) and the full pool (95 features) for multiple logistic regression, indicating that much of the data in the larger pool was unavailable in the smaller pool. This underscores that model performance is contingent on multiple factors, and in certain scenarios, simpler statistical models can rival complex models in performance while offering greater interpretability.^3^ Consequently, it is advisable to evaluate multiple models rather than presuppose the superiority of specific models.

This study has several limitations. The participant recruitment strategy may have attracted runners with recent injuries, potentially introducing selection bias. The predominantly UK-based sample limits external validity across different ethnicities. The ML models employed varied computational loads, allowing for different numbers of hyperparameter combinations, which may favor certain models. For TSNN and TSGNN, some hyperparameters (e.g., attention weights, batch normalization, oversampling) were fixed to facilitate feature number and learning rate exploration, possibly overlooking superior hyperparameter configurations. Due to limitations in data collection, only a small number of history features were available, and the temporal resolution of many behavior features (i.e., nutrition features) were low, potentially resulting in suboptimal TSNN/TSGNN performance.

This study represents the first attempt to design ML source algorithms and interpretation strategies from a prognostic rather than a purely mathematical perspective. TSNN and TSGNN provide unique insights into risk factor interactions while maintaining performance levels comparable to traditional ML models. Both models improve in validity and interpretability with increasing parameterization, particularly in feature influence estimation and pairwise interaction extraction. This suggests that their interpretability could further improve if trained on larger, more complex datasets. Future prognostic modeling studies could consider integrating TSNN and TSGNN alongside conventional ML models for additional assessment.

## Supporting information

Class 1 Dataset

All Features Dataset

## Data and Code Availability

Processed data is shared within the supplementary materials. Raw data is available upon contact, however some metrics (e.g. past performance records) need to remain normalized to ensure participant anonymity. Code is available via GitHub page: <u>henrywu0709/TSNN-TSGNN-for-prognostic-modelling</u>.

## Acknowledgements

This research was supported by the Alan Turing Institute Enrichment Scheme and the China Scholarship Council.

Special thanks to Dr. Hui Fang for his technical support.

## Authors’ Contributions

HW contributed to the ideation of ML, genetic, and biomechanics-related content, plus data collection and analysis. RB contributed to the ideation of study structure and strength, bone, and nutrition-related content, plus participant recruitment. KB advised on data collection and analysis. MB advised on genetic-related and ML-related content. ZA advised on ML-related content. SM advised on genetic-related content. SA advised on biomechanics-related content. All authors contributed to writing the manuscript. All authors have read and approved the final version of the manuscript and have agreed with the order of presentation of their names.

## References

1. Royston P, Moons KGM, Altman DG, Vergouwe Y. Prognosis and prognostic research: Developing a prognostic model. BMJ. 2009;338(mar31 1):b604–b604. doi:10.1136/bmj.b604

2. Dhiman P, Ma J, Andaur Navarro CL, et al. Risk of bias of prognostic models developed using machine learning: a systematic review in oncology. Diagn Progn Res. 2022;6(1):13. doi:10.1186/s41512-022-00126-w

3. Clift AK, Dodwell D, Lord S, et al. Development and internal-external validation of statistical and machine learning models for breast cancer prognostication: cohort study. BMJ. Published online May 10, 2023:e073800. doi:10.1136/bmj-2022-073800

4. Perveen S, Shahbaz M, Keshavjee K, Guergachi A. Prognostic Modeling and Prevention of Diabetes Using Machine Learning Technique. Sci Rep. 2019;9(1):13805. doi:10.1038/s41598-019-49563-6

5. Booth AL, Abels E, McCaffrey P. Development of a prognostic model for mortality in COVID-19 infection using machine learning. Modern Pathology. 2021;34(3):522–531. doi:10.1038/s41379-020-00700-x

6. Yoon CH, Torrance R, Scheinerman N. Machine learning in medicine: should the pursuit of enhanced interpretability be abandoned? J Med Ethics. 2022;48(9):581–585. doi:10.1136/medethics-2020-107102

7. Arora P, Boyne D, Slater JJ, Gupta A, Brenner DR, Druzdzel MJ. Bayesian Networks for Risk Prediction Using Real-World Data: A Tool for Precision Medicine. Value in Health. 2019;22(4):439–445. doi:10.1016/j.jval.2019.01.006

8. Ganian R, Korchemna V. The complexity of bayesian network learning: Revisiting the superstructure. In: ; 2021:430–442.

9. Nagalla S, Chou JW, Willingham MC, et al. Interactions between immunity, proliferation and molecular subtype in breast cancer prognosis. Genome Biol. 2013;14(4):R34. doi:10.1186/gb-2013-14-4-r34

10. Verhagen E. The cost of sports injuries. J Sci Med Sport. 2010;13:e40. doi:10.1016/j.jsams.2010.10.546

11. Van Eetvelde H, Mendonça LD, Ley C, Seil R, Tischer T. Machine learning methods in sport injury prediction and prevention: a systematic review. J Exp Orthop. 2021;8(1):27. doi:10.1186/s40634-021-00346-x

12. Saarela M, Jauhiainen S. Comparison of feature importance measures as explanations for classification models. SN Appl Sci. 2021;3(2):272. doi:10.1007/s42452-021-04148-9

13. Lövdal SS, Den Hartigh RJR, Azzopardi G. Injury Prediction in Competitive Runners With Machine Learning. Int J Sports Physiol Perform. 2021;16(10):1522–1531. doi:10.1123/ijspp.2020-0518

14. Bittencourt NFN, Meeuwisse WH, Mendonça LD, Nettel-Aguirre A, Ocarino JM, Fonseca ST. Complex systems approach for sports injuries: moving from risk factor identification to injury pattern recognition—narrative review and new concept. Br J Sports Med. 2016;50(21):1309–1314. doi:10.1136/bjsports-2015-095850

15. Raghunandan A, Charnoff JN, Matsuwaka ST. The Epidemiology, Risk Factors, and Nonsurgical Treatment of Injuries Related to Endurance Running. Curr Sports Med Rep. 2021;20(6):306–311. doi:10.1249/JSR.0000000000000852

16. Chakravarti A, Little P. Nature, nurture and human disease. Nature. 2003;421(6921):412–414. doi:10.1038/nature01401

17. Lundberg SM, Lee SI. A unified approach to interpreting model predictions. In: Advances in Neural Information Processing Systems. ; 2017:30.

18. Kakouris N, Yener N, Fong DTP. A systematic review of running-related musculoskeletal injuries in runners. J Sport Health Sci. 2021;10(5):513–522. doi:10.1016/j.jshs.2021.04.001

19. Weeks BK, Beck BR. The BPAQ: a bone-specific physical activity assessment instrument. Osteoporosis International. 2008;19(11):1567–1577. doi:10.1007/s00198-008-0606-2

20. Luce KH, Crowther JH. The reliability of the eating disorder examination-Self-report questionnaire version (EDE-Q). International Journal of Eating Disorders. 1999;25(3):349–351. doi:10.1002/(SICI)1098-108X(199904)25:3<349::AID-EAT15>3.0.CO;2-M

21. Melin A, Tornberg ÅB, Skouby S, et al. The LEAF questionnaire: a screening tool for the identification of female athletes at risk for the female athlete triad. Br J Sports Med. 2014;48(7):540–545. doi:10.1136/bjsports-2013-093240

22. Brody DM. Techniques in the evaluation and treatment of the injured runner. Orthop Clin North Am. 1982;13(3):541–558.

23. Clarsen B, Bahr R, Myklebust G, et al. Improved reporting of overuse injuries and health problems in sport: an update of the Oslo Sport Trauma Research Center questionnaires. Br J Sports Med. 2020;54(7):390–396. doi:10.1136/bjsports-2019-101337

24. Pascanu R, Mikolov T, Bengio Y. On the difficulty of training Recurrent Neural Networks. Published online November 21, 2012.

25. Kourou K, Exarchos TP, Exarchos KP, Karamouzis M V., Fotiadis DI. Machine learning applications in cancer prognosis and prediction. Comput Struct Biotechnol J. 2015;13:8–17. doi:10.1016/j.csbj.2014.11.005

26. Nguyen NH, Picetti D, Dulai PS, et al. Machine Learning-based Prediction Models for Diagnosis and Prognosis in Inflammatory Bowel Diseases: A Systematic Review. J Crohns Colitis. 2022;16(3):398–413. doi:10.1093/ecco-jcc/jjab155

27. Lillis C. Exploring Machine Learning, Real-Time Bio-Feedback, and Inertial Sensor Accuracy for the Prevention of Running-Related Injuries. Dublin City University; 2022.

28. Baldassi C, Gerace F, Kappen HJ, et al. Role of Synaptic Stochasticity in Training Low-Precision Neural Networks. Phys Rev Lett. 2018;120(26):268103. doi:10.1103/PhysRevLett.120.268103

29. Spiriev B. World Athletics scoring tables.

30. Steyerberg EW, van der Ploeg T, Van Calster B. Risk prediction with machine learning and regression methods. Biometrical Journal. 2014;56(4):601–606. doi:10.1002/bimj.201300297

