## Supplementary Material for "A time-sequenced approach to machine learning prognostic modelling with implementation on running-related injury prediction"

**Questionnaire Contents**

**Baseline Questionnaire**

**Basic Information**

1. Please enter your first name.

2. Please enter your last name.

3. Please select your date of birth.

4. Please select your biological sex. This refers to your physiological sex at the time of birth.

4.a. If you selected Other, please specify:

5. Please select the date you complete this questionnaire (the date today)

**Health Screen**

6. At present, do you have any health problem for which you are:

6.1.a. (a) on medication, prescribed or otherwise

7. Have you ever had any of the following:

7.1.a. (a) Convulsions/epilepsy

7.2.a. (b) Asthma

7.3.a. (c) Eczema

7.4.a. (d) Diabetes

7.5.a. (e) A blood disorder

7.6.a. (f) Head injury

7.7.a. (g) Digestive problems

7.8.a. (h) Heart problems/chest pains

7.9.a. (i) Problems with muscles, bones or joints

7.10.a. (j) Disturbance of balance/coordination

7.11.a. (k) Numbness in hands or feet

7.12.a. (l) Disturbance of vision

7.13.a. (m) Ear/hearing problems

7.14.a. (n) Thyroid problems

7.15.a. (o) Kidney or liver problems

7.16.a. (p) Problems with blood pressure

7.a. If YES to any question, please describe briefly if you wish (eg to confirm problem was/is short-lived, insignificant or well controlled.)

8. Smoking, physical activity and family history

8.1.a. (a) Are you a current or recent (within the last six months) smoker or vaper?

9. Allergy Information

9.1.a. (a) Are you allergic to any food products?

9.2.a. (b) Are you allergic to any medicines?

9.3.a. (c) Are you allergic to plasters?

9.4.a. (d) Are you allergic to latex?

9.a. If YES to any of the above, please provide additional information on the allergy

10. Additional questions for female participants

10.1.a. (a) Are your periods normal/regular?

10.2.a. (b) Are you on hormonal contraception

10.3.a. (c) Are you taking hormone replacement therapy (HRT)?

11. Stress fracture history

11.1.a. (a) Have you ever been diagnosed with a bone stress injury (stress fracture, stress reaction, stress response)?

11.a. If yes, please provide details of type of injury, age of occurrence, anatomical location, time of year and method of diagnosis.

12. Research involvement

12.1.a. (a) Are you currently involved in any other research studies at the University or elsewhere?

12.a. If yes, please provide details.

13. Please provide the name of your emergency contact in the event of any incident or emergency.

13.a. Telephone number

13.b. Relationship to participant

**Past 12-month Injuries**

14. Please indicate the location of the problem(s) you have experienced over the last 12 months and the number of days you were forced to reduce your normal running routine because of each problem:

14.1.a. 1 - Location of the problem

14.1.b. 1 - Number of days the problem lasted

14.1.c. 1 - Did you visit a healthcare professional (e.g. physiotherapist), to obtain a diagnosis?

14.2.a. 2 - Location of the problem

14.2.b. 2 - Number of days the problem lasted

14.2.c. 2 - Did you visit a healthcare professional (e.g. physiotherapist), to obtain a diagnosis?

14.3.a. 3 - Location of the problem

14.3.b. 3 - Number of days the problem lasted

14.3.c. 3 - Did you visit a healthcare professional (e.g. physiotherapist), to obtain a diagnosis?

14.4.a. 4 - Location of the problem

14.4.b. 4 - Number of days the problem lasted

14.4.c. 4 - Did you visit a healthcare professional (e.g. physiotherapist), to obtain a diagnosis?

14.5.a. 5 - Location of the problem

14.5.b. 5 - Number of days the problem lasted

14.5.c. 5 - Did you visit a healthcare professional (e.g. physiotherapist), to obtain a diagnosis?

14.6.a. 6 - Location of the problem

14.6.b. 6 - Number of days the problem lasted

14.6.c. 6 - Did you visit a healthcare professional (e.g. physiotherapist), to obtain a diagnosis?

14.7.a. 7 - Location of the problem

14.7.b. 7 - Number of days the problem lasted

14.7.c. 7 - Did you visit a healthcare professional (e.g. physiotherapist), to obtain a diagnosis?

14.8.a. 8 - Location of the problem

14.8.b. 8 - Number of days the problem lasted

14.8.c. 8 - Did you visit a healthcare professional (e.g. physiotherapist), to obtain a diagnosis?

14.9.a. 9 - Location of the problem

14.9.b. 9 - Number of days the problem lasted

14.9.c. 9 - Did you visit a healthcare professional (e.g. physiotherapist), to obtain a diagnosis?

14.10.a. 10 - Location of the problem

14.10.b. 10 - Number of days the problem lasted

14.10.c. 10 - Did you visit a healthcare professional (e.g. physiotherapist), to obtain a diagnosis?

14.a. If there is any further information you would like to add, please describe below:

**Bone-specific Physical Activity Questionnaire (BPAQ): standard questionnaire; see references within article**

**Athletic Performance Level**

17. Please list the fastest times you have recorded in races or time trials over the following distances in the last 6 months:

17.1.a. 5000m track - Time

17.2.a. 5km road - Time

17.3.a. 10,000m track - Time

17.4.a. 10km road - Time

17.5.a. Half Marathon - Time

17.6.a. Marathon - Time

17.7.a. Other distance (5km or further): - Time

17.a. If there is any further information you would like to specify, please describe:

**Past 12-month Training and S&C**

18. Typically, how many hours did you run per week over the last 12 months during the following training phases?

18.1.a. Preparatory (off-season) period: - hours per week

18.2.a. Competitive (in-season or tapering) period: - hours per week

19. Typically, how often (runs per week) did you run per week over the last 12 months during the following training phases?

19.1.a. Preparatory (off-season) period: - times per week

19.2.a. Competitive (in-season or tapering) period: - times per week

20. Typically, how often have you performed intensive running sessions (speeds faster than half-marathon intensity, i.e. interval training and tempo running) per week over the last 12 months during the following training phases?

20.1.a. Preparatory (off-season) period: - times per week

20.2.a. Competitive (in-season or tapering) period: - times per week

21. In addition to your running sessions, did you include any other forms of aerobic exercise training over the last 12 months, and if so, typically how many hours and how often per week (sessions per week)?

21.1.a. Swimming - Yes/No

21.1.b. Swimming - If yes, hours per week

21.1.c. Swimming - If yes, how often per week

21.2.a. Cycling - Yes/No

21.2.b. Cycling - If yes, hours per week

21.2.c. Cycling - If yes, how often per week

21.3.a. Rowing - Yes/No

21.3.b. Rowing - If yes, hours per week

21.3.c. Rowing - If yes, how often per week

21.4.a. Cross-trainer - Yes/No

21.4.b. Cross-trainer - If yes, hours per week

21.4.c. Cross-trainer - If yes, how often per week

21.5.a. Other1 - Yes/No

21.5.b. Other1 - If yes, hours per week

21.5.c. Other1 - If yes, how often per week

21.6.a. Other2 - Yes/No

21.6.b. Other2 - If yes, hours per week

21.6.c. Other2 - If yes, how often per week

21.7.a. Other3 - Yes/No

21.7.b. Other3 - If yes, hours per week

21.7.c. Other3 - If yes, how often per week

21.a. Plese specify what "other" exercises refer to, if applicable:

21.a.1.a. other1 - Name of exercise

21.a.2.a. other2 - Name of exercise

21.a.3.a. other3 - Name of exercise

22. Over the last 12 months, have you participated in the following training activities, and if so, typically how many hours and how often per week (sessions per week)?

22.1.a. Resistance training (i.e. free weights, kettlebells, machines, elastic bands, medicine balls) - Yes/No

22.1.b. Resistance training (i.e. free weights, kettlebells, machines, elastic bands, medicine balls) - If yes, hours per week

22.1.c. Resistance training (i.e. free weights, kettlebells, machines, elastic bands, medicine balls) - If yes, how often per week

22.2.a. Plyometrics (i.e. jumping, hopping, bounding) - Yes/No

22.2.b. Plyometrics (i.e. jumping, hopping, bounding) - If yes, hours per week

22.2.c. Plyometrics (i.e. jumping, hopping, bounding) - If yes, how often per week

22.3.a. Core stability (i.e. exercises that specifically target the trunk/abdominal region) - Yes/No

22.3.b. Core stability (i.e. exercises that specifically target the trunk/abdominal region) - If yes, hours per week

22.3.c. Core stability (i.e. exercises that specifically target the trunk/abdominal region) - If yes, how often per week

22.4.a. Bodyweight exercises (i.e. burpees, press-ups, lunges without external load) - Yes/No

22.4.b. Bodyweight exercises (i.e. burpees, press-ups, lunges without external load) - If yes, hours per week

22.4.c. Bodyweight exercises (i.e. burpees, press-ups, lunges without external load) - If yes, how often per week

22.5.a. Stretching or yoga - Yes/No

22.5.b. Stretching or yoga - If yes, hours per week

22.5.c. Stretching or yoga - If yes, how often per week

23. Other than the activities mentioned above, were there any other sports or other physical activities (be as specific as possible) that you participated in regularly over the last 12 months, and if so, typically how many hours and how often per week (sessions per week)?

23.1.a. Activity1 - Name of activity

23.1.b. Activity1 - Hours per week

23.1.c. Activity1 - How often per week

23.2.a. Activity2 - Name of activity

23.2.b. Activity2 - Hours per week

23.2.c. Activity2 - How often per week

23.3.a. Activity3 - Name of activity

23.3.b. Activity3 - Hours per week

23.3.c. Activity3 - How often per week

23.4.a. Activity4 - Name of activity

23.4.b. Activity4 - Hours per week

23.4.c. Activity4 - How often per week

23.5.a. Activity5 - Name of activity

23.5.b. Activity5 - Hours per week

23.5.c. Activity5 - How often per week

23.6.a. Activity6 - Name of activity

23.6.b. Activity6 - Hours per week

23.6.c. Activity6 - How often per week

23.7.a. Activity7 - Name of activity

23.7.b. Activity7 - Hours per week

23.7.c. Activity7 - How often per week

24. If there is additional information you would like to specify, please describe:

**Eating Disorder Examination Questionnaire (EDE-Q) ): standard questionnaire; see references within article**

**Low Energy Availability in Females Questionnaire (LEAF-Q) ): standard questionnaire; see references within article**

### Weekly Questionnaire

Q20_1. How many kilometers did you run at the following intensities during the past 7 days? - Low-moderate intensity (easy/recovery run; can hold conversation while running; 1-4 out of 10 perceived exertion)

Q20_2. How many kilometers did you run at the following intensities during the past 7 days? - Steady/heavy intensity (tempo run or around half-marathon pace; can say no more than 5 words while running; 5-6 out of 10 perceived exertion)

Q20_3. How many kilometers did you run at the following intensities during the past 7 days? - Severe/High intensity (intervals at 10km – 1500m/mile pace; very hard to talk while running; 7-10 out of 10 perceived exertion)

Q20_4. How many kilometers did you run at the following intensities during the past 7 days? - Very high intensity (sprint intervals at faster than 1500m/mile pace)

Q23_1. What is the typical speed you ran at the above-mentioned intensities (kilometers/hour)? - Low-moderate intensity (easy/recovery run; can hold conversation while running)

Q23_2. What is the typical speed you ran at the above-mentioned intensities (kilometers/hour)? - Steady/heavy intensity (tempo run or around half-marathon pace; can say no more than 5 words while running)

Q23_3. What is the typical speed you ran at the above-mentioned intensities (kilometers/hour)? - Severe/High intensity (intervals at 10km – 1500m/mile pace; very hard to talk while running)

Q23_4. What is the typical speed you ran at the above-mentioned intensities (kilometers/hour)? - Very high intensity (sprint intervals at faster than 1500m/mile pace)

Q30_1. How many minutes of non-running exercises did you perform during the past 7 days that could be represented by the descriptions below? - resistance training (e.g. free weights, kettlebells, machines, elastic bands, medicine balls)

Q30_2. How many minutes of non-running exercises did you perform during the past 7 days that could be represented by the descriptions below? - bodyweight exercises (e.g. unweighted lunges, wall squats)

Q30_16. How many minutes of non-running exercises did you perform during the past 7 days that could be represented by the descriptions below? - core stability exercises (e.g. planks, side planks, swiss ball-based, exercises specifically targeting trunk/abdominal region)

Q30_19. How many minutes of non-running exercises did you perform during the past 7 days that could be represented by the descriptions below? - balance training (e.g. exercises on an unstable surface)

Q30_3. How many minutes of non-running exercises did you perform during the past 7 days that could be represented by the descriptions below? - plyometric exercises (e.g. jumping, hopping, bounding)

Q30_18. How many minutes of non-running exercises did you perform during the past 7 days that could be represented by the descriptions below? - running technique drills (e.g. A- and B- drills, dribbles)

Q30_5. How many minutes of non-running exercises did you perform during the past 7 days that could be represented by the descriptions below? - circuit training

Q30_20. How many minutes of non-running exercises did you perform during the past 7 days that could be represented by the descriptions below? - barefoot exercises

Q30_6. How many minutes of non-running exercises did you perform during the past 7 days that could be represented by the descriptions below? - stretching or yoga

Q30_12. How many minutes of non-running exercises did you perform during the past 7 days that could be represented by the descriptions below? - swimming

Q30_13. How many minutes of non-running exercises did you perform during the past 7 days that could be represented by the descriptions below? - cycling

Q30_14. How many minutes of non-running exercises did you perform during the past 7 days that could be represented by the descriptions below? - rowing

Q30_17. How many minutes of non-running exercises did you perform during the past 7 days that could be represented by the descriptions below? - cross-trainer/elliptical machine

Q30_7. How many minutes of non-running exercises did you perform during the past 7 days that could be represented by the descriptions below? - Other (please specify)

Q30_7_TEXT. How many minutes of non-running exercises did you perform during the past 7 days that could be represented by the descriptions below? - Other (please specify) - Text

Q32. If there are any other details regarding the exercises you completed during the past 7 days you would like to specify, please describe below:

Acknowledgement. Please answer all following questions regardless of whether or not you have physical problems. Select the alternative that is most appropriate for you, and in

the case that you are unsure, try to answer as best you can anyway.
The term "physical problems" refers to pain, ache, stiffness, clicking/catching, swelling, instability/giving way, locking, or other complaints related to your joint, bone, tendon, ligament, or muscle.

1_Q3. Have you had any difficulties participating in training and competition due to (other) physical
problems during the past 7 days? ("The past 7 days" refers to the 7 consecutive days on and before the day this questionnaire is released, which is Sunday on each week)

1_Q4. Please select the location that best represents your physical problem. Select one, and you will come back to this question if you have more than one physical problems. - Selected Choice

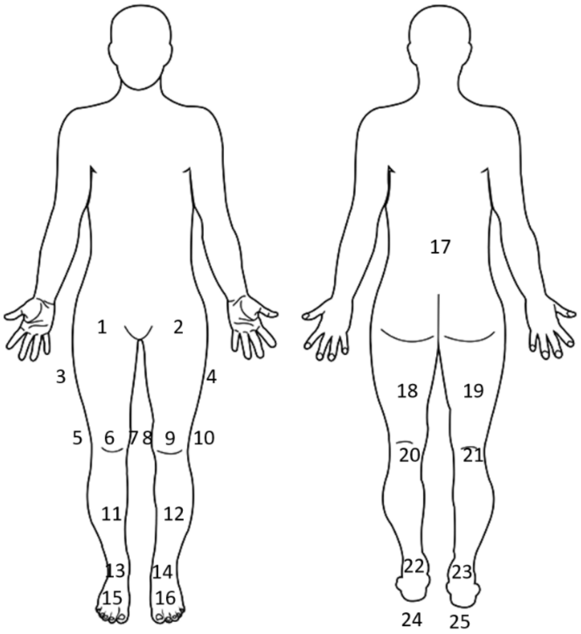

1_Q4_26_TEXT. Please select the location that best represents your physical problem. Select one, and you will come back to this question if you have more than one physical problems. - Other (please specify; you can use descriptions such as "between area X and area X", "at the X part of my body", or use anatomical descriptions if you are familiar with them) – Text

1_Q6. Have you had any difficulties participating in training and competition due to your selected problem during the past 7 days?

1_Q7. To what extent have you modified your training or competition due to your selected problem during the past 7 days?

1_Q8. To what extent has your selected problem affected your performance during the past 7
days?

1_Q9. To what extent have you experienced pain at your selected location related to your sport during the
past 7 days?

1_Q10. How would you describe the cause of your selected physical problem?

1_Q11. Would you describe your selected physical problem as caused by or related to running?

1_Q33. Have you obtained a diagnosis by a qualified healthcare professional (i.e. physician or physiotherapist) for your selected physical problem?

1_Q34. Please specify your diagnosis below.

1_Q12. If there are any other details regarding your selected physical problem you would like to specify, please describe below:

2_Q3. Have you had any difficulties participating in training and competition due to (other) physical
problems during the past 7 days? ("The past 7 days" refers to the 7 consecutive days on and before the day this questionnaire is released, which is Sunday on each week)

Note: all the injury-related questions will be looped if answered ‘yes’ when this question appears. Otherwise, the questionnaire is completed.

**Experimental Protocols**

### Bone Scans

**DXA and pQCT measurement were made according to standard manufacturer guidelines.**

### Posture Measurements

#### Q Angle Measurement SOP

- Ask the participant to take off their shoes, roll up their pants to above the knee, and lie supine on the DXA bench with feet just hanging over the side of the bench.
- Move the participant's legs so that their femurs are horizontal to the midline of the body (or vertical to the line drawn between the two ASISs). Rotate the participant's legs so that their patella face directly anterior.
- Ask the participant to try staying in this position while relaxed. Note that the participant should not contract their quadriceps as it would disturb the patellar angle. If the participant cannot maintain relaxed with patella facing anteriorly, allow a small extent of rotation.
- Use a marker to mark 3 points on each patella: top left, top right, and bottom corner, forming an inverse triangle. Visually determine the midpoint of each triangle and mark an additional point.
- Use the marker to mark the participant's centre of tibial tuberosity on both sides.
- Put a thread through the centre of the goniometer. Put the centre of the goniometer at the marked point in the middle of the left patella.
- Palpate the participant's left ASIS and ask the participant to hold the other end of the thread onto the left ASIS. This line is used as a reference line to align the goniometer arm.
- Keeping the centre of the goniometer still, rotate the two arms to align with: the thread the participant is holding, and the mark at the centre of the left tibial tuberosity. Read the angle of measurement at this point.
- Repeat the same process for the right side. Note down both Q angles.

#### Navicular Drop Measurement SOP

- Ask the participant to take off shoes and socks and then stay in a comfortable seated position.

- Adjust the participant's feet position so that their tibias are vertical to the ground. The participant's mass should be on their butt, meaning they should not be placing weight on their feet.
- Palpate the navicular bone (protruding bone on the medial side of the foot) and use a marker to mark the most prominent protrusion on both sides.
- Place the marking paper vertically on the ground, kneel to get a horizontal view of the navicular, and draw on the paper the level of the marked point. Repeat this for the other side (remember to mark left and right on the paper clearly before measuring to prevent confusion).
- Ask the participant to stand up in a comfortable position. Re-measure the navicular heights in this standing position. Seated and standing navicular heights of the same side are marked on the same edge on the marking paper.
- Use a ruler to measure the seated and standing navicular heights, as well as the difference between the two.

### Treadmill GRF Test SOP

- Turn on the AMTI and the Vicon systems. Create folders for the participant's data, with folder name being the participant's study ID and the trial name being study ID + speed (e.g. BHEAXXX 10km).
- Turn on the treadmill but do not start it. Stand on the treadmill to instruct the participant of the procedures.
- Let the participant see the edges of the measurement forceplate and instruct them to always run within the forceplate's range. The participant should initially stand with feet wide open on the side edges so the forceplate is not disturbed. After the treadmill is started and runs at the target speed, the researcher should zero the forceplate and then give a go signal, after which the participant can step on the forceplate and start running. Instruct the participant to be mindful of the risk of tripping while stepping onto a running treadmill.
- Once the participant is clear of the procedures, instruct them to stand on the treadmill with feet wide open on the edges. Start the treadmill at 10km/h.
- Within Vicon Nexus, click 'Go Live' on the top left corner. Check the tabloid below to ensure the treadmill forceplate data is connected and running properly. If not, try going offline and live again, or restarting the Vicon Nexus.
- On the middle screen, click the options on the top left corner and change from '3D Perspective' to 'Graph'. 3D Perspective is for motion capture which is not needed in this case.
- The graphs should show the real time forces Fx/y/z 1-4 which is data for each sensor's each dimension. If the horizontal/vertical scale of the graph is

inconvenient for observation, click the rescaling options for the respective dimension on the middle top section to rescale.
- On the left tabloid, find the treadmill forceplate data ‘Treadmill (Generic Analog)’. Right click and choose ‘zero level’ to zero the forceplate at 10km/h.
- After zeroing, give the participant a go signal and let them start running.
- Use a timer to time the participant’s run. Data recording should start 1.5-2min after the participant starts running. On the right tabloid, under ‘tools’, click the middle ‘Capture’ option, and select the pre-set trial type for this study (‘Henry’).
- Once 1.5-2min has passed and the participant is accustomed to the run, click ‘start’ on the right-side tabloid to start recording. Data recording should stop automatically after 1min.
- Let the participant stop and stand with feet wide open again the same as before. Adjust the treadmill speed to 12km/h, re-zero, and repeat the above process.
- After finishing the 12km/h trial, let the participant stop and stand on the edges, stop the treadmill, and ask the participant to safely come down the treadmill.

**Data Export**

- On the middle lower tabloid of the screen, select the trial to be exported by double-clicking.
- On the right tabloid of the screen, click the right-most button ‘Pipeline’.
- Choose the pre-set pipeline for the current study (‘Henry’) which exports ASCIII and C3D of the forceplate data.
- Click the start button to export.
- The exported files are saved on disk F. Copy the exported file to a hard drive.

### Isokinetic Dynamometer SOP

**Start-up and preparation**

- Start up the Isomed 2000 dynamometer by first using the key to unlock the emergency stop button on the right side of the start-up switches, then twist the left start-up switch from ‘off’ to ‘on’, and finally press the middle green button. Sometimes the green button needs to be pressed twice to start.
- Wait for the attached computer to start up, when prompted whether to create a new backup, choose ‘2’ to proceed directly to starting up.

**Hip Abduction/Adduction Measurements**

All directional arguments (such as superior/inferior) below are relative to the participant's subjective position.

- At the initial menu of the computer, go to 'testing/training' – 'library' – 'new library training' – 'patient' – 'henry' – 'test-program' – 'henry pros study'. This will load the pre-established testing sequence created for this study.
- Choose 'edit' – 'assume-all' to load the testing sequence to the testing interface.
- Choose 'starting testing/training', and then enter the participant's identifier (BHEA+number) onto patient name. This will automatically create a new patient profile using the unique identifier.
- Choose 'test-program' – 'automatical process' to initiate the testing sequence.
- Adjust the testing bench to a flat surface. The upper end of the bench could be adjusted using the 'back up' and 'back down' buttons. The lower end of the bench could be adjusted using the adjustment pin below the inferior end of the bench.
- Ask the participant to lie on their left side on the bench and then measure the participant's thigh length. Thigh length is measured using a tape measure, starting from the lateral side of the greater trochanter to the middle of the patella. Write down the participant's thigh length for later use.
- Move the computer screen to a position in front of the participant to provide real-time feedback.
- Press and hold the blue 'swivel arm brake open/close' button to unlock swivel arm rotation, move the swivel arm to the posterior side of the participant while observing the 'current position' shown on the swivel arm aligns with the 'referred position'.
- Instruct the participant to move superiorly/inferiorly so that the greater trochanter of the participant aligns with the pivot on the swivel arm.
- Use the 'dynamometer height' button to adjust the swivel arm height so that the pivot of the swivel arm is approximately 5cm higher than the participant's greater trochanter.
- Attach the fixation tools '3' and 'D' onto the swivel arm. Adjust the exposed length of '3' so that the lever arm length is equal to the participant's thigh length. At 'MAX', lever arm length is 40cm, and each number below 'MAX' refers to 1cm shorter than 40cm. If the participant's thigh length is greater than 40cm, pull the tool 2-3cm longer than 'MAX' which is the closest possible approximation of thigh length.
- Fix the participant's knee with tool 'D' so that the middle adjustment pin is aligned with the middle of the participant's patella. The participant can further adjust position superiorly/inferiorly at this point to find a comfortable position. 'D' needs to be tightly strapped to enable adduction at full force.

- Inform the participant to move the left thigh 45 degrees forward (anteriorly). Use the fixation tool 'B' and the unlabelled thigh fixation tool to stabilise the participant's thigh. The tools can be adjusted at different dimensions to comfortably fit the participant. Avoid fixing (pushing down) on the participant's knee joint or distal to the knee joint as doing so will cause instability and pain during adduction. Fix only on the participant's thigh. Push down the fixation tool tightly to enable adduction at full force.
- Press 'esc' – 'esc' to not make modifications to default positional values.
- The screen will now instruct to move the adaptor to the end of range of motion 'E'. Press and hold the white 'dynamometer lever release' button to unlock adaptor motion. Hold the adaptor's end up (in the direction of abduction) while also letting the participant perform abduction. Make sure that the participant can reach the end of the ROM. Otherwise adjust the participant's position.
- After reaching 'E', return to the middle of ROM and release the 'dynamometer lever release' button. The screen now prompts to move the adaptor to the starting position 'A' and to set mechanical stoppers in place to prevent over motion. Set mechanical stoppers first as moving to position 'A' will directly initiate testing.
- Before moving the adaptor to 'A', inform the participant of the testing procedures, as the automatic testing sequence cannot be paused easily.
- The testing procedure involves 2 practice sets and 5 testing sets. Each set includes 4 reps of alternating abduction and adduction. Before each set begins, the swivel arm will exert an abduction force slightly larger than the participant's leg weight. The participant should forcefully hold their leg in an adduction starting position until the countdown is finished and the lifting force disappears. During the countdown, a place indicator is shown on the computer screen. The participant should try to keep the indicator within the middle green range which indicates the adducted starting position.
- During the practice set, the participant should use ~80% of maximum force to get used to the movement. Instruct the participant to forcefully push both up and down in case some may think only one of them is being tested. Between each set, there will be a 1 minute break.
- After putting mechanical stoppers on and instructing participants, press the 'dynamometer lever release' button and move the participant's leg to the 'A' position.
- Before the first set, a prompt will appear to measure the participant's leg weight. Hold the 'dynamometer lever release' button to move the participant's leg to a horizontal position, and then release the button to let the participant's leg stay in that position. Instruct the participant to fully relax the leg, and then press 'enter' to measure leg weight. After weight measurement, the first set will automatically begin.
- During the practice sets, if the participant is not used to holding the testing-leg down forcefully, help the participant push the adaptor down and verbally walk

through the process with the participant. Release the adaptor once the countdown ends and let the participant complete the set.

- If the participant does not feel familiar with the process after the practice sets, exit the testing sequence (press 'esc' to exist and then press 'enter' to interrupt training, and then press 'esc' to exit to main screen) and re-initiate to give 2 more practice sets.
- During testing sets, observe whether the participant's leg hits the bench and bounces at the bottom. Excessive bouncing can cause the dynamometer to mistakenly recognise a bounce as a rep. If bouncing happens frequently, stop the testing sequence, adjust the participant's position inferiorly, and re-initiate.
- During testing sets, the participant's performance typically increases during the first 2 sets due to familiarisation effect and stabilises during sets 3-5. If the participant's performance kept increasing through sets 4 and 5, ask the participant whether they feel like they could do better with additional sets, and if yes, exit and re-initiate the testing sequence and discard the previous sets. Caution should be placed when taking extra sets to prevent fatigue.
- After completing all testing sets, the screen will prompt for the next exercise, which is the same abduction/adduction exercise for the left. Switch everything to the other side and repeat the testing procedure.

**Knee Flexion/Extension Measurements**

After hip abduction/addition measurements, the testing sequence will automatically prompt for right knee flexion/extension measurement.

- Ask the participant to come down from the bench to take a break.
- Press and hold the 'back up' button to move the upper part of the bench up, stop at about 80 degrees as shown on the swivel arm screen. Lower the fixation stick at the back and then press and hold 'back down' to let the upper part of the bench go down and be stopped by the fixation stick at 75 degrees.
- Let the participant sit on the lower part of the bench. Press and hold the 'forward' button to move the back of the bench forward. Let the participant indicate when their back is fully against the back of the bench and stop at that point. The participant should have their knee hanging on the edge of the bench and there should be no space between the back of knee and the edge.
- Measure the participant's lower leg length with a tape measure. The lower leg length is the distance between the knee joint space and the middle of the lateral malleolus. Write down the measurement result for later use.
- Move the swivel arm to align the 'current position' with the 'referred position'. Differently than with abduction/adduction, since different people have different leg and knee sizes, the position should differ slightly among individuals and thus

should be adjusted away from 'referred position' on an individual basis. Attach the laser shooter at the top of the swivel arm to the pivot point of the lever and use the laser to find an appropriate position. The laser light should be positioned in the middle of the knee joint shooting at knee joint space. The laser shooter should be horizontal to the edge of the bench.

- Once the swivel arm position is settled, remove the laser shooter. Attach fixation tools '3' and 'F' to the swivel arm. Through testing, a typical comfortable position is at a lever arm length of lower leg length – 9cm. For instance, for a person whose lower leg length is 43cm, the length of the lever arm should be 35cm, which is 5cm shorter than the 'MAX' mark. The fixation tool should be strapped tightly right above the ankle joint.
- Insert and fixate the 2 handles 'I' on the left and right sides of the participant to provide holding objects while exerting forces. Adjust the handles' position so that the participant can comfortably exert forces while holding the handles.
- Use a thigh strap to strap the participant's thigh onto the edge of the bench. Strap tightly to enable forceful knee flexion.
- Instruct the participant to 'point toes to the sky' while performing knee flexion. This is to ensure a standardised gastrocnemius position during knee flexion to control for calf muscle participation.
- Once the participant is fully prepared, press 'enter' to initiate the testing sequence. Prompts will appear again to move the participant's leg to 'E' position, put mechanical stoppers on, and then move to 'A' position. Before the first practice set, the screen will prompt to measure weight of the lower leg. Make sure the participant's lower leg is lifted to a horizontal position, freeze the adaptor, let the participant fully relax, and then initiate lower leg weight measurement.
- The testing sequence is generally the same as the abduction/adduction sequence. The thigh strap will likely become loose during testing, especially with stronger participants. Re-strap the thigh strap between sets if they come loose.
- Participants may slide anteriorly to compensate. If the participant's butt moves forward and is not against the back of the bench, instruct the participant to move back and have their whole back against the back of the bench.
- After right leg flexion/extension measurements, move everything to the other side and repeat the process with the left leg.

**Data Export**

- At the main screen, go to 'data evaluation' – 'depiction mode' – 'reconstruction' and enter the participant's ID.
- Go to 'choice filters' and select the target exercise. The exercises are listed in time sequence. For each unique movement, there will typically be a 2 sets practice exercise and a 5 sets testing exercise. Choose the latter. If there are more than 2

exercises for a specific movement, this is due to the participant repeating the testing sets. Only take the last exercise with 5 sets.
- After selecting the exercise, go to ‘activate’. Write down the max torque and degree at max torque for each rep, then go to the next set and do the same. Repeat for all 5 sets.
- Repeat the previous step for all 4 movements (hip abduction/adduction for both sides, knee flexion/extension for both sides).

### Food Diary Instructions

- Between this visit and the next, we would like to ask you to do 3 full days of food diary.
- The 3 days should be 2 weekdays and 1 weekend day. These 3 days do not need to be consecutive, so they can be any day as long as they fall between the 2 visits.
- The reason we ask for 2 weekdays and 1 weekend day is because people have different dietary habits during weekdays versus weekends. If you have a fixed diet for both weekdays and weekends, you can pick any day you like.
- If you are on an intermittent fasting diet, please make sure the 3 days you include are representative of your dietary cycle.
- During a food diary day, weigh **everything** that you eat/drink (including beverages and pills) and input them onto the Libro app. The app has food searching function. If you cannot find exactly what you are eating, find the closest match (for instance, the same microwave meal from a different brand), or more ideally, input each ingredient of the meal if you are cooking your own meal.
- You do not have to input pure water as we are not tracking hydration.
- When you are eating from a dish or a bowl, weigh the whole dish/bowl before you eat, and again after you eat, and by subtraction you can get the food weight. Same goes with weighing beverages, 1g of beverage roughly equals to 1ml.

### DNA Extraction SOP

**See Manufacturer Isolation Protocol for GFX-02 4ml GeneFix™ saliva sample** (2ml saliva collected into 2ml lysis buffer)

### Feature Description and Evidence Appraisal

#### Evidence Appraisal Criteria

Criteria were separated for genetic features vs. non-genetic features. Since we were only looking at genotypes without obtaining any information on gene expression, all the genetic evidence we gathered was essentially prospective evidence since genetic makeup is determined at birth. As a result, a separate set of criteria that do not distinguish between prospective and cross-sectional evidence was established for genetic features.

#### Genetic Features

- Class 1: Gene associated with the target SNP has theoretical connections with soft tissue injuries/musculoskeletal injuries (such as type I collagen), plus research has shown a direct link between the target SNP and common running-related injuries (not necessarily within a runner population, but the type of injury is also common in runners).
- Class 1: Fulfill 1 of the 2 conditions above (theoretical connection of associated gene; research showing direct link with RRI).

- Class 3: Fulfill none of the 2 conditions or has >90% overlap in genotypic distribution with another SNP within the study population. When an overlapping case was discovered, a subjective judgement was made to downgrade the one with relatively lower quality of evidence.

Non-genetic Features

- Class 1: There is prospective evidence showing a direct link between the feature and running-related injuries. The study must be conducted directly on runners.
- Class 2: There is cross-sectional/retrospective evidence showing a direct link between the feature and running-related injuries. Studies conducted with non-runner populations were included as long as the injury type was common among runners.
- Class 3: There is only circumferential evidence supporting the feature (such as opinion article), or the feature makes theoretical sense but does not have research evidence to support. If a feature is considered to be highly correlated with other features, subjective judgements were made to keep 1 of all correlated features and leave all the rest to class 3.

Below are some examples of class assignments of non-genetic features:

1. A feature with high quality prospective evidence on runners, but some results showing no correlation. Pooled results showed no correlation. For instance, pooled meta-analysis showed no difference in injury risk between sexes, but there were differences on specific injury types between sexes. Class 1
2. A feature that has no prospective evidence on runners, but has prospective evidence on other sports. Class 2
3. A feature that has no prospective evidence on runners, but has retrospective/cross-sectional evidence. Class 2
4. A feature that has no evidence on runners, but has evidence linking it to common running-related injury types (such as PFPS, plantar fasciitis). Class 2 or 3 depending on quality of evidence
5. A feature that has no original study to support directly, but could make sense via inference from original studies on similar topics (such as the same asymmetry study cited for all the asymmetry features). Class 3
6. A feature that has no original study to support, but appeared in an opinion article potentially due to some circumstantial evidence (such as most of the nutrient features). Class 3
7. A feature that has no study to support, but the reason for that was likely because the measurement method is complex, and researchers often use simpler replacement methods (such as thigh and calf FFMI vs. BMI). Class 3

**Baseline Questionnaire**

| Feature Name | Explanation | References | Evidence Appraisal |
|---|---|---|---|
| Sex | | Jacobsson, J., Timpka, T., Kowalski, J., Nilsson, S., Ekberg, J., Dahlström, Ö., & Renström, P. A. (2013). Injury patterns in Swedish elite athletics: annual incidence, injury types and risk factors. *British Journal of Sports Medicine*, *47*(15), 941-952. Rauh, M. J., Koepsell, T. D., Rivara, F. P., Margherita, A. J., & Rice, S. G. (2006). Epidemiology of musculoskeletal injuries among high school cross-country runners. *American journal of epidemiology*, *163*(2), 151-159. Messier, S. P., Martin, D. F., Mihalko, S. L., Ip, E., DeVita, P., Cannon, D. W., ... & Seay, J. F. (2018). A 2-year prospective cohort study of overuse running injuries: | Multiple prospective studies with conflicting results. Different types of injuries also seem to have different variations between sexes. The last article is a systematic review and meta-analysis that contains a comprehensive summarisation of |

| | | | |
|---|---|---|---|
| | | the runners and injury longitudinal study (TRAILS). *The American journal of sports medicine*, *46*(9), 2211-2221. Hollander, K., Rahlf, A. L., Wilke, J., Edler, C., Steib, S., Junge, A., & Zech, A. (2021). Sex-specific differences in running injuries: a systematic review with meta-analysis and meta-regression. *Sports Medicine*, *51*, 1011-1039. | existing evidence. |
| Age | Age as of the date of completing the baseline questionnaire | Taunton, J. E., Ryan, M. B., Clement, D. B., McKenzie, D. C., Lloyd-Smith, D. R., & Zumbo, B. D. (2003). A prospective study of running injuries: the Vancouver Sun Run “In Training” clinics. *British journal of sports medicine*, *37*(3), 239-244. Nielsen, R. O., Buist, I., Parner, E. T., Nohr, E. A., Sørensen, H., Lind, M., & Rasmussen, S. (2013). Predictors of running-related injuries among 930 novice runners: a 1-year prospective follow-up study. *Orthopaedic* | Prospective investigations showed some link (one only found in women, the other not statistically significant) |

| | | | |
|---|---|---|---|
| | | *journal of sports medicine*, *1*(1), 2325967113487316. Taunton, J. E., Ryan, M. B., Clement, D. B., McKenzie, D. C., Lloyd-Smith, D. R., & Zumbo, B. D. (2002). A retrospective case-control analysis of 2002 running injuries. *British journal of sports medicine*, *36*(2), 95-101. | |
| Past_stress_injury | Y/N on being diagnosed with stress injury in the past | Kelsey, J. L., Bachrach, L. K., Procter-Gray, E., Nieves, J. E. R. I., Greendale, G. A., Sowers, M., ... & Cobb, K. L. (2007). Risk factors for stress fracture among young female cross-country runners. *Medicine & Science in Sports & Exercise*, *39*(9), 1457-1463. Tenforde, A. S., Sayres, L. C., McCurdy, M. L., Sainani, K. L., & Fredericson, M. I. C. H. A. E. L. (2013). Identifying sex-specific risk factors for stress fractures in adolescent runners. *Medicine & Science in Sports & Exercise*, *45*(10), 1843-1851. | Prospective evidence showing link between previous stress frature and future stress fracture. However, as stress fracture only forms a fraction of all injuries, this feature is downgraded. |

| | | | |
|---|---|---|---|
| | | Wright, A. A., Taylor, J. B., Ford, K. R., Siska, L., & Smoliga, J. M. (2015). Risk factors associated with lower extremity stress fractures in runners: a systematic review with meta-analysis. *British Journal of Sports Medicine*, *49*(23), 1517-1523. | |
| lower_limb_days_total[1] | Total number of days affected by lower limb injuries during the past year | Buist, I., Bredeweg, S. W., Lemmink, K. A., Van Mechelen, W., & Diercks, R. L. (2010). Predictors of running-related injuries in novice runners enrolled in a systematic training program: a prospective cohort study. *The American journal of sports medicine*, *38*(2), 273-280.<br>Theisen, D., Malisoux, L., Genin, J., Delattre, N., Seil, R., & Urhausen, A. (2014). Influence of midsole hardness of standard cushioned shoes on running-related injury risk. *British Journal of Sports Medicine*, *48*(5), 371-376. | Ample evidence in prospective studies showing links between previous RRI and future RRI. |

| | | | |
|---|---|---|---|
| | | Saragiotto, B. T., Yamato, T. P., Hespanhol Junior, L. C., Rainbow, M. J., Davis, I. S., & Lopes, A. D. (2014). What are the main risk factors for running-related injuries?. *Sports medicine*, *44*, 1153-1163.<br>Hulme, A., Nielsen, R. O., Timpka, T., Verhagen, E., & Finch, C. (2017). Risk and protective factors for middle- and long-distance running-related injury. *Sports Medicine*, *47*, 869-886. | |
| Athlete_Score<br>[2] | Athlete score based on best performance reported (or gathered online) during the past year | Van Mechelen, W. (1992). Running injuries: a review of the epidemiological literature. *Sports medicine*, *14*, 320-335.<br>Fredette, A., Roy, J. S., Perreault, K., Dupuis, F., Napier, C., & Esculier, J. F. (2022). The association between running injuries and training parameters: a systematic review. *Journal of Athletic Training*, *57*(7), 650-671. | Comparison between studies show large differences among cohorts with different performance levels. However, it is suspected that this |

| | | | |
|---|---|---|---|
| | | | features correlates with training volume, thus it is downgraded. |
| average_run_hours | Average hours of running per week during the past year | Junior, L. C. H., Costa, L. O. P., & Lopes, A. D. (2013). Previous injuries and some training characteristics predict running-related injuries in recreational runners: a prospective cohort study. *Journal of Physiotherapy*, *59*(4), 263-269.<br>Kluitenberg, B., van der Worp, H., Huisstede, B. M., Hartgens, F., Diercks, R., Verhagen, E., & van Middelkoop, M. (2016). The NLstart2run study: Training-related factors associated with running-related injuries in novice runners. *Journal of Science and Medicine in Sport*, *19*(8), 642-646.<br>Fredette, A., Roy, J. S., Perreault, K., Dupuis, F., Napier, | Prospective tracking studies show conflicting evidence. Most show no significant correlation between duration and injuries, some show significant correlations. |

| | | | |
|---|---|---|---|
| | | C., & Esculier, J. F. (2022). The association between running injuries and training parameters: a systematic review. *Journal of Athletic Training*, *57*(7), 650-671. | |
| average_run_frequency | Average number of running sessions per week during the past year | Taunton, J. E., Ryan, M. B., Clement, D. B., McKenzie, D. C., Lloyd-Smith, D. R., & Zumbo, B. D. (2003). A prospective study of running injuries: the Vancouver Sun Run "In Training" clinics. *British journal of sports medicine*, *37*(3), 239-244.<br>Malisoux, L., Ramesh, J., Mann, R., Seil, R., Urhausen, A., & Theisen, D. (2015). Can parallel use of different running shoes decrease running-related injury risk?. *Scandinavian journal of medicine & science in sports*, *25*(1), 110-115.<br>Fredette, A., Roy, J. S., Perreault, K., Dupuis, F., Napier, C., & Esculier, J. F. (2022). The association between running injuries and training parameters: a systematic | Prospective evidence show conflicting results, however this feature is correlated with average_run_hours. |

| | | | |
|---|---|---|---|
| | | review. *Journal of Athletic Training*, *57*(7), 650-671. | |
| average_interval_training_frequency | Average number of interval training sessions per week during the past year | Junior, L. C. H., Costa, L. O. P., & Lopes, A. D. (2013). Previous injuries and some training characteristics predict running-related injuries in recreational runners: a prospective cohort study. *Journal of Physiotherapy*, *59*(4), 263-269. | Prospective evidence shows correlation. |
| EDEQ_total | EDEQ score | Rauh, M. J., Barrack, M., & Nichols, J. F. (2014). Associations between the female athlete triad and injury among high school runners. *International journal of sports physical therapy*, *9*(7), 948.<br>Rauh, M. J., Nichols, J. F., & Barrack, M. T. (2010). Relationships among injury and disordered eating, menstrual dysfunction, and low bone mineral density in high school athletes: a prospective study. *Journal of athletic training*, *45*(3), 243-252. | Prospective studies showed association between EDEQ overall and some subscale scores and RRIs. |

| | | | |
|---|---|---|---|
| | | Hamstra-Wright, K. L., Bliven, K. C. H., Coumbe-Lilley, J. E., Djelovic, E., & Patel, J. (2023). The relationship between eating disorders, disordered eating, and injury in athletes: a critically appraised topic. *Journal of Sport Rehabilitation*, *32*(4), 474-481. | |
| LEAF-Q[3] | LEAF-Q score | Barrack, M. T., Gibbs, J. C., De Souza, M. J., Williams, N. I., Nichols, J. F., Rauh, M. J., & Nattiv, A. (2014). Higher incidence of bone stress injuries with increasing female athlete triad–related risk factors: a prospective multisite study of exercising girls and women. *The American journal of sports medicine*, *42*(4), 949-958. Holtzman, B., Popp, K. L., Tenforde, A. S., Parziale, A. L., Taylor, K., & Ackerman, K. E. (2022). Low energy availability surrogates associated with lower bone mineral density and bone stress injury site. *PM&R*, *14*(5), 587-596. | Prospective studies showed association between LEAF-Q and stress fratures in females. However, stress fratures only make up a fraction of all injuires, and females make up less than 1/2 of the cohort. |

| | | |
|---|---|---|
| | | Hamstra-Wright, K. L., Bliven, K. C. H., Coumbe-Lilley, J. E., Djelovic, E., & Patel, J. (2023). The relationship between eating disorders, disordered eating, and injury in athletes: a critically appraised topic. *Journal of Sport Rehabilitation*, *32*(4), 474-481. |

### Isokinetic Strength

| Feature Name | Explanation | References | Evidence Appraisal |
|---|---|---|---|
| Hip_abduction_peak_torque | The average peak torque of hip abduction for the top 3-7 reps (total of 4*5 reps, taking away 13 familiarisation and 2 potential compensation) | Becker, J. A. M. E. S., Nakajima, M. I. M. I., & Wu, W. F. (2018). Factors Contributing to Medial Tibial Stress Syndrome in Runners: A Prospective Study. *Medicine and science in sports and exercise*, *50*(10), 2092-2100. Luedke, L. E., Heiderscheit, B. C., Williams, D. B., & Rauh, M. J. (2015). Association of isometric strength of | Prospective studies show correlations between hip abductor strength and RRIs. |

| | | |
|---|---|---|
| | | hip and knee muscles with injury risk in high school cross country runners. *International journal of sports physical therapy*, *10*(6), 868.<br>Finnoff, J. T., Hall, M. M., Kyle, K., Krause, D. A., Lai, J., & Smith, J. (2011). Hip strength and knee pain in high school runners: a prospective study. *PM&R*, *3*(9), 792-801.<br>Mucha, M. D., Caldwell, W., Schlueter, E. L., Walters, C., & Hassen, A. (2017). Hip abductor strength and lower extremity running related injury in distance runners: a systematic review. *Journal of science and medicine in sport*, *20*(4), 349-355.<br>Christopher, S. M., McCullough, J., Snodgrass, S. J., & Cook, C. (2019). Do alterations in muscle strength, flexibility, range of motion, and alignment predict lower extremity injury in runners: a systematic review. *Archives* |

| | | | |
|---|---|---|---|
| | | *of Physiotherapy*, *9*, 1-14. de Marche Baldon, R., Nakagawa, T. H., Muniz, T. B., Amorim, C. F., Maciel, C. D., & Serrão, F. V. (2009). Eccentric hip muscle function in females with and without patellofemoral pain syndrome. *Journal of athletic training*, *44*(5), 490-496. Neal, B. S., Lack, S. D., Lankhorst, N. E., Raye, A., Morrissey, D., & Van Middelkoop, M. (2019). Risk factors for patellofemoral pain: a systematic review and meta-analysis. *British Journal of Sports Medicine*, *53*(5), 270-281. | |
| Hip_abduction_peak_angle | The average joint angle at peak torque of the 5 reps used above, measured as the angle of the isokinetic rotor arm | Brughelli, M., Cronin, J., & Nosaka, K. (2010). Muscle architecture and optimum angle of the knee flexors and extensors: a comparison between cyclists and Australian Rules football players. *The Journal of Strength &* | Mostly speculative. |

| | | | |
|---|---|---|---|
| | | *Conditioning Research*, *24*(3), 717-721. | |
| hip_abduction_peak_torque_asymmetry | The absolute value of the difference between the left and right hip abduction peak torque divided by their total value | De Blaiser, C., Roosen, P., Willems, T., De Bleecker, C., Vermeulen, S., Danneels, L., & De Ridder, R. (2021). The role of core stability in the development of non-contact acute lower extremity injuries in an athletic population: A prospective study. *Physical Therapy in Sport*, *47*, 165-172.<br>Hietamo, J., Pasanen, K., Leppänen, M., Steffen, K., Kannus, P., Heinonen, A., ... & Parkkari, J. (2021). Association between lower extremity muscle strength and acute ankle injury in youth team-sports athletes. *Physical Therapy in Sport*, *48*, 188-195.<br>Niemuth, P. E., Johnson, R. J., Myers, M. J., & Thieman, T. J. (2005). Hip muscle weakness and overuse injuries in recreational runners. *Clinical Journal of Sport Medicine*, *15*(1), 14-21. | Prospective studies show correlations in sporting populations but not runners specifically. |

| | | | |
|---|---|---|---|
| | | Guan, Y., Bredin, S. S., Taunton, J., Jiang, Q., Wu, N., & Warburton, D. E. (2022). Association between inter-limb asymmetries in lower-limb functional performance and sport injury: a systematic review of prospective cohort studies. *Journal of clinical medicine*, *11*(2), 360. | |
| hip_abduction_peak_angle_asymmetry | The difference between the left and right hip abduction peak angle, NOT normalised by the mean | | Speculative. |
| Hip_adduction_peak_torque | | de Marche Baldon, R., Nakagawa, T. H., Muniz, T. B., Amorim, C. F., Maciel, C. D., & Serrão, F. V. (2009). Eccentric hip muscle function in females with and without patellofemoral pain syndrome. *Journal of athletic training*, *44*(5), 490-496. | Cross-sectional evidence links hip adduction strength to PFPS which is a common RRI. |
| Hip_adduction_peak_angle | | | Speculative |

| | | | |
|---|---|---|---|
| hip_adduction_peak_torque_asymmetry | | Niemuth, P. E., Johnson, R. J., Myers, M. J., & Thieman, T. J. (2005). Hip muscle weakness and overuse injuries in recreational runners. *Clinical Journal of Sport Medicine*, *15*(1), 14-21.<br>Guan, Y., Bredin, S. S., Taunton, J., Jiang, Q., Wu, N., & Warburton, D. E. (2022). Association between inter-limb asymmetries in lower-limb functional performance and sport injury: a systematic review of prospective cohort studies. *Journal of clinical medicine*, *11*(2), 360. | Cross-sectional study shows injured runners had greater asymmetry in isometric hip adduction strength. |
| hip_adduction_peak_angle_asymmetry | | | Speculative |
| Total_ad_ab_ratio | The ratio between the left and right hip adduction and abduction average peak torque | Jungmalm, J., Nielsen, R. Ø., Desai, P., Karlsson, J., Hein, T., & Grau, S. (2020). Associations between biomechanical and clinical/anthropometrical factors and | Prospective studies shows link with RRIs. |

| | | | |
|---|---|---|---|
| | | running-related injuries among recreational runners: a 52-week prospective cohort study. *Injury epidemiology*, *7*, 1-9. Finnoff, J. T., Hall, M. M., Kyle, K., Krause, D. A., Lai, J., & Smith, J. (2011). Hip strength and knee pain in high school runners: a prospective study. *PM&R*, *3*(9), 792-801. Ferber, R., Hreljac, A., & Kendall, K. D. (2009). Suspected mechanisms in the cause of overuse running injuries: a clinical review. *Sports health*, *1*(3), 242-246. | |
| Ad_ab_ratio_asymmetry | The difference between the left and right adduction and abduction average peak torque ratio | | Speculative. |
| Knee_extension_peak_torque | | Luedke, L. E., Heiderscheit, B. C., Williams, D. B., & Rauh, M. J. (2015). Association of isometric strength of hip and knee muscles with injury risk in high school cross country runners. *International journal of sports physical* | Prospective study shows correlation, pooled analysis from a meta-analysis also |

| | | | |
|---|---|---|---|
| | | *therapy*, *10*(6), 868.<br>Peterson, B., Hawke, F., Spink, M., Sadler, S., Hawes, M., Callister, R., & Chuter, V. (2022). Biomechanical and musculoskeletal measurements as risk factors for running-related injury in non-elite runners: A systematic review and meta-analysis of prospective studies. *Sports medicine-open*, *8*(1), 38.<br>McGuire, B., & King, B. (2021). Neuromuscular risk factors for non-contact knee injury: a systematic review and meta-analysis. *medRxiv*, 2021-09.<br>Neal, B. S., Lack, S. D., Lankhorst, N. E., Raye, A., Morrissey, D., & Van Middelkoop, M. (2019). Risk factors for patellofemoral pain: a systematic review and meta-analysis. *British Journal of Sports Medicine*, *53*(5), 270-281. | shows significant correlation despite each included study not significant. |

| | | | |
|---|---|---|---|
| Knee_extension_peak_angle | | | Speculative. |
| Knee_extension_peak_torque_asymmetry | | Fousekis, K., Tsepis, E., Poulmedis, P., Athanasopoulos, S., & Vagenas, G. (2011). Intrinsic risk factors of non-contact quadriceps and hamstring strains in soccer: a prospective study of 100 professional players. *British journal of sports medicine*, *45*(9), 709-714. Guan, Y., Bredin, S. S., Taunton, J., Jiang, Q., Wu, N., & Warburton, D. E. (2022). Association between inter-limb asymmetries in lower-limb functional performance and sport injury: a systematic review of prospective cohort studies. *Journal of clinical medicine*, *11*(2), 360. | Prospective study shows association within soccer players. |
| Knee_extension_peak_angl | | | Speculative. |

| | | | |
|---|---|---|---|
| e_asymmetry | | | |
| Knee_flexion_peak_torque | | Luedke, L. E., Heiderscheit, B. C., Williams, D. B., & Rauh, M. J. (2015). Association of isometric strength of hip and knee muscles with injury risk in high school cross country runners. *International journal of sports physical therapy*, *10*(6), 868.<br>Hein, T., Janssen, P., Wagner-Fritz, U., Haupt, G., & Grau, S. (2014). Prospective analysis of intrinsic and extrinsic risk factors on the development of achilles tendon pain in runners. *Scandinavian Journal of Medicine & Science in Sports*, *24*(3), e201-e212. | Prospective studies show association with RRIs and achilles tendon pain. |
| Knee_flexion_peak_angle | | Timmins, R. G., Shield, A. J., Williams, M. D., & Opar, D. A. (2016). Is there evidence to support the use of the angle of peak torque as a marker of hamstring injury and re-injury risk?. *Sports Medicine*, *46*, 7-13. | Some theoretical basis with sprinters but not robust. |

| | | | |
|---|---|---|---|
| Knee_flexion_peak_torque_asymmetry | | Knapik, J. J., Bauman, C. L., Jones, B. H., Harris, J. M., & Vaughan, L. (1991). Preseason strength and flexibility imbalances associated with athletic injuries in female collegiate athletes. *The American journal of sports medicine*, *19*(1), 76-81.<br>Fousekis, K., Tsepis, E., Poulmedis, P., Athanasopoulos, S., & Vagenas, G. (2011). Intrinsic risk factors of non-contact quadriceps and hamstring strains in soccer: a prospective study of 100 professional players. *British journal of sports medicine*, *45*(9), 709-714.<br>Grazioli, R., Sobieski, N., Wilhelm, E. N., Brusco, C. M., & Rech, A. (2022). Divergent isokinetic muscle strength deficits in street running athletes. *Sport Sciences for Health*, 1-8.<br>Guan, Y., Bredin, S. S., Taunton, J., Jiang, Q., Wu, N., & | Prospective evidence present in general sporting population and soccer players. |

| | | | |
|---|---|---|---|
| | | Warburton, D. E. (2022). Association between inter-limb asymmetries in lower-limb functional performance and sport injury: a systematic review of prospective cohort studies. *Journal of clinical medicine*, *11*(2), 360. | |
| Knee_flexion_peak_angle_asymmetry | | | Speculative |
| Total_fl_ex_ratio | | Knapik, J. J., Bauman, C. L., Jones, B. H., Harris, J. M., & Vaughan, L. (1991). Preseason strength and flexibility imbalances associated with athletic injuries in female collegiate athletes. *The American journal of sports medicine*, *19*(1), 76-81.<br>Padasala, M., Joksimovic, M., Bruno, C., Melino, D., & Manzi, V. (2020). Muscle injuries in athletes. The relationship between H/Q ratio (hamstring/quadriceps | Prospective evidence present in general sporting population. |

| | | | |
|---|---|---|---|
| | | ratio). *Ita J Sports Reh Po*, *7*(1), 1478-1498. McGuire, B., & King, B. (2021). Neuromuscular risk factors for non-contact knee injury: a systematic review and meta-analysis. *medRxiv*, 2021-09. | |
| Fl_ex_ratio_asymmetry | | | Speculative. |

### Biomechanics

| Feature Name | Explanation | References | Evidence Appraisal |
|---|---|---|---|
| Navicular_drop | The average navicular drop between left and right | Buist, I., Bredeweg, S. W., Lemmink, K. A., Van Mechelen, W., & Diercks, R. L. (2010). Predictors of running-related injuries in novice runners enrolled in a systematic training program: a prospective cohort study. *The American journal of sports medicine*, *38*(2), | Multiple prospective studies showing correlation with RRIs. |

| | | |
|---|---|---|
| | | 273-280.<br><br>Bennett, J. E., Reinking, M. F., & Rauh, M. J. (2012). The relationship between isotonic plantar flexor endurance, navicular drop, and exercise-related leg pain in a cohort of collegiate cross-country runners. *International journal of sports physical therapy*, *7*(3), 267.<br><br>Raissi, G. R. D., Cherati, A. D. S., Mansoori, K. D., & Razi, M. D. (2009). The relationship between lower extremity alignment and Medial Tibial Stress Syndrome among non-professional athletes. *BMC Sports Science, Medicine and Rehabilitation*, *1*, 1-8.<br><br>Zifchock, R. A., Davis, I., Higginson, J., McCaw, S., & Royer, T. (2008). Side-to-side differences in overuse running injury susceptibility: a retrospective study. *Human movement science*, *27*(6), 888-902. |

| | | | |
|---|---|---|---|
| | | Christopher, S. M., McCullough, J., Snodgrass, S. J., & Cook, C. (2019). Do alterations in muscle strength, flexibility, range of motion, and alignment predict lower extremity injury in runners: a systematic review. *Archives of Physiotherapy*, *9*, 1-14. | |
| Navicular_drop_asymmetry | The absolute value of the difference between left and right navicular drop | Raissi, G. R. D., Cherati, A. D. S., Mansoori, K. D., & Razi, M. D. (2009). The relationship between lower extremity alignment and Medial Tibial Stress Syndrome among non-professional athletes. *BMC Sports Science, Medicine and Rehabilitation*, *1*, 1-8. | Prospective study shows runners sustaining MTSS have higher navicular drop asymmetry. |
| Q_angle | The average Q angle between left and right | Rauh, M. J., Koepsell, T. D., Rivara, F. P., Rice, S. G., & Margherita, A. J. (2007). Quadriceps angle and risk of injury among high school cross-country runners. *Journal of Orthopaedic & Sports Physical Therapy*, *37*(12), 725-733. Puckree, T., Govender, A., Govender, K., & Naidoo, P. | Prospective studies show association with RRIs. |

| | | | |
|---|---|---|---|
| | | (2007). The quadriceps angle and the incidence of knee injury in Indian long-distance runners. *South African Journal of Sports Medicine*, *19*(1), 9-11.<br>Ellapen, T. J., Satyendra, S., Morris, J., & Van Heerden, H. J. (2013). Common running musculoskeletal injuries among recreational half-marathon runners in KwaZulu-Natal. *South African Journal of Sports Medicine*, *25*(2), 39-43. | |
| Q_angle_asymmetry | The absolute value of the difference between left and right Q angle | Rauh, M. J., Koepsell, T. D., Rivara, F. P., Rice, S. G., & Margherita, A. J. (2007). Quadriceps angle and risk of injury among high school cross-country runners. *Journal of Orthopaedic & Sports Physical Therapy*, *37*(12), 725-733. | Prospective study shows association with RRIs. |
| VILR_10[4] | Vertical impact loading rate during the 10km/h run | Davis, I. S., Bowser, B. J., & Mullineaux, D. R. (2016). Greater vertical impact loading in female runners with medically diagnosed injuries: a prospective | Ample prospective evidence on runners. However since |

| | | | |
|---|---|---|---|
| | | investigation. *British journal of sports medicine*, *50*(14), 887-892.<br>Johnson, C. D., Tenforde, A. S., Outerleys, J., Reilly, J., & Davis, I. S. (2020). Impact-related ground reaction forces are more strongly associated with some running injuries than others. *The American journal of sports medicine*, *48*(12), 3072-3080.<br>Bredeweg, S. W., Kluitenberg, B., Bessem, B., & Buist, I. (2013). Differences in kinetic variables between injured and noninjured novice runners: a prospective cohort study. *Journal of Science and Medicine in Sport*, *16*(3), 205-210. | features under 10km/h highly correlate with features under 12km/h, they are downgraded. |
| VALR_10[4] | Vertical average loading rate during the 10km/h run | Davis, I. S., Bowser, B. J., & Mullineaux, D. R. (2016). Greater vertical impact loading in female runners with medically diagnosed injuries: a prospective | Same as above. |

| | | | |
|---|---|---|---|
| | | investigation. *British journal of sports medicine*, *50*(14), 887-892.<br>Johnson, C. D., Tenforde, A. S., Outerleys, J., Reilly, J., & Davis, I. S. (2020). Impact-related ground reaction forces are more strongly associated with some running injuries than others. *The American journal of sports medicine*, *48*(12), 3072-3080.<br>Bredeweg, S. W., Kluitenberg, B., Bessem, B., & Buist, I. (2013). Differences in kinetic variables between injured and noninjured novice runners: a prospective cohort study. *Journal of Science and Medicine in Sport*, *16*(3), 205-210. | |
| VILR_asymmetry_10 | The absolute value of the difference between alternating steps on VILR normalised by the mean | | Speculative. |

| | | | |
|---|---|---|---|
| VALR_asymmetry_10 | The absolute value of the difference between alternating steps on VALR normalised by the mean | | Speculative. |
| Impact_peak_10[5] | The average value of peak force experienced during each step during the 10km/h run | Davis, I. S., Bowser, B. J., & Mullineaux, D. R. (2016). Greater vertical impact loading in female runners with medically diagnosed injuries: a prospective investigation. *British journal of sports medicine*, *50*(14), 887-892. | Downgraded due to high correlation with feature under 12km/h. |
| Impact_peak_asymmetry_10 | The absolute value of the difference between alternating steps on average value of peak force for the 10km/h run, normalised by the mean | | Speculative |
| Flight_time_10[5] | Average amount of time during each step when the participant is not in contact with the ground for the | Malisoux, L., Gette, P., Delattre, N., Urhausen, A., & Theisen, D. (2022). Spatiotemporal and ground-reaction force characteristics as risk factors for running-related | Downgraded due to high correlation with feature under 12km/h. |

| | | | |
|---|---|---|---|
| | 10km/h run. | injury: A secondary analysis of a randomized trial including 800+ recreational runners. *The American Journal of Sports Medicine*, *50*(2), 537-544.<br>Winter, S. C., Gordon, S., Brice, S. M., Lindsay, D., & Barrs, S. (2020). A multifactorial approach to overuse running injuries: a 1-year prospective study. *Sports Health*, *12*(3), 296-303. | |
| Contact_time_10[5] | Average amount of time during each step when the participant is in contact with the ground for the 10km/h run. | Malisoux, L., Gette, P., Delattre, N., Urhausen, A., & Theisen, D. (2022). Spatiotemporal and ground-reaction force characteristics as risk factors for running-related injury: A secondary analysis of a randomized trial including 800+ recreational runners. *The American Journal of Sports Medicine*, *50*(2), 537-544. | Downgraded due to high correlation with feature under 12km/h. |
| Duty_factor_10 | Ratio between contact time and stride time (flight time + contact time) for | Malisoux, L., Gette, P., Delattre, N., Urhausen, A., & Theisen, D. (2022). Spatiotemporal and ground-reaction | Downgraded due to high correlation with |

| | | | |
|---|---|---|---|
| | the 10km/h run. | force characteristics as risk factors for running-related injury: A secondary analysis of a randomized trial including 800+ recreational runners. *The American Journal of Sports Medicine*, *50*(2), 537-544. | feature under 12km/h. |
| Step_frequency_10 | Number of steps per minutes during the 10km/h run | Winter, S. C., Gordon, S., Brice, S. M., Lindsay, D., & Barrs, S. (2020). A multifactorial approach to overuse running injuries: a 1-year prospective study. *Sports Health*, *12*(3), 296-303. Anderson, L. M., Martin, J. F., Barton, C. J., & Bonanno, D. R. (2022). What is the effect of changing running step rate on injury, performance and biomechanics? A systematic review and meta-analysis. *Sports Medicine-Open*, *8*(1), 112. | Downgraded due to high correlation with feature under 12km/h. |
| Cadence_asymmetry_10 | The absolute value of the difference between alternating steps on average | | Speculative. |

| | | | |
|---|---|---|---|
| | time taken for each step for the 10km/h run, normalised by the mean. | | |
| Duty_factor_asymmetry_10 | The absolute value of the difference between the duty factors of alternating steps for the 10km/h run, normalised by the mean. | | Speculative. |
| VILR_12 | | Davis, I. S., Bowser, B. J., & Mullineaux, D. R. (2016). Greater vertical impact loading in female runners with medically diagnosed injuries: a prospective investigation. *British journal of sports medicine*, *50*(14), 887-892.<br>Johnson, C. D., Tenforde, A. S., Outerleys, J., Reilly, J., & Davis, I. S. (2020). Impact-related ground reaction forces are more strongly associated with some running injuries than others. *The American journal of sports* | Prospective studies link VILR with RRIs, however since VILR correlates highly with VALR, it is downgraded. |

| | | | |
|---|---|---|---|
| | | *medicine*, *48*(12), 3072-3080.<br>Bredeweg, S. W., Kluitenberg, B., Bessem, B., & Buist, I. (2013). Differences in kinetic variables between injured and noninjured novice runners: a prospective cohort study. *Journal of Science and Medicine in Sport*, *16*(3), 205-210. | |
| VALR_12 | | Davis, I. S., Bowser, B. J., & Mullineaux, D. R. (2016). Greater vertical impact loading in female runners with medically diagnosed injuries: a prospective investigation. *British journal of sports medicine*, *50*(14), 887-892.<br>Johnson, C. D., Tenforde, A. S., Outerleys, J., Reilly, J., & Davis, I. S. (2020). Impact-related ground reaction forces are more strongly associated with some running injuries than others. *The American journal of sports* | VALR is chosen over VILR because within the prospective studies VALR present with higher effect sizes when both variables show significant correlation with RRIs. |

| | | | |
|---|---|---|---|
| | | *medicine*, *48*(12), 3072-3080. Bredeweg, S. W., Kluitenberg, B., Bessem, B., & Buist, I. (2013). Differences in kinetic variables between injured and noninjured novice runners: a prospective cohort study. *Journal of Science and Medicine in Sport*, *16*(3), 205-210. | |
| VILR_asymmetry_12 | | | Speculative. |
| VALR_asymmetry_12 | | | Speculative. |
| Impact_peak_12 | | Davis, I. S., Bowser, B. J., & Mullineaux, D. R. (2016). Greater vertical impact loading in female runners with medically diagnosed injuries: a prospective investigation. *British journal of sports medicine*, *50*(14), 887-892. | Prospective study shows correlations with RRIs. |

| | | | |
|---|---|---|---|
| Impact_peak_asymmetry_12 | | | Speculative. |
| Flight_time_12 | Average amount of time during each step when the participant is not in contact with the ground for the 12km/h run. | Malisoux, L., Gette, P., Delattre, N., Urhausen, A., & Theisen, D. (2022). Spatiotemporal and ground-reaction force characteristics as risk factors for running-related injury: A secondary analysis of a randomized trial including 800+ recreational runners. *The American Journal of Sports Medicine*, *50*(2), 537-544.<br>Winter, S. C., Gordon, S., Brice, S. M., Lindsay, D., & Barrs, S. (2020). A multifactorial approach to overuse running injuries: a 1-year prospective study. *Sports Health*, *12*(3), 296-303. | Downgraded due to high correlation with duty factor. |
| Contact_time_12 | Average amount of time during each step when the participant is in contact | Malisoux, L., Gette, P., Delattre, N., Urhausen, A., & Theisen, D. (2022). Spatiotemporal and ground-reaction | Downgraded due to high correlation with |

| | | | |
|---|---|---|---|
| | with the ground for the 12km/h run. | force characteristics as risk factors for running-related injury: A secondary analysis of a randomized trial including 800+ recreational runners. *The American Journal of Sports Medicine*, *50*(2), 537-544. | duty factor. |
| Duty_factor_12 | Ratio between contact time and stride time (flight time + contact time) for the 12km/h run. | Malisoux, L., Gette, P., Delattre, N., Urhausen, A., & Theisen, D. (2022). Spatiotemporal and ground-reaction force characteristics as risk factors for running-related injury: A secondary analysis of a randomized trial including 800+ recreational runners. *The American Journal of Sports Medicine*, *50*(2), 537-544. | Prospective study shows correlation with larger efffect sizes than flight time and contact time. |
| Step_frequency_12 | Number of steps per minutes during the 12km/h run | Winter, S. C., Gordon, S., Brice, S. M., Lindsay, D., & Barrs, S. (2020). A multifactorial approach to overuse running injuries: a 1-year prospective study. *Sports Health*, *12*(3), 296-303.<br>Anderson, L. M., Martin, J. F., Barton, C. J., & Bonanno, | Downgraded due to high correlation with flight time, contact time, and duty factor. |

| | | | |
|---|---|---|---|
| | | D. R. (2022). What is the effect of changing running step rate on injury, performance and biomechanics? A systematic review and meta-analysis. *Sports Medicine-Open*, *8*(1), 112. | |
| Cadence_asymmetry_12 | The absolute value of the difference between alternating steps on average time taken for each step for the 12km/h run, normalised by the mean | | Speculative. |
| Duty_factor_asymmetry_12 | The absolute value of the difference between the duty factors of alternating steps for the 12km/h run, normalised by the mean.. | | Speculative. |
| Alt_strike[6] | Alternating striker, meaning that for the most part of either the 10km/h or the 12km/h trial, the participant shows | | Speculative. |

| | |
|---|---|
| | a rearfoot strike pattern with one leg and a forefoot strike pattern with the other leg |

### Nutrition

| Feature Name | Explanation | References | Evidence Appraisal |
|---|---|---|---|
| Fat_intake_avg[7] | Average fat intake per day in grams | Gerlach, K. E., Burton, H. W., Dorn, J. M., Leddy, J. J., & Horvath, P. J. (2008). Fat intake and injury in female runners. *Journal of the International society of sports nutrition*, *5*, 1-8. | Prospective study shows link between fat intake and RRIs. |
| Fat_intake_BW[7] [9] | Average fat intake per day in grams / lean body mass | | Speculative. |
| Fat_percentage_avg | Percentage of calories obtained from fat relative to total energy intake | Gerlach, K. E., Burton, H. W., Dorn, J. M., Leddy, J. J., & Horvath, P. J. (2008). Fat intake and injury in female | Downgraded due to high correlation with |

| | | | |
|---|---|---|---|
| | | runners. *Journal of the International society of sports nutrition*, *5*, 1-8. | fat intake. |
| Average_energy_availability[8] | (Caloric intake - exercise energy expenditure + resting metabolic rate during exercising hours) / lean body mass, averaged over each 3-day food diary | Heikura, I. A., Uusitalo, A. L., Stellingwerff, T., Bergland, D., Mero, A. A., & Burke, L. M. (2018). Low energy availability is difficult to assess but outcomes have large impact on bone injury rates in elite distance athletes. *International journal of sport nutrition and exercise metabolism*, *28*(4), 403-411. Gerlach, K. E., Burton, H. W., Dorn, J. M., Leddy, J. J., & Horvath, P. J. (2008). Fat intake and injury in female runners. *Journal of the International society of sports nutrition*, *5*, 1-8. Edama, M., Inaba, H., Hoshino, F., Natsui, S., Maruyama, S., & Omori, G. (2021). The relationship between the female athlete triad and injury rates in collegiate female | Prospective evidence seems to show LEAF-Q to be a better predictor than energy availability measured via food diary in predicting stress fracutres. Lack of evidence for EA's direct correlation with sports injuries other than BSI. |

| | | | |
|---|---|---|---|
| | | athletes. *PeerJ*, *9*, e11092.<br><br>Close, G. L., Sale, C., Baar, K., & Bermon, S. (2019). Nutrition for the prevention and treatment of injuries in track and field athletes. *International journal of sport nutrition and exercise metabolism*, *29*(2), 189-197.<br><br>Griffin, K. L., Knight, K. B., Bass, M. A., & Valliant, M. W. (2021). Predisposing risk factors for stress fractures in collegiate cross-country runners. *The Journal of Strength & Conditioning Research*, *35*(1), 227-232.<br><br>Curtis, L. (2016). Nutritional research may be useful in treating tendon injuries. *Nutrition*, *32*(6), 617-619. | |
| Protein_intake_BW | Average daily protein intake in g/kg lean body mass | Close, G. L., Sale, C., Baar, K., & Bermon, S. (2019). Nutrition for the prevention and treatment of injuries in track and field athletes. *International journal of sport nutrition and exercise metabolism*, *29*(2), 189-197. | Speculative. |

| | | | |
|---|---|---|---|
| | | Curtis, L. (2016). Nutritional research may be useful in treating tendon injuries. *Nutrition*, *32*(6), 617-619. | |
| Omega3_intake_BW | Average daily omega-3 intake in g/kg lean body mass | Close, G. L., Sale, C., Baar, K., & Bermon, S. (2019). Nutrition for the prevention and treatment of injuries in track and field athletes. *International journal of sport nutrition and exercise metabolism*, *29*(2), 189-197. | Speculative. |
| vitaminD_intake_BW | Average daily vitamin D intake in in ug/kg lean body mass | Close, G. L., Sale, C., Baar, K., & Bermon, S. (2019). Nutrition for the prevention and treatment of injuries in track and field athletes. *International journal of sport nutrition and exercise metabolism*, *29*(2), 189-197.<br>Griffin, K. L., Knight, K. B., Bass, M. A., & Valliant, M. W. (2021). Predisposing risk factors for stress fractures in collegiate<br>Curtis, L. (2016). Nutritional research may be useful in treating tendon injuries. *Nutrition*, *32*(6), 617-619.cross- | Speculative. |

| | | | |
|---|---|---|---|
| | | country runners. *The Journal of Strength & Conditioning Research*, *35*(1), 227-232. | |
| vitaminC_intake_BW | Average daily vitamin C intake in in mg/kg lean body mass | Close, G. L., Sale, C., Baar, K., & Bermon, S. (2019). Nutrition for the prevention and treatment of injuries in track and field athletes. *International journal of sport nutrition and exercise metabolism*, *29*(2), 189-197. Curtis, L. (2016). Nutritional research may be useful in treating tendon injuries. *Nutrition*, *32*(6), 617-619. | Speculative. |
| vitaminE_intake_BW | Average daily vitamin E intake in in mg/kg lean body mass | Close, G. L., Sale, C., Baar, K., & Bermon, S. (2019). Nutrition for the prevention and treatment of injuries in track and field athletes. *International journal of sport nutrition and exercise metabolism*, *29*(2), 189-197. | Speculative. |
| Calcium_intake_BW | Average daily calcium intake in in mg/kg lean body mass | Close, G. L., Sale, C., Baar, K., & Bermon, S. (2019). Nutrition for the prevention and treatment of injuries in track and field athletes. *International journal of sport* | Speculative. |

| | | | |
|---|---|---|---|
| | | *nutrition and exercise metabolism*, *29*(2), 189-197. Griffin, K. L., Knight, K. B., Bass, M. A., & Valliant, M. W. (2021). Predisposing risk factors for stress fractures in collegiate cross-country runners. *The Journal of Strength & Conditioning Research*, *35*(1), 227-232. | |
| copper_intake_BW | Average daily copper intake in in mg/kg lean body mass | Close, G. L., Sale, C., Baar, K., & Bermon, S. (2019). Nutrition for the prevention and treatment of injuries in track and field athletes. *International journal of sport nutrition and exercise metabolism*, *29*(2), 189-197. | Speculative. |
| iron_intake_BW | Average daily iron intake in in mg/kg lean body mass | Close, G. L., Sale, C., Baar, K., & Bermon, S. (2019). Nutrition for the prevention and treatment of injuries in track and field athletes. *International journal of sport nutrition and exercise metabolism*, *29*(2), 189-197. | Speculative. |
| Glycine_intake_BW | Average daily glycine intake in in g/kg lean body mass | Close, G. L., Sale, C., Baar, K., & Bermon, S. (2019). Nutrition for the prevention and treatment of injuries in | Speculative. |

| | | | |
|---|---|---|---|
| | | track and field athletes. *International journal of sport nutrition and exercise metabolism*, *29*(2), 189-197. | |
| arginine_intake_BW | Average daily arginine intake in in g/kg lean body mass | Close, G. L., Sale, C., Baar, K., & Bermon, S. (2019). Nutrition for the prevention and treatment of injuries in track and field athletes. *International journal of sport nutrition and exercise metabolism*, *29*(2), 189-197. Curtis, L. (2016). Nutritional research may be useful in treating tendon injuries. *Nutrition*, *32*(6), 617-619. | Speculative. |

**Bone Scans**[9]

| Feature Name | Explanation | References | Evidence Appraisal |
|---|---|---|---|
| height | Average of height measured at all scans | Duffey, M. J., Martin, D. F., Cannon, D. W., Craven, T., & Messier, S. P. (2000). Etiologic factors associated with anterior knee pain in distance runners. *Medicine and science* | Has prospective evidence but downgraded due to |

| | | | |
|---|---|---|---|
| | | *in sports and exercise*, *32*(11), 1825-1832. | high correlation with BMI. |
| Mass[9] | | Shiotani, H., Mizokuchi, T., Yamashita, R., Naito, M., & Kawakami, Y. (2023). Influence of Body Mass on Running-Induced Changes in Mechanical Properties of Plantar Fascia. *The Journal of Strength & Conditioning Research*, *37*(11), e588-e592. | Downgraded due to high correlation with BMI. |
| BMI | mass / (height^2) | Buist, I., Bredeweg, S. W., Lemmink, K. A., Van Mechelen, W., & Diercks, R. L. (2010). Predictors of running-related injuries in novice runners enrolled in a systematic training program: a prospective cohort study. *The American journal of sports medicine*, *38*(2), 273-280.<br>Theisen, D., Malisoux, L., Genin, J., Delattre, N., Seil, R., & Urhausen, A. (2014). Influence of midsole hardness of | Numerous prospective studies showing associations between BMI and RRIs. |

| | | |
|---|---|---|
| | | standard cushioned shoes on running-related injury risk. *British Journal of Sports Medicine*, *48*(5), 371-376.<br>Taunton, J. E., Ryan, M. B., Clement, D. B., McKenzie, D. C., Lloyd-Smith, D. R., & Zumbo, B. D. (2003). A prospective study of running injuries: the Vancouver Sun Run "In Training" clinics. *British journal of sports medicine*, *37*(3), 239-244.<br>Malisoux, L., Nielsen, R. O., Urhausen, A., & Theisen, D. (2015). A step towards understanding the mechanisms of running-related injuries. *Journal of Science and Medicine in Sport*, *18*(5), 523-528.<br>Taunton, J. E., Ryan, M. B., Clement, D. B., McKenzie, D. C., Lloyd-Smith, D. R., & Zumbo, B. D. (2002). A retrospective case-control analysis of 2002 running injuries. *British journal of sports medicine*, *36*(2), 95-101. |

| | | | |
|---|---|---|---|
| | | Van Leeuwen, K. D. B., Rogers, J., Winzenberg, T., & van Middelkoop, M. (2016). Higher body mass index is associated with plantar fasciopathy/‘plantar fasciitis’: systematic review and meta-analysis of various clinical and imaging risk factors. *British journal of sports medicine*, *50*(16), 972-981. | |
| Thigh_lean_mass[10] | Lean mass of the thigh region | | Speculative. |
| Thigh_FFMI | Thigh lean mass / (height^2) | | Speculative. |
| Lower_leg_lean_mass[10] | Lean mass of the lower leg region | | Speculative. |
| Lower_leg_FFMI | Lower leg lean mass / (height^2) | | Speculative. |
| Leg_lean_mass[10] | Thigh + lower leg lean mass | Liew, B. X., Zhu, X., Zhai, X., McErlain-Naylor, S. A., & McManus, C. (2024). Association between fat and fat-free | Some evidence pointing to its |

| | | | |
|---|---|---|---|
| | | body mass indices on shock attenuation during running. *Journal of Biomechanics*, *165*, 112025. Carbuhn, A. F., Yu, D., Magee, L. M., McCulloch, P. C., & Lambert, B. S. (2022). Anthropometric factors associated with bone stress injuries in collegiate distance runners: new risk metrics and screening tools?. *Orthopaedic Journal of Sports Medicine*, *10*(2), 23259671211070308. | associations with injury-related factors and with BSIs. |
| Leg_FFMI | Leg lean mass / (height^2) | | Speculative. |
| Total_lean_mass | Lean mass of the entire body | Carbuhn, A. F., Yu, D., Magee, L. M., McCulloch, P. C., & Lambert, B. S. (2022). Anthropometric factors associated with bone stress injuries in collegiate distance runners: new risk metrics and screening tools?. *Orthopaedic Journal of Sports Medicine*, *10*(2), 23259671211070308. | Downgraded due to high correlation with leg lean mass. |

| | | | |
|---|---|---|---|
| Total_FFMI | Total lean mass / (height^2) | Domaradzki, J., & Koźlenia, D. (2022). The performance of body mass component indices in detecting risk of musculoskeletal injuries in physically active young men and women. *PeerJ*, *10*, e12745. | Downgraded due to high correlation with BMI.n |
| Calf_size | Calf muscle cross sectional area at 66% tibial length | | Speculative. |
| BMD_spine | Bone mineral density of L1-L4 spine measured via DXA. | Rauh, M. J., Barrack, M., & Nichols, J. F. (2014). Associations between the female athlete triad and injury among high school runners. *International journal of sports physical therapy*, *9*(7), 948.<br>Rauh, M. J., Nichols, J. F., & Barrack, M. T. (2010). Relationships among injury and disordered eating, menstrual dysfunction, and low bone mineral density in high school athletes: a prospective study. *Journal of athletic training*, *45*(3), 243-252. | Prospective studies showing associations with RRIs. |

| | | | |
|---|---|---|---|
| BMD_hip | Bone mineral density of both hips measured via DXA. | Rauh, M. J., Barrack, M., & Nichols, J. F. (2014). Associations between the female athlete triad and injury among high school runners. *International journal of sports physical therapy*, *9*(7), 948. | Downgraded to to correlation with BMD_spine. |
| BMD_body | Bone mineral density of the entire body measured via DXA. | Rauh, M. J., Barrack, M., & Nichols, J. F. (2014). Associations between the female athlete triad and injury among high school runners. *International journal of sports physical therapy*, *9*(7), 948.<br>Rauh, M. J., Nichols, J. F., & Barrack, M. T. (2010). Relationships among injury and disordered eating, menstrual dysfunction, and low bone mineral density in high school athletes: a prospective study. *Journal of athletic training*, *45*(3), 243-252. | Downgraded to to correlation with BMD_spine. |

**Training (weekly questionnaire)**

| Feature Name | Explanation | References | Evidence Appraisal |
|---|---|---|---|
| Past_month_distance | Average running volume in km during the past 4 weeks | Hootman, J. M., Macera, C. A., Ainsworth, B. E., Martin, M., Addy, C. L., & Blair, S. N. (2002). Predictors of lower extremity injury among recreationally active adults. *Clinical Journal of Sport Medicine*, *12*(2), 99-106.<br>Lysholm, J., & Wiklander, J. (1987). Injuries in runners. *The American journal of sports medicine*, *15*(2), 168-171.<br>Walter, S. D., Hart, L. E., McIntosh, J. M., & Sutton, J. R. (1989). The Ontario cohort study of running-related injuries. *Archives of internal medicine*, *149*(11), 2561-2564.<br>Macera, C. A., Pate, R. R., Powell, K. E., Jackson, K. L., Kendrick, J. S., & Craven, T. E. (1989). Predicting lower- | Prospective studies show correlation between running distance and RRIs. |

| | | | |
|---|---|---|---|
| | | extremity injuries among habitual runners. *Archives of internal medicine*, *149*(11), 2565-2568.<br>Koplan, J. P., Rothenberg, R. B., & Jones, E. L. (1995). The natural history of exercise: a 10-yr follow-up of a cohort of runners. *Medicine and Science in Sports and Exercise*, *27*(8), 1180-1184. | |
| Past_month_min | Average running volume in min during the past 4 weeks | Besomi, M., Leppe, J., Mauri-Stecca, M., Hooper, T., & Sizer, P. (2019). Training volume and previous injury as associated factors for running-related injuries by race distance: a cross-sectional study.<br>Rasmussen, C. H., Nielsen, R. O., Juul, M. S., & Rasmussen, S. (2013). Weekly running volume and risk of running-related injuries among marathon runners. *International journal of sports physical therapy*, *8*(2), 111. | Lack of prospective evidence, plus high correlation with distance volume, thus downgraded. |

| | | | |
|---|---|---|---|
| | | Malisoux, L., Nielsen, R. O., Urhausen, A., & Theisen, D. (2015). A step towards understanding the mechanisms of running-related injuries. *Journal of Science and Medicine in Sport*, *18*(5), 523-528.<br>Wen, D. Y. (2007). Risk factors for overuse injuries in runners. *Current sports medicine reports*, *6*(5), 307-313. | |
| Past_week_ratio | Total running volume in min during the past week divided by the total running volume in min during the week prior | Nielsen, R. Ø., Parner, E. T., Nohr, E. A., Sørensen, H., Lind, M., & Rasmussen, S. (2014). Excessive progression in weekly running distance and risk of running-related injuries: an association which varies according to type of injury. *journal of orthopaedic & sports physical therapy*, *44*(10), 739-747.<br>Nielsen, R. O., Cederholm, P., Buist, I., Sørensen, H., Lind, M., & Rasmussen, S. (2013). Can GPS be used to detect deleterious progression in training volume among | Downgraded due to high correlation with past_month_ratio. The latter is in line with the IOC's definition of acute:chronic workload. |

| | | | |
|---|---|---|---|
| | | runners?. *The Journal of Strength & Conditioning Research*, *27*(6), 1471-1478. Winter, S. C., Gordon, S., Brice, S. M., Lindsay, D., & Barrs, S. (2020). A multifactorial approach to overuse running injuries: a 1-year prospective study. *Sports Health*, *12*(3), 296-303. | |
| Past_month_ratio | Total running volume in min during the past week divided by the average running volume in min during the past 4 weeks | Dijkhuis, T. B., Otter, R., Aiello, M., Velthuijsen, H., & Lemmink, K. (2020). Increase in the acute: chronic workload ratio relates to injury risk in competitive runners. *International journal of sports medicine*, *41*(11), 736-743. Nakaoka, G., Barboza, S. D., Verhagen, E., Van Mechelen, W., & Hespanhol, L. (2021). The association between the acute: chronic workload ratio and running-related injuries in Dutch runners: a prospective cohort study. *Sports* | Prospective studies show correlation with RRIs. |

| | | | |
|---|---|---|---|
| | | *medicine*, *51*, 2437-2447. | |
| Past_month_volume_low | Average low intensity running volume in min during the past 4 weeks | | Speculative. |
| Past_week_ratio_low | Total low intensity running volume in min during the past week divided by the total running volume in min during the week prior | | Speculative. |
| Past_month_ratio_low | Total low intensity running volume in min during the past week divided by the average running volume in min during the past 4 weeks | | Speculative. |
| Past_month_volume_moderate | | | Speculative. |
| Past_week_ra | | | Speculative. |

| | | | |
|---|---|---|---|
| tio_moderate | | | |
| Past_month_ratio_moderate | | | Speculative. |
| Past_month_volume_high | | | Speculative. |
| Past_week_ratio_high | | | Speculative. |
| Past_month_ratio_high | | | Speculative. |
| Past_month_volume_very_high | | | Speculative. |
| Past_week_ratio_very_high | | | Speculative. |
| Past_month_r | | | Speculative. |

| | | | |
|---|---|---|---|
| atio_very_high | | | |
| Past_month_calculated_volume | Average running volume during the past 4 weeks, calculated via “minutes under intensity * RPE” | Same as above with a different method of volume calculation | Speculative. Method of calculation is invented in this study. |
| Past_week_ratio_calculated_volume | | Same as above with a different method of volume calculation | Speculative. |
| Past_month_ratio_calculated_volume | | Same as above with a different method of volume calculation | Speculative. |
| Resistance_training_past_month | Total minutes of resistance training conducted during the past 4 weeks | | Correlated with the past season feature. |
| Resistance_tr | Total minutes of resistance training | Leppänen, M., Viiala, J., Kaikkonen, P., Tokola, K., | Downgraded due to |

| | | | |
|---|---|---|---|
| aining_past_s eason | conducted during the past 12 weeks | Vasankari, T., Nigg, B. M., ... & Pasanen, K. (2024). Hip and core exercise programme prevents running-related overuse injuries in adult novice recreational runners: a three-arm randomised controlled trial (Run RCT). *British Journal of Sports Medicine*, *58*(13), 722-732. Desai, P., Jungmalm, J., Börjesson, M., Karlsson, J., & Grau, S. (2023). Effectiveness of an 18-week general strength and foam-rolling intervention on running-related injuries in recreational runners. *Scandinavian Journal of Medicine & Science in Sports*, *33*(5), 766-775. Mendez-Rebolledo, G., Figueroa-Ureta, R., Moya-Mura, F., Guzmán-Muñoz, E., Ramirez-Campillo, R., & Lloyd, R. S. (2021). The protective effect of neuromuscular training on the medial tibial stress syndrome in youth female track-and-field athletes: a clinical trial and cohort | correlation with total SC. |

| | | | |
|---|---|---|---|
| | | study. *Journal of sport rehabilitation*, *30*(7), 1019-1027. Wu, H., Brooke-Wavell, K., Fong, D. T., Paquette, M. R., & Blagrove, R. C. (2024). Do Exercise-Based Prevention Programs Reduce Injury in Endurance Runners? A Systematic Review and Meta-Analysis. *Sports Medicine*, 1-19. | |
| Bodyweight_exercises_past_month | Total minutes of bodyweight exercises conducted during the past 4 weeks | | Correlated with the past season feature. |
| Bodyweight_exercises_past_season | Total minutes of bodyweight exercises conducted during the past 12 weeks | Mendez-Rebolledo, G., Figueroa-Ureta, R., Moya-Mura, F., Guzmán-Muñoz, E., Ramirez-Campillo, R., & Lloyd, R. S. (2021). The protective effect of neuromuscular training on the medial tibial stress syndrome in youth female track-and-field athletes: a clinical trial and cohort study. *Journal of sport rehabilitation*, *30*(7), 1019-1027. | Downgraded due to correlation with total SC. |

| | | | |
|---|---|---|---|
| | | Wu, H., Brooke-Wavell, K., Fong, D. T., Paquette, M. R., & Blagrove, R. C. (2024). Do Exercise-Based Prevention Programs Reduce Injury in Endurance Runners? A Systematic Review and Meta-Analysis. *Sports Medicine*, 1-19. | |
| Core_stability _past_month | Total minutes of core stability exercises conducted during the past 4 weeks | | Correlated with the past season feature. |
| Core_stability _past_season | Total minutes of core stability exercises conducted during the past 12 weeks | Leppänen, M., Viiala, J., Kaikkonen, P., Tokola, K., Vasankari, T., Nigg, B. M., ... & Pasanen, K. (2024). Hip and core exercise programme prevents running-related overuse injuries in adult novice recreational runners: a three-arm randomised controlled trial (Run RCT). *British Journal of Sports Medicine*, *58*(13), 722-732. Mendez-Rebolledo, G., Figueroa-Ureta, R., Moya-Mura, | Downgraded due to correlation with total SC. |

| | | | |
|---|---|---|---|
| | | F., Guzmán-Muñoz, E., Ramirez-Campillo, R., & Lloyd, R. S. (2021). The protective effect of neuromuscular training on the medial tibial stress syndrome in youth female track-and-field athletes: a clinical trial and cohort study. *Journal of sport rehabilitation*, *30*(7), 1019-1027. Wu, H., Brooke-Wavell, K., Fong, D. T., Paquette, M. R., & Blagrove, R. C. (2024). Do Exercise-Based Prevention Programs Reduce Injury in Endurance Runners? A Systematic Review and Meta-Analysis. *Sports Medicine*, 1-19. | |
| Balance_training_past_month | Total minutes of balance training conducted during the past 4 weeks | | Correlated with the past season feature. |
| Balance_training_past_seas | Total minutes of balance training conducted during the past 12 weeks | Mendez-Rebolledo, G., Figueroa-Ureta, R., Moya-Mura, F., Guzmán-Muñoz, E., Ramirez-Campillo, R., & Lloyd, | Downgraded due to correlation with total |

| | | | |
|---|---|---|---|
| on | | R. S. (2021). The protective effect of neuromuscular training on the medial tibial stress syndrome in youth female track-and-field athletes: a clinical trial and cohort study. *Journal of sport rehabilitation*, *30*(7), 1019-1027. Wu, H., Brooke-Wavell, K., Fong, D. T., Paquette, M. R., & Blagrove, R. C. (2024). Do Exercise-Based Prevention Programs Reduce Injury in Endurance Runners? A Systematic Review and Meta-Analysis. *Sports Medicine*, 1-19. | SC. |
| plyometrics_past_month | Total minutes of plyometrics conducted during the past 4 weeks | | Correlated with the past season feature. |
| plyometrics_past_season | Total minutes of plyometrics conducted during the past 12 weeks | Mendez-Rebolledo, G., Figueroa-Ureta, R., Moya-Mura, F., Guzmán-Muñoz, E., Ramirez-Campillo, R., & Lloyd, R. S. (2021). The protective effect of neuromuscular training on the medial tibial stress syndrome in youth | Downgraded due to correlation with total SC. |

| | | | |
|---|---|---|---|
| | | female track-and-field athletes: a clinical trial and cohort study. *Journal of sport rehabilitation*, *30*(7), 1019-1027. Wu, H., Brooke-Wavell, K., Fong, D. T., Paquette, M. R., & Blagrove, R. C. (2024). Do Exercise-Based Prevention Programs Reduce Injury in Endurance Runners? A Systematic Review and Meta-Analysis. *Sports Medicine*, 1-19. | |
| drills_past_month | Total minutes of running technique drills conducted during the past 4 weeks | | Correlated with the past season feature. |
| drills_past_season | Total minutes of running technique drills conducted during the past 12 weeks | Mendez-Rebolledo, G., Figueroa-Ureta, R., Moya-Mura, F., Guzmán-Muñoz, E., Ramirez-Campillo, R., & Lloyd, R. S. (2021). The protective effect of neuromuscular training on the medial tibial stress syndrome in youth female track-and-field athletes: a clinical trial and cohort | Downgraded due to correlation with total SC. |

| | | | |
|---|---|---|---|
| | | study. *Journal of sport rehabilitation*, *30*(7), 1019-1027.<br>Wu, H., Brooke-Wavell, K., Fong, D. T., Paquette, M. R., & Blagrove, R. C. (2024). Do Exercise-Based Prevention Programs Reduce Injury in Endurance Runners? A Systematic Review and Meta-Analysis. *Sports Medicine*, 1-19. | |
| Circuit_training_past_month | Total minutes of circuit training conducted during the past 4 weeks | | Correlated with the past season feature. |
| Circuit_training_past_season | Total minutes of circuit training conducted during the past 12 weeks | | Speculative. |
| barefoot_past_month | Total minutes of barefoot exercises conducted during the past 4 weeks | | Correlated with the past season feature. |
| barefoot_past | Total minutes of barefoot exercises | | Speculative. |

| | | | |
|---|---|---|---|
| _season | conducted during the past 12 weeks | | |
| stretching_past_month | Total minutes of stretching or yoga conducted during the past 4 weeks | | Correlated with the past season feature. |
| stretching_past_season | Total minutes of stretching or yoga conducted during the past 12 weeks | Weldon, S. M., & Hill, R. H. (2003). The efficacy of stretching for prevention of exercise-related injury: a systematic review of the literature. *Manual therapy*, *8*(3), 141-150. | Downgraded due to correlation with total SC. |
| SC_past_month | Total minutes of all of the above exercises conducted during the past 4 weeks | | Correlated with the past season feature. |
| SC_past_season | Total minutes of all of the above exercises conducted during the past 12 weeks | Leppänen, M., Viiala, J., Kaikkonen, P., Tokola, K., Vasankari, T., Nigg, B. M., ... & Pasanen, K. (2024). Hip and core exercise programme prevents running-related overuse injuries in adult novice recreational runners: a three-arm randomised controlled trial (Run RCT). *British* | Prospective intervention studies show supervised S&C has an effect on RRIs. |

| | | |
|---|---|---|
| | | *Journal of Sports Medicine*, *58*(13), 722-732.<br>Mendez-Rebolledo, G., Figueroa-Ureta, R., Moya-Mura, F., Guzmán-Muñoz, E., Ramirez-Campillo, R., & Lloyd, R. S. (2021). The protective effect of neuromuscular training on the medial tibial stress syndrome in youth female track-and-field athletes: a clinical trial and cohort study. *Journal of sport rehabilitation*, *30*(7), 1019-1027.<br>Desai, P., Jungmalm, J., Börjesson, M., Karlsson, J., & Grau, S. (2023). Effectiveness of an 18-week general strength and foam-rolling intervention on running-related injuries in recreational runners. *Scandinavian Journal of Medicine & Science in Sports*, *33*(5), 766-775.<br>Taddei, U. T., Matias, A. B., Duarte, M., & Sacco, I. C. (2020). Foot core training to prevent running-related injuries: a survival analysis of a single-blind, randomized |

| | | | |
|---|---|---|---|
| | | controlled trial. *The American journal of sports medicine*, *48*(14), 3610-3619.<br>Wu, H., Brooke-Wavell, K., Fong, D. T., Paquette, M. R., & Blagrove, R. C. (2024). Do Exercise-Based Prevention Programs Reduce Injury in Endurance Runners? A Systematic Review and Meta-Analysis. *Sports Medicine*, 1-19. | |
| Non_running_past_month | Total minutes of non-running (and non-S&C) exercises conducted during the past 4 weeks | | Correlated with the past season feanture. |
| Non_running_past_season | Total minutes of all of non-running (and non-S&C) exercises conducted during the past 12 weeks | This is to control for total training volume by taking into account sports other than running: some people are pure runners, while some do other sports. | Included as a complementary factor for running training volume. |
| Past_month_i | Whether injury was detected within | | It has been observed |

| | | | |
|---|---|---|---|
| njury | the 4 weeks prior to the target week | | that participants who keep training after a recent injury tend to worsen the extent of the injury (re-injuries). |
| Tracking_period_injury | The participant's injury severity score summed up over all previous weeks / total number of previous weeks relative to current week (if multiple injuries occurred during a week, severity score is calculated by highest region's score + ((100 - highest region's score) * second highest region's score) / 100), and so on for additional regions) | Buist, I., Bredeweg, S. W., Lemmink, K. A., Van Mechelen, W., & Diercks, R. L. (2010). Predictors of running-related injuries in novice runners enrolled in a systematic training program: a prospective cohort study. *The American journal of sports medicine*, *38*(2), 273-280.<br>Theisen, D., Malisoux, L., Genin, J., Delattre, N., Seil, R., & Urhausen, A. (2014). Influence of midsole hardness of standard cushioned shoes on running-related injury risk. *British Journal of Sports Medicine*, *48*(5), 371-376. | Ample evidence in prospective studies showing links between previous RRI and future RRI. |

| | | |
|---|---|---|
| | | Saragiotto, B. T., Yamato, T. P., Hespanhol Junior, L. C., Rainbow, M. J., Davis, I. S., & Lopes, A. D. (2014). What are the main risk factors for running-related injuries?. *Sports medicine*, *44*, 1153-1163.<br>Hulme, A., Nielsen, R. O., Timpka, T., Verhagen, E., & Finch, C. (2017). Risk and protective factors for middle- and long-distance running-related injury. *Sports Medicine*, *47*, 869-886. |

### Genetics

| rsID | Gene and Function | Risky Allele | References | Evidence Appraisal |
|---|---|---|---|---|
| rs3753841[11] | COL11A1, collagen XI fiber | Ref/alt | Alakhdar Mohmara, Y., Cook, J., Benítez-Martínez, J. C., McPeek, E. R., Aguilar, A. A., Olivas, E. S., & Hernandez-Sanchez, S. (2020). Influence of genetic factors in elbow | Not RRI, weak evidence. |

| | | | | |
|---|---|---|---|---|
| | | | tendon pathology: a case-control study. *Scientific Reports*, *10*(1), 6503. | |
| rs7528684[12] | FCRL3, immunoglobulin receptor | Alt | Salles, J. I., Lopes, L. R., Duarte, M. E. L., Morrissey, D., Martins, M. B., Machado, D. E., ... & Perini, J. A. (2018). Fc receptor-like 3 (− 169T> C) polymorphism increases the risk of tendinopathy in volleyball athletes: a case control study. *BMC medical genetics*, *19*, 1-10. | Function points more towards autoimmune conditions (rheumatoid arthritis) but linkage with lower limb tendinopathy. |
| rs57104447 | CACNA1E, calcium voltage-gated channel subunit | Alt | Kim, S. K., Roos, T. R., Roos, A. K., Kleimeyer, J. P., Ahmed, M. A., Goodlin, G. T., ... & Dragoo, J. L. (2017). Genome-wide association screens for Achilles tendon and ACL tears and tendinopathy. *PloS one*, *12*(3), e0170422. | Function does not point to physical structrual integrity of tendon but linkage with lower limb tendinopathy. **rs183364169 |
| rs1887632 | SOST, provides instructions for making the protein | Alt | Varley, I., Hughes, D. C., Greeves, J. P., Stellingwerff, T., Ranson, C., Fraser, W. D., & Sale, C. (2018). The association of novel polymorphisms with stress fracture | Related to stress fracture, very few cases in current study. |

| | | | | |
|---|---|---|---|---|
| | sclerostin | | injury in Elite Athletes: Further insights from the SFEA cohort. *Journal of science and medicine in sport*, *21*(6), 564-568. | |
| rs4654760 | TNAP, non-specific alkaline phosphatase | Ref/alt | Peach, C. A., Zhang, Y., Dunford, J. E., Brown, M. A., & Carr, A. J. (2007). Cuff tear arthropathy: evidence of functional variation in pyrophosphate metabolism genes. *Clinical Orthopaedics and Related Research (1976-2007)*, *462*, 67-72. | Function does not seem to be injury-related, reference is not RRI. |
| rs1137101 | LEPR, leptin receptor | Alt | Wang, Y., Meng, F., Wu, J., Long, H., Li, J., Wu, Z., ... & Xie, D. (2022). Associations between adipokines gene polymorphisms and knee osteoarthritis: a meta-analysis. *BMC musculoskeletal disorders*, *23*(1), 166. | Function does not seem to be injury-related, reference is not RRI. |
| rs4919510[13] | MIR608, microRNA-related | Ref/ref | Abrahams, Y., Laguette, M. J., Prince, S., & Collins, M. (2013). Polymorphisms within the COL5A1 3′-UTR that alters mRNA structure and the MIR608 gene are associated | Function does not seem to link to injury, but association found for |

| | | | | |
|---|---|---|---|---|
| | | | with Achilles tendinopathy. *Annals of human genetics*, *77*(3), 204-214. | achilles tendinopathy which is a common RRI. |
| rs1937810 | MPP7, establishment of epithelial cell polarity | Alt | Kim, S. K., Nguyen, C., Avins, A. L., & Abrams, G. D. (2021). Identification of Three Loci Associated with Achilles Tendon Injury Risk from a Genome-wide Association Study. *Medicine and science in sports and exercise*, *53*(8), 1748.<br>Kim, S. K., Roos, T. R., Roos, A. K., Kleimeyer, J. P., Ahmed, M. A., Goodlin, G. T., ... & Dragoo, J. L. (2017). Genome-wide association screens for Achilles tendon and ACL tears and tendinopathy. *PloS one*, *12*(3), e0170422.<br>Kang, X., Tian, B., Zhang, L., Ge, Z., Zhao, Y., & Zhang, Y. (2019). Relationship of common variants in MPP7, TIMP2 and CASP8 genes with the risk of chronic achilles tendinopathy. *Scientific Reports*, *9*(1), 17627. | Function does not seem to link to injury, but association found for achilles tendinopathy which is a common RRI. |

| | | | | |
|---|---|---|---|---|
| rs6481512 | MPP7, establishment of epithelial cell polarity | Alt | Kim, S. K., Nguyen, C., Avins, A. L., & Abrams, G. D. (2021). Identification of Three Loci Associated with Achilles Tendon Injury Risk from a Genome-wide Association Study. *Medicine and science in sports and exercise*, *53*(8), 1748. | Function does not seem to link to injury, but association found for achilles tendinopathy which is a common RRI. |
| rs1249269 | MPP7, establishment of epithelial cell polarity | Ref | Kim, S. K., Nguyen, C., Avins, A. L., & Abrams, G. D. (2021). Identification of Three Loci Associated with Achilles Tendon Injury Risk from a Genome-wide Association Study. *Medicine and science in sports and exercise*, *53*(8), 1748. | Function does not seem to link to injury, but association found for achilles tendinopathy which is a common RRI. |
| rs11225395 | MMP8, matrix metalloproteinase, degradation of type I, II and III collagens | Ref | Godoy-Santos, A., Ortiz, R. T., Junior, R. M., Fernandes, T. D., & Santos, M. C. L. G. (2014). MMP-8 polymorphism is genetic marker to tendinopathy primary posterior tibial tendon. *Scandinavian journal of medicine & science in sports*, *24*(1), 220-223. | Both function and references point to potential RRIs. |

| | | | | |
|---|---|---|---|---|
| | | | de Araujo Munhoz, F. B., Baroneza, J. E., Godoy-Santos, A., Fernandes, T. D., Branco, F. P., Alle, L. F., ... & Dos Santos, M. C. L. G. (2016). Posterior tibial tendinopathy associated with matrix metalloproteinase 13 promoter genotype and haplotype. *The Journal of Gene Medicine*, *18*(11-12), 325-330.<br>Godoy-Santos, A., Cunha, M. V., Ortiz, R. T., Fernandes, T. D., Mattar Jr, R., & dos Santos, M. C. L. (2013). MMP-1 promoter polymorphism is associated with primary tendinopathy of the posterior tibial tendon. *Journal of Orthopaedic Research*, *31*(7), 1103-1107. | |
| rs1144393 | MMP1, matrix metalloproteinase, breaks down the interstitial collagens | Alt | de Araujo Munhoz, F. B., Baroneza, J. E., Godoy-Santos, A., Fernandes, T. D., Branco, F. P., Alle, L. F., ... & Dos Santos, M. C. L. G. (2016). Posterior tibial tendinopathy associated with matrix metalloproteinase 13 promoter | Both function and references point to potential RRIs. |

| | | | | |
|---|---|---|---|---|
| | | | genotype and haplotype. *The Journal of Gene Medicine*, *18*(11-12), 325-330.<br>Baroneza, J. E., Godoy-Santos, A., Massa, B. F., de Araujo Munhoz, F. B., Fernandes, T. D., & dos Santos, M. C. L. G. (2014). MMP-1 promoter genotype and haplotype association with posterior tibial tendinopathy. *Gene*, *547*(2), 334-337. | |
| rs650108 | MMP3, matrix metalloproteinase, degrades fibronectin, laminin, collagens III, IV, IX, and X | Alt/alt | Raleigh, S. M., Van der Merwe, L., & Ribbans, W. J. (2009). Variants within the MMP3 gene are associated with Achilles tendinopathy. *possible interaction with the COL5A1 gene*, *2009*, 43.<br>Briški, N., Vrgoč, G., Knjaz, D., Janković, S., Ivković, A., Pećina, M., & Lauc, G. (2021). Association of the matrix metalloproteinase 3 (MMP3) single nucleotide polymorphisms with tendinopathies: case-control study in | Both function and references point to potential RRIs. |

| | | | | |
|---|---|---|---|---|
| | | | high-level athletes. *International orthopaedics*, *45*, 1163-1168. | |
| rs679620 / rs591058* | MMP3, matrix metalloproteinase, degrades fibronectin, laminin, collagens III, IV, IX, and X | Alt | Raleigh, S. M., Van der Merwe, L., & Ribbans, W. J. (2009). Variants within the MMP3 gene are associated with Achilles tendinopathy. *possible interaction with the COL5A1 gene*, *2009*, 43.<br>Nie, G., Wen, X., Liang, X., Zhao, H., Li, Y., & Lu, J. (2019). Additional evidence supports association of common genetic variants in MMP3 and TIMP2 with increased risk of chronic Achilles tendinopathy susceptibility. *Journal of science and medicine in sport*, *22*(10), 1074-1078.<br>Briški, N., Vrgoč, G., Knjaz, D., Janković, S., Ivković, A., Pećina, M., & Lauc, G. (2021). Association of the matrix metalloproteinase 3 (MMP3) single nucleotide | Both function and references point to potential RRIs. |

| | | | | |
|---|---|---|---|---|
| | | | polymorphisms with tendinopathies: case-control study in high-level athletes. *International orthopaedics*, *45*, 1163-1168.<br>Figueiredo, E. A., Loyola, L. C., Belangero, P. S., Campos Ribeiro-dos-Santos, Â. K., Emanuel Batista Santos, S., Cohen, C., ... & Leal, M. F. (2020). Rotator cuff tear susceptibility is associated with variants in genes involved in tendon extracellular matrix homeostasis. *Journal of Orthopaedic Research®*, *38*(1), 192-201. | |
| rs2252070 | MMP13, matrix metalloproteinase, cleaves type II collagen more efficiently than types I and III | Alt | de Araujo Munhoz, F. B., Baroneza, J. E., Godoy-Santos, A., Fernandes, T. D., Branco, F. P., Alle, L. F., ... & Dos Santos, M. C. L. G. (2016). Posterior tibial tendinopathy associated with matrix metalloproteinase 13 promoter genotype and haplotype. *The Journal of Gene Medicine*, *18*(11-12), 325-330. | Both function and references point to potential RRIs. |

| | | | | |
|---|---|---|---|---|
| rs2306033 | LRP4, LDL receptor protein | Ref/ref | Yanovich, R., Friedman, E., Milgrom, R., Oberman, B., Freedman, L., & Moran, D. S. (2012). Candidate gene analysis in Israeli soldiers with stress fractures. *Journal of Sports Science & Medicine*, *11*(1), 147. | Function does not seem to directly cause injury, reference is stress fracture, very few cases in current study. |
| rs2277268 | LRP5, LDL receptor protein | Alt | Korvala, J., Hartikka, H., Pihlajamäki, H., Solovieva, S., Ruohola, J. P., Sahi, T., ... & Männikkö, M. (2010). Genetic predisposition for femoral neck stress fractures in military conscripts. *BMC genetics*, *11*, 1-9. | Function does not seem to directly cause injury, reference is stress fracture, very few cases in current study. |
| rs4988321 | LRP5, LDL receptor protein | Alt | Korvala, J., Hartikka, H., Pihlajamäki, H., Solovieva, S., Ruohola, J. P., Sahi, T., ... & Männikkö, M. (2010). Genetic predisposition for femoral neck stress fractures in military conscripts. *BMC genetics*, *11*, 1-9. | Function does not seem to directly cause injury, reference is stress fracture, very few cases in current study. |

| | | | | |
|---|---|---|---|---|
| rs12574452 | FGF3, fibroblast growth factor, embryonic development, cell growth, morphogenesis, tissue repair, tumor growth | Alt/alt | da Rocha Motta, G., Amaral, M. V., Rezende, E., Pitta, R., dos Santos Vieira, T. C., Duarte, M. E. L., ... & Casado, P. L. (2014). Evidence of genetic variations associated with rotator cuff disease. *Journal of shoulder and elbow surgery*, *23*(2), 227-235.<br>Tashjian, R. Z., Kim, S. K., Roche, M. D., Jones, K. B., & Teerlink, C. C. (2021). Genetic variants associated with rotator cuff tearing utilizing multiple population-based genetic resources. *Journal of Shoulder and Elbow Surgery*, *30*(3), 520-531. | Function links to connective tissue repair, but references not with RRI. |
| rs11232681 | RP11-664H7.2, pseudogene | Alt | Sood, R. F., Westenberg, R. F., Winograd, J. M., Eberlin, K. R., & Chen, N. C. (2020). Genetic risk of trigger finger: results of a genomewide association study. *Plastic and reconstructive surgery*, *146*(2), 165e-176e. | Function unclear and reference not RRI. |
| rs1718119 | P2RX7, ATP- | Ref/alt | Varley, I., Greeves, J. P., Sale, C., Friedman, E., Moran, D. | Function does not seem to |

| | | | | |
|---|---|---|---|---|
| | dependent lysis of macrophages | | S., Yanovich, R., ... & Gallagher, J. A. (2016). Functional polymorphisms in the P2X7 receptor gene are associated with stress fracture injury. *Purinergic signalling*, *12*, 103-113. | directly cause injury, reference is stress fracture, very few cases in current study. |
| rs3751143 | P2RX7, ATP-dependent lysis of macrophages | Ref/alt | Varley, I., Greeves, J. P., Sale, C., Friedman, E., Moran, D. S., Yanovich, R., ... & Gallagher, J. A. (2016). Functional polymorphisms in the P2X7 receptor gene are associated with stress fracture injury. *Purinergic signalling*, *12*, 103-113. | Function does not seem to directly cause injury, reference is stress fracture, very few cases in current study. |
| rs1544410 | VDR, vitamin D receptor | Ref | Chatzipapas, C., Boikos, S., Drosos, G. I., Kazakos, K., Tripsianis, G., Serbis, A., ... & Stratakis, C. A. (2009). Polymorphisms of the vitamin D receptor gene and stress fractures. *Hormone and Metabolic Research*, *41*(08), 635-640. | Function and reference both point to stress fracture, very few cases in current study. |
| rs2228570 | VDR, vitamin D | Ref | Chatzipapas, C., Boikos, S., Drosos, G. I., Kazakos, K., | Function and reference both |

| | | | | |
|---|---|---|---|---|
| | receptor | | Tripsianis, G., Serbis, A., ... & Stratakis, C. A. (2009). Polymorphisms of the vitamin D receptor gene and stress fractures. *Hormone and Metabolic Research*, *41*(08), 635-640. | point to stress fracture, very few cases in current study. |
| rs4328262 | VDR, vitamin D receptor | Ref/ref, Alt/alt | Yanovich, R., Friedman, E., Milgrom, R., Oberman, B., Freedman, L., & Moran, D. S. (2012). Candidate gene analysis in Israeli soldiers with stress fractures. *Journal of Sports Science & Medicine*, *11*(1), 147. | Function and reference both point to stress fracture, very few cases in current study. |
| rs1021188 | RANKL, osteoclastogenesis | Alt/alt | Varley, I., Hughes, D. C., Greeves, J. P., Stellingwerff, T., Ranson, C., Fraser, W. D., & Sale, C. (2015). RANK/RANKL/OPG pathway: genetic associations with stress fracture period prevalence in elite athletes. *Bone*, *71*, 131-136. | Function and reference both point to stress fracture, very few cases in current study. |
| rs74544784 | KLHL1, actin-organizing proteins | Alt | Sood, R. F., Westenberg, R. F., Winograd, J. M., Eberlin, K. R., & Chen, N. C. (2020). Genetic risk of trigger finger: | Function seems close to musculoskeletal problems |

| | | | | |
|---|---|---|---|---|
| | | | results of a genomewide association study. *Plastic and reconstructive surgery*, *146*(2), 165e-176e. | but reference not with RRI. **rs12429486 |
| rs12429486 / rs113732656 / rs113033795 / rs111237638 / rs113796757 / rs59988404 / rs113390517 / rs12429555 / rs12429576 / rs12428036 / rs76743567 / rs117409997 / rs74092661* | KLHL1, actin-organizing proteins | Alt | Sood, R. F., Westenberg, R. F., Winograd, J. M., Eberlin, K. R., & Chen, N. C. (2020). Genetic risk of trigger finger: results of a genomewide association study. *Plastic and reconstructive surgery*, *146*(2), 165e-176e. | Function seems close to musculoskeletal problems but reference not with RRI. |

| | | | | |
|---|---|---|---|---|
| rs78391032 | KLHL1, actin-organizing proteins | Alt | Sood, R. F., Westenberg, R. F., Winograd, J. M., Eberlin, K. R., & Chen, N. C. (2020). Genetic risk of trigger finger: results of a genomewide association study. *Plastic and reconstructive surgery*, *146*(2), 165e-176e. | Function seems close to musculoskeletal problems but reference not with RRI. **rs12429486 |
| rs77569527 | KLHL1, actin-organizing proteins | Alt | Sood, R. F., Westenberg, R. F., Winograd, J. M., Eberlin, K. R., & Chen, N. C. (2020). Genetic risk of trigger finger: results of a genomewide association study. *Plastic and reconstructive surgery*, *146*(2), 165e-176e. | Function seems close to musculoskeletal problems but reference not with RRI. **rs12429486 |
| rs117544024 | KLHL1, actin-organizing proteins | Alt | Sood, R. F., Westenberg, R. F., Winograd, J. M., Eberlin, K. R., & Chen, N. C. (2020). Genetic risk of trigger finger: results of a genomewide association study. *Plastic and reconstructive surgery*, *146*(2), 165e-176e. | Function seems close to musculoskeletal problems but reference not with RRI. **rs12429486 |
| rs4454832 | SOX21, encode a family of DNA-binding proteins | Ref | Kim, S. K., Nguyen, C., Avins, A. L., & Abrams, G. D. (2021). Identification of Three Loci Associated with Achilles Tendon Injury Risk from a Genome-wide | Function does not relate to RRI, but reference on Achilles tendon injury. |

| | | | | |
|---|---|---|---|---|
| | | | Association Study. *Medicine and science in sports and exercise*, *53*(8), 1748. | |
| rs912336 | STK24, encodes a serine/threonine protein kinase | Ref | Kim, S. K., Nguyen, C., Jones, K. B., & Tashjian, R. Z. (2021). A genome-wide association study for shoulder impingement and rotator cuff disease. *Journal of shoulder and elbow surgery*, *30*(9), 2134-2145. | Neither function nor reference directly point to RRI. |
| rs3218791 / rs8003305* | POLE2, DNA polymerase epsilon, involved in DNA repair and replication | Alt | Sood, R. F., Westenberg, R. F., Winograd, J. M., Eberlin, K. R., & Chen, N. C. (2020). Genetic risk of trigger finger: results of a genomewide association study. *Plastic and reconstructive surgery*, *146*(2), 165e-176e. | Neither function nor reference directly point to RRI. |
| rs2761884 | BMP4, bone morphogentic protein, heart development and | Alt | Salles, J. I., Amaral, M. V., Aguiar, D. P., Lira, D. A., Quinelato, V., Bonato, L. L., ... & Casado, P. L. (2015). BMP4 and FGF3 haplotypes increase the risk of tendinopathy in volleyball athletes. *Journal of science and* | Function does not point to physical structrual integrity of tendon but linkage with lower limb tendinopathy. |

| | | | | |
|---|---|---|---|---|
| | adipogenesis | | *medicine in sport*, *18*(2), 150-155. | |
| rs4986938 | ESR2, estrogen receptor | Alt | Nogara, P. R. B., Godoy-Santos, A. L., Fonseca, F. C. P., Cesar-Netto, C., Carvalho, K. C., Baracat, E. C., ... & Santos, M. C. L. (2020). Association of estrogen receptor β polymorphisms with posterior tibial tendon dysfunction. *Molecular and cellular biochemistry*, *471*, 63-69. | Both function and reference seem to link to common RRIs especially in females. |
| rs911263 | RAD51B, DNA repair by homologous recombination | Alt | Zhi, L., Yao, S., Ma, W., Zhang, W., Chen, H., Li, M., & Ma, J. (2017). Polymorphisms of RAD51B are associated with rheumatoid arthritis and erosion in rheumatoid arthritis patients. *Scientific Reports*, *7*(1), 45876. | Neither function nor reference directly point to RRI. |
| rs2525504 | RAD51B, DNA repair by homologous recombination | Alt | Zhi, L., Yao, S., Ma, W., Zhang, W., Chen, H., Li, M., & Ma, J. (2017). Polymorphisms of RAD51B are associated with rheumatoid arthritis and erosion in rheumatoid arthritis patients. *Scientific Reports*, *7*(1), 45876. | Neither function nor reference directly point to RRI. |

| | | | | |
|---|---|---|---|---|
| rs17756404 | RAD51B, DNA repair by homologous recombination | Ref | Zhi, L., Yao, S., Ma, W., Zhang, W., Chen, H., Li, M., & Ma, J. (2017). Polymorphisms of RAD51B are associated with rheumatoid arthritis and erosion in rheumatoid arthritis patients. *Scientific Reports*, *7*(1), 45876. | Neither function nor reference directly point to RRI. |
| rs4903399 | ESRRB, estrogen-realted receptor beta | Ref/ref | da Rocha Motta, G., Amaral, M. V., Rezende, E., Pitta, R., dos Santos Vieira, T. C., Duarte, M. E. L., ... & Casado, P. L. (2014). Evidence of genetic variations associated with rotator cuff disease. *Journal of shoulder and elbow surgery*, *23*(2), 227-235.<br>Bonato, L. L., Quinelato, V., Amaral, M. V. G., de Souza, F. N., Lobo, J. C., Aguiar, D. P., ... & Casado, P. L. (2016). ESRRB polymorphisms are associated with comorbidity of temporomandibular disorders and rotator cuff disease. *International Journal of Oral and Maxillofacial Surgery*, *45*(3), 323-331. | Neither function nor reference directly point to RRI (‘estrogen-related’ receptor is not actually an estrogen receptor). |

| | | | | |
|---|---|---|---|---|
| rs10132091 | ESRRB, estrogen-realted receptor beta | Ref | Bonato, L. L., Quinelato, V., Amaral, M. V. G., de Souza, F. N., Lobo, J. C., Aguiar, D. P., ... & Casado, P. L. (2016). ESRRB polymorphisms are associated with comorbidity of temporomandibular disorders and rotator cuff disease. *International Journal of Oral and Maxillofacial Surgery*, *45*(3), 323-331. | Neither function nor reference directly point to RRI. |
| rs17583842 | ESRRB, estrogen-realted receptor beta | Alt | Teerlink, C. C., Cannon-Albright, L. A., & Tashjian, R. Z. (2015). Significant association of full-thickness rotator cuff tears and estrogen-related receptor-β (ESRRB). *Journal of shoulder and elbow surgery*, *24*(2), e31-e35. | Neither function nor reference directly point to RRI. |
| rs1676303 | ESRRB, estrogen-realted receptor beta | Ref | da Rocha Motta, G., Amaral, M. V., Rezende, E., Pitta, R., dos Santos Vieira, T. C., Duarte, M. E. L., ... & Casado, P. L. (2014). Evidence of genetic variations associated with rotator cuff disease. *Journal of shoulder and elbow surgery*, *23*(2), 227-235. | Neither function nor reference directly point to RRI. |

| | | | | |
|---|---|---|---|---|
| | | | Bonato, L. L., Quinelato, V., Amaral, M. V. G., de Souza, F. N., Lobo, J. C., Aguiar, D. P., ... & Casado, P. L. (2016). ESRRB polymorphisms are associated with comorbidity of temporomandibular disorders and rotator cuff disease. *International Journal of Oral and Maxillofacial Surgery*, *45*(3), 323-331. | |
| rs11629171 | SERPINA6, serine proteinase inhibitor, glucorticoids and progestins transport | Alt | Yanovich, R., Friedman, E., Milgrom, R., Oberman, B., Freedman, L., & Moran, D. S. (2012). Candidate gene analysis in Israeli soldiers with stress fractures. *Journal of Sports Science & Medicine*, *11*(1), 147. | Function does not seem to link to RRIs, and reference is in stress fracture, too few cases in current study. |
| rs2281518 | SERPINA6, serine proteinase inhibitor, glucorticoids and progestins transport | Ref/alt | Yanovich, R., Friedman, E., Milgrom, R., Oberman, B., Freedman, L., & Moran, D. S. (2012). Candidate gene analysis in Israeli soldiers with stress fractures. *Journal of Sports Science & Medicine*, *11*(1), 147. | Function does not seem to link to RRIs, and reference is in stress fracture, too few cases in current study. |
| rs2285053 | MMP2, matrix | Alt | Figueiredo, E. A., Loyola, L. C., Belangero, P. S., Campos | Neither function nor |

| | | | | |
|---|---|---|---|---|
| | metalloproteinase, gelatinase A, type IV collagenase | | Ribeiro-dos-Santos, Â. K., Emanuel Batista Santos, S., Cohen, C., ... & Leal, M. F. (2020). Rotator cuff tear susceptibility is associated with variants in genes involved in tendon extracellular matrix homeostasis. *Journal of Orthopaedic Research®*, *38*(1), 192-201. | reference directly point to RRI. |
| rs71404070 | CDH8, cell adhesion | Alt | Roos, T. R., Roos, A. K., Avins, A. L., Ahmed, M. A., Kleimeyer, J. P., Fredericson, M., ... & Kim, S. K. (2017). Genome-wide association study identifies a locus associated with rotator cuff injury. *PLoS One*, *12*(12), e0189317. | Neither function nor reference directly point to RRI. |
| rs62051384 | WWP2, protein ubiquitination | Alt | Kim, S. K., Ioannidis, J. P., Ahmed, M. A., Avins, A. L., Kleimeyer, J. P., Fredericson, M., & Dragoo, J. L. (2018). Two genetic variants associated with plantar fascial disorders. *International journal of sports medicine*, *39*(04), 314-321. | Function doesn’t seem directly linked to injuries, but reference on common RRI. |
| rs4362400 | VAT1L, | Alt | Rodas, G., Osaba, L., Arteta, D., Pruna, R., Fernández, D., | Function doesn’t seem |

| | | | | |
|---|---|---|---|---|
| | oxidoreductase activity and zinc ion binding activity | | & Lucia, A. (2019). Genomic prediction of tendinopathy risk in elite team sports. *International Journal of Sports Physiology and Performance*, *15*(4), 489-495. | directly linked to injuries, but reference on tendinopathy. |
| rs710079 | MPG, alkylbase DNA N-glycosylase activity | Ref | Chen, S. Y., Wan, L., Huang, C. M., Huang, Y. C., Sheu, J. J. C., Lin, Y. J., ... & Tsai, F. J. (2010). Genetic polymorphisms of the DNA repair gene MPG may be associated with susceptibility to rheumatoid arthritis. *Journal of applied genetics*, *51*, 519-521. | Neither function nor reference directly point to RRI. |
| rs2858056 | MPG, alkylbase DNA N-glycosylase activity | Ref/ref | Chen, S. Y., Wan, L., Huang, C. M., Huang, Y. C., Sheu, J. J. C., Lin, Y. J., ... & Tsai, F. J. (2010). Genetic polymorphisms of the DNA repair gene MPG may be associated with susceptibility to rheumatoid arthritis. *Journal of applied genetics*, *51*, 519-521. | Neither function nor reference directly point to RRI. |
| rs2586488 | COL1A1, pro-alpha1 chains of | Ref/ref | Korvala, J., Hartikka, H., Pihlajamäki, H., Solovieva, S., Ruohola, J. P., Sahi, T., ... & Männikkö, M. (2010). Genetic | Function seems tendon-related but reference only |

| | | | | |
|---|---|---|---|---|
| | type I collagen | | predisposition for femoral neck stress fractures in military conscripts. *BMC genetics*, *11*, 1-9. | with stress fracture, too few cases in current study. |
| rs1800012 | COL1A1, pro-alpha1 chains of type I collagen | Ref | Wang, C., Li, H., Chen, K., Wu, B., & Liu, H. (2017). Association of polymorphisms rs1800012 in COL1A1 with sports-related tendon and ligament injuries: a meta-analysis. *Oncotarget*, *8*(16), 27627.<br>Leźnicka, K., Żyżniewska-Banaszak, E., Gębska, M., Machoy-Mokrzyńska, A., Krajewska-Pędzik, A., Maciejewska-Skrendo, A., & Leońska-Duniec, A. (2021). Interactions between gene variants within the COL1A1 and COL5A1 genes and musculoskeletal injuries in physically active Caucasian. *Genes*, *12*(7), 1056. | Both function and referenced injuries seem close to RRI. |
| rs820218 | SAP30BP, modulation by host of symbiont | Ref | Tashjian, R. Z., Granger, E. K., Farnham, J. M., Cannon-Albright, L. A., & Teerlink, C. C. (2016). Genome-wide association study for rotator cuff tears identifies two | Neither function nor reference directly point to RRI. |

| | | | | |
|---|---|---|---|---|
| | transcription | | significant single-nucleotide polymorphisms. *Journal of shoulder and elbow surgery*, *25*(2), 174-179. | |
| rs2277698 | TIMP2, natural inhibitors of the matrix metalloproteinases | Alt | Tashjian, R. Z., Kim, S. K., Roche, M. D., Jones, K. B., & Teerlink, C. C. (2021). Genetic variants associated with rotator cuff tearing utilizing multiple population-based genetic resources. *Journal of Shoulder and Elbow Surgery*, *30*(3), 520-531.<br>Figueiredo, E. A., Loyola, L. C., Belangero, P. S., Campos Ribeiro-dos-Santos, Â. K., Emanuel Batista Santos, S., Cohen, C., ... & Leal, M. F. (2020). Rotator cuff tear susceptibility is associated with variants in genes involved in tendon extracellular matrix homeostasis. *Journal of Orthopaedic Research®*, *38*(1), 192-201. | Function links to tissue homeostasis, but reference not with RRI. |
| rs4789932 | TIMP2, natural inhibitors of the | Ref | El Khoury, L., Posthumus, M., Collins, M., Handley, C. J., Cook, J., & Raleigh, S. M. (2013). Polymorphic variation | Function on tissue homeostasis and references |

| | | | | |
|---|---|---|---|---|
| | matrix metalloproteinases | | within the ADAMTS2, ADAMTS14, ADAMTS5, ADAM12 and TIMP2 genes and the risk of Achilles tendon pathology: a genetic association study. *Journal of science and medicine in sport*, *16*(6), 493-498.<br>El Khoury, L., Ribbans, W. J., & Raleigh, S. M. (2016). MMP3 and TIMP2 gene variants as predisposing factors for Achilles tendon pathologies: Attempted replication study in a British case–control cohort. *Meta gene*, *9*, 52-55.<br>Kim, S. K., Roos, T. R., Roos, A. K., Kleimeyer, J. P., Ahmed, M. A., Goodlin, G. T., ... & Dragoo, J. L. (2017). Genome-wide association screens for Achilles tendon and ACL tears and tendinopathy. *PloS one*, *12*(3), e0170422.<br>Kang, X., Tian, B., Zhang, L., Ge, Z., Zhao, Y., & Zhang, Y. (2019). Relationship of common variants in MPP7, TIMP2 and CASP8 genes with the risk of chronic achilles | on Achilles tendinopathy. |

| | | | | |
|---|---|---|---|---|
| | | | tendinopathy. *Scientific Reports*, *9*(1), 17627. Nie, G., Wen, X., Liang, X., Zhao, H., Li, Y., & Lu, J. (2019). Additional evidence supports association of common genetic variants in MMP3 and TIMP2 with increased risk of chronic Achilles tendinopathy susceptibility. *Journal of science and medicine in sport*, *22*(10), 1074-1078. | |
| rs3018362 | TNFRSF11A, tumor necrosis factor receptor | Ref | Varley, I., Hughes, D. C., Greeves, J. P., Stellingwerff, T., Ranson, C., Fraser, W. D., & Sale, C. (2015). RANK/RANKL/OPG pathway: genetic associations with stress fracture period prevalence in elite athletes. *Bone*, *71*, 131-136. | Function does not link to RRI and reference on stress fracture, too few cases in current study. |
| rs1800470 | TGFB1, transforming growth factor beta 1 | Alt/alt | Figueiredo, E. A., Loyola, L. C., Belangero, P. S., Campos Ribeiro-dos-Santos, Â. K., Emanuel Batista Santos, S., Cohen, C., ... & Leal, M. F. (2020). Rotator cuff tear | Neither function nor reference directly point to RRI. |

| | | | | |
|---|---|---|---|---|
| | | | susceptibility is associated with variants in genes involved in tendon extracellular matrix homeostasis. *Journal of Orthopaedic Research®*, *38*(1), 192-201. | |
| rs1800469 | TGFB1, transforming growth factor beta 1 | Alt/alt | Figueiredo, E. A., Loyola, L. C., Belangero, P. S., Campos Ribeiro-dos-Santos, Â. K., Emanuel Batista Santos, S., Cohen, C., ... & Leal, M. F. (2020). Rotator cuff tear susceptibility is associated with variants in genes involved in tendon extracellular matrix homeostasis. *Journal of Orthopaedic Research®*, *38*(1), 192-201. | Neither function nor reference directly point to RRI. |
| rs25487 | XRCC1, repair of DNA single-strand breaks | Ref | Mohamed, R. H., Amal, S., El-Shahawy, E. E., & Galil, S. M. A. (2016). Association of XRCC1 and OGG1 DNA repair gene polymorphisms with rheumatoid arthritis in Egyptian patients. *Gene*, *578*(1), 112-116. | Neither function nor reference directly point to RRI. |
| rs25489 | XRCC1, repair of DNA single-strand | Alt | Mohamed, R. H., Amal, S., El-Shahawy, E. E., & Galil, S. M. A. (2016). Association of XRCC1 and OGG1 DNA | Neither function nor reference directly point to |

| | | | | |
|---|---|---|---|---|
| | breaks | | repair gene polymorphisms with rheumatoid arthritis in Egyptian patients. *Gene*, *578*(1), 112-116. | RRI. |
| rs1045485 | CASP8, execution-phase of cell apoptosis | Alt | Nell, E. M., Van Der Merwe, L., Cook, J., Handley, C. J., Collins, M., & September, A. V. (2012). The apoptosis pathway and the genetic predisposition to Achilles tendinopathy. *Journal of Orthopaedic Research*, *30*(11), 1719-1724.<br>Kim, S. K., Roos, T. R., Roos, A. K., Kleimeyer, J. P., Ahmed, M. A., Goodlin, G. T., ... & Dragoo, J. L. (2017). Genome-wide association screens for Achilles tendon and ACL tears and tendinopathy. *PloS one*, *12*(3), e0170422. | Evidence points to Achilles tendinopathy but function does not directly link to RRI. |
| rs2289360 | EMILIN1, associates with elastic fibers at the interface between | Alt/alt | Hall, E. C., Baumert, P., Larruskain, J., Gil, S. M., Lekue, J. A., Rienzi, E., ... & Erskine, R. M. (2022). The genetic association with injury risk in male academy soccer players depends on maturity status. *Scandinavian Journal of* | Neither function nor reference directly point to RRI. |

| | | | | |
|---|---|---|---|---|
| | elastin and microfibrils | | *Medicine & Science in Sports*, *32*(2), 338-350. | |
| rs143383 | GDF5, growth differentiation factor | Alt | Zhao, L., Chang, Q., Huang, T., & Huang, C. (2016). Prospective cohort study of the risk factors for stress fractures in Chinese male infantry recruits. *Journal of International Medical Research*, *44*(4), 787-795.<br>Posthumus, M., Collins, M., Cook, J., Handley, C. J., Ribbans, W. J., Smith, R. K., ... & Raleigh, S. M. (2010). Components of the transforming growth factor-β family and the pathogenesis of human Achilles tendon pathology—a genetic association study. *Rheumatology*, *49*(11), 2090-2097.<br>Vaes, R. B. A., Rivadeneira, F., Kerkhof, J. M., Hofman, A., Pols, H. A. P., Uitterlinden, A. G., & van Meurs, J. B. J. (2009). Genetic variation in the GDF5 region is associated | Function does not seem to link to RRI but reference contains Achilles tendinopathy. |

| | | | | |
|---|---|---|---|---|
| | | | with osteoarthritis, height, hip axis length and fracture risk: the Rotterdam study. *Annals of the rheumatic diseases*, *68*(11), 1754-1760. Ge, W., Mu, J., & Huang, C. (2014). The GDF5 SNP is associated with meniscus injury and function recovery in male Chinese soldiers. *International journal of sports medicine*, *35*(07), 625-628. | |
| rs17576 | MMP9, degrades type IV and V collagens | Alt | Figueiredo, E. A., Loyola, L. C., Belangero, P. S., Campos Ribeiro-dos-Santos, Â. K., Emanuel Batista Santos, S., Cohen, C., ... & Leal, M. F. (2020). Rotator cuff tear susceptibility is associated with variants in genes involved in tendon extracellular matrix homeostasis. *Journal of Orthopaedic Research®*, *38*(1), 192-201. | Function seems relevant but reference not with RRI. |
| rs183364169 | CDCP1, tumor invasion and | Alt | Kim, S. K., Nguyen, C., Avins, A. L., & Abrams, G. D. (2021). Identification of Three Loci Associated with | Function does not seem relevant but reference with |

| | | | | |
|---|---|---|---|---|
| | metastasis | | Achilles Tendon Injury Risk from a Genome-wide Association Study. *Medicine and science in sports and exercise*, *53*(8), 1748. | Achilles tendon injury. **rs149047058 |
| rs11177 | GNL3, may be involved in tumorigenesis and stem cell proliferation | Alt | Liu, B., Cheng, H., Ma, W., Gong, F., Wang, X., Duan, N., & Dang, X. (2018). Common variants in the GNL3 contribute to the increasing risk of knee osteoarthritis in Han Chinese population. *Scientific reports*, *8*(1), 9610. | Neither function nor reference directly point to RRI. |
| rs6617 | GNL3, may be involved in tumorigenesis and stem cell proliferation | Alt | Liu, B., Cheng, H., Ma, W., Gong, F., Wang, X., Duan, N., & Dang, X. (2018). Common variants in the GNL3 contribute to the increasing risk of knee osteoarthritis in Han Chinese population. *Scientific reports*, *8*(1), 9610. | Neither function nor reference directly point to RRI. |
| rs3219008 | OGG1, excision of 8-oxoguanine, a | Ref/alt | Mohamed, R. H., Amal, S., El-Shahawy, E. E., & Galil, S. M. A. (2016). Association of XRCC1 and OGG1 DNA | Neither function nor reference directly point to |

| | | | | |
|---|---|---|---|---|
| | mutagenic base byproduct | | repair gene polymorphisms with rheumatoid arthritis in Egyptian patients. *Gene*, *578*(1), 112-116. | RRI. |
| rs13107325 | SLC39A8, cellular import of zinc at the onset of inflammation | Alt | Kim, S. K., Nguyen, C., Jones, K. B., & Tashjian, R. Z. (2021). A genome-wide association study for shoulder impingement and rotator cuff disease. *Journal of shoulder and elbow surgery*, *30*(9), 2134-2145. | Neither function nor reference directly point to RRI. |
| rs60713544 | TRIML1, function of the encoded protein has not been determined | Alt | Kim, S. K., Roos, T. R., Roos, A. K., Kleimeyer, J. P., Ahmed, M. A., Goodlin, G. T., ... & Dragoo, J. L. (2017). Genome-wide association screens for Achilles tendon and ACL tears and tendinopathy. *PloS one*, *12*(3), e0170422. | Neither function nor reference directly point to RRI. |
| rs2305948 | KDR, endothelial proliferation, survival, migration, tubular morphogenesis and | Ref/ref | Salles, J. I., Duarte, M. E. L., Guimarães, J. M., Lopes, L. R., Vilarinho Cardoso, J., Aguiar, D. P., ... & Perini, J. A. (2016). Vascular endothelial growth factor receptor-2 polymorphisms have protective effect against the development of tendinopathy in volleyball athletes. *PLoS* | Function does not seem to be relevant but reference covers lower limb tendinopathy. |

| | | | | |
|---|---|---|---|---|
| | sprouting | | *One*, *11*(12), e0167717. | |
| rs145648292 | C5orf63, open reading frame, glutaredoxin-like protein | Alt | Kim, S. K., Nguyen, C., Jones, K. B., & Tashjian, R. Z. (2021). A genome-wide association study for shoulder impingement and rotator cuff disease. *Journal of shoulder and elbow surgery*, *30*(9), 2134-2145. | Neither function nor reference directly point to RRI. |
| rs4244032 | NR3C1, glucocorticoid receptor, inflammatory responses | Ref/ref, Alt/alt | Yanovich, R., Friedman, E., Milgrom, R., Oberman, B., Freedman, L., & Moran, D. S. (2012). Candidate gene analysis in Israeli soldiers with stress fractures. *Journal of Sports Science & Medicine*, *11*(1), 147. | Function does not seem relevant and reference in stress fractures, too few cases in current study. |
| rs12656106 | NR3C1, glucocorticoid receptor, inflammatory responses | Ref/ref | Yanovich, R., Friedman, E., Milgrom, R., Oberman, B., Freedman, L., & Moran, D. S. (2012). Candidate gene analysis in Israeli soldiers with stress fractures. *Journal of Sports Science & Medicine*, *11*(1), 147. | Function does not seem relevant and reference in stress fractures, too few cases in current study. |

| | | | | |
|---|---|---|---|---|
| rs3045 | ANKH, controls pyrophosphate levels | Alt | Peach, C. A., Zhang, Y., Dunford, J. E., Brown, M. A., & Carr, A. J. (2007). Cuff tear arthropathy: evidence of functional variation in pyrophosphate metabolism genes. *Clinical Orthopaedics and Related Research (1976-2007)*, *462*, 67-72. | Neither function nor reference directly point to RRI. |
| rs187483 | ANKH, controls pyrophosphate levels | Ref/alt | Peach, C. A., Zhang, Y., Dunford, J. E., Brown, M. A., & Carr, A. J. (2007). Cuff tear arthropathy: evidence of functional variation in pyrophosphate metabolism genes. *Clinical Orthopaedics and Related Research (1976-2007)*, *462*, 67-72. | Neither function nor reference directly point to RRI. |
| rs4701616 | ANKH, controls pyrophosphate levels | Ref | Yanovich, R., Friedman, E., Milgrom, R., Oberman, B., Freedman, L., & Moran, D. S. (2012). Candidate gene analysis in Israeli soldiers with stress fractures. *Journal of Sports Science & Medicine*, *11*(1), 147. | Function does not seem relevant and reference in stress fractures, too few cases in current study. |
| rs144414988 | LSP1P3, | Alt | Kim, S. K., Nguyen, C., Jones, K. B., & Tashjian, R. Z. | Neither function nor |

| | | | | |
|---|---|---|---|---|
| | pseudogene | | (2021). A genome-wide association study for shoulder impingement and rotator cuff disease. *Journal of shoulder and elbow surgery*, *30*(9), 2134-2145. | reference directly point to RRI. |
| rs1011814 / rs900379* | FGF10, embryonic development, cell growth, morphogenesis, tissue repair, tumor growth | Alt | Salles, J. I., Amaral, M. V., Aguiar, D. P., Lira, D. A., Quinelato, V., Bonato, L. L., ... & Casado, P. L. (2015). BMP4 and FGF3 haplotypes increase the risk of tendinopathy in volleyball athletes. *Journal of science and medicine in sport*, *18*(2), 150-155. da Rocha Motta, G., Amaral, M. V., Rezende, E., Pitta, R., dos Santos Vieira, T. C., Duarte, M. E. L., ... & Casado, P. L. (2014). Evidence of genetic variations associated with rotator cuff disease. *Journal of shoulder and elbow surgery*, *23*(2), 227-235. | Function does not seem relevant but reference with lower limb tendinopathy. |
| rs11154027 | GJA1, gap junction protein | Ref | Rodas, G., Osaba, L., Arteta, D., Pruna, R., Fernández, D., & Lucia, A. (2019). Genomic prediction of tendinopathy | Function does not seem relevant but reference with |

| | | | | |
|---|---|---|---|---|
| | | | risk in elite team sports. *International Journal of Sports Physiology and Performance*, *15*(4), 489-495. | lower limb tendinopathy. |
| rs2234693 | ESR1, estrogen receptor 1 | Ref | Lian, K., Lui, L., Zmuda, J. M., Nevitt, M. C., Hochberg, M. C., Lee, J. M., ... & Lane, N. E. (2007). Estrogen receptor alpha genotype is associated with a reduced prevalence of radiographic hip osteoarthritis in elderly Caucasian women. *Osteoarthritis and Cartilage*, *15*(8), 972-978. | Estrogen seems to play a significant role in joint laxity in females, but the reference is not with RRI. |
| rs9340799 | ESR1, estrogen receptor 1 | Ref/ref | Pontin, P. A., Nogara, P. R. B., Fonseca, F. C. P., Cesar Netto, C., Carvalho, K. C., Soares Junior, J. M., ... & Godoy-Santos, A. (2018). ERα PvuII and XbaI polymorphisms in postmenopausal women with posterior tibial tendon dysfunction: a case control study. *Journal of orthopaedic surgery and research*, *13*, 1-5. | Both function and reference seem to link with potential RRI especially in females. |
| rs1643821 | ESR1, estrogen receptor 1 | Ref/ref | Dalewski, B., Kamińska, A., Białkowska, K., Jakubowska, A., & Sobolewska, E. (2020). Association of estrogen | Estrogen seems to play a significant role in joint |

| | | | | |
|---|---|---|---|---|
| | | | receptor 1 and tumor necrosis factor α polymorphisms with temporomandibular joint anterior disc displacement without reduction. *Disease Markers*, *2020*(1), 6351817. | laxity in females, but the reference is not with RRI. |
| rs1800629 | TNF, tumor necrosis factor | Alt | Furquim, B. D. A., Flamengui, L. M. S. P., Repeke, C. E. P., Cavalla, F., Garlet, G. P., & Conti, P. C. R. (2016). Influence of TNF-α-308 G/A gene polymorphism on temporomandibular disorder. *American Journal of Orthodontics and Dentofacial Orthopedics*, *149*(5), 692-698. | Neither function nor reference directly point to RRI. |
| rs2010963 | VEGFA, vascular endothelial growth factor A | Ref/ref | Hall, E. C., Baumert, P., Larruskain, J., Gil, S. M., Lekue, J. A., Rienzi, E., ... & Erskine, R. M. (2022). The genetic association with injury risk in male academy soccer players depends on maturity status. *Scandinavian Journal of Medicine & Science in Sports*, *32*(2), 338-350. | Function does not seem relevant but reference links to tendon/ligament injuries. |
| rs10484958 | SASH1, scaffold | Alt | Tashjian, R. Z., Granger, E. K., Farnham, J. M., Cannon- | Neither function nor |

| | | | | |
|---|---|---|---|---|
| | protein involved in the TLR4 signaling pathway | | Albright, L. A., & Teerlink, C. C. (2016). Genome-wide association study for rotator cuff tears identifies two significant single-nucleotide polymorphisms. *Journal of shoulder and elbow surgery*, *25*(2), 174-179. | reference directly point to RRI. |
| rs970547 | COL12A1, alpha chain of type XII collagen | Alt/alt | Bell, R. D., Shultz, S. J., Wideman, L., & Henrich, V. C. (2012). Collagen gene variants previously associated with anterior cruciate ligament injury risk are also associated with joint laxity. *Sports Health*, *4*(4), 312-318. | Both function and reference seem relevant to RRI. |
| rs4730153 | NAMPT, catalyzes the condensation of nicotinamide | Alt/alt | Wang, Y., Meng, F., Wu, J., Long, H., Li, J., Wu, Z., ... & Xie, D. (2022). Associations between adipokines gene polymorphisms and knee osteoarthritis: a meta-analysis. *BMC musculoskeletal disorders*, *23*(1), 166. | Neither function nor reference directly point to RRI. |
| rs10263021 | CNTNAP2, functions in the vertebrate nervous system as | Alt | Rodas, G., Osaba, L., Arteta, D., Pruna, R., Fernández, D., & Lucia, A. (2019). Genomic prediction of tendinopathy risk in elite team sports. *International Journal of Sports* | Function does not seem relevant but reference with tendinopathy. |

| | | | | |
|---|---|---|---|---|
| | cell adhesion molecules and receptors | | *Physiology and Performance*, *15*(4), 489-495. | |
| rs1800797 | IL6-AS1, interleukin 6, cytokine that functions in inflammation and the maturation of B cells | Alt | Li, J., Jiang, L., Zhou, X., Wu, L., Li, D., & Chen, G. (2020). The association between Interleukin-6 rs1800795/rs1800797 polymorphisms and risk of rotator cuff tear in a Chinese population. *Bioscience Reports*, *40*(4), BSR20200193. | Inflammatory function is closely linked to overuse injuries but reference is not RRI. **rs1800795 |
| rs1800795 | IL6-AS1, interleukin 6, cytokine that functions in inflammation and | Ref/ref | Hall, E. C., Baumert, P., Larruskain, J., Gil, S. M., Lekue, J. A., Rienzi, E., ... & Erskine, R. M. (2022). The genetic association with injury risk in male academy soccer players depends on maturity status. *Scandinavian Journal of Medicine & Science in Sports*, *32*(2), 338-350. | Inflammatory function is closely linked to overuse injuries and reference shows link for any (soccer) injuries and muscle injuries |

| | | | | |
|---|---|---|---|---|
| | the maturation of B cells | | Li, J., Jiang, L., Zhou, X., Wu, L., Li, D., & Chen, G. (2020). The association between Interleukin-6 rs1800795/rs1800797 polymorphisms and risk of rotator cuff tear in a Chinese population. *Bioscience Reports*, *40*(4), BSR20200193. | |
| rs1554606 | IL6, interleukin 6, cytokine that functions in inflammation and the maturation of B cells | Ref/ref | Yanovich, R., Friedman, E., Milgrom, R., Oberman, B., Freedman, L., & Moran, D. S. (2012). Candidate gene analysis in Israeli soldiers with stress fractures. *Journal of Sports Science & Medicine*, *11*(1), 147. | Inflammatory function is closely linked to overuse injuries but reference is stress fracture, too few cases in current study. **rs1800795 |
| rs2237352 / rs12700903* | CREB5, functions as a CRE-dependent trans-activator | Ref | Yanik, E. L., Keener, J. D., Lin, S. J., Colditz, G. A., Wright, R. W., Evanoff, B. A., ... & Saccone, N. L. (2021). Identification of a novel genetic marker for risk of degenerative rotator cuff disease surgery in the UK | Neither function nor reference directly point to RRI. |

| | | | | |
|---|---|---|---|---|
| | | | biobank. *JBJS*, *103*(14), 1259-1267. | |
| rs149047058 | COA1, mitochondrial cytochrome c oxidase assembly and mitochondrial respiratory chain complex I assembly | Alt | Kim, S. K., Nguyen, C., Horton, B. H., Avins, A. L., & Abrams, G. D. (2021). Association of COA1 with patellar tendonitis: a genome-wide association analysis. *Med. Sci. Sports Exerc*, *53*, 2419-2424. | Function does not seem relevant but reference with patellar tendonitis. |
| rs4725069 | GLCCI1, may be an early marker for glucocorticoid-induced apoptosis | Alt | Tashjian, R. Z., Kim, S. K., Roche, M. D., Jones, K. B., & Teerlink, C. C. (2021). Genetic variants associated with rotator cuff tearing utilizing multiple population-based genetic resources. *Journal of Shoulder and Elbow Surgery*, *30*(3), 520-531. | Neither function nor reference directly point to RRI. |
| rs12154667 | CALCR, maintaining | Ref/ref | Yanovich, R., Friedman, E., Milgrom, R., Oberman, B., Freedman, L., & Moran, D. S. (2012). Candidate gene | Both function and reference relevant to stress fracture, |

| | | | | |
|---|---|---|---|---|
| | calcium homeostasis and regulating osteoclast-mediated bone resorption | | analysis in Israeli soldiers with stress fractures. *Journal of Sports Science & Medicine*, *11*(1), 147. | too few cases in current study. |
| rs1548456 | CALCR, maintaining calcium homeostasis and regulating osteoclast-mediated bone resorption | Ref/ref | Yanovich, R., Friedman, E., Milgrom, R., Oberman, B., Freedman, L., & Moran, D. S. (2012). Candidate gene analysis in Israeli soldiers with stress fractures. *Journal of Sports Science & Medicine*, *11*(1), 147. | Both function and reference relevant to stress fracture, too few cases in current study. |
| rs420257 | COL1A2, pro-alpha2 chain of type I collagen | Ref/ref | Yanovich, R., Friedman, E., Milgrom, R., Oberman, B., Freedman, L., & Moran, D. S. (2012). Candidate gene analysis in Israeli soldiers with stress fractures. *Journal of* | Function seems relevant but reference is with stress fracture, too few cases in |

| | | | | |
|---|---|---|---|---|
| | | | *Sports Science & Medicine*, *11*(1), 147. | current study. |
| rs42517 | COL1A2, pro-alpha2 chain of type I collagen | Alt/alt | Yanovich, R., Friedman, E., Milgrom, R., Oberman, B., Freedman, L., & Moran, D. S. (2012). Candidate gene analysis in Israeli soldiers with stress fractures. *Journal of Sports Science & Medicine*, *11*(1), 147. | Function seems relevant but reference is with stress fracture, too few cases in current study. |
| rs42522 | COL1A2, pro-alpha2 chain of type I collagen | Ref/ref | Yanovich, R., Friedman, E., Milgrom, R., Oberman, B., Freedman, L., & Moran, D. S. (2012). Candidate gene analysis in Israeli soldiers with stress fractures. *Journal of Sports Science & Medicine*, *11*(1), 147. | Function seems relevant but reference is with stress fracture, too few cases in current study. |
| rs3216902 | COL1A2, pro-alpha2 chain of type I collagen | Ref | Korvala, J., Hartikka, H., Pihlajamäki, H., Solovieva, S., Ruohola, J. P., Sahi, T., ... & Männikkö, M. (2010). Genetic predisposition for femoral neck stress fractures in military conscripts. *BMC genetics*, *11*, 1-9. | Function seems relevant but reference is with stress fracture, too few cases in current study. **rs149047058 |
| rs42531 | COL1A2, pro- | Ref/ref, | Yanovich, R., Friedman, E., Milgrom, R., Oberman, B., | Function seems relevant but |

| | | | | |
|---|---|---|---|---|
| | alpha2 chain of type I collagen | Alt/alt | Freedman, L., & Moran, D. S. (2012). Candidate gene analysis in Israeli soldiers with stress fractures. *Journal of Sports Science & Medicine*, *11*(1), 147. | reference is with stress fracture, too few cases in current study. |
| rs413826 | COL1A2, pro-alpha2 chain of type I collagen | Ref/ref | Yanovich, R., Friedman, E., Milgrom, R., Oberman, B., Freedman, L., & Moran, D. S. (2012). Candidate gene analysis in Israeli soldiers with stress fractures. *Journal of Sports Science & Medicine*, *11*(1), 147. | Function seems relevant but reference is with stress fracture, too few cases in current study. |
| rs35360670 | MTSS1, cellular response to fluid shear stress | Alt | Kim, S. K., Ahmed, M. A., Avins, A. L., & Ioannidis, J. P. (2017). A genetic marker associated with De Quervain's tenosynovitis. *International journal of sports medicine*, *38*(12), 942-948. | Neither function nor reference directly point to RRI. |
| rs13317 | FGFR1, binds both acidic and basic fibroblast growth factors and is | Ref/ref | da Rocha Motta, G., Amaral, M. V., Rezende, E., Pitta, R., dos Santos Vieira, T. C., Duarte, M. E. L., ... & Casado, P. L. (2014). Evidence of genetic variations associated with rotator cuff disease. *Journal of shoulder and elbow* | Neither function nor reference directly point to RRI. |

| | | | | |
|---|---|---|---|---|
| | involved in limb induction | | *surgery*, *23*(2), 227-235. | |
| rs1800972 | DEFB1, antimicrobial peptide implicated in the resistance of epithelial surfaces to microbial colonization | Alt/alt | da Rocha Motta, G., Amaral, M. V., Rezende, E., Pitta, R., dos Santos Vieira, T. C., Duarte, M. E. L., ... & Casado, P. L. (2014). Evidence of genetic variations associated with rotator cuff disease. *Journal of shoulder and elbow surgery*, *23*(2), 227-235. | Neither function nor reference directly point to RRI. |
| rs7035322 | TNC, guidance of migrating neurons as well as axons during development, synaptic plasticity, | Ref | Kluger, R., Burgstaller, J., Vogl, C., Brem, G., Skultety, M., & Mueller, S. (2017). Candidate gene approach identifies six SNPs in tenascin-C (TNC) associated with degenerative rotator cuff tears. *Journal of Orthopaedic Research*, *35*(4), 894-901. | Neither function nor reference directly point to RRI. |

| | | | | |
|---|---|---|---|---|
| | and neuronal regeneration | | | |
| rs7021589 | TNC, guidance of migrating neurons as well as axons during development, synaptic plasticity, and neuronal regeneration | Alt | Tashjian, R. Z., Kim, S. K., Roche, M. D., Jones, K. B., & Teerlink, C. C. (2021). Genetic variants associated with rotator cuff tearing utilizing multiple population-based genetic resources. *Journal of Shoulder and Elbow Surgery*, *30*(3), 520-531. Kluger, R., Burgstaller, J., Vogl, C., Brem, G., Skultety, M., & Mueller, S. (2017). Candidate gene approach identifies six SNPs in tenascin-C (TNC) associated with degenerative rotator cuff tears. *Journal of Orthopaedic Research*, *35*(4), 894-901. | Neither function nor reference directly point to RRI. |
| rs72758637 | TNC, guidance of migrating neurons as well as axons | Alt | Tashjian, R. Z., Kim, S. K., Roche, M. D., Jones, K. B., & Teerlink, C. C. (2021). Genetic variants associated with rotator cuff tearing utilizing multiple population-based | Neither function nor reference directly point to RRI. |

| | | | | |
|---|---|---|---|---|
| | during development, synaptic plasticity, and neuronal regeneration | | genetic resources. *Journal of Shoulder and Elbow Surgery*, *30*(3), 520-531.<br>Kluger, R., Burgstaller, J., Vogl, C., Brem, G., Skultety, M., & Mueller, S. (2017). Candidate gene approach identifies six SNPs in tenascin-C (TNC) associated with degenerative rotator cuff tears. *Journal of Orthopaedic Research*, *35*(4), 894-901. | |
| rs10759753 | TNC, guidance of migrating neurons as well as axons during development, synaptic plasticity, and neuronal regeneration | Ref | Kluger, R., Burgstaller, J., Vogl, C., Brem, G., Skultety, M., & Mueller, S. (2017). Candidate gene approach identifies six SNPs in tenascin-C (TNC) associated with degenerative rotator cuff tears. *Journal of Orthopaedic Research*, *35*(4), 894-901. | Neither function nor reference directly point to RRI. |

| | | | | |
|---|---|---|---|---|
| rs2104772 | TNC, guidance of migrating neurons as well as axons during development, synaptic plasticity, and neuronal regeneration | Alt | Saunders, C. J., van der Merwe, L., Posthumus, M., Cook, J., Handley, C. J., Collins, M., & September, A. V. (2013). Investigation of variants within the COL27A1 and TNC genes and Achilles tendinopathy in two populations. *Journal of Orthopaedic Research*, *31*(4), 632-637. | Function does not seem relevant but reference shows link with Achilles tendinopathy. |
| rs1330363 | TNC, guidance of migrating neurons as well as axons during development, synaptic plasticity, and neuronal | Ref | Saunders, C. J., van der Merwe, L., Posthumus, M., Cook, J., Handley, C. J., Collins, M., & September, A. V. (2013). Investigation of variants within the COL27A1 and TNC genes and Achilles tendinopathy in two populations. *Journal of Orthopaedic Research*, *31*(4), 632-637. | Function does not seem relevant but reference shows link with Achilles tendinopathy. |

| | | | | |
|---|---|---|---|---|
| | regeneration | | | |
| rs3789870 | TNC, guidance of migrating neurons as well as axons during development, synaptic plasticity, and neuronal regeneration | Ref | Kluger, R., Burgstaller, J., Vogl, C., Brem, G., Skultety, M., & Mueller, S. (2017). Candidate gene approach identifies six SNPs in tenascin-C (TNC) associated with degenerative rotator cuff tears. *Journal of Orthopaedic Research*, *35*(4), 894-901. | Neither function nor reference directly point to RRI. |
| rs1138545 | TNC, guidance of migrating neurons as well as axons during development, synaptic plasticity, | Alt | Tashjian, R. Z., Kim, S. K., Roche, M. D., Jones, K. B., & Teerlink, C. C. (2021). Genetic variants associated with rotator cuff tearing utilizing multiple population-based genetic resources. *Journal of Shoulder and Elbow Surgery*, *30*(3), 520-531.<br>Kluger, R., Burgstaller, J., Vogl, C., Brem, G., Skultety, M., | Neither function nor reference directly point to RRI. |

| | | | | |
|---|---|---|---|---|
| | and neuronal regeneration | | & Mueller, S. (2017). Candidate gene approach identifies six SNPs in tenascin-C (TNC) associated with degenerative rotator cuff tears. *Journal of Orthopaedic Research*, *35*(4), 894-901. | |
| rs13946 | COL5A1, collagen type V alpha 1 chain | Alt | Guo, R., Ji, Z., Gao, S., Aizezi, A., Fan, Y., Wang, Z., & Ning, K. (2022). Association of COL5A1 gene polymorphisms and musculoskeletal soft tissue injuries: a meta-analysis based on 21 observational studies. *Journal of Orthopaedic Surgery and Research*, *17*(1), 129.<br>Altinisik, J., Meric, G., Erduran, M., Ates, O., Ulusal, A. E., & Akseki, D. (2015). The BstUI and DpnII variants of the COL5A1 gene are associated with tennis elbow. *The American journal of sports medicine*, *43*(7), 1784-1789. | Both function and reference seem relevant to RRI. |
| rs12722 | COL5A1, collagen type V alpha 1 | Alt | Lv, Z. T., Gao, S. T., Cheng, P., Liang, S., Yu, S. Y., Yang, Q., & Chen, A. M. (2018). Association between | Both function and reference seem relevant to RRI. |

| | | | |
|---|---|---|---|
| | chain | | polymorphism rs12722 in COL5A1 and musculoskeletal soft tissue injuries: a systematic review and meta-analysis. *Oncotarget*, *9*(20), 15365.<br>September, A. V., Cook, J., Handley, C. J., van der Merwe, L., Schwellnus, M. P., & Collins, M. (2009). Variants within the COL5A1 gene are associated with Achilles tendinopathy in two populations. *British journal of sports medicine*, *43*(5), 357-365.<br>Hall, E. C., Baumert, P., Larruskain, J., Gil, S. M., Lekue, J. A., Rienzi, E., ... & Erskine, R. M. (2022). The genetic association with injury risk in male academy soccer players depends on maturity status. *Scandinavian Journal of Medicine & Science in Sports*, *32*(2), 338-350.<br>Guo, R., Ji, Z., Gao, S., Aizezi, A., Fan, Y., Wang, Z., & Ning, K. (2022). Association of COL5A1 gene |

| | | | |
|---|---|---|---|
| | | | polymorphisms and musculoskeletal soft tissue injuries: a meta-analysis based on 21 observational studies. *Journal of Orthopaedic Surgery and Research, 17*(1), 129.<br>Dalewski, B., Białkowska, K., Pałka, Ł., Jakubowska, A., Kiczmer, P., & Sobolewska, E. (2021). COL5A1 RS12722 is associated with temporomandibular joint anterior disc displacement without reduction in polish caucasians. *Cells, 10*(9), 2423.<br>Bell, R. D., Shultz, S. J., Wideman, L., & Henrich, V. C. (2012). Collagen gene variants previously associated with anterior cruciate ligament injury risk are also associated with joint laxity. *Sports Health, 4*(4), 312-318.<br>Altinisik, J., Meric, G., Erduran, M., Ates, O., Ulusal, A. E., & Akseki, D. (2015). The BstUI and DpnII variants of the COL5A1 gene are associated with tennis elbow. *The* |

| | | | | |
|---|---|---|---|---|
| | | | *American journal of sports medicine*, *43*(7), 1784-1789. | |
| rs3196378 | COL5A1, collagen type V alpha 1 chain | Alt | September, A. V., Cook, J., Handley, C. J., van der Merwe, L., Schwellnus, M. P., & Collins, M. (2009). Variants within the COL5A1 gene are associated with Achilles tendinopathy in two populations. *British journal of sports medicine*, *43*(5), 357-365.<br>Guo, R., Ji, Z., Gao, S., Aizezi, A., Fan, Y., Wang, Z., & Ning, K. (2022). Association of COL5A1 gene polymorphisms and musculoskeletal soft tissue injuries: a meta-analysis based on 21 observational studies. *Journal of Orthopaedic Surgery and Research*, *17*(1), 129.<br>Figueiredo, E. A., Loyola, L. C., Belangero, P. S., Campos Ribeiro-dos-Santos, Â. K., Emanuel Batista Santos, S., Cohen, C., ... & Leal, M. F. (2020). Rotator cuff tear susceptibility is associated with variants in genes involved | Both function and reference seem relevant to RRI.<br>**rs12722 |

| | | | | |
|---|---|---|---|---|
| | | | in tendon extracellular matrix homeostasis. *Journal of Orthopaedic Research®*, *38*(1), 192-201. | |
| rs1134170 | COL5A1, collagen type V alpha 1 chain | Alt/alt | Abrahams, Y., Laguette, M. J., Prince, S., & Collins, M. (2013). Polymorphisms within the COL5A1 3′-UTR that alters mRNA structure and the MIR608 gene are associated with Achilles tendinopathy. *Annals of human genetics*, *77*(3), 204-214. | Both function and reference seem relevant to RRI. **rs13946 |
| rs10992075 | ROR2, early formation of the chondrocytes | Ref/alt | Yanovich, R., Friedman, E., Milgrom, R., Oberman, B., Freedman, L., & Moran, D. S. (2012). Candidate gene analysis in Israeli soldiers with stress fractures. *Journal of Sports Science & Medicine*, *11*(1), 147. | Function does not seem relevant and reference is with stress fracture, too few cases in current study. |
| rs1590 | TGFBR1, serine/threonine protein kinase | Alt | Figueiredo, E. A., Loyola, L. C., Belangero, P. S., Campos Ribeiro-dos-Santos, Â. K., Emanuel Batista Santos, S., Cohen, C., ... & Leal, M. F. (2020). Rotator cuff tear susceptibility is associated with variants in genes involved | Neither function nor reference directly point to RRI. |

| | | | | |
|---|---|---|---|---|
| | | | in tendon extracellular matrix homeostasis. *Journal of Orthopaedic Research®*, *38*(1), 192-201. | |
| rs144371252 | FRMPD4, positive regulator of dendritic spine morphogenesis and density | Alt | Kim, S. K., Nguyen, C., Jones, K. B., & Tashjian, R. Z. (2021). A genome-wide association study for shoulder impingement and rotator cuff disease. *Journal of shoulder and elbow surgery*, *30*(9), 2134-2145. | Neither function nor reference directly point to RRI. |
| rs761804508 | ACOT9, mitochondrial acyl-CoA thioesterase of unknown function | Alt | Kim, S. K., Nguyen, C., Jones, K. B., & Tashjian, R. Z. (2021). A genome-wide association study for shoulder impingement and rotator cuff disease. *Journal of shoulder and elbow surgery*, *30*(9), 2134-2145. | Neither function nor reference directly point to RRI. |
| Class1_SNP_risk_score | Combined risk score of all class 1 SNPs based on risky alleles | | | |

| | |
|---|---|
| Class12_SNP_risk_score | Combined risk score of all class 1 and 2 SNPs based on risky alleles |
| Class123_SNP_risk_score | Combined risk score of all SNPs based on risky alleles |

*These SNPs have identical genotype distributions among the study population.

**This SNP is classified as class 3 because it has >90% identical genotype information within the study population compared to another SNP, while the other SNP is considered more relevant to RRI.

[1] Separate questions were asked for number of days injured at different body regions during the past 12 months. For each region, answers >365 were assumed to be 365. Lower limb days total was the added value from all lower body regions consisting of knee, ankle/Achilles, calf/shin, foot/toes, hip/groin, and thigh (total of 6 regions).

[2] Scoring was based on the World Athletics Scoring Table 2022 version (Technical Information | Official Documents). Within the baseline questionnaire, participants were asked for their best performance during the past 6 months for 5km track, 5km road, 10km track, 10km road, half marathon, and marathon separately. For those who answered more than one questions, the highest score was chosen. For those who did not have recorded performance for any of the above (some competed at other distances, some recorded performance outside of the 6-month time frame), search was conducted on thePowerof10 website (Power of 10) for their closest scorable performance.

[3] For all male participants and for female participants <18 or >39 years of age at the date of questionnaire completion, LEAF-Q score was assumed to be 0. This is in accordance with the original LEAF-Q guidance (The LEAF questionnaire: a screening tool for the identification of female athletes at risk for the female athlete triad | British Journal of Sports Medicine).

[4] VILR and VALR were only calculated for participants whose rearfoot strikes consisted more than 70% of the total number of strikes during the running trial (Differences in kinetic variables between injured and noninjured novice runners: A prospective cohort study - ScienceDirect). Rearfoot strike was defined as any strike that presented with >1 peaks after vertical GRF were passed through a lowpass filter. A peak was defined as a point where the frame both immediately before and after it had lower vertical GRFs. A more common approach to define rearfoot runners is through visual inspection during kinematic analysis. However, since this study did not include kinematic analysis, such an approach was not viable. The region of interest was defined as the duration between landing

initiation and the first point during a foot strike where GRF > 75% BW and slope < 15 * BW/s; VILR was defined as the highest slope between 20%-100% of the region of interest; VALR was defined as the average slope between 20%-80% of the region of interest (Impact-Related Ground Reaction Forces Are More Strongly Associated With Some Running Injuries Than Others - Caleb D. Johnson, Adam S. Tenforde, Jereme Outerleys, Julia Reilly, Irene S. Davis, 2020). VALR and VILR were only calculated for rearfoot strikes, meaning that forefoot strikes from rearfoot strikers were not average in during calculation. For forefoot strikers, VILR and VALR were assumed to be 0.

[5] A step was defined as a landing phase and an ensuing flight phase. A landing phase was defined as a period of time where vertical GRF rises from 0 to above bodyweight and then returns to 0, and a flight phase is the period of time after each landing phase and before the initiation of the next landing phase.

[6] An alt_strike participant was defined as anyone whose number of rearfoot strikes was between 40%-60% of their total number of strikes. It was observed during data analysis that these participants consistently presented with 2 peaks and 1 peak during alternating strikes, meaning that they were landing with rearfoot using one leg while landing with forefoot/midfoot using the other leg. This may indicate some form of asymmetry so was included as an additional class 3 feature.

[7] Each food diary consisted of 3 days of food log, and no specification was given on how close the 3 days should be as long as they fall within the 4-month period between the participant’s 2 visits. It was recommended that the participants do 2*weekdays and

1*weekend day for each log since many people eat differently during weekdays vs. weekends. For each nutrient, the value was averaged among the 3 days within each log. The 3 dates within each food log were averaged and mapped against the Sunday of the average date's week, and the average nutrient value was placed on that week's weekly questionnaire. If the participant only completed 1 round of 3-day food diary, their nutrient value would be the averaged value for every week. If the participant completed more than 1 round of food diaries, all weeks before the first placed average nutrient value took the value of the first average nutrient value; all weeks after the last placed average nutrient value took the value of the last average nutrient value; all weeks in between 2 average nutrient values were placed on a linear slope between the previous and the next placed nutrient values (X axis time, Y axis value), and were interpolated based on the linear equation. This was to prevent sudden dramatic changes in nutritional intake values between 2 weeks.

[8] Caloric expenditure was calculated separately for every week based on the participants' answers in each weekly questionnaire. Different running speeds, strength and conditioning exercises, and non-running exercises were mapped against the Compendium of Physical Activities (Compendium of Physical Activities – Quantifying Physical Activity Energy Expenditure) for their respective METs. Participants' masses during the corresponding week and weekly MET were used to calculate total energy expenditure, which was then divided by 7 to get a daily average energy expenditure value for each week.

[9] All measurements during bone scan sessions except for height were linearly interpolated similar to nutrient values (mentioned above). Each scanning session was mapped against the Sunday during that week, and the scan/mass measurement results were placed on the weekly questionnaire during that week. All weeks prior to the first scanning session (typically none since weekly questionnaires start after the first visit) and after the last scanning session took values of the initial and the last session, respectively. Weeks between 2 sessions were mapped onto a slope drawn between the previous and the next sessions' values (X axis time, Y axis value). This was to prevent sudden dramatic change in values between 2 weeks.

[10] Thigh and lower leg regions were custom-drawn for each scan. The top point for the thigh region was defined as the horizontal extension from the anterior-superior iliac spine. Another point on the side was defined close to the horizontal extension of the greater trochanter so that the region can fully contain all thigh tissue. A middle point was defined at the tip of the coccyx, and if the tip of the coccyx was not visible, a visual approximation was used. The bottom of the thigh region/top of the lower leg region was defined as a horizontal line drawn at the knee joint space. Vertical lines were then extended to the bottom of the ~~picture~~ scan region to define the lower leg regions. The best effort was made so that the vertical lines in the middle could separate tissues from the two legs. An example of custom-defined regions is shown below:

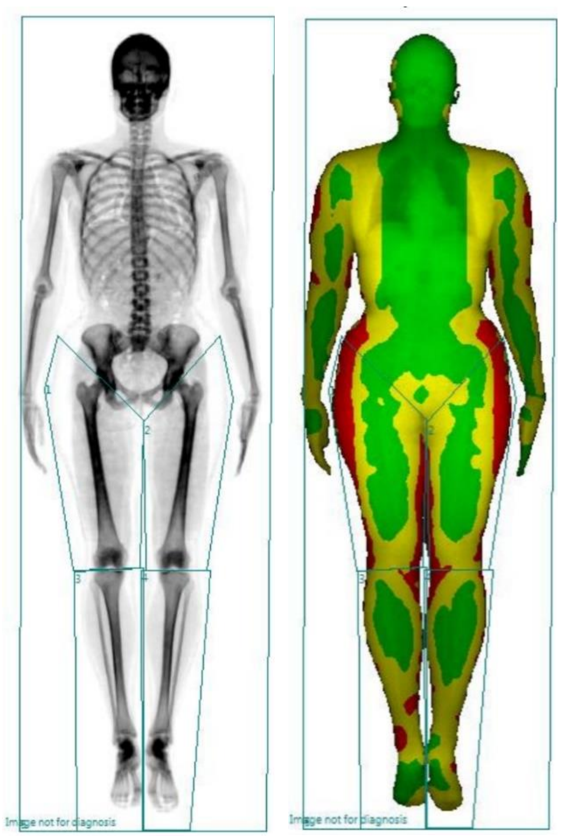

Image not for diagnosis
Image not for diagnosis

Regions 1, 2, 3, and 4 are right thigh, left thigh, right lower leg, and left lower leg, respectively.

[11] When the risky genotype is ‘ref/alt’, meaning it is heterozygotic, then genotype ‘ref/alt’ is given a risk score of 1, and ‘ref/ref’ and ‘alt/alt’ are both given a risk score of 0. Same goes the other way around (if the risky genotype is ‘ref/ref, alt/alt’, meaning both ‘ref/ref’ and ‘alt/alt’ increase risk, while ‘ref/alt’ reduces risk).

[12] When the risky genotype is ‘alt’, meaning the alternative allele increases the risk of injury, genotype ‘alt/alt’ receives a risk score of 1, genotype ‘ref/alt’ receives a risk score of 0.5, and genotype ‘ref/ref’ receives a risk score of 0. Same goes the other way around (if the risky genotype is ‘ref’).

[13] When the risky genotype is ‘ref/ref’, meaning the reference allele is recessive and increases the risk of injury, genotype ‘ref/ref’ receives a risk score of 1, while genotype ‘ref/alt’ and ‘alt/alt’ both receive a risk score of 0. Same goes the other way around (if the risky genotype is ‘alt/alt’).

**Descriptive Statistics and Data Cleaning**

**Descriptive Statistics (with notes)**

**Baseline Questionnaire**

Q4. Please select your biological sex. This refers to your physiological sex at the time of birth.

male 81

female 67

One person (male) conducted baseline testing and was non-responsive afterwards (one of the three people who had 0 weekly questionnaire responses). It was later discovered that he didn't even complete the baseline questionnaire, so the total number of participants here is 148.

Q11.1.a. (a) Have you ever been diagnosed with a bone stress injury (stress fracture, stress reaction, stress response)?

no 108

yes 40

Q14. Please indicate the location of the problem(s) you have experienced over the last 12 months and the number of days you were forced to reduce your normal running routine because of each problem.

Location choices include knee, ankle/achilles, calf/shin, hip/groin, foot/toes, thigh, lower back, and upper body. Only lower limb regions are reported here. Answers >365 days for a certain location were assumed to be 365. Answers that reported positive for a body location but did not report number of days were assumed to be 0 days.

Knee (count refers to number of people who experienced problems at the knee; descriptive statistics below refer to number of days problem lasted)

count 22.000000

mean 52.272727

std 74.892419

min 0.000000

25% 20.000000

50% 30.000000

75% 60.000000

max 365.000000

Visited healthcare professional for diagnosis?

yes 17

no 4

nan 1

Ankle/Achilles

count 34.000000

mean 43.000000

std 65.799512

min 3.000000

25% 14.000000

50% 23.000000

75% 52.000000

max 365.000000

Diagnosis?

yes 24

no 8

nan 2

Calf/shin

count 33.000000

mean 45.454545

std 65.851676

min 2.000000

25% 7.000000

50% 20.000000

75% 62.000000

max 356.000000

Diagnosis?

yes 20

no 14

The number for diagnosis here is 34 while the count above is 33. This is because one of the participants answered yes to injury but did not put in number of days affected. This resulted in that person not being counted.

Hip/groin

count 29.000000

mean 41.517241

std 72.629992

min 0.000000

25% 7.000000

50% 20.000000

75% 40.000000

max 365.000000

Diagnosis?

yes 21

no 7

nan 1

foot/toes

count 30.000000

mean 41.533333

std 60.273362

min 0.000000

25% 6.250000

50% 14.000000

75%      42.000000

max      270.000000

Diagnosis?

yes    24

no      4

nan     2

Thigh

count    12.000000

mean     34.583333

std      45.790945

min       3.000000

25%       6.500000

50%      14.000000

75%      33.000000

max      150.000000

Diagnosis?

yes    11

no 1

lower_limb_days_total (see feature description)

count 148.000000

mean 47.141892

std 90.297393

min 0.000000

25% 1.500000

50% 16.500000

75% 55.250000

max 721.000000

Number of people who had lower limb problems:

no 111

yes 37

athlete_score (see feature description)

count 148.000000

mean 686.952703

std 242.822080

min 27.000000

25% 555.500000

50% 718.000000

75% 865.250000

max 1167.000000

Q18. Typically, how many hours did you run per week over the last 12 months during the following training phases?

18.1.a. Preparatory (off-season) period: - hours per week

count 146.000000

mean 6.635616

std 2.550429

min 2.000000

25% 5.000000

50% 6.000000

75% 8.000000

max 15.000000

18.2.a. Competitive (in-season or tapering) period: - hours per week

count 139.000000

mean 6.802158

std 2.630382

min 2.000000

25% 5.000000

50% 6.500000

75% 8.250000

max 18.000000

average_run_hours (see feature description)

count 148.000000

mean 6.659797

std 2.288651

min 2.750000

25% 5.000000

50% 6.000000

75%      8.000000

max      13.500000

Q19. Typically, how often (runs per week) did you run per week over the last 12 months during the following training phases?

19.1.a. Preparatory (off-season) period: - times per week

count    145.000000

mean      5.789655

std       1.887154

min       2.500000

25%       4.500000

50%       6.000000

75%       6.000000

max      13.000000

19.2.a. Competitive (in-season or tapering) period: - times per week

count    139.000000

mean      5.917266

std 1.961216

min 2.000000

25% 5.000000

50% 6.000000

75% 6.750000

max 13.000000

average_run_frequency (see feature description)

count 146.000000

mean 5.825342

std 1.795677

min 2.500000

25% 5.000000

50% 5.500000

75% 6.250000

max 13.000000

Q20. Typically, how often have you performed intensive running sessions (speeds faster than half-marathon intensity, i.e. interval training and tempo running) per week over the last 12 months during the following training phases?

20.1.a. Preparatory (off-season) period: - times per week

count 145.000000

mean 1.858621

std 0.796484

min 0.000000

25% 1.000000

50% 2.000000

75% 2.500000

max 4.000000

20.2.a. Competitive (in-season or tapering) period: - times per week

count 139.000000

mean 2.000000

std 0.714751

min 0.000000

25% 1.750000

50% 2.000000

75% 2.250000

max 5.000000

average_interval_training_frequency (see feature description)

count 146.000000

mean 1.928082

std 0.668219

min 0.000000

25% 1.500000

50% 2.000000

75% 2.250000

max 4.500000

Age

count 148.000000

mean 31.344595

std 9.811636

min 14.000000

25% 23.000000

50% 30.000000

75% 39.250000

max 50.000000

pBPAQ: past BPAQ scores derived from questions 15 and 16

count 148.000000

mean 41.350662

std 33.871018

min 0.147000

25% 19.844875

50% 34.959000

75% 54.094250

max 210.414000

cBPAQ: current BPAQ scores derived from all exercise-related information during the past 12 months (including running, S&C, and non-running exercise questions)

count 146.000000

mean 5.083632

std 3.066868

min 1.549444

25% 3.064542

50% 4.471461

75% 6.277229

max 20.254000

tBPAQ: total BPAQ score

count 146.000000

mean 23.422187

std 17.157555

min 1.210500

25% 12.226970

50% 20.607447

75%       30.481406

max      106.285733

EDEQ metric on restraint:

| | |
|---|---|
| 0.0 | 89 |
| 1.2 | 14 |
| 0.4 | 9 |
| 0.2 | 5 |
| 0.6 | 5 |
| 3.2 | 3 |
| 0.8 | 3 |
| 1.8 | 2 |
| 4.6 | 2 |
| 1.4 | 2 |
| 1.0 | 2 |
| 2.8 | 2 |
| 2.6 | 2 |
| 2.2 | 2 |

| | |
|---|---|
| 1.6 | 2 |
| 2.4 | 2 |
| 3.4 | 1 |
| 3.0 | 1 |

EDEQ metric on eating concern:

| | |
|---|---|
| 0.0 | 94 |
| 0.2 | 25 |
| 0.4 | 9 |
| 1.2 | 4 |
| 1.0 | 3 |
| 0.6 | 3 |
| 1.6 | 2 |
| 0.8 | 2 |
| 1.4 | 2 |
| 2.4 | 1 |
| 3.6 | 1 |
| 1.8 | 1 |

| | |
|---|---|
| 3.2 | 1 |

EDEQ metric on shape concern:

| | |
|---|---|
| 0.000 | 45 |
| 0.125 | 16 |
| 0.375 | 11 |
| 0.250 | 11 |
| 0.750 | 8 |
| 0.875 | 8 |
| 0.500 | 7 |
| 0.625 | 4 |
| 1.125 | 4 |
| 1.000 | 4 |
| 1.375 | 3 |
| 1.875 | 3 |
| 1.250 | 3 |
| 2.125 | 3 |
| 1.625 | 2 |

| | |
|---|---|
| 2.625 | 2 |
| 2.875 | 2 |
| 2.500 | 2 |
| 1.750 | 1 |
| 3.625 | 1 |
| 2.000 | 1 |
| 2.250 | 1 |
| 4.250 | 1 |
| 2.375 | 1 |
| 3.000 | 1 |
| 1.500 | 1 |
| 3.750 | 1 |
| 5.500 | 1 |

EDEQ metric on weight concern:

| | |
|---|---|
| 0.0 | 66 |
| 0.6 | 16 |
| 0.2 | 15 |

| | |
|---|---|
| 0.4 | 10 |
| 0.8 | 6 |
| 1.2 | 5 |
| 1.0 | 5 |
| 1.6 | 4 |
| 1.4 | 4 |
| 2.8 | 3 |
| 2.2 | 2 |
| 1.8 | 2 |
| 2.6 | 2 |
| 3.0 | 2 |
| 2.4 | 2 |
| 2.0 | 1 |
| 3.4 | 1 |
| 3.2 | 1 |
| 6.0 | 1 |

EDEQ_total (see feature description)

count    148.000000

mean       0.535980

std        0.711912

min        0.000000

25%        0.031250

50%        0.225000

75%        0.739062

max        3.875000

LEAF-Q (see feature description)

count    46.000000

mean      8.608696

std       4.312660

min       0.000000

25%       5.250000

50%       8.000000

75%      11.000000

max      19.000000

### Isokinetic Strength

Each exercise was conducted 5 sets * 4 reps. During pilot testing it was discovered that that this setup is best to achieve familiarisation without causing unintended fatigue (see SOPs). The top 3-7 reps from the 20 reps were taken out for analysis to screen away compensation-induced outliers and familiarisation-induced underperformances.

The Isomed 2000 was not able to export data automatically, so we had to jot down data from screen, which was a painful process. Two participants were already confirmed dropouts with 0 completed weekly questionnaires after we finished baseline testing, so we decided not to jot down data from those two, resulting in a total count of 147.

hip_abduction_peak_torque

count 147.000000

mean 103.759864

std 28.764801

min 42.500000

25% 84.450000

50% 100.300000

75% 120.450000

max 205.800000

hip_abduction_peak_angle

count 147.000000

mean 5.044898

std 6.732758

min -1.800000

25% 1.000000

50% 3.000000

75% 5.800000

max 36.400000

hip_abduction_peak_torque_asymmetry

count 147.000000

mean 0.063352

std 0.050138

min 0.000000

25% 0.025288

50% 0.049945

75%        0.091118

max        0.261290

hip_abduction_peak_angle_asymmetry

count    147.000000

mean       3.771429

std        4.880602

min        0.000000

25%        1.000000

50%        2.200000

75%        4.500000

max       32.800000

hip_adduction_peak_torque

count    147.000000

mean     116.083673

std       28.470032

min       53.300000

25%     96.400000

50%    114.400000

75%    136.900000

max    200.100000

hip_adduction_peak_angle

count   147.000000

mean     25.073469

std      6.170936

min      8.500000

25%     20.900000

50%     25.000000

75%     29.400000

max     49.400000

hip_adduction_peak_torque_asymmetry

count   147.000000

mean      0.047624

std 0.044147

min 0.000789

25% 0.016510

50% 0.036551

75% 0.067725

max 0.263717

hip_adduction_peak_angle_asymmetry

count 147.000000

mean 5.118367

std 5.360165

min 0.000000

25% 2.000000

50% 4.000000

75% 6.700000

max 48.800000

total_ad_ab_ratio

| | |
|---|---|
| count | 147.000000 |
| mean | 1.147940 |
| std | 0.223227 |
| min | 0.638368 |
| 25% | 1.002757 |
| 50% | 1.136296 |
| 75% | 1.271513 |
| max | 2.019685 |

ad_ab_ratio_asymmetry

| | |
|---|---|
| count | 147.000000 |
| mean | 0.160970 |
| std | 0.140855 |
| min | 0.001860 |
| 25% | 0.055755 |
| 50% | 0.122505 |
| 75% | 0.212636 |
| max | 0.775640 |

knee_extension_peak_torque

count 147.000000

mean 137.810204

std 41.375411

min 53.400000

25% 108.950000

50% 131.800000

75% 160.150000

max 309.900000

knee_extension_peak_angle

count 147.000000

mean 62.444218

std 5.902184

min 39.600000

25% 60.150000

50% 63.200000

75% 66.550000

max 75.000000

knee_extension_peak_torque_asymmetry

count 147.000000

mean 0.046990

std 0.039311

min 0.000766

25% 0.016232

50% 0.036630

75% 0.066559

max 0.207632

knee_extension_peak_angle_asymmetry

count 147.000000

mean 3.827211

std 3.235136

min 0.000000

25%        1.300000

50%        3.400000

75%        5.700000

max       19.600000

knee_flexion_peak_torque

count    147.000000

mean      78.445578

std       22.262504

min       37.000000

25%       63.050000

50%       75.300000

75%       89.650000

max      150.700000

knee_flexion_peak_angle

count    147.000000

mean      42.397959

std 11.677830

min 19.800000

25% 33.050000

50% 40.300000

75% 50.700000

max 78.200000

knee_flexion_peak_torque_asymmetry

count 147.000000

mean 0.048856

std 0.040965

min 0.000000

25% 0.017974

50% 0.038926

75% 0.064933

max 0.226316

knee_flexion_peak_angle_asymmetry

| | |
|---|---|
| count | 147.00000 |
| mean | 6.67619 |
| std | 6.54884 |
| min | 0.00000 |
| 25% | 2.10000 |
| 50% | 4.60000 |
| 75% | 8.40000 |
| max | 34.60000 |

total_fl_ex_ratio

| | |
|---|---|
| count | 147.000000 |
| mean | 0.579159 |
| std | 0.094829 |
| min | 0.406265 |
| 25% | 0.511655 |
| 50% | 0.565865 |
| 75% | 0.633868 |
| max | 1.055556 |

fl_ex_ratio_asymmetry

count 147.000000

mean 0.076320

std 0.060792

min 0.002431

25% 0.031166

50% 0.066574

75% 0.102536

max 0.338052

### Biomechanics

navicular_drop

count 149.000000

mean 0.503356

std 0.223203

min 0.000000

25% 0.350000

50%        0.500000

75%        0.600000

max        1.300000

navicular_drop_asymmetry

count    149.000000

mean       0.170470

std        0.144495

min        0.000000

25%        0.100000

50%        0.100000

75%        0.200000

max        0.800000

Q_angle

count    149.000000

mean      13.238255

std        4.361062

min 3.500000

25% 10.000000

50% 13.000000

75% 16.000000

max 26.500000

Q_angle_asymmetry

count 149.000000

mean 3.496644

std 2.808347

min 0.000000

25% 1.000000

50% 3.000000

75% 5.000000

max 12.000000

The instrumented treadmill lab experienced a power outage one afternoon during baseline testing, which resulted in 3 participants not having their GRF measured. 1 of the 3 participants took the treadmill test during the second visit, and unfortunately the other 2 participants dropped out soon after the baseline test and did not conduct revisits (they contributed to only a few valid samples). As a result, the total number of measured cases is 147.

VILR_10

0-value (mid/forefoot strikers) 78

Non-0 value (rearfoot strikers) 69

count 69.000000

mean 80.156836

std 11.177799

min 54.461105

25% 72.000725

50% 81.030558

75% 88.320589

max 105.574642

VALR_10

count 69.000000

mean 70.400512

std 9.807378

min 48.014808

25% 63.551402

50% 70.346940

75% 78.247243

max 91.422770

VILR_asymmetry_10

count 69.000000

mean 0.043977

std 0.046565

min 0.000466

25% 0.013921

50% 0.026364

75% 0.067045

max 0.265231

VALR_asymmetry_10

count 69.000000

mean 0.043376

std 0.045925

min 0.000238

25% 0.010744

50% 0.024928

75% 0.065101

max 0.250974

Impact_peak_10

count 147.000000

mean 2.529939

std 0.226041

min 2.096892

25% 2.379742

50% 2.496879

75% 2.649728

max 3.239484

Impact_peak_asymmetry_10

count 147.000000

mean 0.024762

std 0.021309

min 0.000030

25% 0.008877

50% 0.019963

75% 0.034403

max 0.102915

Flight_time_10

count 147.000000

mean 0.122376

std 0.022665

min 0.063815

25% 0.107787

50%        0.122908

75%        0.137732

max        0.182038

Contact_time_10

count    147.000000

mean       0.240310

std        0.020574

min        0.189757

25%        0.225443

50%        0.242816

75%        0.255873

max        0.302865

Duty_factor_10

count    147.000000

mean       0.663467

std        0.055179

min 0.510875

25% 0.629320

50% 0.666804

75% 0.696739

max 0.812416

Step_frequency_10

count 147.000000

mean 165.923966

std 9.275687

min 149.582796

25% 158.910724

50% 164.568391

75% 170.460520

max 210.009087

Cadence_asymmetry_10

count 147.000000

mean 0.016345

std 0.012310

min 0.000236

25% 0.006879

50% 0.013935

75% 0.021913

max 0.059104

Duty_factor_asymmetry_10

count 147.000000

mean 0.019227

std 0.016625

min 0.000469

25% 0.006047

50% 0.015929

75% 0.027818

max 0.096717

VILR_12

0-value (mid/forefoot strikers) 68

Non-0 value (rearfoot strikers) 79

count 79.000000

mean 92.260413

std 12.632410

min 62.304330

25% 84.046500

50% 92.657439

75% 100.807154

max 125.648040

VALR_12

count 79.000000

mean 80.847378

std 11.090926

min 55.289046

25% 73.606304

50%       80.752109

75%       88.941514

max      108.596942

VILR_asymmetry_12

count    79.000000

mean      0.054029

std       0.049525

min       0.000477

25%       0.014296

50%       0.043468

75%       0.083976

max       0.229408

VALR_asymmetry_12

count    79.000000

mean      0.056534

std       0.049791

min 0.000014

25% 0.014666

50% 0.046524

75% 0.084459

max 0.227726

Impact_peak_12

count 147.000000

mean 2.660064

std 0.229354

min 2.196389

25% 2.513279

50% 2.630061

75% 2.793848

max 3.356592

Impact_peak_asymmetry_12

count 147.000000

mean 0.025228

std 0.021321

min 0.000219

25% 0.010361

50% 0.019979

75% 0.033967

max 0.098902

Flight_time_12

count 147.000000

mean 0.133772

std 0.021307

min 0.081044

25% 0.121267

50% 0.133884

75% 0.148289

max 0.187942

Contact_time_12

count 147.000000

mean 0.219458

std 0.017421

min 0.179415

25% 0.208446

50% 0.220825

75% 0.229347

max 0.272302

Duty_factor_12

count 147.000000

mean 0.622397

std 0.049367

min 0.498203

25% 0.590601

50% 0.623202

75% 0.647293

max 0.754855

Step_frequency_12

count 147.000000

mean 170.412270

std 9.908495

min 150.692782

25% 163.070664

50% 169.899620

75% 175.408986

max 214.603860

Cadence_asymmetry_12

count 147.000000

mean 0.016256

std 0.012239

min 0.000102

25% 0.006550

50%        0.013651

75%        0.022313

max        0.064059

Duty_factor_asymmetry_12

count    147.000000

mean       0.018393

std        0.015465

min        0.000003

25%        0.006382

50%        0.012816

75%        0.027468

max        0.074562

Alt_strike

For this feature, 0 means negative and 1 means positive.

0    127

1     20

### Nutrition

Within the machine learning models, nutritional data was averaged for every 3-day diary and was linearly extrapolated to fit every week's value (see feature descriptions). Body weight information was also linearly extrapolated based on weight measurements during visits and was used to calculate some of the nutritional features. Descriptive statistics cannot account for these operations. Here the statistics are for all person-days of food diaries without accounting for bodyweight.

protein_intake

count 698.000000

mean 109.650798

std 40.240483

min 9.599000

25% 81.172350

50% 105.757050

75% 133.029300

max 283.786600

fat_intake

count 698.000000

mean 92.010683

std 37.739716

min 13.616000

25% 64.804125

50% 86.700600

75% 110.276125

max 315.397100

energy_intake (KJ)

count 698.000000

mean 10145.795313

std 3141.288727

min 1804.484000 (intermittent fasting day)

25% 8203.458925

50% 9659.379500

75% 11840.562050

max 26937.526400

omega3_intake

count 673.000000

mean 6.290894

std 44.066484

min 0.000600

25% 0.163500

50% 0.505600

75% 1.450000

max 483.262500

vitaminD_intake

count 663.000000

mean 9.382870

std 29.638041

min 0.000400

25% 0.846900

50% 2.882500

75%        6.816700

max      500.691000

vitaminC_intake

count     692.000000

mean      183.310866

std       525.205806

min         0.000100

25%        34.183000

50%        86.019750

75%       166.292850

max      8755.200000

vitaminE_intake

count     694.000000

mean       10.395129

std        40.211375

min         0.007500

25%         3.184225

50%         6.286100

75%        10.142800

max      1016.081700

calcium_intake

count     696.000000

mean      716.855899

std       449.976630

min         1.125000

25%       391.310825

50%       640.252050

75%       952.681975

max      2793.489300

copper_intake

count    695.000000

mean       1.148538

std 0.881791

min 0.003100

25% 0.628200

50% 0.941700

75% 1.434350

max 8.237600

iron_intake

count 694.000000

mean 15.537530

std 30.022110

min 0.000300

25% 5.870750

50% 9.076600

75% 13.779975

max 213.344800

glycine_intake

count 680.000000

mean 1333.324441

std 1290.429847

min 1.350000

25% 387.107900

50% 955.916950

75% 1981.851425

max 11186.658000

arginine_intake

count 680.000000

mean 1899.037696

std 1780.752333

min 2.430000

25% 544.349850

50% 1437.972400

75% 2809.234175

max 15468.900000

fat_percentage

count    698.000000

mean      33.852067

std        7.621479

min       14.525690

25%       28.781698

50%       34.280014

75%       38.737028

max       62.124885

More nutrient data can be exported through Nutritics. The above only contains information that pertains to the feature pool of this study. If you are interested in descriptive statistics of other nutrients, you can contact us for them.

### Anthropometry and Bone Scans

Anthropometry and bone scan (DXA and pQCT) data were collected together at each bone scan session. For these measurements, each participant's data is averaged across sessions to produce descriptive statistics.

Height

| | |
|---|---|
| count | 149.000000 |
| mean | 173.163087 |
| std | 8.986276 |
| min | 146.400000 |
| 25% | 165.600000 |
| 50% | 174.000000 |
| 75% | 178.700000 |
| max | 191.100000 |

Mass

| | |
|---|---|
| count | 149.000000 |
| mean | 64.787919 |
| std | 11.098039 |
| min | 41.800000 |
| 25% | 55.900000 |
| 50% | 64.500000 |
| 75% | 71.900000 |

max 105.700000

DXA and pQCT

L1 BMD

count 149.000000

mean 1.103886

std 0.127660

min 0.838674

25% 1.012741

50% 1.092006

75% 1.188871

max 1.583956

L2 BMD

count 149.000000

mean 1.217313

std 0.137320

min 0.937092

25%        1.117335

50%        1.207081

75%        1.297864

max        1.608861

L3 BMD

count    149.000000

mean       1.243875

std        0.140121

min        1.011572

25%        1.131937

50%        1.238896

75%        1.342449

max        1.593036

L4 BMD

count    149.000000

mean       1.186651

std 0.136850

min 0.936064

25% 1.089068

50% 1.170211

75% 1.291911

max 1.555480

L1-L4 BMD

count 149.000000

mean 1.190123

std 0.129939

min 0.943225

25% 1.091423

50% 1.183298

75% 1.275761

max 1.546079

L1 BMC

count 149.000000

mean 14.480363

std 2.812467

min 8.789660

25% 12.380159

50% 14.090760

75% 16.507135

max 22.578841

L2 BMC

count 149.000000

mean 16.987467

std 3.292715

min 10.345271

25% 14.678363

50% 16.286943

75% 19.326917

max 25.620317

L3 BMC

count 149.000000

mean 19.498225

std 3.686093

min 12.254614

25% 16.851381

50% 18.979872

75% 22.211820

max 29.102868

L4 BMC

count 149.000000

mean 20.818285

std 3.848734

min 11.010394

25% 18.081004

50% 20.335205

75%       23.052864

max       32.514064

L1-L4 BMC

count    149.000000

mean      71.784341

std       13.275408

min       45.189680

25%       62.171194

50%       70.020337

75%       81.158485

max      105.421256

Femoral neck BMD

count    149.000000

mean       1.103437

std        0.143146

min        0.803729

25% 0.994950

50% 1.089596

75% 1.185979

max 1.479664

Femoral wards BMD

count 149.000000

mean 0.963785

std 0.174452

min 0.545895

25% 0.851556

50% 0.961300

75% 1.079881

max 1.367406

Trochanteric region BMD

count 149.000000

mean 0.911293

std 0.139615

min 0.592352

25% 0.819209

50% 0.897350

75% 1.016837

max 1.240481

Femoral Shaft BMD

count 149.000000

mean 1.351958

std 0.164420

min 0.982813

25% 1.235216

50% 1.344824

75% 1.463925

max 1.768130

Femoral total BMD

| | |
|---|---|
| count | 149.000000 |
| mean | 1.129665 |
| std | 0.140324 |
| min | 0.854704 |
| 25% | 1.023756 |
| 50% | 1.127222 |
| 75% | 1.221423 |
| max | 1.450660 |

Femoral neck BMC

| | |
|---|---|
| count | 149.000000 |
| mean | 5.640289 |
| std | 0.947395 |
| min | 3.790915 |
| 25% | 4.875273 |
| 50% | 5.637317 |
| 75% | 6.301275 |
| max | 8.489792 |

Femoral wards BMC

count 149.000000

mean 2.812164

std 0.725507

min 1.416662

25% 2.237707

50% 2.728359

75% 3.277907

max 5.339239

Trochanteric region BMC

count 149.000000

mean 13.118812

std 3.362207

min 5.325329

25% 10.508588

50% 13.034139

75%       15.436450

max       21.906373

Femoral shaft BMC

count    149.000000

mean      19.662536

std       2.614601

min       14.188957

25%       17.670427

50%       19.382218

75%       21.966693

max       26.982843

Femoral total BMC

count    149.000000

mean      38.421637

std       6.634774

min       23.439122

25% 33.372112

50% 37.707752

75% 43.535966

max 57.193303

Thigh lean mass (g)

count 149.000000

mean 14714.827508

std 2815.428379

min 9253.328802

25% 12158.941882

50% 14732.884040

75% 16927.536743

max 22299.326833

Lower leg lean mass (g)

| | |
|---|---|
| count | 149.000000 |
| mean | 5467.032415 |
| std | 1094.484516 |
| min | 3578.300239 |
| 25% | 4616.578622 |
| 50% | 5493.744239 |
| 75% | 6345.075239 |
| max | 8668.302020 |

Thigh fat mass (g)

| | |
|---|---|
| count | 149.000000 |
| mean | 3118.946294 |

| | |
|---|---|
| std | 1364.992793 |
| min | 790.690781 |
| 25% | 1944.230308 |
| 50% | 2876.375102 |
| 75% | 3898.558002 |
| max | 7237.194256 |

Lower leg fat mass (g)

| | |
|---|---|
| count | 149.000000 |
| mean | 1474.068251 |
| std | 341.423088 |
| min | 730.588959 |
| 25% | 1264.662254 |

50%     1439.270001

75%     1699.617029

max     2725.229287

Total body lean mass (g)

count     149.000000

mean     51649.937402

std     9122.384942

min     35369.183750

25%     43068.685776

50%     52496.553487

75%     59372.665748

max     72248.785164

Total body fat mass (g)

count     149.000000

mean     10761.344071

std 4158.225075

min 3624.260970

25% 8051.307879

50% 9900.102065

75% 12354.355978

max 28719.458746

Total body bone mass (g)

count 149.000000

mean 2850.343985

std 472.431968

min 1910.292401

25% 2431.552345

50% 2849.110164

75% 3224.778852

max 4005.035576

Total body composition

| | |
|---|---|
| count | 149.000000 |
| mean | 0.164813 |
| std | 0.052627 |
| min | 0.073684 |
| 25% | 0.122821 |
| 50% | 0.159491 |
| 75% | 0.198379 |
| max | 0.329868 |

Muscle cross sectional area at calf (66% tibial length distal to proximal)

| | |
|---|---|
| count | 147.000000 |
| mean | 7706.286848 |
| std | 1351.894855 |
| min | 5124.062500 |
| 25% | 6739.906250 |
| 50% | 7640.937500 |
| 75% | 8638.250000 |
| max | 11631.375000 |

### Training and Injuries

Each participant's weekly values are averaged to get a personal average running volume throughout the tracking period.

Weekly running volume (min)

count 146.000000

mean 285.089845

std 130.642426

min 21.600000

25% 187.104582

50% 285.697154

75% 365.992134

max 666.051373

Weekly running volume at low intensity (min)

count 146.000000

mean 228.201052

std 122.515206

min 14.480299

25% 129.444911

50% 227.912437

75% 305.274090

max 585.069101

Weekly running volume at moderate intensity (min)

count 146.000000

mean 35.340877

std 28.395151

min 0.000000

25% 17.079176

50% 31.881355

75% 43.768844

max 222.081209

Weekly running volume at high intensity (min)

count 146.000000

| | |
|---|---|
| mean | 18.677303 |
| std | 12.573574 |
| min | 0.000000 |
| 25% | 9.714171 |
| 50% | 16.935444 |
| 75% | 24.757881 |
| max | 60.000000 |

Weekly running volume at very high intensity (min)

| | |
|---|---|
| count | 146.000000 |
| mean | 2.870612 |
| std | 4.299868 |
| min | 0.000000 |
| 25% | 0.189651 |
| 50% | 1.237273 |
| 75% | 3.519767 |
| max | 22.117201 |

Weekly resistance training volume (min)

count 146.000000

mean 36.847702

std 36.140562

min 0.000000

25% 9.311622

50% 27.022007

75% 51.610577

max 172.538462

Weekly bodyweight exercises volume (min)

count 146.000000

mean 10.191470

std 13.486219

min 0.000000

25% 0.907491

50% 4.266667

75% 15.942308

max 72.500000

Weekly core stability exercises volume (min)

count 146.000000

mean 13.170576

std 21.739545

min 0.000000

25% 1.548583

50% 7.532145

75% 16.053092

max 180.000000

Weekly balance training volume (min)

count 146.000000

mean 1.790701

std 6.557516

min 0.000000

25% 0.000000

50%        0.000000

75%        0.988095

max       67.962963

Weekly plyometrics volume (min)

count    146.000000

mean       3.189966

std        6.331710

min        0.000000

25%        0.000000

50%        0.489804

75%        3.053750

max       38.557692

Weekly drills volume (min)

count    146.000000

mean       5.265703

std        9.311122

min 0.000000

25% 0.000000

50% 0.444314

75% 6.041166

max 45.000000

Weekly circuit training volume (min)

count 146.000000

mean 2.287055

std 11.093199

min 0.000000

25% 0.000000

50% 0.000000

75% 0.179467

max 95.000000

Weekly barefoot exercises volume (min)

count 146.000000

mean 1.950165

std 8.388745

min 0.000000

25% 0.000000

50% 0.000000

75% 0.076019

max 80.000000

Weekly stretching volume (min)

count 146.000000

mean 22.413061

std 31.556338

min 0.000000

25% 0.000000

50% 10.283019

75% 31.690668

max 184.915254

Number of injured weeks experienced

count 142.000000

mean 3.971831

std 4.679734

min 0.000000

25% 0.250000

50% 3.000000

75% 6.000000

max 27.000000

Number of injured weeks for every individual:

[ 0 2 6 0 4 0 24 2 1 0 3 8 0 1 1 1 0 0 9 6 2 0 3 6

6 5 2 1 0 1 7 5 0 4 4 7 0 9 4 5 8 7 5 8 0 2 9 4

5 0 0 11 2 7 9 2 7 4 8 3 3 4 3 2 2 4 16 3 3 4 1 0

0 2 6 2 1 19 11 5 6 2 3 8 1 11 5 1 6 14 2 0 1 7 27 0

0 5 3 1 0 0 1 4 0 6 0 0 0 0 1 0 11 4 0 0 4 0 4 12

1 2 16 3 14 0 1 2 11 0 3 1 4 11 0 0 0 0 1 6 4 3]

Number of injured weeks divided by total number of weeks for that participant

| | |
|---|---|
| count | 142.000000 |
| mean | 0.101129 |
| std | 0.146392 |
| min | 0.000000 |
| 25% | 0.003521 |
| 50% | 0.063859 |
| 75% | 0.127201 |
| max | 1.000000 |

Note: max of 1 refers to an individual who got injured right at the beginning of the study and soon dropped out, leaving only a few injured weeks.

### Data Imputation

**N/A values are filled with '0'**: ['VILR_10', 'VALR_10', "VILR_asymmetry_10", "VALR_asymmetry_10", "VILR_12", "VALR_12", "VILR_asymmetry_12", "VALR_asymmetry_12", 'Alt_strike', "past_week_ratio", "past_month_ratio", "past_week_ratio_low", "past_month_ratio_low", "past_week_ratio_moderate", "past_month_ratio_moderate", "past_week_ratio_high", "past_month_ratio_high", "past_week_ratio_very_high", "past_month_ratio_very_high", "past_week_ratio_calculated_volume", "past_month_ratio_calculated_volume"]

Rationale: VALR and VILR data are predicated on the participant being a rearfoot striker, so it would not be appropriate to assume a positive value for a participant whose strike pattern is unknown. Alt_strike is a rare pattern identified only on a handful of participants, so it would be appropriate to assume 0 on anyone unknown. For ratio data, nan often refers to division by 0 or unavailable data (the second weekly questionnaire where data from 2 weeks prior is not available), and were thus filled with 0.

**N/A values are filled with the median of all available values**: ["fat_intake_avg", "fat_intake_BW", "fat_percentage_avg", "average_energy_availability", "protein_intake_BW", "omega3_intake_BW", "vitaminD_intake_BW", "vitaminC_intake_BW", "vitaminE_intake_BW", "calcium_intake_BW", "copper_intake_BW", "iron_intake_BW", "glycine_intake_BW", "arginine_intake_BW", "class1_SNP_risk_score", "class12_SNP_risk_score", "class123_SNP_risk_score"]

Rationale: Food intake data is assumed to be the median among all other participants. Risk scores calculated via groups of SNPs are continuous and are thus suitable for median imputation.

**N/A values are filled with the mean of all available values**: ["rs3753841", "rs7528684", "rs57104447", "rs1887632", "rs4654760", "rs1137101", "rs4919510", "rs1937810", "rs6481512", "rs1249269", "rs11225395", "rs1144393", "rs650108", "rs591058", "rs2252070", "rs2306033", "rs2277268", "rs4988321", "rs12574452", "rs11232681", "rs1718119", "rs3751143", "rs1544410", "rs2228570", "rs4328262", "rs1021188", "rs74544784", "rs12429486", "rs78391032", "rs77569527", "rs117544024", "rs4454832", "rs912336", "rs3218791", "rs2761884", "rs4986938", "rs911263", "rs2525504", "rs17756404", "rs4903399", "rs10132091", "rs17583842", "rs1676303", "rs11629171", "rs2281518", "rs2285053", "rs71404070", "rs62051384", "rs4362400", "rs710079", "rs2858056", "rs2586488", "rs1800012", "rs820218", "rs2277698", "rs4789932", "rs3018362", "rs1800470", "rs1800469", "rs25487", "rs25489", "rs1045485", "rs2289360", "rs143383", "rs17576", "rs183364169", "rs11177", "rs6617", "rs3219008", "rs13107325", "rs60713544", "rs2305948", "rs145648292", "rs4244032", "rs12656106", "rs3045", "rs187483", "rs4701616", "rs144414988", "rs1011814", "rs11154027", "rs2234693", "rs9340799", "rs1643821", "rs1800629", "rs2010963", "rs10484958", "rs970547", "rs4730153", "rs10263021", "rs1800797", "rs1800795", "rs1554606", "rs2237352", "rs149047058", "rs4725069", "rs12154667", "rs1548456", "rs420257", "rs42517", "rs42522", "rs3216902", "rs42531", "rs413826", "rs35360670", "rs13317", "rs1800972", "rs7035322", "rs7021589", "rs72758637", "rs10759753", "rs2104772", "rs1330363", "rs3789870", "rs1138545", "rs13946", "rs12722", "rs3196378", "rs1134170", "rs10992075", "rs1590", "rs144371252", "rs761804508"]

Rationale: Individual SNP information is not continuous and typically adapt 0/1 or 0/0.5/1 values. As a result, the mean would better reflect the total risk expectancy provided that the individual's information is not available.

**N/A values are filled with the median of all other available values from participants of the same sex**: ["hip_abduction_peak_torque", "hip_abduction_peak_angle", "hip_abduction_peak_torque_asymmetry", "hip_abduction_peak_angle_asymmetry", "hip_adduction_peak_torque", "hip_adduction_peak_angle", "hip_adduction_peak_torque_asymmetry", "hip_adduction_peak_angle_asymmetry", "total_ad_ab_ratio", "ad_ab_ratio_asymmetry", "knee_extension_peak_torque", "knee_extension_peak_angle", "knee_extension_peak_torque_asymmetry", "knee_extension_peak_angle_asymmetry", "knee_flexion_peak_torque", "knee_flexion_peak_angle", "knee_flexion_peak_torque_asymmetry", "knee_flexion_peak_angle_asymmetry", "total_fl_ex_ratio", "fl_ex_ratio_asymmetry", "Impact_peak_10", "Impact_peak_asymmetry_10", "Flight_time_10", "Contact_time_10", "Duty_factor_10", "Step_frequency_10", "Cadence_asymmetry_10", "Duty_factor_asymmetry_10", "Impact_peak_12", "Impact_peak_asymmetry_12", "Flight_time_12", "Contact_time_12", "Duty_factor_12", "Step_frequency_12", "Cadence_asymmetry_12", "Duty_factor_asymmetry_12", "calf_size"]

Rationale: For any features related to strength/muscle size/bodyweight, there is expected to be a distinct sex difference, so imputation is conducted using the median of all other same-sex participants.

### Time-sequenced Feature Categories

**Genotype**: ['rs11225395', 'rs1144393', 'rs650108', 'rs591058', 'rs2252070', 'rs4986938', 'rs1800012', 'rs4789932', 'rs9340799', 'rs970547', 'rs1800795', 'rs13946', 'rs12722', 'class1_SNP_risk_score', 'rs7528684', 'rs4919510', 'rs1937810', 'rs6481512', 'rs1249269', 'rs12574452', 'rs12429486', 'rs4454832', 'rs2761884', 'rs62051384', 'rs4362400', 'rs2586488', 'rs2277698', 'rs1045485', 'rs143383', 'rs17576', 'rs2305948', 'rs1011814', 'rs11154027', 'rs2234693', 'rs1643821', 'rs2010963', 'rs10263021', 'rs149047058', 'rs420257', 'rs42517', 'rs42522', 'rs42531', 'rs413826', 'rs2104772', 'rs1330363', 'class12_SNP_risk_score', 'rs3753841', 'rs57104447', 'rs1887632', 'rs4654760', 'rs1137101', 'rs2306033', 'rs2277268', 'rs4988321', 'rs11232681', 'rs1718119', 'rs3751143', 'rs1544410', 'rs2228570', 'rs4328262', 'rs1021188', 'rs74544784', 'rs78391032', 'rs77569527', 'rs117544024', 'rs912336', 'rs3218791', 'rs911263', 'rs2525504', 'rs17756404', 'rs4903399', 'rs10132091', 'rs17583842', 'rs1676303', 'rs11629171', 'rs2281518', 'rs2285053', 'rs71404070', 'rs710079', 'rs2858056', 'rs820218', 'rs3018362', 'rs1800470', 'rs1800469', 'rs25487', 'rs25489', 'rs2289360', 'rs183364169', 'rs11177', 'rs6617', 'rs3219008', 'rs13107325', 'rs60713544', 'rs145648292', 'rs4244032', 'rs12656106', 'rs3045', 'rs187483', 'rs4701616', 'rs144414988', 'rs1800629', 'rs10484958', 'rs4730153', 'rs1800797', 'rs1554606', 'rs2237352', 'rs4725069', 'rs12154667', 'rs1548456', 'rs3216902', 'rs35360670', 'rs13317', 'rs1800972', 'rs7035322', 'rs7021589', 'rs72758637', 'rs10759753', 'rs3789870', 'rs1138545', 'rs3196378', 'rs1134170', 'rs10992075', 'rs1590', 'rs144371252', 'rs761804508', 'class123_SNP_risk_score', 'sex']

History: ['Age', 'lower_limb_days_total', 'average_run_hours', 'average_interval_training_frequency', 'EDEQ_total', 'tracking_period_injury', 'past_stress_injury', 'LEAF-Q', 'Athlete_Score', 'average_run_frequency', 'past_month_injury']

**Phenotype**: ['hip_abduction_peak_torque', 'total_ad_ab_ratio', 'knee_extension_peak_torque', 'knee_flexion_peak_torque', 'navicular_drop', 'navicular_drop_asymmetry', 'Q_angle', 'Q_angle_asymmetry', 'VALR_12', 'Impact_peak_12', 'Duty_factor_12', 'BMI', 'BMD_spine', 'hip_abduction_peak_torque_asymmetry', 'hip_adduction_peak_torque', 'hip_adduction_peak_torque_asymmetry', 'knee_extension_peak_torque_asymmetry', 'knee_flexion_peak_torque_asymmetry', 'total_fl_ex_ratio', 'leg_lean_mass', 'hip_abduction_peak_angle', 'hip_abduction_peak_angle_asymmetry', 'hip_adduction_peak_angle', 'hip_adduction_peak_angle_asymmetry', 'ad_ab_ratio_asymmetry', 'knee_extension_peak_angle', 'knee_extension_peak_angle_asymmetry', 'knee_flexion_peak_angle', 'knee_flexion_peak_angle_asymmetry', 'fl_ex_ratio_asymmetry', 'VILR_10', 'VALR_10', 'VILR_asymmetry_10', 'VALR_asymmetry_10', 'Impact_peak_10', 'Impact_peak_asymmetry_10', 'Flight_time_10', 'Contact_time_10', 'Duty_factor_10', 'Step_frequency_10', 'Cadence_asymmetry_10', 'Duty_factor_asymmetry_10', 'VILR_12', 'VILR_asymmetry_12', 'VALR_asymmetry_12', 'Impact_peak_asymmetry_12', 'Flight_time_12', 'Contact_time_12', 'Step_frequency_12', 'Cadence_asymmetry_12',

'Duty_factor_asymmetry_12', 'Alt_strike', 'height', 'Mass', 'thigh_lean_mass', 'thigh_ffmi', 'lower_leg_lean_mass', 'lower_leg_ffmi', 'leg_ffmi', 'total_lean_mass', 'total_ffmi', 'calf_size', 'BMD_hip', 'BMD_body']

**Behaviour**: ['fat_intake_avg', 'past_month_distance', 'past_month_ratio', 'SC_past_season', 'non_running_past_season', 'fat_intake_BW', 'fat_percentage_avg', 'average_energy_availability', 'protein_intake_BW', 'omega3_intake_BW', 'vitaminD_intake_BW', 'vitaminC_intake_BW', 'vitaminE_intake_BW', 'calcium_intake_BW', 'copper_intake_BW', 'iron_intake_BW', 'glycine_intake_BW', 'arginine_intake_BW', 'past_month_min', 'past_week_ratio', 'past_month_volume_low', 'past_week_ratio_low', 'past_month_ratio_low', 'past_month_volume_moderate', 'past_week_ratio_moderate', 'past_month_ratio_moderate', 'past_month_volume_high', 'past_week_ratio_high', 'past_month_ratio_high', 'past_month_volume_very_high', 'past_week_ratio_very_high', 'past_month_ratio_very_high', 'past_month_calculated_volume', 'past_week_ratio_calculated_volume', 'past_month_ratio_calculated_volume', 'resistance_training_past_month', 'resistance_training_past_season', 'bodyweight_exercises_past_month', 'bodyweight_exercises_past_season', 'core_stability_past_month', 'core_stability_past_season', 'balance_training_past_month', 'balance_training_past_season', 'plyometrics_past_month', 'plyometrics_past_season', 'drills_past_month', 'drills_past_season', 'circuit_training_past_month', 'circuit_training_past_season', 'barefoot_past_month', 'barefoot_past_season', 'stretching_past_month', 'stretching_past_season', 'SC_past_month',

'non_running_past_month]

### Custom Hyperparameters Description

**TSNN**

Feature attention

Code:

```
class FeatureAttention(nn.Module):
    def __init__(self, feature_dim):
        super(FeatureAttention, self).__init__()
        self.attention = nn.Sequential(
            nn.Linear(feature_dim, max(1, feature_dim // 2)),
            nn.ReLU(),
            nn.Linear(max(1, feature_dim // 2), feature_dim),
            nn.Sigmoid()
        )

    def forward(self, x):
        weights = self.attention(x)
        return x * weights
```

Explanation:

Attention weights were applied by reducing the dimensionality to half and then expanding back for each layer (including genotype input layer). Attention weights improved model performance by about 0.01 AUC during pilot testing and were thus retained as default during hyperparameter tuning.

Learnable mask (not used)

Code:

```
class MaskedLinear(nn.Module):
    def __init__(self, in_features, out_features, bias=True):
        super(MaskedLinear, self).__init__()
        # Initialize the linear layer
        self.linear = nn.Linear(in_features, out_features, bias)
        # Initialize mask parameters with the same shape as weights
        self.mask_param = nn.Parameter(torch.ones_like(self.linear.weight))
        if bias:
            self.bias = self.linear.bias
        else:
            self.register_parameter('bias', None)

    def forward(self, x):
        # Apply sigmoid to mask parameters to get gating probabilities
        mask = torch.sigmoid(self.mask_param)
```

```
        # Apply the mask to the weights
        masked_weight = self.linear.weight * mask
        return nn.functional.linear(x, masked_weight, self.bias)

    def get_binary_mask(self, threshold=0.5):
        """
        Returns a binary mask based on the gating parameters and a specified threshold.
        """
        with torch.no_grad():
            mask = torch.sigmoid(self.mask_param)
            binary_mask = (mask > threshold).float()
        return binary_mask
```

Explanation:

A learnable mask was designed during pilot testing with the hope to prune away non-useful features in addition to feature selection via relief/lasso. Each feature is attached with a gating mechanism where mask values above a certain threshold after passing through a sigmoid function get retained, otherwise they get dropped and the features remain inactive. It did not improve performance during pilot testing and was thus dropped during hyperparameter tuning. It could potentially be added if computation resources allow for more extensive exploration of feature space.

Batch normalisation

Explanation:

Batch nromalisation was added after timing the feature value at each middle node. This is to prevent extreme value fluctuations caused by timing a feature value to the sum of all previous layer contributions. Batch normalisation improved model performance by about 0.01 AUC during pilot testing and was thus retained as default during hyperparameter tuning.

All other commonly used hyperparameters in conventional neural networks (e.g. loss function, optimiser, L1 regularisation, oversampling) are applicable to TSNN and were not included in this study as part of the hyperparameter tuning process.

**TSGNN**

Both learnable mask and batch normalisation were applicable to TSGNN, but learnable mask did not improve model performance, so only batch normalisation was used after timing each node’s feature value with the sum of incoming values.

For more details regarding model structure and hyperparameters employed, see Github page for full code.

### Feature Selection Results Comparison

#### Class 1

| | LASSO-AUC | Relief-AUC | LASSO-features | LASSO-alpha | Relief-features |
|---|---|---|---|---|---|
| decision tree | 0.584473348 | 0.729555428 | 24, 20, 16, 38, 15, 12, 3, 10, 13, 21, 9, 22, 19, 26, 28, 33, 2 | 0.006951928 | 37, 30, 25, 26, 29, 28, 32, 38, 36, 10 |
| random forest | 0.767295761 | 0.765169092 | 24, 20, 16, 15, 12, 3, 10, 13, 21, 9, 22, 28, 29, 19, 38, 2, 6, 26, 31, 5, 33, 11, 35, 34, 1, 8, 36, 32, 30, 23, 14, 0, 4, 27, 37, 18, 17 | 0.037926902 | 37, 30, 25, 26, 29, 28, 32, 38, 36, 10, 7, 13, 33, 12, 34, 24, 14, 18, 3, 15, 6, 16, 8, 11, 17, 4, 1, 9, 19, 31, 5, 0, 20, 35, 22, 23, 2, 27, 21 |
| SVM | 0.708094201 | 0.70662508 | 24, 20, 16, 38, 15, 12, 3, 10, 13, 21, 9, 22, 19, 26, 28, 33, 2, 29, 6, 5, 35, 32, 30, 11, 31, 25, 8, 1, 23, 37, 27, 14, 0, 4, 18, 17, 36 | 0.006951928 | 37, 30, 25, 26, 29, 28, 32, 38, 36, 10, 7, 13, 33, 12, 34, 24, 14, 18, 3, 15, 6, 16, 8, 11, 17, 4, 1, 9, 19, 31, 5, 0, 20, 35, 22, 23, 2 |
| KNN | 0.69234721 | 0.694794175 | 24, 20, 16, 15, 12, 3, 10, 13, 21, 9, 22, 28, 29, 19, 38, 2, 6, 26, 31, 5, 33, 11, 35, 34, 1, 8, 36, 32, 30, 23, 14, 0, 4, 27, 37, 18, 17 | 0.037926902 | 37, 30, 25, 26, 29, 28, 32, 38, 36, 10, 7, 13, 33, 12, 34, 24, 14, 18, 3, 15, 6, 16, 8, 11, 17, 4, 1, 9, 19, 31, 5, 0, 20, 35, 22, 23 |
| naïve bayes | 0.662029907 | 0.66105154 | 24 | 0.1 | 37, 30, 25, 26, 29, 28, 32, 38, 36, 10, 7, 13, 33, 12, 34, 24, 14, 18, 3, 15, 6 |
| adaboost | 0.695538385 | 0.694519778 | 24, 20, 16, 15, 12, 3, 10, 13, 21, 9, 22, 28, 29, 19, 38, 2, 6, 26, 31 | 0.037926902 | 37, 30, 25, 26, 29, 28, 32, 38, 36, 10, 7, 13, 33, 12, 34, 24, 14, 18, 3, 15, 6, 16, 8, 11 |
| gradient boosting | 0.714393733 | 0.720638444 | 24, 20, 16, 15, 12, 3, 10, 13, 21, 9, 22, 28, 29, 19, 38, 2, 6, 26, 31, 5, 33, 11, 35, 34, 1, 8, 36 | 0.037926902 | 37, 30, 25, 26, 29, 28, 32, 38, 36, 10, 7, 13, 33, 12 |
| MLP | 0.734657131 | 0.733912176 | 24, 20, 16, 15, 12, 3, 10, 13, 21, 22, 9, 28, 29, 19, 31, 2, 6, 5, 26, 34, 33, 36, 11, 1, 27, 37, 8, 25, 14, 23, 0, 35, 4, 32, 30, 18 | 0.078475997 | 37, 30, 25, 26, 29, 28, 32, 38, 36, 10, 7, 13, 33, 12, 34, 24, 14, 18, 3, 15, 6, 16, 8, 11, 17, 4, 1, 9, 19, 31, 5, 0, 20, 35, 22, 23 |
| logistic regression | 0.671088876 | 0.6604 | 24, 38, 20, 16, 15, 12, 3, 10, 13, 21, 9, 22, 26, 33, 19, 2, 35, 32, 28, 6, 30 | 0.011288379 | 37, 30, 25, 26, 29, 28, 32, 38, 36, 10, 7, 13, 33, 12, 34, 24, 14, 18, 3, 15, 6, 16, 8, 11, 17, 4, 1, 9, 19, 31, 5, 0, 20, 35, 22, 23, 2, 27, 21 |
| Baysian network | 0.6494 | 0.6494 | 24, 30, 28, 9, 3, 12, 13, 8, 2, 10, 7, 6, 27, 25, 36, 20, 33, 19, 15 | 0.00001 | 29, 15, 10, 5, 11, 36, 12, 3, 38, 33, 2, 16, 14, 13, 4, 7, 6, 24 |

### Class 1-3

| | LASSO-AUC | Relief-AUC | LASSO-features | LASSO-alpha | Relief-features |
|---|---|---|---|---|---|
| decision tree | 0.601006395 | 0.7276 | 138, 244, 224, 137, 118, 103, 204, 230, 3, 86, 42, 24, 13, 21, 78, 33, 88, 183, 123, 119, 126, 210, 122, 65, 201, 5, 253, 30, 109, 227, 90, 49, 214, 52, 80, 25, 247, 217, 22, 229, 157, 115, 68, 155, 206, 182, 173, 59, 142, 44, 26, 28, 231, 232, 82, 50, 156, 116, 243, 1, 202, 61, 251, 174, 58, 240, 154, 74, 4, 145, 55, 170, 57, 255, 11, 199, 172, 133, 62, 29, 32, 81, 9, 222, 213, 67, 73, 40, 190, 203, 205, 223, 168, 165, 221, 0, 69, 184, 71, 148, 20, 189, 121, 151, 64, 92, 171, 167, 197, 185, 147, 188, 187, 226, 177, 176, 180, 196, 51, 124, 56, 117, 215, 27, 105, 18, 72, 163, 200, 31, 38, 238, 35, 160, 95, 12, 146, 166, 128, 41, 70, 77, 225, 175, 113, 234, 130, 120, 39 | 0.1 | 28, 234, 36, 215, 189, 37, 77, 26, 250, 188, 236, 181, 70, 55, 48, 67, 202, 244, 78, 65, 52, 237, 211, 57, 185, 229, 231, 219, 62, 220, 51, 200, 30, 242, 233, 198, 34, 25, 212, 205, 252, 32, 29, 248, 253, 75, 66, 247, 56, 58, 12, 221, 41, 197, 7, 63, 217, 1, 251, 199 |
| random forest | 0.767986759 | 0.7733 | 138, 244, 137, 224, 118, 103, 230, 42, 78, 204, 13, 33, 86, 123, 24, 3, 5, 21, 183, 126, 119, 65, 201, 30, 253, 122, 88, 49, 210, 80, 214, 217, 52, 206, 173, 90, 44, 59, 71, 229, 25, 157, 68, 247, 227, 182, 28 | 0.061584821 | 28, 234, 36, 215, 189, 37, 77, 26, 250, 188, 236, 181, 70, 55, 48, 67, 202, 244, 78, 65, 52, 237, 211, 57, 185, 229, 231, 219, 62, 220, 51, 200, 30, 242, 233, 198, 34, 25, 212, 205, 252, 32, 29, 248, 253, 75, 66, 247, 56, 58, 12, 221, 41, 197, 7, 63, 217, 1, 251, 199, 114, 49, 24, 50, 101, 80, 186, 232, 207, 130, 79, 115, 43, 243, 190, 137, 136, 108, 46, 214, 17, 10, 110, 88, 125, 182, 33, 203, 225, 68, 213, 227, 126, 93, 81, 14, 13, 104, 92, 42, 201, 129, 177, |

| | | | | | |
|---|---|---|---|---|---|
| | | | | | 122, 134, 133, 60, 19, 31, 9, 135, 98, 8, 45, 3, 47, 4, 6, 226, 116, 106, 90, 15, 105, 138, 18, 89, 84, 100, 44, 228, 131, 38, 112, 103, 96, 16, 127, 206, 117, 11, 102, 176, 128, 97, 0, 132, 5, 256, 61, 235, 87, 91, 193, 39, 111, 64, 180, 99, 95, 179, 35, 191, 246, 94, 238, 109, 22, 249, 187, 204, 245, 174, 53, 40, 86, 107, 119, 222, 239, 183, 157, 141, 123, 20, 196, 85, 83, 82, 167, 23, 139, 241, 72, 159, 2, 192, 175, 223, 54, 156, 73, 69, 208, 161, 120, 195, 158 |
| SVM | 0.716759342 | 0.7074 | 138, 244, 224, 137, 118, 103, 204, 230, 3, 86, 42, 24, 13, 21, 78, 33, 88, 183, 123, 119, 126, 210, 122, 65, 201, 5, 253, 30, 109, 227, 90, 49, 214, 52, 80, 25, 247, 217, 22, 229, 157, 115 | 0.1 | 28, 234, 36, 215, 189, 37, 77, 26, 250, 188, 236, 181, 70, 55, 48, 67, 202, 244, 78, 65, 52, 237, 211, 57, 185, 229, 231, 219, 62, 220, 51, 200, 30, 242, 233, 198, 34, 25, 212, 205, 252, 32, 29, 248, 253, 75, 66, 247, 56, 58, 12, 221, 41, 197, 7, 63, 217, 1, 251, 199, 114, 49, 24, 50, 101, 80, 186, 232, 207, 130, 79, 115, 43, 243, 190, 137, 136, 108, 46, 214, 17, 10, 110, 88, 125, 182, 33, 203, 225, 68, 213, 227, 126, 93, 81, 14, 13, 104, 92, 42, 201, 129, 177, 122, 134, 133, 60, 19, 31, 9, 135, 98, 8, 45, 3, 47, 4, 6, 226, 116, 106, 90, 15, 105, 138, 18, 89, 84, 100, 44, 228, 131, 38, 112, 103, 96, 16, 127, 206, 117, 11, 102, 176, 128, 97, 0, 132, 5, 256, 61, 235, 87, 91, 193, 39, 111, 64, 180, 99, 95, 179, 35, 191, 246, 94, 238, 109, 22, 249, 187, 204, 245, 174, 53, 40, 86, 107, 119, 222, 239, 183, 157, 141, 123, 20, 196, 85, 83, 82, 167, 23, 139, 241, 72, 159, 2, 192, 175, 223 |
| KNN | 0.700922586 | 0.7011 | 138, 244, 224, 137, 118, 103, 230, 204, 42 | 0.078475997 | 28, 234, 36, 215, 189, 37, 77, 26, 250, 188, 236, 181, 70, 55, 48, 67, 202, 244, 78, 65, 52, |

| | | | | | |
|---|---|---|---|---|---|
| | | | | | 237, 211, 57, 185, 229, 231, 219, 62, 220, 51, 200, 30, 242, 233, 198, 34, 25, 212, 205, 252, 32, 29, 248, 253, 75, 66, 247, 56, 58, 12, 221, 41, 197, 7, 63, 217, 1, 251, 199, 114, 49, 24, 50, 101, 80, 186, 232, 207, 130, 79, 115, 43, 243, 190, 137, 136, 108, 46, 214, 17, 10, 110, 88, 125, 182, 33, 203, 225, 68, 213, 227, 126, 93, 81, 14, 13, 104, 92, 42, 201, 129, 177, 122, 134, 133, 60, 19, 31, 9, 135, 98, 8, 45, 3, 47, 4, 6, 226, 116, 106, 90, 15 |
| naïve bayes | 0.682788866 | 0.6949 | 137, 118, 244, 224, 78, 138, 5, 154, 13, 86, 140, 183, 103, 33, 126, 230, 42, 71, 201, 65, 49, 55, 240, 253, 217, 206, 204, 74, 30, 59, 4, 123, 102, 119, 187, 28, 82, 73, 24, 52, 214, 199, 203, 202, 84, 190, 3, 0, 44, 61, 251, 182, 255, 21, 9, 189, 50, 80, 108, 128, 134, 11, 58, 232, 184, 212, 173, 194, 226, 213, 122, 209, 210, 68, 125, 243, 109, 142, 41, 85, 1, 145, 67, 7, 100, 157, 170, 38, 238, 40, 220, 25, 60, 121, 229, 193, 22, 155, 116, 191, 215, 177, 223, 114, 247, 156, 26, 168, 18, 88, 196, 179, 115, 29, 90, 227, 19, 180, 133, 231, 172, 72, 96, 87, 135, 171, 132, 17, 165, 56, 45, 62, 127, 221, 31, 53, 35, 32, 219, 48, 174, 57, 130, 104, 245, 97, 148, 228, 185, 151, 99, 34, 188, 160, 167, 66, 254, 163, 234, 10, 195, 81, 14, 222, 20, 98, 169, 198, 77, 200, 12, 249, 225, 51, 15, 69, 162, 207, 131, 205, 147, 70, 2, 208, 113, 101, 186, 47, 76, 176, 161, 39, 124, 211, 256, 252, 166, 75, 250, 248, 91, 237, 54, 233, 117, 43, 112, 216 | 0.005455595 | 28, 234, 36, 215, 189, 37, 77, 26, 250, 188, 236, 181, 70, 55, 48, 67, 202, 244, 78, 65, 52, 237, 211, 57, 185, 229, 231, 219, 62, 220, 51, 200, 30, 242, 233, 198, 34, 25, 212, 205, 252, 32, 29, 248, 253, 75, 66, 247, 56, 58, 12, 221, 41, 197, 7, 63, 217, 1, 251, 199, 114, 49, 24, 50, 101, 80, 186, 232, 207, 130, 79, 115, 43, 243, 190, 137, 136, 108, 46, 214, 17, 10, 110, 88, 125, 182, 33, 203, 225, 68, 213, 227, 126, 93, 81, 14, 13, 104, 92, 42, 201, 129, 177, 122, 134, 133, 60, 19, 31, 9, 135, 98, 8, 45, 3, 47, 4, 6, 226, 116, 106, 90, 15, 105, 138, 18, 89, 84, 100, 44, 228, 131, 38, 112, 103, 96, 16, 127, 206, 117, 11, 102, 176, 128, 97, 0, 132, 5, 256, 61, 235, 87, 91, 193, 39 |

| | | | | | |
|---|---|---|---|---|---|
| adaboost | 0.714989998 | 0.7089 | 137, 118, 244, 224, 5, 78, 154, 13, 138, 140, 86, 183, 103, 33, 126, 230, 42, 201, 71, 65, 49, 55, 240, 74, 206, 217, 253, 59, 102, 30, 4, 204, 187, 28, 82, 84, 73, 119, 123, 52, 199, 214, 203, 190, 202 | 0.001274275 | 28, 234, 36, 215, 189, 37, 77, 26, 250, 188, 236, 181, 70, 55, 48, 67, 202, 244, 78, 65, 52, 237, 211, 57, 185, 229, 231, 219, 62, 220, 51, 200, 30, 242, 233, 198, 34, 25, 212, 205, 252, 32, 29, 248, 253, 75, 66, 247, 56, 58, 12, 221, 41, 197, 7, 63, 217, 1, 251, 199, 114, 49, 24, 50, 101, 80, 186, 232, 207, 130, 79, 115, 43, 243, 190, 137, 136, 108, 46, 214, 17, 10, 110, 88, 125, 182, 33, 203, 225, 68, 213, 227, 126, 93, 81, 14, 13, 104, 92, 42, 201, 129, 177, 122, 134, 133, 60, 19, 31, 9, 135, 98, 8, 45, 3, 47, 4, 6, 226, 116, 106, 90, 15, 105, 138, 18, 89, 84, 100, 44, 228, 131, 38, 112, 103, 96, 16, 127, 206, 117, 11, 102, 176, 128, 97, 0, 132, 5, 256, 61, 235, 87, 91, 193, 39, 111, 64, 180, 99 |
| gradient boosting | 0.731445851 | 0.7266 | 138, 244, 224, 137, 118, 103, 230, 204, 42, 13, 86, 3 | 0.078475997 | 28, 234, 36, 215, 189, 37, 77, 26, 250, 188, 236, 181, 70, 55, 48, 67, 202, 244, 78, 65, 52, 237, 211, 57, 185, 229, 231, 219, 62, 220, 51, 200, 30, 242, 233, 198, 34, 25, 212, 205, 252, 32, 29, 248, 253, 75, 66, 247, 56, 58, 12 |
| MLP | 0.741557538 | 0.7404 | 137, 118, 244, 224, 78, 5, 154, 13, 138, 140, 86, 183, 103, 33, 126, 230, 42, 201, 71, 65, 49, 55, 240, 253, 74, 206, 217, 59, 30, 102, 204, 4, 187, 28, 82, 73, 119, 84, 123, 52, 199, 214, 203, 190, 24, 202, 3, 44, 251, 61, 189, 0, 255, 182, 134, 9, 209 | 0.002069138 | 28, 234, 36, 215, 189, 37, 77, 26, 250, 188, 236, 181, 70, 55, 48, 67, 202, 244, 78, 65, 52, 237, 211, 57, 185, 229, 231, 219, 62, 220, 51, 200, 30, 242, 233, 198, 34, 25, 212, 205, 252, 32, 29, 248, 253, 75, 66, 247, 56, 58, 12, 221, 41, 197, 7, 63, 217, 1, 251, 199, 114, 49, 24, 50, 101, 80, 186, 232, 207, 130, 79, 115, 43, 243, 190, 137, 136, 108, 46, 214, 17, 10, 110, 88, 125, 182, 33, 203, 225, 68, 213, 227, 126, 93, 81, 14, 13, 104, 92, 42, 201, 129, 177, 122, 134, 133, 60, 19, 31, 9, 135, 98, 8, 45, 3, 47, 4, 6, 226, 116, 106, 90, 15, 105, 138, 18, |

| | | | | | |
|---|---|---|---|---|---|
| | | | | | 89, 84, 100, 44, 228, 131, 38, 112, 103, 96, 16, 127, 206, 117, 11, 102, 176, 128 |
| logistic regression | 0.754261739 | 0.7376 | 224, 3, 204, 138, 88, 21, 183, 22, 227, 142, 24, 244, 13, 103, 119, 184, 86, 26, 38, 155, 229, 222, 122, 253, 126, 174, 25, 82, 77, 137, 157, 182, 42, 216, 49, 232, 115, 133, 196, 231, 217, 90, 210, 116, 202, 59, 156, 109, 65, 20, 52, 201, 113, 68, 243, 214, 206, 185, 57, 197, 78, 64, 1, 221, 69, 80, 91, 11, 176, 29, 35, 123, 117, 33, 67, 147, 215, 61, 154, 207, 51, 252, 62, 190, 179, 30, 124, 167, 148, 28, 203, 168, 53, 146, 5 | 0.379269019 | 28, 234, 36, 215, 189, 37, 77, 26, 250, 188, 236, 181, 70, 55, 48, 67, 202, 244, 78, 65, 52, 237, 211, 57, 185, 229, 231, 219, 62, 220, 51, 200, 30, 242, 233, 198, 34, 25, 212, 205, 252, 32, 29, 248, 253, 75, 66, 247, 56, 58, 12, 221, 41, 197, 7, 63, 217, 1, 251, 199, 114, 49, 24, 50, 101, 80, 186, 232, 207, 130, 79, 115, 43, 243, 190, 137, 136, 108, 46, 214, 17, 10, 110, 88, 125, 182, 33, 203, 225, 68, 213, 227, 126, 93, 81, 14, 13, 104, 92, 42, 201, 129, 177, 122, 134, 133, 60, 19, 31, 9, 135, 98, 8, 45, 3, 47, 4, 6, 226, 116, 106, 90, 15, 105, 138, 18, 89, 84, 100, 44, 228, 131, 38, 112, 103, 96, 16, 127, 206, 117, 11, 102, 176, 128, 97, 0, 132, 5, 256, 61, 235, 87, 91, 193, 39, 111, 64, 180, 99, 95, 179, 35, 191, 246, 94, 238, 109, 22, 249, 187, 204, 245, 174, 53, 40, 86, 107, 119, 222, 239, 183, 157, 141, 123, 20, 196, 85, 83, 82, 167, 23, 139, 241, 72, 159, 2, 192, 175, 223, 54, 156, 73, 69, 208, 161, 120, 195, 158, 166, 59, 153, 147, 160, 172, 216, 173, 165, 76, 27, 178, 74, 209, 255, 224, 124, 194, 164, 171, 155, 254, 184, 113, 143, 71, 230, 144, 146, 210, 21, 240, 170, 218, 149, 163, 154 |
| Baysian network | 0.5892 | 0.6124 | 254, 224, 3, 208, 241, 202, 25, 67, 74, 210, 216, 60, 246, 245, 53, 66, 48, 33, 57, 229, 212, 81, 88, 102, 7, 78, 58, 244, 207, 181, 112, 232, 76, 197, 13, 52 | LASSO-alpha | 198, 11, 244, 103, 55, 45, 253, 234, 177, 200, 39, 247, 248, 43, 62, 112, 97, 80, 98, 6, 217, 29, 7, 87, 86, 5, 231, 102, 232, 137, 213, 227, 111 |

### Hyperparameter Tuning Results

#### Class 1

**Decision tree:**

feature_indexes = [37, 30, 25, 26, 29, 28, 32, 38, 36, 10]

Best Average AUC: 0.7317 ± 0.0455

Average Accuracy: 0.9073 ± 0.0018

Average Precision: 0.0619 ± 0.1243

Average F1 Score: 0.0128 ± 0.0256

Best Hyperparameters:

criterion: entropy

max_depth: 12

max_features: None

min_samples_leaf: 1

min_samples_split: 10

splitter: best

=== Optimized Threshold ===

Threshold for Maximum F1 Score: 0.16

F1 Score: 0.3086 ± 0.0535

Accuracy: 0.8256 ± 0.0192

Precision: 0.2429 ± 0.0437

With oversampling:

Best Average AUC: 0.7339 ± 0.0437

Average Accuracy: 0.7457 ± 0.0316

Average Precision: 0.1932 ± 0.0333

Average F1 Score: 0.2855 ± 0.0438

Best Hyperparameters:

classifier__criterion: entropy

classifier__max_depth: 19

classifier__max_features: None

classifier__min_samples_leaf: 4

classifier__min_samples_split: 5

classifier__splitter: best

sampler: RandomOverSampler()

sampler__sampling_strategy: 0.75

=== Optimized Threshold ===

Average AUPRC: 0.2390 ± 0.0520

Optimal Threshold (F1-maximizing): 0.6289

F1 Score: 0.2959 ± 0.0576

Accuracy: 0.8431 ± 0.0206

Precision: 0.2535 ± 0.0550

Sensitivity (Recall): 0.3581 ± 0.0628

Specificity: 0.8918 ± 0.0194

**Random Forest:**

Feature_indexes (relief): [37, 30, 25, 26, 29, 28, 32, 38, 36, 10, 7, 13, 33, 12, 34, 24, 14, 18, 3, 15, 6, 16, 8, 11, 17, 4, 1, 9, 19, 31, 5, 0, 20, 35, 22, 23, 2, 27, 21]

Best Average AUC: 0.7745 ± 0.0170

Average Accuracy: 0.8186 ± 0.0046

Average Precision: 0.2533 ± 0.0803

Average F1 Score: 0.2083 ± 0.0559

Best Hyperparameters:

bootstrap: False

criterion: gini

max_depth: 30

max_features: log2

min_samples_leaf: 2

min_samples_split: 5

n_estimators: 200

=== Optimized Threshold ===

Threshold for Maximum F1 Score: 0.16

F1 Score: 0.3376 ± 0.0215

Accuracy: 0.8186 ± 0.0100

Precision: 0.2536 ± 0.0170

With Oversampling:

Best Average AUC: 0.7810 ± 0.0204

Average Accuracy: 0.8672 ± 0.0078

Average Precision: 0.3042 ± 0.0311

Average F1 Score: 0.3256 ± 0.0339

Best Hyperparameters:

classifier__bootstrap: False

classifier__criterion: gini

classifier__max_depth: 30

classifier__max_features: log2

classifier__min_samples_leaf: 2

classifier__min_samples_split: 2

classifier__n_estimators: 250

sampler: ADASYN()

sampler__sampling_strategy: auto

=== Optimized Threshold ===

Threshold for Maximum F1 Score: 0.32

F1 Score: 0.3274 ± 0.0311

Accuracy: 0.8212 ± 0.0103

Precision: 0.2495 ± 0.0237

Feature index (LASSO): [24, 20, 16, 15, 12, 3, 10, 13, 21, 9, 22, 28, 29, 19, 38, 2, 6, 26, 31, 5, 33, 11, 35, 34, 1, 8, 36, 32, 30, 23, 14, 0, 4, 27, 37, 18, 17]

Best Average AUC: 0.7752 ± 0.0163

Average Accuracy: 0.9018 ± 0.0030

Average Precision: 0.3952 ± 0.0585

Average F1 Score: 0.2223 ± 0.0535

Best Hyperparameters:

bootstrap: False

criterion: gini

max_depth: None

max_features: log2

min_samples_leaf: 2

min_samples_split: 2

n_estimators: 100

=== Optimized Threshold ===

Threshold for Maximum F1 Score: 0.16

F1 Score: 0.3435 ± 0.0217

Accuracy: 0.8249 ± 0.0065

Precision: 0.2612 ± 0.0135

With Oversampling:

Best Average AUC: 0.7814 ± 0.0223

Average Accuracy: 0.8702 ± 0.0073

Average Precision: 0.3067 ± 0.0313

Average F1 Score: 0.3185 ± 0.0318

Best Hyperparameters:

classifier__bootstrap: False

classifier__criterion: gini

classifier__max_depth: None

classifier__max_features: log2

classifier__min_samples_leaf: 2

classifier__min_samples_split: 5

classifier__n_estimators: 200

sampler: SMOTE()

sampler__sampling_strategy: 0.75

=== Optimized Threshold ===

Optimal Threshold (F1-maximizing): 0.3605

Average AUPRC: 0.2788 ± 0.0406

F1 Score: 0.3384 ± 0.0224

Accuracy: 0.8413 ± 0.0082

Precision: 0.2734 ± 0.0178

Sensitivity (Recall): 0.4451 ± 0.0382

Specificity: 0.8811 ± 0.0096

**SVM:**

Feature indexes (relief): [37, 30, 25, 26, 29, 28, 32, 38, 36, 10, 7, 13, 33, 12, 34, 24, 14, 18, 3, 15, 6, 16, 8, 11, 17, 4, 1, 9, 19, 31, 5, 0, 20, 35, 22, 23, 2]

Best Average AUC: 0.7355 ± 0.0265

Average Accuracy: 0.7125 ± 0.0112

Average Precision: 0.1827 ± 0.0103

Average F1 Score: 0.2820 ± 0.0157

Best Hyperparameters:

C: 100

class_weight: balanced

gamma: scale

kernel: rbf

tol: 0.001

=== Optimized Threshold ===

Threshold for Maximum F1 Score: 0.18

F1 Score: 0.3171 ± 0.0334

Accuracy: 0.8283 ± 0.0170

Precision: 0.2504 ± 0.0288

With Oversampling:

Best Average AUC: 0.7386 ± 0.0259

Average Accuracy: 0.7730 ± 0.0095

Average Precision: 0.2152 ± 0.0143

Average F1 Score: 0.3111 ± 0.0207

Best Hyperparameters:

classifier__C: 125

classifier__class_weight: None

classifier__gamma: scale

classifier__kernel: rbf

classifier__tol: 0.001

sampler: RandomOverSampler

sampler__sampling_strategy: 0.75

=== Optimized Threshold ===

Optimal Threshold (F1-maximizing): 0.6612

Average AUPRC: 0.2494 ± 0.0412

F1 Score: 0.3104 ± 0.0244

Accuracy: 0.8227 ± 0.0112

Precision: 0.2411 ± 0.0184

Sensitivity (Recall): 0.4378 ± 0.0452

Specificity: 0.8613 ± 0.0137

Feature Indexes (LASSO): [24, 20, 16, 38, 15, 12, 3, 10, 13, 21, 9, 22, 19, 26, 28, 33, 2, 29, 6, 5, 35, 32, 30, 11, 31, 25, 8, 1, 23, 37, 27, 14, 0, 4, 18, 17, 36]

Best Average AUC: 0.7235 ± 0.0256

Average Accuracy: 0.7221 ± 0.0107

Average Precision: 0.1848 ± 0.0114

Average F1 Score: 0.2824 ± 0.0168

Best Hyperparameters:

C: 100

class_weight: balanced

gamma: scale

kernel: rbf

tol: 0.0001

=== Optimized Threshold ===

Threshold for Maximum F1 Score: 0.17

F1 Score: 0.2989 ± 0.0329

Accuracy: 0.8311 ± 0.0179

Precision: 0.2427 ± 0.0312

With Oversampling:

Best Average AUC: 0.7279 ± 0.0302

Average Accuracy: 0.7801 ± 0.0148

Average Precision: 0.2123 ± 0.0196

Average F1 Score: 0.3008 ± 0.0257

Best Hyperparameters:

classifier__C: 100

classifier__class_weight: None

classifier__gamma: scale

classifier__kernel: rbf

classifier__max_iter: 10000

classifier__tol: 0.001

sampler: RandomOverSampler

sampler__sampling_strategy: 0.75

=== Optimized Threshold ===

Threshold for Maximum F1 Score: 0.47

F1 Score: 0.3027 ± 0.0261

Accuracy: 0.7843 ± 0.0088

Precision: 0.2148 ± 0.0175

**KNN:**

Feature Indexes (relief): [37, 30, 25, 26, 29, 28, 32, 38, 36, 10, 7, 13, 33, 12, 34, 24, 14, 18, 3, 15, 6, 16, 8, 11, 17, 4, 1, 9, 19, 31, 5, 0, 20, 35, 22, 23]

Best Average AUC: 0.7586 ± 0.0267

Average Accuracy: 0.9057 ± 0.0039

Average Precision: 0.3829 ± 0.1674

Average F1 Score: 0.0837 ± 0.0445

Best Hyperparameters:

algorithm: auto

leaf_size: 20

metric: manhattan

n_neighbors: 28

weights: distance

=== Optimized Threshold ===

Threshold for Maximum F1 Score: 0.15

F1 Score: 0.3394 ± 0.0473

Accuracy: 0.8216 ± 0.0186

Precision: 0.2574 ± 0.0395

With Oversampling:

Best Average AUC: 0.7600 ± 0.0165

Average Accuracy: 0.7572 ± 0.0156

Average Precision: 0.2104 ± 0.0196

Average F1 Score: 0.3116 ± 0.0270

Best Hyperparameters:

classifier__algorithm: kd_tree

classifier__leaf_size: 50

classifier__metric: minkowski

classifier__n_neighbors: 28

classifier__p: 1

classifier__weights: distance

oversampler: RandomOverSampler()

oversampler__sampling_strategy: 0.9

=== Optimized Threshold ===

Optimal Threshold (F1-maximizing): 0.6068

Average AUPRC: 0.2583 ± 0.0406

F1 Score: 0.3276 ± 0.0329

Accuracy: 0.8139 ± 0.0118

Precision: 0.2447 ± 0.0255

Sensitivity (Recall): 0.4962 ± 0.0494

Specificity: 0.8458 ± 0.0110

Feature Indexes (LASSO): [24, 20, 16, 15, 12, 3, 10, 13, 21, 9, 22, 28, 29, 19, 38, 2, 6, 26, 31, 5, 33, 11, 35, 34, 1, 8, 36, 32, 30, 23, 14, 0, 4, 27, 37, 18, 17]

Best Average AUC: 0.7538 ± 0.0283

Average Accuracy: 0.9076 ± 0.0030

Average Precision: 0.4681 ± 0.1483

Average F1 Score: 0.0888 ± 0.0324

Best Hyperparameters:

algorithm: auto

leaf_size: 20

metric: manhattan

n_neighbors: 24

p: 1

weights: distance

=== Optimized Threshold ===

Threshold for Maximum F1 Score: 0.18

F1 Score: 0.3360 ± 0.0496

Accuracy: 0.8491 ± 0.0155

Precision: 0.2823 ± 0.0470

With Oversampling:

Best Average AUC: 0.7594 ± 0.0223

Average Accuracy: 0.7305 ± 0.0156

Average Precision: 0.1980 ± 0.0142

Average F1 Score: 0.3021 ± 0.0195

Best Hyperparameters:

classifier__algorithm: brute

classifier__leaf_size: 15

classifier__metric: manhattan

classifier__n_neighbors: 24

classifier__weights: distance

oversampler: RandomOverSampler()

oversampler__sampling_strategy: auto

=== Optimized Threshold ===

Threshold for Maximum F1 Score: 0.76

F1 Score: 0.3214 ± 0.0506

Accuracy: 0.8620 ± 0.0127

Precision: 0.2930 ± 0.0492

**Naïve Bayes:**

Feature Indexes (LASSO): [24]

Best Average AUC: 0.6620 ± 0.0440

Average Accuracy: 0.8905 ± 0.0046

Average Precision: 0.2123 ± 0.0665

Average F1 Score: 0.1095 ± 0.0364

Best Hyperparameters:

classifier: GaussianNB()

classifier__var_smoothing: 1e-12

=== Optimized Threshold ===

Threshold for Maximum F1 Score: 0.08

F1 Score: 0.2441 ± 0.0469

Accuracy: 0.7651 ± 0.0158

Precision: 0.1729 ± 0.0335

With Oversampling:

Best Average AUC: 0.6620 ± 0.0440

Average Accuracy: 0.8524 ± 0.0129

Average Precision: 0.2111 ± 0.0520

Average F1 Score: 0.2154 ± 0.0488

Best Hyperparameters:

classifier: GaussianNB()

classifier__var_smoothing: 1e-12

sampler: RandomOverSampler()

sampler__sampling_strategy: 0.5

=== Optimized Threshold ===

Threshold for Maximum F1 Score: 0.30

F1 Score: 0.2420 ± 0.0462

Accuracy: 0.7617 ± 0.0152

Precision: 0.1705 ± 0.0325

Feature Indexes (relief): [37, 30, 25, 26, 29, 28, 32, 38, 36, 10, 7, 13, 33, 12, 34, 24, 14, 18, 3, 15, 6]

Best Average AUC: 0.6611 ± 0.0419

Average Accuracy: 0.8887 ± 0.0063

Average Precision: 0.2311 ± 0.0736

Average F1 Score: 0.1309 ± 0.0391

Best Hyperparameters:

classifier: GaussianNB()

classifier__var_smoothing: 1e-12

=== Optimized Threshold ===

Threshold for Maximum F1 Score: 0.13

F1 Score: 0.2672 ± 0.0461

Accuracy: 0.7894 ± 0.0191

Precision: 0.1965 ± 0.0361

With Oversampling:

Best Average AUC: 0.6626 ± 0.0414

Average Accuracy: 0.8215 ± 0.0181

Average Precision: 0.2062 ± 0.0438

Average F1 Score: 0.2532 ± 0.0495

Best Hyperparameters:

classifier: GaussianNB()

classifier__var_smoothing: 1e-11

sampler: RandomOverSampler()

sampler__sampling_strategy: 0.5

=== Optimized Threshold ===

Optimal Threshold (F1-maximizing): 0.4740

Average AUPRC: 0.1788 ± 0.0337

F1 Score: 0.2703 ± 0.0458

Accuracy: 0.8157 ± 0.0216

Precision: 0.2139 ± 0.0428

Sensitivity (Recall): 0.3706 ± 0.0544

Specificity: 0.8604 ± 0.0213

**Adaboost:**

Feature Indexes (LASSO): [24, 20, 16, 15, 12, 3, 10, 13, 21, 9, 22, 28, 29, 19, 38, 2, 6, 26, 31]

Best Average AUC: 0.7259 ± 0.0293

Average Accuracy: 0.9018 ± 0.0044

Average Precision: 0.3514 ± 0.0955

Average F1 Score: 0.1285 ± 0.0277

Best Hyperparameters:

ada__algorithm: SAMME

ada__base_estimator__max_depth: 4

ada__base_estimator__max_features: sqrt

ada__base_estimator__min_samples_leaf: 1

ada__base_estimator__min_samples_split: 2

ada__learning_rate: 1.0

ada__n_estimators: 200

=== Optimized Threshold ===

Threshold for Maximum F1 Score: 0.47

F1 Score: 0.2879 ± 0.0469

Accuracy: 0.8293 ± 0.0149

Precision: 0.2334 ± 0.0404

With Oversampling:

Best Average AUC: 0.7704 ± 0.0232

Average Accuracy: 0.8725 ± 0.0064

Average Precision: 0.3036 ± 0.0314

Average F1 Score: 0.3044 ± 0.0331

Best Hyperparameters:

ada__algorithm: SAMME

ada__estimator__max_depth: 25

ada__estimator__max_features: sqrt

ada__estimator__min_samples_leaf: 1

ada__estimator__min_samples_split: 5

ada__learning_rate: 0.0025

ada__n_estimators: 300

sampler: ADASYN()

sampler__sampling_strategy: 0.5

=== Optimized Threshold ===

Threshold for Maximum F1 Score: 0.34

F1 Score: 0.3261 ± 0.0281

Accuracy: 0.8368 ± 0.0103

Precision: 0.2622 ± 0.0249

Feature Indexes (relief): [37, 30, 25, 26, 29, 28, 32, 38, 36, 10, 7, 13, 33, 12, 34, 24, 14, 18, 3, 15, 6, 16, 8, 11]

Best Average AUC: 0.7327 ± 0.0341

Average Accuracy: 0.9057 ± 0.0027

Average Precision: 0.1948 ± 0.1804

Average F1 Score: 0.0321 ± 0.0285

Best Hyperparameters:

ada__algorithm: SAMME

ada__estimator__max_depth: 3

ada__estimator__max_features: sqrt

ada__estimator__min_samples_leaf: 4

ada__estimator__min_samples_split: 10

ada__learning_rate: 1.0

ada__n_estimators: 300

=== Optimized Threshold ===

Threshold for Maximum F1 Score: 0.46

F1 Score: 0.3113 ± 0.0498

Accuracy: 0.8337 ± 0.0210

Precision: 0.2529 ± 0.0451

With Oversampling:

Best Average AUC: 0.7711 ± 0.0221

Average Accuracy: 0.8306 ± 0.0098

Average Precision: 0.2530 ± 0.0223

Average F1 Score: 0.3201 ± 0.0285

Best Hyperparameters:

ada__algorithm: SAMME

ada__estimator__max_depth: 15

ada__estimator__max_features: sqrt

ada__estimator__min_samples_leaf: 2

ada__estimator__min_samples_split: 3

ada__learning_rate: 0.001

ada__n_estimators: 400

sampler: ADASYN()

sampler__sampling_strategy: 0.5

=== Optimized Threshold ===

Optimal Threshold (F1-maximizing): 0.4211

Average AUPRC: 0.2588 ± 0.0434

F1 Score: 0.3263 ± 0.0314

Accuracy: 0.8094 ± 0.0162

Precision: 0.2417 ± 0.0250

Sensitivity (Recall): 0.5050 ± 0.0535

Specificity: 0.8400 ± 0.0181

**Gradient Boosting:**

Feature Indexes (LASSO): [24, 20, 16, 15, 12, 3, 10, 13, 21, 9, 22, 28, 29, 19, 38, 2, 6, 26, 31, 5, 33, 11, 35, 34, 1, 8, 36]

Best Average AUC: 0.7421 ± 0.0357

Average Accuracy: 0.9029 ± 0.0057

Average Precision: 0.3689 ± 0.2441

Average F1 Score: 0.0993 ± 0.0386

Best Hyperparameters:

gbc__learning_rate: 0.1

gbc__loss: exponential

gbc__max_depth: 4

gbc__max_features: log2

gbc__min_samples_leaf: 4

gbc__min_samples_split: 10

gbc__n_estimators: 400

gbc__subsample: 0.6

=== Optimized Threshold ===

Threshold for Maximum F1 Score: 0.16

F1 Score: 0.3220 ± 0.0467

Accuracy: 0.8685 ± 0.0138

Precision: 0.3074 ± 0.0550

With Oversampling:

Best Average AUC: 0.7564 ± 0.0209

Average Accuracy: 0.8696 ± 0.0104

Average Precision: 0.2960 ± 0.0449

Average F1 Score: 0.3005 ± 0.0414

Best Hyperparameters:

gbc__learning_rate: 0.05

gbc__loss: exponential

gbc__max_depth: None

gbc__max_features: log2

gbc__min_samples_leaf: 1

gbc__min_samples_split: 14

gbc__n_estimators: 75

gbc__subsample: 0.4

resampler: RandomOverSampler()

resampler__sampling_strategy: auto

=== Optimized Threshold ===

Optimal Threshold (F1-maximizing): 0.2179

Average AUPRC: 0.2588 ± 0.0446

F1 Score: 0.3304 ± 0.0251

Accuracy: 0.8359 ± 0.0097

Precision: 0.2638 ± 0.0217

Sensitivity (Recall): 0.4433 ± 0.0352

Specificity: 0.8754 ± 0.0101

Feature Indexes (relief): [37, 30, 25, 26, 29, 28, 32, 38, 36, 10, 7, 13, 33, 12]

Best Average AUC: 0.7307 ± 0.0446

Average Accuracy: 0.9071 ± 0.0017

Average Precision: 0.1952 ± 0.2056

Average F1 Score: 0.0293 ± 0.0336

Best Hyperparameters:

gbc__learning_rate: 0.2

gbc__loss: exponential

gbc__max_depth: 5

gbc__max_features: sqrt

gbc__min_samples_leaf: 1

gbc__min_samples_split: 2

gbc__n_estimators: 100

gbc__subsample: 0.6

=== Optimized Threshold ===

Threshold for Maximum F1 Score: 0.16

F1 Score: 0.3003 ± 0.0487

Accuracy: 0.8237 ± 0.0189

Precision: 0.2367 ± 0.0410

With Oversampling:

Best Average AUC: 0.7337 ± 0.0433

Average Accuracy: 0.8073 ± 0.0198

Average Precision: 0.2198 ± 0.0433

Average F1 Score: 0.3039 ± 0.0530

Best Hyperparameters:

gbc__learning_rate: 0.05

gbc__loss: exponential

gbc__max_depth: 6

gbc__max_features: None

gbc__min_samples_leaf: 2

gbc__min_samples_split: 2

gbc__n_estimators: 200

gbc__subsample: 0.8

resampler: RandomOverSampler()

resampler__sampling_strategy: 0.5

=== Optimized Threshold ===

Threshold for Maximum F1 Score: 0.47

F1 Score: 0.2849 ± 0.0522

Accuracy: 0.8033 ± 0.0200

Precision: 0.2138 ± 0.0388

**Multi-layer Perceptron:**

Feature Indexes: [24, 20, 16, 15, 12, 3, 10, 13, 21, 22, 9, 28, 29, 19, 31, 2, 6, 5, 26, 34, 33, 36, 11, 1, 27, 37, 8, 25, 14, 23, 0, 35, 4, 32, 30, 18]

Best Average AUC: 0.7347 ± 0.0413

Average Accuracy: 0.9052 ± 0.0036

Average Precision: 0.3448 ± 0.1775

Average F1 Score: 0.0829 ± 0.0463

Best Hyperparameters:

activation: relu

alpha: 0.0001

batch_size: auto

early_stopping: False

hidden_layer_sizes: (100,)

learning_rate: constant

learning_rate_init: 0.001

solver: adam

=== Optimized Threshold ===

Threshold for Maximum F1 Score: 0.19

F1 Score: 0.3266 ± 0.0646

Accuracy: 0.8525 ± 0.0276

Precision: 0.2888 ± 0.0714

With Oversampling:

Best Average AUC: 0.7359 ± 0.0412

Average Accuracy: 0.8900 ± 0.0090

Average Precision: 0.3459 ± 0.0667

Average F1 Score: 0.2682 ± 0.0597

Best Hyperparameters:

classifier__activation: relu

classifier__alpha: 1e-06

classifier__batch_size: auto

classifier__early_stopping: False

classifier__hidden_layer_sizes: (100,)

classifier__learning_rate: adaptive

classifier__learning_rate_init: 0.0005

classifier__max_iter: 1500

classifier__solver: adam

sampler: RandomOverSampler()

sampler__sampling_strategy: 0.25

=== Optimized Threshold ===

Threshold for Maximum F1 Score: 0.38

F1 Score: 0.3112 ± 0.0510

Accuracy: 0.8602 ± 0.0196

Precision: 0.2881 ± 0.0525

Feature Indexes (relief): [37, 30, 25, 26, 29, 28, 32, 38, 36, 10, 7, 13, 33, 12, 34, 24, 14, 18, 3, 15, 6, 16, 8, 11, 17, 4, 1, 9, 19, 31, 5, 0, 20, 35, 22, 23]

Best Average AUC: 0.7428 ± 0.0396

Average Accuracy: 0.9049 ± 0.0043

Average Precision: 0.3570 ± 0.1506

Average F1 Score: 0.1094 ± 0.0492

Best Hyperparameters:

activation: relu

alpha: 0.0025

batch_size: auto

early_stopping: False

hidden_layer_sizes: (35, 70, 140)

learning_rate: constant

learning_rate_init: 0.001

max_iter: 200

solver: adam

=== Optimized Threshold ===

Threshold for Maximum F1 Score: 0.17

F1 Score: 0.3266 ± 0.0535

Accuracy: 0.8444 ± 0.0273

Precision: 0.2801 ± 0.0671

With Oversampling:

Best Average AUC: 0.7483 ± 0.0208

Average Accuracy: 0.7288 ± 0.0300

Average Precision: 0.1929 ± 0.0165

Average F1 Score: 0.2923 ± 0.0189

Best Hyperparameters:

classifier__activation: relu

classifier__alpha: 1e-05

classifier__batch_size: auto

classifier__early_stopping: True

classifier__hidden_layer_sizes: (150, 75, 35)

classifier__learning_rate: constant

classifier__learning_rate_init: 0.001

classifier__max_iter: 1000

classifier__solver: adam

sampler: RandomOverSampler()

sampler__sampling_strategy: 1.0

=== Optimized Threshold ===

Optimal Threshold (F1-maximizing): 0.7091

Average AUPRC: 0.2610 ± 0.0337

F1 Score: 0.3178 ± 0.0486

Accuracy: 0.8434 ± 0.0177

Precision: 0.2660 ± 0.0410

Sensitivity (Recall): 0.4005 ± 0.0705

Specificity: 0.8878 ± 0.0198

**Bayesian Network:**

Feature Indexes (lasso): [24, 30, 28, 9, 3, 12, 13, 8, 2, 10, 7, 6, 27, 25, 36, 20, 33, 19, 15]

scoring_method BIC

max_parents NaN

oversampler passthrough

sampling_rate NaN

avg_auc 0.649448

std_auc 0.038856

AUPRC: 0.1359 ± 0.0196

Optimal Threshold: 0.1015

F1 Score: 0.2401 ± 0.0401

Accuracy: 0.6837 ± 0.0662

Precision: 0.1555 ± 0.0281

Sensitivity: 0.5423 ± 0.1117

Specificity: 0.6979 ± 0.0789

Feature Indexes (relief): [29, 15, 10, 5, 11, 36, 12, 3, 38, 33, 2, 16, 14, 13, 4, 7, 6, 24]

scoring_method BIC

max_parents NaN

oversampler passthrough

sampling_rate NaN

avg_auc 0.649448

std_auc 0.038856

Accuracy: 0.7021 ± 0.0551

Precision: 0.1629 ± 0.0255

F1 Score: 0.2484 ± 0.0310

**Logistic Regression:**

Feature Indexes (lasso): [24, 38, 20, 16, 15, 12, 3, 10, 13, 21, 9, 22, 26, 33, 19, 2, 35, 32, 28, 6, 30]

Best Average AUC: 0.6739 ± 0.0469

Average Accuracy: 0.6657 ± 0.0219

Average Precision: 0.1552 ± 0.0202

Average F1 Score: 0.2463 ± 0.0315

Best Hyperparameters:

lr__C: 30

lr__l1_ratio: None

lr__penalty: l2

lr__solver: sag

resampler: RandomOverSampler()

resampler__sampling_strategy: auto

=== Optimized Threshold ===

Optimal Threshold (F1-maximizing): 0.6390

Average AUPRC: 0.2031 ± 0.0460

F1 Score: 0.2764 ± 0.0603

Accuracy: 0.8338 ± 0.0169

Precision: 0.2303 ± 0.0512

Sensitivity (Recall): 0.3474 ± 0.0783

Specificity: 0.8827 ± 0.0143

Feature Indexes (relief): [37, 30, 25, 26, 29, 28, 32, 38, 36, 10, 7, 13, 33, 12, 34, 24, 14, 18, 3, 15, 6, 16, 8, 11, 17, 4, 1, 9, 19, 31, 5, 0, 20, 35, 22, 23, 2, 27, 21]

Best Average AUC: 0.6668 ± 0.0509

Average Accuracy: 0.8547 ± 0.0148

Average Precision: 0.2352 ± 0.0604

Average F1 Score: 0.2459 ± 0.0612

Best Hyperparameters:

lr__C: 0.1

lr__l1_ratio: 0.5

lr__penalty: elasticnet

lr__solver: saga

resampler: SMOTE()

resampler__sampling_strategy: 0.5

=== Optimized Threshold ===

Threshold for Maximum F1 Score: 0.43

F1 Score: 0.2724 ± 0.0432

Accuracy: 0.8133 ± 0.0148

Precision: 0.2119 ± 0.0344

**TSGNN:**

global relief

Best trial:

Value (Mean AUC):  0.7455567712393181

Params:

n_epochs: 1175

lr: 0.002652887105202934

weight_decay: 1.1942523033711955e-07

batch_size: 64

n_genotype: 12

n_history: 5

n_phenotype: 11

n_behaviour: 4

Average AUC: 0.7456 ± 0.0312

Average F1 Score: 0.1408 ± 0.0673

Average Accuracy: 0.9058 ± 0.0083

Average Precision: 0.4434 ± 0.2184

Optimal Threshold: 0.1703

AUPRC: 0.2668 ± 0.0566

F1 Score: 0.3187 ± 0.0410

Accuracy: 0.8223 ± 0.0209

Precision: 0.2471 ± 0.0344

Sensitivity (Recall): 0.4553 ± 0.0713

Specificity: 0.8591 ± 0.0252

Separate relief:

Best trial:

Value (Mean AUC): 0.7394404100047507

Params:

n_epochs: 2080

lr: 0.00033145861044404 68

weight_decay: 0.0005417687953694567

batch_size: 128

n_genotype: 6

n_history: 2

n_phenotype: 13

n_behaviour: 1

Average AUC: 0.7394 ± 0.0405

Average F1 Score: 0.0334 ± 0.0297

Average Accuracy: 0.9079 ± 0.0031

Average Precision: 0.4571 ± 0.4554

Average F1 Score: 0.3331 ± 0.0619

Average Accuracy: 0.8162 ± 0.0740

Average Precision: 0.2721 ± 0.0782

Global lasso:

Best trial:

Value (Mean AUC): 0.7377684524348088

Params:

n_epochs: 1042

lr: 0.002295940611801856

weight_decay: 1.3988230833611534e-07

batch_size: 32

n_genotype: 13

n_history: 6

n_phenotype: 12

n_behaviour: 5

Average AUC: 0.7378 ± 0.0317

Average F1 Score: 0.1673 ± 0.0557

Average Accuracy: 0.9073 ± 0.0062

Average Precision: 0.4841 ± 0.1696

Average F1 Score: 0.3375 ± 0.0662

Average Accuracy: 0.8424 ± 0.0609

Average Precision: 0.3393 ± 0.1543

Separate lasso:

Best trial:

Value (Mean AUC): 0.7433285096690063

Params:

n_epochs: 1175

lr: 0.0004464002613441988

weight_decay: 1.4153526263149175e-08

batch_size: 128

n_genotype: 10

n_history: 3

n_phenotype: 13

n_behaviour: 2

Average AUC: 0.7433 ± 0.0368

Average F1 Score: 0.0718 ± 0.0559

Average Accuracy: 0.9073 ± 0.0049

Average Precision: 0.3878 ± 0.3015

Average F1 Score: 0.3459 ± 0.0665

Average Accuracy: 0.8327 ± 0.0723

Average Precision: 0.3171 ± 0.1140

**TSNN:**

global relief

Best Trial:

AUC: 0.7336598151821246 ± 0.0361

Params:

n_genotype: 14

n_history: 5

n_phenotype: 10

n_behaviour: 4

learning_rate: 0.0014641867315396168

epochs: 1546

batch_size: 16

Best Threshold to maximize F1 Score: 0.16

F1 Score: 0.3010 ± 0.0464

Accuracy: 0.8230 ± 0.0178

Precision: 0.2363 ± 0.0384

separate relief

Best Trial:

AUC: 0.7357700263311236

Params:

n_genotype: 13

n_history: 3

n_phenotype: 11

n_behaviour: 3

learning_rate: 0.0013776008234454911

epochs: 2382

batch_size: 32

Optimal Threshold: 0.1833

AUPRC: 0.2508 ± 0.0570

F1 Score: 0.3102 ± 0.0605

Accuracy: 0.8487 ± 0.0121

Precision: 0.2653 ± 0.0476

Sensitivity (Recall): 0.3756 ± 0.0836

Specificity: 0.8962 ± 0.0110

global lasso:

Best Trial:

AUC: 0.7242 (±0.0508)

F1 Score: 0.0411 (Std: 0.0449)

Accuracy: 0.9068 (Std: 0.0029)

Precision: 0.1867 (Std: 0.2059)

Params:

n_genotype: 12

n_history: 6

n_phenotype: 10

n_behaviour: 1

learning_rate: 0.0007849256583952668

epochs: 1674

batch_size: 64

Best Threshold to maximize F1 Score: 0.68

F1 Score: 0.2874 ± 0.0547

Accuracy: 0.8194 ± 0.0244

Precision: 0.2273 ± 0.0500

separate lasso:

Best Trial:

AUC: 0.7293 (±0.0259)

F1 Score: 0.0192 (±0.0321)

Accuracy: 0.9058 (±0.0024)

Precision: 0.1550 (±0.3020)

Params:

n_genotype: 13

n_history: 5

n_phenotype: 10

n_behaviour: 2

learning_rate: 0.0009955474543238663

epochs: 1353

batch_size: 64

Best Threshold to maximize F1 Score: 0.15

F1 Score: 0.2984 ± 0.0445

Accuracy: 0.8059 ± 0.0200

Precision: 0.2239 ± 0.0372

### Class 1-3

**Decision Tree**

Feature Indexes (relief): [28, 234, 36, 215, 189, 37, 77, 26, 250, 188, 236, 181, 70, 55, 48, 67, 202, 244, 78, 65, 52, 237, 211, 57, 185, 229, 231, 219, 62, 220, 51, 200, 30, 242, 233, 198, 34, 25, 212, 205, 252, 32, 29, 248, 253, 75, 66, 247, 56, 58, 12, 221, 41, 197, 7, 63, 217, 1, 251, 199]

Best Average AUC: 0.7337 ± 0.0400

Average Accuracy: 0.8966 ± 0.0101

Average Precision: 0.3636 ± 0.1096

Average F1 Score: 0.2262 ± 0.0686

Best Hyperparameters:

classifier__criterion: entropy

classifier__max_depth: 21

classifier__max_features: sqrt

classifier__min_samples_leaf: 2

classifier__min_samples_split: 7

classifier__splitter: best

sampler: RandomOverSampler()

sampler__sampling_strategy: 0.25

=== Optimized Threshold ===

Average AUPRC: 0.2303 ± 0.0437

Optimal Threshold (F1-maximizing): 0.3676

F1 Score: 0.2948 ± 0.0586

Accuracy: 0.8478 ± 0.0208

Precision: 0.2595 ± 0.0625

Sensitivity (Recall): 0.3438 ± 0.0527

Specificity: 0.8984 ± 0.0188

**Random Forest**

Feature indexes (lasso): [138, 244, 137, 224, 118, 103, 230, 42, 78, 204, 13, 33, 86, 123, 24, 3, 5, 21, 183, 126, 119, 65, 201, 30, 253, 122, 88, 49, 210, 80, 214, 217, 52, 206, 173, 90, 44, 59, 71, 229, 25, 157, 68, 247, 227, 182, 28]

Best Average AUC: 0.7825 ± 0.0206

Average Accuracy: 0.8830 ± 0.0059

Average Precision: 0.3273 ± 0.0365

Average F1 Score: 0.2942 ± 0.0357

Best Hyperparameters:

classifier__bootstrap: False

classifier__class_weight: None

classifier__criterion: gini

classifier__max_depth: None

classifier__max_features: sqrt

classifier__min_samples_leaf: 2

classifier__min_samples_split: 2

classifier__n_estimators: 290

sampler: ADASYN()

sampler__sampling_strategy: 0.25

=== Optimized Threshold ===

Threshold for Maximum F1 Score: 0.30

F1 Score: 0.3328 ± 0.0315

Accuracy: 0.8448 ± 0.0095

Precision: 0.2744 ± 0.0282

Feature indexes (relief): [28, 234, 36, 215, 189, 37, 77, 26, 250, 188, 236, 181, 70, 55, 48, 67, 202, 244, 78, 65, 52, 237, 211, 57, 185, 229, 231, 219, 62, 220, 51, 200, 30, 242, 233, 198, 34, 25, 212, 205, 252, 32, 29, 248, 253, 75, 66, 247, 56, 58, 12, 221, 41, 197, 7, 63, 217, 1, 251, 199, 114, 49, 24, 50, 101, 80, 186, 232, 207, 130, 79, 115, 43, 243, 190, 137, 136, 108, 46, 214, 17, 10, 110, 88, 125, 182, 33, 203, 225, 68, 213, 227, 126, 93, 81, 14, 13, 104, 92, 42, 201, 129, 177, 122, 134, 133, 60, 19, 31, 9, 135, 98, 8, 45, 3, 47, 4, 6, 226, 116, 106, 90, 15, 105, 138, 18, 89, 84, 100, 44, 228, 131, 38, 112, 103, 96, 16, 127, 206, 117, 11, 102, 176, 128, 97, 0, 132, 5, 256, 61, 235, 87, 91, 193, 39, 111, 64, 180, 99, 95, 179, 35, 191, 246, 94, 238, 109, 22, 249, 187, 204, 245, 174, 53, 40, 86, 107, 119, 222, 239, 183, 157, 141, 123, 20, 196, 85, 83, 82, 167, 23, 139, 241, 72, 159, 2, 192, 175, 223, 54, 156, 73, 69, 208, 161, 120, 195, 158]

Best Average AUC: 0.7842 ± 0.0189

Average Accuracy: 0.8845 ± 0.0056

Average Precision: 0.3068 ± 0.0497

Average F1 Score: 0.2532 ± 0.0564

Best Hyperparameters:

classifier__bootstrap: False

classifier__class_weight: None

classifier__criterion: entropy

classifier__max_depth: None

classifier__max_features: sqrt

classifier__min_samples_leaf: 2

classifier__min_samples_split: 7

classifier__n_estimators: 225

sampler: RandomOverSampler()

sampler__sampling_strategy: auto

=== Optimized Threshold ===

Optimal Threshold (F1-maximizing): 0.1938

Average AUPRC: 0.2734 ± 0.0402

F1 Score: 0.3408 ± 0.0348

Accuracy: 0.8350 ± 0.0110

Precision: 0.2686 ± 0.0270

Sensitivity (Recall): 0.4680 ± 0.0547

Specificity: 0.8718 ± 0.0111

**Support Vector Machine**

Feature indexes (LASSO): [138, 244, 224, 137, 118, 103, 204, 230, 3, 86, 42, 24, 13, 21, 78, 33, 88, 183, 123, 119, 126, 210, 122, 65, 201, 5, 253, 30, 109, 227, 90, 49, 214, 52, 80, 25, 247, 217, 22, 229, 157, 115]

Best Average AUC: 0.7500 ± 0.0193

Average Accuracy: 0.8882 ± 0.0100

Average Precision: 0.3663 ± 0.0599

Average F1 Score: 0.3294 ± 0.0515

Best Hyperparameters:

classifier__C: 10

classifier__class_weight: None

classifier__coef0: 0.5

classifier__degree: 5

classifier__gamma: scale

classifier__kernel: poly

classifier__max_iter: 20000

classifier__tol: 1e-05

sampler: RandomOverSampler

sampler__sampling_strategy: 0.25

=== Optimized Threshold ===

Optimal Threshold (F1-maximizing): 0.2328

Average AUPRC: 0.2746 ± 0.0406

F1 Score: 0.3462 ± 0.0336

Accuracy: 0.8426 ± 0.0128

Precision: 0.2799 ± 0.0322

Sensitivity (Recall): 0.4556 ± 0.0422

Specificity: 0.8814 ± 0.0133

Feature Indexes (relief): [28, 234, 36, 215, 189, 37, 77, 26, 250, 188, 236, 181, 70, 55, 48, 67, 202, 244, 78, 65, 52, 237, 211, 57, 185, 229, 231, 219, 62, 220, 51, 200, 30, 242, 233, 198, 34, 25, 212, 205, 252, 32, 29, 248, 253, 75, 66, 247, 56, 58, 12, 221, 41, 197, 7, 63, 217, 1, 251, 199, 114, 49, 24, 50, 101, 80, 186, 232, 207, 130, 79, 115, 43, 243, 190, 137, 136, 108, 46, 214, 17, 10, 110, 88, 125, 182, 33, 203, 225, 68, 213, 227, 126, 93, 81, 14, 13, 104, 92, 42, 201, 129, 177, 122, 134, 133, 60, 19, 31, 9, 135, 98, 8, 45, 3, 47, 4, 6, 226, 116, 106, 90, 15, 105, 138, 18, 89, 84, 100, 44, 228, 131,

38, 112, 103, 96, 16, 127, 206, 117, 11, 102, 176, 128, 97, 0, 132, 5, 256, 61, 235, 87, 91, 193, 39, 111, 64, 180, 99, 95, 179, 35, 191, 246, 94, 238, 109, 22, 249, 187, 204, 245, 174, 53, 40, 86, 107, 119, 222, 239, 183, 157, 141, 123, 20, 196, 85, 83, 82, 167, 23, 139, 241, 72, 159, 2, 192, 175, 223]

Best Average AUC: 0.7415 ± 0.0316

Average Accuracy: 0.8076 ± 0.0153

Average Precision: 0.2294 ± 0.0283

Average F1 Score: 0.3072 ± 0.0354

Best Hyperparameters:

classifier__C: 10

classifier__class_weight: balanced

classifier__coef0: 1.0

classifier__degree: 4

classifier__gamma: scale

classifier__kernel: poly

classifier__max_iter: 20000

classifier__tol: 1e-05

sampler: SMOTE

sampler__sampling_strategy: 0.5

=== Optimized Threshold ===

Threshold for Maximum F1 Score: 0.56

F1 Score: 0.3231 ± 0.0734

Accuracy: 0.8544 ± 0.0203

Precision: 0.2835 ± 0.0710

**KNN:**

feature_indexes (lasso): [138, 244, 224, 137, 118, 103, 230, 204, 42]

Best Average AUC: 0.7592 ± 0.0266

Average Accuracy: 0.9071 ± 0.0031

Average Precision: 0.4590 ± 0.1331

Average F1 Score: 0.1110 ± 0.0262

Best Hyperparameters:

classifier__algorithm: auto

classifier__leaf_size: 50

classifier__metric: manhattan

classifier__n_neighbors: 30

classifier__weights: distance

oversampler: passthrough

=== Optimized Threshold ===

Optimal Threshold (F1-maximizing): 0.1876

Average AUPRC: 0.2693 ± 0.0404

F1 Score: 0.3312 ± 0.0387

Accuracy: 0.8348 ± 0.0146

Precision: 0.2637 ± 0.0345

Sensitivity (Recall): 0.4467 ± 0.0477

Specificity: 0.8738 ± 0.0138

Feature Indexes (relief): [28, 234, 36, 215, 189, 37, 77, 26, 250, 188, 236, 181, 70, 55, 48, 67, 202, 244, 78, 65, 52, 237, 211, 57, 185, 229, 231, 219, 62, 220, 51, 200, 30, 242, 233, 198, 34, 25, 212, 205, 252, 32, 29, 248, 253, 75, 66, 247, 56, 58, 12, 221, 41, 197, 7, 63, 217, 1, 251, 199, 114, 49, 24, 50, 101, 80, 186, 232, 207, 130, 79, 115, 43, 243, 190, 137, 136, 108, 46, 214, 17, 10, 110, 88, 125, 182, 33, 203, 225, 68, 213, 227, 126, 93, 81, 14, 13, 104, 92, 42, 201, 129, 177, 122, 134, 133, 60, 19, 31, 9, 135, 98, 8, 45, 3, 47, 4, 6, 226, 116, 106, 90, 15]

Best Average AUC: 0.7527 ± 0.0291

Average Accuracy: 0.8963 ± 0.0076

Average Precision: 0.3706 ± 0.0821

Average F1 Score: 0.2485 ± 0.0488

Best Hyperparameters:

classifier__algorithm: auto

classifier__leaf_size: 20

classifier__metric: manhattan

classifier__n_neighbors: 49

classifier__weights: distance

oversampler: ADASYN

oversampler__sampling_strategy: 0.2

=== Optimized Threshold ===

Threshold for Maximum F1 Score: 0.29

F1 Score: 0.3371 ± 0.0445

Accuracy: 0.8466 ± 0.0164

Precision: 0.2804 ± 0.0429

**Naïve Bayes**

Feature Indexes (relief): [28, 234, 36, 215, 189, 37, 77, 26, 250, 188, 236, 181, 70, 55, 48, 67, 202, 244, 78, 65, 52, 237, 211, 57, 185, 229, 231, 219, 62, 220, 51, 200, 30, 242, 233, 198, 34, 25, 212, 205, 252, 32, 29, 248, 253, 75, 66, 247, 56, 58, 12, 221, 41, 197, 7, 63, 217, 1, 251, 199, 114, 49, 24, 50, 101, 80, 186, 232, 207, 130, 79, 115, 43, 243, 190, 137, 136, 108, 46, 214, 17, 10, 110, 88, 125, 182, 33, 203, 225, 68, 213, 227, 126, 93, 81, 14, 13, 104, 92, 42, 201, 129, 177, 122, 134, 133, 60, 19, 31, 9, 135, 98, 8, 45, 3, 47, 4, 6, 226, 116, 106, 90, 15, 105, 138, 18, 89, 84, 100, 44, 228, 131, 38, 112, 103, 96, 16, 127, 206, 117, 11, 102, 176, 128, 97, 0, 132, 5, 256, 61, 235, 87, 91, 193, 39]

Best Average AUC: 0.6977 ± 0.0444

Average Accuracy: 0.6790 ± 0.0257

Average Precision: 0.1623 ± 0.0240

Average F1 Score: 0.2553 ± 0.0357

Best Hyperparameters:

classifier: BernoulliNB()

classifier__alpha: 25.75

classifier__binarize: 0.0

classifier__fit_prior: False

classifier__force_alpha: True

sampler: RandomOverSampler()

sampler__sampling_strategy: 0.5

=== Optimized Threshold ===

Optimal Threshold (F1-maximizing): 0.7810

Average AUPRC: 0.2153 ± 0.0444

F1 Score: 0.2680 ± 0.0392

Accuracy: 0.8330 ± 0.0114

Precision: 0.2237 ± 0.0300

Sensitivity (Recall): 0.3367 ± 0.0590

Specificity: 0.8829 ± 0.0136

**Adaboost:**

Feature Indexes (lasso): [137, 118, 244, 224, 5, 78, 154, 13, 138, 140, 86, 183, 103, 33, 126, 230, 42, 201, 71, 65, 49, 55, 240, 74, 206, 217, 253, 59, 102, 30, 4, 204, 187, 28, 82, 84, 73, 119, 123, 52, 199, 214, 203, 190, 202]

Best Average AUC: 0.7461 ± 0.0251

Average Accuracy: 0.9073 ± 0.0045

Average Precision: 0.4738 ± 0.1492

Average F1 Score: 0.1357 ± 0.0333

Best Hyperparameters:

ada__algorithm: SAMME

ada__estimator__max_depth: 10

ada__estimator__max_features: log2

ada__estimator__min_samples_leaf: 1

ada__estimator__min_samples_split: 12

ada__learning_rate: 0.01

ada__n_estimators: 250

sampler: passthrough

=== Optimized Threshold ===

Threshold for Maximum F1 Score: 0.24

F1 Score: 0.3279 ± 0.0425

Accuracy: 0.8415 ± 0.0158

Precision: 0.2690 ± 0.0402

Feature Indexes (relief): [28, 234, 36, 215, 189, 37, 77, 26, 250, 188, 236, 181, 70, 55, 48, 67, 202, 244, 78, 65, 52, 237, 211, 57, 185, 229, 231, 219, 62, 220, 51, 200, 30, 242, 233, 198, 34, 25, 212, 205, 252, 32, 29, 248, 253, 75, 66, 247, 56, 58, 12, 221, 41, 197, 7, 63, 217, 1, 251, 199, 114, 49, 24, 50, 101, 80, 186, 232, 207, 130, 79, 115, 43, 243, 190, 137, 136, 108, 46, 214, 17, 10, 110, 88, 125, 182, 33, 203, 225, 68, 213, 227, 126, 93, 81, 14, 13, 104, 92, 42, 201, 129, 177, 122, 134, 133, 60, 19, 31, 9, 135, 98, 8, 45, 3, 47, 4, 6, 226, 116, 106, 90, 15, 105, 138, 18, 89, 84, 100, 44, 228, 131, 38, 112, 103, 96, 16, 127, 206, 117, 11, 102, 176, 128, 97, 0, 132, 5, 256, 61, 235, 87, 91, 193, 39, 111, 64, 180, 99]

Best Average AUC: 0.7811 ± 0.0248

Average Accuracy: 0.8855 ± 0.0090

Average Precision: 0.3202 ± 0.0554

Average F1 Score: 0.2616 ± 0.0476

Best Hyperparameters:

ada__algorithm: SAMME

ada__estimator__max_depth: 20

ada__estimator__max_features: sqrt

ada__estimator__min_samples_leaf: 2

ada__estimator__min_samples_split: 15

ada__learning_rate: 1e-05

ada__n_estimators: 250

sampler: ADASYN

sampler__sampling_strategy: 0.2

=== Optimized Threshold ===

Optimal Threshold (F1-maximizing): 0.2399

Average AUPRC: 0.2664 ± 0.0432

F1 Score: 0.3195 ± 0.0309

Accuracy: 0.7973 ± 0.0166

Precision: 0.2310 ± 0.0257

Sensitivity (Recall): 0.5194 ± 0.0384

Specificity: 0.8252 ± 0.0163

**Gradient boosting:**

Feature Indexes (lasso): [138, 244, 224, 137, 118, 103, 230, 204, 42, 13, 86, 3]

Best Average AUC: 0.7590 ± 0.0283

Average Accuracy: 0.8796 ± 0.0147

Average Precision: 0.3179 ± 0.0816

Average F1 Score: 0.2908 ± 0.0708

Best Hyperparameters:

gbc__learning_rate: 0.01

gbc__loss: exponential

gbc__max_depth: 20

gbc__max_features: log2

gbc__min_samples_leaf: 2

gbc__min_samples_split: 5

gbc__n_estimators: 125

gbc__subsample: 0.1

resampler: RandomOverSampler

resampler__sampling_strategy: 0.5

=== Optimized Threshold ===

Optimal Threshold (F1-maximizing): 0.4279

Average AUPRC: 0.2684 ± 0.0452

F1 Score: 0.3059 ± 0.0500

Accuracy: 0.8536 ± 0.0138

Precision: 0.2709 ± 0.0450

Sensitivity (Recall): 0.3544 ± 0.0654

Specificity: 0.9037 ± 0.0147

Feature Indexes (relief): [28, 234, 36, 215, 189, 37, 77, 26, 250, 188, 236, 181, 70, 55, 48, 67, 202, 244, 78, 65, 52, 237, 211, 57, 185, 229, 231, 219, 62, 220, 51, 200, 30, 242, 233, 198, 34, 25, 212, 205, 252, 32, 29, 248, 253, 75, 66, 247, 56, 58, 12]

Best Average AUC: 0.7332 ± 0.0428

Average Accuracy: 0.8313 ± 0.0163

Average Precision: 0.2460 ± 0.0176

Average F1 Score: 0.2653 ± 0.0282

Best Hyperparameters:

gbc__learning_rate: 0.1

gbc__loss: exponential

gbc__max_depth: 5

gbc__max_features: sqrt

gbc__min_samples_leaf: 1

gbc__min_samples_split: 2

gbc__n_estimators: 100

gbc__subsample: 0.8

resampler: RandomOverSampler

resampler__sampling_strategy: auto

=== Optimized Threshold ===

Threshold for Maximum F1 Score: 0.29

F1 Score: 0.2893 ± 0.0596

Accuracy: 0.8272 ± 0.0170

Precision: 0.2322 ± 0.0460

**MLP:**

Feature Indexes (lasso): [137, 118, 244, 224, 78, 5, 154, 13, 138, 140, 86, 183, 103, 33, 126, 230, 42, 201, 71, 65, 49, 55, 240, 253, 74, 206, 217, 59, 30, 102, 204, 4, 187, 28, 82, 73, 119, 84, 123, 52, 199, 214, 203, 190, 24, 202, 3, 44, 251, 61, 189, 0, 255, 182, 134, 9, 209]

Best Average AUC: 0.7429 ± 0.0421

Average Accuracy: 0.8102 ± 0.0172

Average Precision: 0.2322 ± 0.0205

Average F1 Score: 0.2724 ± 0.0319

Best Hyperparameters:

classifier__activation: tanh

classifier__alpha: 0.0001

classifier__batch_size: 64

classifier__early_stopping: False

classifier__hidden_layer_sizes: (50, 100, 50)

classifier__learning_rate: adaptive

classifier__learning_rate_init: 0.001

classifier__max_iter: 500

classifier__solver: sgd

sampler: RandomOverSampler

sampler__sampling_strategy: 1.0

=== Optimized Threshold ===

Threshold for Maximum F1 Score: 0.64

F1 Score: 0.3000 ± 0.0441

Accuracy: 0.8057 ± 0.0154

Precision: 0.2239 ± 0.0339

Feature Indexes (relief): [28, 234, 36, 215, 189, 37, 77, 26, 250, 188, 236, 181, 70, 55, 48, 67, 202, 244, 78, 65, 52, 237, 211, 57, 185, 229, 231, 219, 62, 220, 51, 200, 30, 242, 233, 198, 34, 25, 212, 205, 252, 32, 29, 248, 253, 75, 66, 247, 56, 58, 12, 221, 41, 197, 7, 63, 217, 1, 251, 199, 114, 49, 24, 50, 101, 80, 186, 232, 207, 130, 79, 115, 43, 243, 190, 137, 136, 108, 46, 214, 17, 10, 110, 88, 125, 182, 33, 203, 225, 68, 213, 227, 126, 93, 81, 14, 13, 104, 92, 42, 201, 129, 177, 122, 134, 133, 60, 19, 31, 9, 135, 98, 8, 45, 3, 47, 4, 6, 226, 116, 106, 90, 15, 105, 138, 18, 89, 84, 100, 44, 228, 131, 38, 112, 103, 96, 16, 127, 206, 117, 11, 102, 176, 128]

Best Average AUC: 0.7455 ± 0.0406

Average Accuracy: 0.8225 ± 0.0191

Average Precision: 0.2445 ± 0.0234

Average F1 Score: 0.2795 ± 0.0360

Best Hyperparameters:

classifier__activation: tanh

classifier__alpha: 1e-05

classifier__batch_size: auto

classifier__early_stopping: False

classifier__hidden_layer_sizes: (100,)

classifier__learning_rate: constant

classifier__learning_rate_init: 0.0001

classifier__max_iter: 500

classifier__solver: adam

sampler: RandomOverSampler

sampler__sampling_strategy: auto

=== Optimized Threshold ===

Optimal Threshold (F1-maximizing): 0.7083

Average AUPRC: 0.2634 ± 0.0596

F1 Score: 0.3209 ± 0.0543

Accuracy: 0.8492 ± 0.0189

Precision: 0.2754 ± 0.0517

Sensitivity (Recall): 0.3881 ± 0.0651

Specificity: 0.8955 ± 0.0182

**Bayesian Network:**

Feature Indexes (relief): [29, 15, 10, 5, 11, 36, 12, 3, 38, 33, 2, 16, 14, 13, 4, 7, 6, 24]

Best Hyperparameters Identified:

scoring_method BIC

max_parents NaN

oversampler passthrough

sampling_rate NaN

avg_auc 0.649448

std_auc 0.038856

AUPRC: 0.1344 ± 0.0197

Optimal Threshold: 0.1302

F1 Score: 0.2387 ± 0.0383

Accuracy: 0.6615 ± 0.0878

Precision: 0.1533 ± 0.0294

Sensitivity: 0.5677 ± 0.1088

Specificity: 0.6710 ± 0.1037

**Logistic Regression:**

Feature Indexes (lasso): [224, 3, 204, 138, 88, 21, 183, 22, 227, 142, 24, 244, 13, 103, 119, 184, 86, 26, 38, 155, 229, 222, 122, 253, 126, 174, 25, 82, 77, 137, 157, 182, 42, 216, 49, 232, 115, 133, 196, 231, 217, 90, 210, 116, 202, 59, 156, 109, 65, 20, 52, 201, 113, 68, 243, 214, 206, 185, 57, 197, 78, 64, 1, 221, 69, 80, 91, 11, 176, 29, 35, 123, 117, 33, 67, 147, 215, 61, 154, 207, 51, 252, 62, 190, 179, 30, 124, 167, 148, 28, 203, 168, 53, 146, 5]

Best Average AUC: 0.7620 ± 0.0274

Average Accuracy: 0.8358 ± 0.0153

Average Precision: 0.2622 ± 0.0429

Average F1 Score: 0.3272 ± 0.0494

Best Hyperparameters:

lr__C: 1

lr__l1_ratio: 0.3

lr__penalty: elasticnet

lr__solver: saga

resampler: RandomOverSampler()

resampler__sampling_strategy: 0.5

=== Optimized Threshold ===

Optimal Threshold (F1-maximizing): 0.5458

Average AUC: 0.7505 ± 0.0327

Average AUPRC: 0.2759 ± 0.0592

F1 Score: 0.3278 ± 0.0551

Accuracy: 0.8565 ± 0.0156

Precision: 0.2883 ± 0.0523

Sensitivity (Recall): 0.3826 ± 0.0650

Specificity: 0.9040 ± 0.0149

Feature Indexes (relief): [28, 234, 36, 215, 189, 37, 77, 26, 250, 188, 236, 181, 70, 55, 48, 67, 202, 244, 78, 65, 52, 237, 211, 57, 185, 229, 231, 219, 62, 220, 51, 200, 30, 242, 233, 198, 34, 25, 212, 205, 252, 32, 29, 248, 253, 75, 66, 247, 56, 58, 12, 221, 41, 197, 7, 63, 217, 1, 251, 199, 114, 49, 24, 50, 101, 80, 186, 232, 207, 130, 79, 115, 43, 243, 190, 137, 136, 108, 46, 214, 17, 10, 110, 88, 125, 182, 33, 203, 225, 68, 213, 227, 126, 93, 81, 14, 13, 104, 92, 42, 201, 129, 177, 122, 134, 133, 60, 19, 31, 9, 135, 98, 8, 45, 3, 47, 4, 6, 226, 116, 106, 90, 15, 105, 138, 18, 89, 84, 100, 44, 228, 131, 38, 112, 103, 96, 16, 127, 206, 117, 11, 102, 176, 128, 97, 0, 132, 5, 256, 61, 235, 87, 91, 193, 39, 111, 64, 180, 99, 95, 179, 35, 191, 246, 94, 238, 109, 22, 249, 187, 204, 245, 174, 53, 40, 86, 107, 119, 222, 239, 183, 157, 141, 123, 20, 196, 85, 83, 82, 167, 23, 139, 241, 72, 159, 2, 192, 175, 223, 54, 156, 73, 69, 208, 161, 120, 195, 158, 166, 59, 153, 147, 160, 172, 216, 173, 165, 76, 27, 178, 74, 209, 255, 224, 124, 194, 164, 171, 155, 254, 184, 113, 143, 71, 230, 144, 146, 210, 21, 240, 170, 218, 149, 163, 154]

Best Average AUC: 0.7421 ± 0.0375

Average Accuracy: 0.6895 ± 0.0138

Average Precision: 0.1757 ± 0.0171

Average F1 Score: 0.2766 ± 0.0268

Best Hyperparameters:

lr__C: 1

lr__l1_ratio: None

lr__penalty: l1

lr__solver: saga

resampler: RandomOverSampler()

resampler__sampling_strategy: auto

=== Optimized Threshold ===

Threshold for Maximum F1 Score: 0.70

F1 Score: 0.3072 ± 0.0529

Accuracy: 0.8491 ± 0.0130

Precision: 0.2648 ± 0.0458

**TSNN:**

Global relief:

Best Trial:

AUC: 0.7531640655561265 ± 0.0330

Params:

n_genotype: 68

n_history: 1

n_phenotype: 59

n_behaviour: 7

learning_rate: 0.0023343464127852007

epochs: 1568

batch_size: 128

AUC: 0.7532 ± 0.0330

F1 Score: 0.1198 ± 0.0304

Accuracy: 0.9055 ± 0.0055

Precision: 0.4348 ± 0.1277

Optimal Threshold: 0.1869

AUC: 0.7405 ± 0.0223

AUPRC: 0.2633 ± 0.0342

F1 Score: 0.3292 ± 0.0394

Accuracy: 0.8340 ± 0.0205

Precision: 0.2633 ± 0.0379

Sensitivity (Recall): 0.4430 ± 0.0438

Specificity: 0.8732 ± 0.0217

separate relief:

Best Trial:

AUC:  0.752471120396032

parameters: {'n_genotype': 1, 'n_history': 7, 'n_phenotype': 52, 'n_behaviour': 4,

'learning_rate': 0.0051175419520232135,

'epochs': 2654,

'batch_size': 256}

AUC: 0.7525 ± 0.0275

F1 Score: 0.1247 ± 0.0445

Accuracy: 0.9065 ± 0.0050

Precision: 0.4666 ± 0.1263

Best Threshold to maximize F1 Score: 0.19

--- Per-Fold Metrics after Threshold Optimization ---

F1 Score: 0.3220 ± 0.0518

Accuracy: 0.8379 ± 0.0171

Precision: 0.2617 ± 0.0450

Global lasso:

Best Trial:

AUC: 0.7417 ± 0.0357

F1 Score: 0.0456 (Std: 0.0528)

Accuracy: 0.9075 (Std: 0.0046)

Precision: 0.3000 (Std: 0.3253)

Params:

n_genotype: 36

n_history: 4

n_phenotype: 8

n_behaviour: 2

learning_rate: 0.00013536078428508602

epochs: 2045

batch_size: 16

Best Threshold to maximize F1 Score: 0.60

F1 Score: 0.2816 ± 0.0400

Accuracy: 0.7588 ± 0.0279

Precision: 0.1948 ± 0.0325

Separate lasso:

Best Trial:

AUC: 0.7311 ± 0.0484

F1 Score: 0.0775 (Std: 0.0450)

Accuracy: 0.9047 (Std: 0.0062)

Precision: 0.3852 (Std: 0.2947)

Params:

n_genotype: 89

n_history: 11

n_phenotype: 49

n_behaviour: 18

learning_rate: 1.0102030709795042e-05

epochs: 1640

batch_size: 32

Best Threshold to maximize F1 Score: 0.22

F1 Score: 0.2978 ± 0.0456

Accuracy: 0.8641 ± 0.0117

Precision: 0.2832 ± 0.0464

**TSGNN:**

Global relief:

Value (Mean AUC): 0.7532256092485068 ± 0.0225

Params:

n_epochs: 2059

lr: 1.185885489910591e-05

weight_decay: 9.916744984817729e-07

batch_size: 64

n_genotype: 125

n_history: 3

n_phenotype: 44

n_behaviour: 20

Best Trial Metrics:

Selected Features: ['rs591058', 'rs1800797', 'rs13946', 'rs25487', 'rs2228570', 'rs12722', 'rs2104772', 'rs1144393', 'rs3196378', 'rs1544410', 'rs2237352', 'rs1137101', 'rs10263021', 'rs4454832', 'rs7528684', 'rs2234693', 'rs10132091', 'rs7035322', 'rs1330363', 'rs1011814', 'rs1249269', 'rs4725069', 'rs820218', 'rs62051384', 'rs11232681', 'rs4701616', 'rs1800629', 'rs11177', 'rs143383', 'rs6617', 'rs6481512', 'rs17756404', 'rs4986938', 'rs13317', 'rs4730153', 'rs911263', 'rs970547', 'rs11225395', 'rs3018362', 'rs11629171', 'rs10992075', 'rs4789932', 'rs2252070', 'rs3789870', 'rs1590', 'rs42531', 'rs11154027', 'rs10759753', 'rs2761884', 'rs4362400', 'rs3219008', 'rs3218791', 'rs17576', 'rs2289360', 'rs1134170', 'rs2525504', 'rs4919510', 'rs1937810', 'rs1718119', 'rs10484958', 'rs2285053', 'class12_SNP_risk_score', 'rs1800972', 'rs4328262', 'rs1800469', 'rs2306033', 'rs9340799', 'rs17583842', 'rs4244032', 'rs1643821', 'rs1800470', 'rs3045', 'rs4903399', 'rs3753841', 'rs2277698', 'rs1800012', 'rs12656106', 'rs187483', 'class1_SNP_risk_score', 'rs2281518', 'sex', 'class123_SNP_risk_score', 'rs1045485', 'rs1554606', 'rs78391032', 'rs2305948', 'rs4654760', 'rs1887632', 'rs1800795', 'rs1021188', 'rs72758637', 'rs12154667', 'rs1138545', 'rs3751143', 'rs1676303', 'rs7021589', 'rs12574452', 'rs13107325', 'rs1548456', 'rs2277268', 'rs912336', 'rs35360670', 'rs420257', 'rs74544784', 'rs60713544', 'rs12429486', 'rs42517', 'rs2010963', 'rs71404070', 'rs117544024', 'rs2586488', 'rs25489', 'rs413826', 'rs650108', 'rs57104447', 'rs42522', 'rs710079', 'rs761804508', 'rs145648292', 'rs77569527', 'rs144371252', 'rs4988321', 'rs149047058', 'rs144414988', 'rs2858056', 'Age', 'tracking_period_injury', 'Athlete_Score', 'Q_angle', 'hip_abduction_peak_torque_asymmetry', 'total_ad_ab_ratio', 'Step_frequency_10', 'thigh_ffmi', 'hip_adduction_peak_torque_asymmetry', 'BMD_hip', 'calf_size', 'Flight_time_12', 'total_fl_ex_ratio', 'navicular_drop', 'Step_frequency_12', 'knee_extension_peak_angle_asymmetry', 'VALR_10', 'VALR_12',

'Q_angle_asymmetry', 'VILR_12', 'VILR_10', 'hip_adduction_peak_torque', 'thigh_lean_mass', 'total_lean_mass', 'leg_ffmi', 'BMD_spine', 'knee_flexion_peak_torque', 'total_ffmi', 'Flight_time_10', 'knee_extension_peak_torque', 'knee_flexion_peak_torque_asymmetry', 'leg_lean_mass', 'hip_abduction_peak_torque', 'VALR_asymmetry_12', 'knee_flexion_peak_angle_asymmetry', 'Impact_peak_12', 'VILR_asymmetry_12', 'BMD_body', 'BMI', 'knee_flexion_peak_angle', 'hip_adduction_peak_angle', 'Duty_factor_10', 'knee_extension_peak_torque_asymmetry', 'lower_leg_lean_mass', 'Duty_factor_asymmetry_12', 'Duty_factor_asymmetry_10', 'Impact_peak_10', 'fat_intake_BW', 'fat_percentage_avg', 'fat_intake_avg', 'glycine_intake_BW', 'arginine_intake_BW', 'calcium_intake_BW', 'average_energy_availability', 'protein_intake_BW', 'SC_past_season', 'SC_past_month', 'vitaminD_intake_BW', 'resistance_training_past_season', 'past_month_volume_low', 'copper_intake_BW', 'past_month_distance', 'drills_past_season', 'non_running_past_season', 'past_month_min', 'bodyweight_exercises_past_season', 'non_running_past_month']

Average AUC: 0.7532 ± 0.0225

Average F1 Score: 0.2214 ± 0.0522

Average Accuracy: 0.9073 ± 0.0063

Average Precision: 0.4877 ± 0.1276

Optimal Threshold: 0.1899

AUPRC: 0.2876 ± 0.0519

F1 Score: 0.3282 ± 0.0515

Accuracy: 0.8403 ± 0.0183

Precision: 0.2681 ± 0.0461

Sensitivity (Recall): 0.4253 ± 0.0611

Specificity: 0.8819 ± 0.0171

Separate relief:

Best trial:

Value (Mean AUC): 0.7530759131312909

Params:

n_epochs: 730

lr: 4.86593416133986e-05

weight_decay: 0.0003438008978072548

batch_size: 256

n_genotype: 126

n_history: 10

n_phenotype: 32

n_behaviour: 9

Best Trial Metrics:

Selected Features: ['rs591058', 'rs2104772', 'rs25487', 'rs1249269', 'rs1800797', 'rs1137101', 'rs1330363', 'rs1144393', 'rs7528684', 'rs4701616', 'rs10132091', 'rs2237352', 'rs143383', 'rs4789932', 'rs2228570', 'rs13946', 'rs10263021', 'rs6481512', 'rs2234693', 'rs6617', 'rs4730153', 'rs7035322', 'rs11177', 'rs12722', 'rs820218', 'rs17756404', 'rs3018362', 'rs4725069', 'rs13317', 'rs4454832', 'rs3196378', 'rs1544410', 'rs62051384', 'rs970547', 'rs4986938', 'rs42531', 'rs17576', 'rs11232681', 'rs911263', 'rs10992075', 'rs1011814', 'rs2252070', 'rs11225395', 'rs11154027', 'rs1800629', 'rs1590', 'rs2761884', 'rs1800469', 'rs4362400', 'rs2525504', 'rs4903399', 'rs3219008', 'rs1800972', 'rs1937810', 'rs3753841', 'rs4919510', 'rs10759753', 'rs3045', 'rs4328262', 'rs1718119', 'rs2306033', 'rs3789870', 'rs4244032', 'rs11629171', 'rs9340799', 'rs2285053', 'rs2289360', 'rs3218791', 'rs17583842', 'rs1800470', 'rs2281518', 'rs10484958', 'class12_SNP_risk_score', 'rs1643821', 'rs12656106', 'sex', 'rs2277698', 'rs1134170', 'class1_SNP_risk_score', 'rs187483', 'rs1800012', 'rs1045485', 'rs1554606', 'class123_SNP_risk_score', 'rs72758637', 'rs1887632', 'rs2305948', 'rs1676303', 'rs1800795', 'rs12154667', 'rs4654760', 'rs1138545', 'rs1021188', 'rs78391032', 'rs3751143', 'rs7021589', 'rs1548456', 'rs2277268', 'rs13107325', 'rs12574452', 'rs912336', 'rs420257', 'rs35360670', 'rs42517', 'rs2010963', 'rs2586488', 'rs60713544', 'rs74544784', 'rs71404070', 'rs12429486', 'rs25489', 'rs117544024', 'rs650108', 'rs761804508', 'rs57104447', 'rs145648292', 'rs42522', 'rs710079', 'rs413826', 'rs4988321', 'rs77569527', 'rs144371252', 'rs144414988', 'rs149047058', 'rs2858056', 'rs3216902', 'Age', 'Athlete_Score', 'average_run_hours', 'average_run_frequency', 'average_interval_training_frequency', 'EDEQ_total', 'LEAF-Q', 'tracking_period_injury', 'lower_limb_days_total', 'past_stress_injury', 'Duty_factor_10', 'Flight_time_10', 'Step_frequency_10', 'Contact_time_10', 'Q_angle_asymmetry', 'Impact_peak_10', 'Cadence_asymmetry_10', 'total_ad_ab_ratio', 'knee_extension_peak_torque', 'hip_abduction_peak_torque', 'hip_adduction_peak_torque', 'Impact_peak_12', 'Duty_factor_12', 'Q_angle', 'Flight_time_12', 'BMI', 'BMD_body', 'knee_flexion_peak_angle', 'knee_flexion_peak_angle_asymmetry', 'hip_adduction_peak_angle', 'Contact_time_12', 'knee_flexion_peak_torque', 'hip_abduction_peak_torque_asymmetry', 'Impact_peak_asymmetry_10', 'knee_flexion_peak_torque_asymmetry', 'Duty_factor_asymmetry_10', 'Cadence_asymmetry_12', 'knee_extension_peak_angle_asymmetry', 'knee_extension_peak_torque_asymmetry', 'knee_extension_peak_angle', 'BMD_hip', 'height', 'fat_intake_BW', 'fat_percentage_avg', 'fat_intake_avg', 'arginine_intake_BW', 'protein_intake_BW', 'average_energy_availability', 'glycine_intake_BW', 'calcium_intake_BW', 'past_month_distance']

Average AUC: 0.7531 ± 0.0378

Average F1 Score: 0.0925 ± 0.0424

Average Accuracy: 0.9068 ± 0.0033

Average Precision: 0.4604 ± 0.2347

Average F1 Score: 0.3619 ± 0.0468

Average Accuracy: 0.8562 ± 0.0304

Average Precision: 0.3162 ± 0.0572

Global lasso:

Best trial:

Value (Mean AUC): 0.7503470913074771

Params:

n_epochs: 1142

lr: 3.5784877937744905e-05

weight_decay: 8.830400761450904e-08

batch_size: 128

n_genotype: 54

n_history: 6

n_phenotype: 12

n_behaviour: 7

Best Trial Metrics:

Selected Features: ['rs7035322', 'rs145648292', 'rs1330363', 'rs144414988', 'rs9340799', 'rs1676303', 'rs2277268', 'rs1011814', 'rs4903399', 'rs1590', 'rs4986938', 'rs4919510', 'rs149047058', 'rs2289360', 'rs1800469', 'rs1249269', 'rs2281518', 'rs2586488', 'rs4454832', 'rs42522', 'rs3216902', 'rs2858056', 'rs591058', 'rs4362400', 'rs10132091', 'rs1937810', 'rs2306033', 'rs1643821', 'rs10484958', 'rs1045485', 'rs42517', 'rs4701616', 'rs2525504', 'rs761804508', 'rs1134170', 'rs11225395', 'rs10759753', 'sex', 'rs1800629', 'rs1144393', 'rs3045', 'rs17583842', 'rs1800972', 'rs4328262', 'rs2234693', 'rs3751143', 'rs1800470', 'rs4988321', 'rs2228570', 'rs78391032', 'rs12656106', 'rs60713544', 'rs143383', 'rs62051384', 'EDEQ_total', 'tracking_period_injury', 'average_run_hours', 'Athlete_Score', 'Age', 'average_interval_training_frequency', 'BMD_body', 'BMD_hip', 'Duty_factor_asymmetry_10', 'Q_angle_asymmetry', 'hip_adduction_peak_torque', 'ad_ab_ratio_asymmetry', 'knee_extension_peak_torque_asymmetry', 'knee_extension_peak_angle_asymmetry', 'knee_flexion_peak_torque', 'hip_abduction_peak_angle', 'knee_flexion_peak_angle_asymmetry', 'navicular_drop_asymmetry', 'omega3_intake_BW', 'copper_intake_BW', 'arginine_intake_BW', 'vitaminD_intake_BW', 'past_month_ratio', 'past_week_ratio_calculated_volume', 'calcium_intake_BW']

Average AUC: 0.7503 ± 0.0355

Average F1 Score: 0.0385 ± 0.0357

Average Accuracy: 0.9062 ± 0.0030

Average Precision: 0.2822 ± 0.2255

Average F1 Score: 0.3472 ± 0.0537

Average Accuracy: 0.8403 ± 0.0495

Average Precision: 0.3085 ± 0.0972

Separate lasso:

Best trial:

Value (Mean AUC): 0.7415440699513642

Params:

n_epochs: 2163

lr: 4.600641941225531e-06

weight_decay: 3.128639842545911e-08

batch_size: 256

n_genotype: 67

n_history: 4

n_phenotype: 47

n_behaviour: 37

Best Trial Metrics:

Selected Features: ['rs144414988', 'rs117544024', 'rs77569527', 'rs78391032', 'rs12722', 'rs3196378', 'rs57104447', 'rs144371252', 'class123_SNP_risk_score', 'rs11177', 'rs2277268', 'rs6617', 'rs4988321', 'rs1676303', 'rs25487', 'rs11154027', 'rs35360670', 'rs6481512', 'rs710079', 'class12_SNP_risk_score', 'rs420257', 'rs42522', 'rs71404070', 'rs1249269', 'rs591058', 'rs72758637', 'rs413826', 'rs74544784', 'rs1330363', 'rs25489', 'rs149047058', 'rs3218791', 'rs4903399', 'rs10263021', 'rs761804508', 'rs2234693', 'rs7528684', 'rs17576', 'rs12154667', 'rs2858056', 'rs17583842', 'rs1137101', 'rs2289360', 'rs2525504', 'rs4701616', 'rs1554606', 'rs1887632', 'rs820218', 'rs2252070', 'rs143383', 'rs1011814', 'rs2586488', 'rs42517', 'rs2277698', 'rs4919510', 'rs3753841', 'rs2104772', 'rs13107325', 'rs2306033', 'rs1800012', 'rs1800972', 'rs11232681', 'rs1800469', 'rs2285053', 'rs12574452', 'rs62051384', 'rs3045', 'tracking_period_injury', 'past_month_injury', 'average_run_hours', 'average_run_frequency', 'Mass', 'BMI', 'VILR_asymmetry_10', 'height', 'VALR_asymmetry_10', 'VALR_10', 'calf_size', 'VILR_10', 'ad_ab_ratio_asymmetry', 'VALR_asymmetry_12', 'Duty_factor_10', 'Impact_peak_12', 'VILR_asymmetry_12', 'Duty_factor_12', 'hip_abduction_peak_torque', 'knee_extension_peak_torque', 'lower_leg_lean_mass', 'knee_extension_peak_angle', 'hip_abduction_peak_torque_asymmetry', 'hip_adduction_peak_torque', 'Step_frequency_12', 'Step_frequency_10', 'hip_adduction_peak_angle_asymmetry', 'knee_flexion_peak_torque', 'Impact_peak_asymmetry_12', 'Impact_peak_10', 'Q_angle', 'total_lean_mass', 'Flight_time_12', 'leg_ffmi', 'total_fl_ex_ratio', 'hip_abduction_peak_angle', 'total_ffmi', 'hip_abduction_peak_angle_asymmetry', 'hip_adduction_peak_torque_asymmetry', 'Alt_strike', 'knee_flexion_peak_angle', 'Q_angle_asymmetry', 'navicular_drop', 'Cadence_asymmetry_10', 'Impact_peak_asymmetry_10', 'Contact_time_10', 'fl_ex_ratio_asymmetry', 'knee_extension_peak_torque_asymmetry', 'total_ad_ab_ratio', 'lower_leg_ffmi', 'Cadence_asymmetry_12', 'past_week_ratio_calculated_volume', 'past_week_ratio', 'arginine_intake_BW', 'glycine_intake_BW', 'stretching_past_season', 'barefoot_past_season', 'past_week_ratio_low', 'circuit_training_past_month', 'past_month_ratio', 'resistance_training_past_season', 'stretching_past_month', 'past_month_distance', 'drills_past_season', 'past_month_ratio_calculated_volume', 'vitaminE_intake_BW', 'past_month_volume_very_high', 'circuit_training_past_season', 'protein_intake_BW', 'iron_intake_BW', 'vitaminD_intake_BW', 'resistance_training_past_month', 'barefoot_past_month', 'past_month_min', 'past_month_volume_low', 'SC_past_month', 'past_week_ratio_moderate', 'vitaminC_intake_BW', 'past_week_ratio_high', 'non_running_past_season', 'fat_percentage_avg', 'non_running_past_month', 'core_stability_past_season', 'calcium_intake_BW', 'omega3_intake_BW', 'past_month_calculated_volume', 'drills_past_month', 'plyometrics_past_season']

Average AUC: 0.7415 ± 0.0366

Average F1 Score: 0.0352 ± 0.0355

Average Accuracy: 0.9068 ± 0.0016

Average Precision: 0.2119 ± 0.1837

Average F1 Score: 0.3479 ± 0.0506

Average Accuracy: 0.8267 ± 0.1040

Average Precision: 0.3057 ± 0.0855

### Selected Feature Names

### Class 1

#### Decision Tree

rs12722, rs4986938, rs11225395, rs1144393, rs2252070, rs591058, rs4789932, class1_SNP_risk_score, rs13946, navicular_drop

#### Random Forest

tracking_period_injury, past_month_distance, Duty_factor_12, Impact_peak_12, Q_angle, average_run_hours, navicular_drop, Q_angle_asymmetry, past_month_ratio, knee_flexion_peak_torque, SC_past_season, rs591058, rs2252070, BMD_spine, class1_SNP_risk_score, lower_limb_days_total, hip_abduction_peak_torque, rs1144393, rs1800012, EDEQ_total, rs9340799, navicular_drop_asymmetry, rs1800795, rs970547, Age, knee_extension_peak_torque, rs13946, rs4789932, rs4986938, non_running_past_season, VALR_12, sex, average_interval_training_frequency, rs650108, rs12722, BMI, fat_intake_avg

**SVM**

rs12722, rs4986938, rs11225395, rs1144393, rs2252070, rs591058, rs4789932, class1_SNP_risk_score, rs13946, navicular_drop, total_ad_ab_ratio, Q_angle_asymmetry, rs9340799, Q_angle, rs970547, tracking_period_injury, VALR_12, BMI, average_run_hours, Impact_peak_12, hip_abduction_peak_torque, Duty_factor_12, knee_extension_peak_torque, navicular_drop_asymmetry, fat_intake_avg, average_interval_training_frequency, Age, knee_flexion_peak_torque, BMD_spine, rs1800012, EDEQ_total, sex, past_month_distance, rs1800795, SC_past_season, non_running_past_season, lower_limb_days_total

**KNN**

rs12722, rs4986938, rs11225395, rs1144393, rs2252070, rs591058, rs4789932, class1_SNP_risk_score, rs13946, navicular_drop, total_ad_ab_ratio, Q_angle_asymmetry, rs9340799, Q_angle, rs970547, tracking_period_injury, VALR_12, BMI, average_run_hours, Impact_peak_12, hip_abduction_peak_torque, Duty_factor_12, knee_extension_peak_torque, navicular_drop_asymmetry, fat_intake_avg, average_interval_training_frequency, Age, knee_flexion_peak_torque, BMD_spine, rs1800012, EDEQ_total, sex, past_month_distance, rs1800795, SC_past_season, non_running_past_season

**Naïve Bayes**

rs12722, rs4986938, rs11225395, rs1144393, rs2252070, rs591058, rs4789932, class1_SNP_risk_score, rs13946, navicular_drop, total_ad_ab_ratio, Q_angle_asymmetry, rs9340799, Q_angle, rs970547, tracking_period_injury, VALR_12, BMI, average_run_hours, Impact_peak_12, hip_abduction_peak_torque

**Adaboost**

rs12722, rs4986938, rs11225395, rs1144393, rs2252070, rs591058, rs4789932, class1_SNP_risk_score, rs13946, navicular_drop, total_ad_ab_ratio, Q_angle_asymmetry, rs9340799, Q_angle, rs970547, tracking_period_injury, VALR_12, BMI, average_run_hours, Impact_peak_12, hip_abduction_peak_torque, Duty_factor_12, knee_extension_peak_torque, navicular_drop_asymmetry

**Gradient Boosting**

tracking_period_injury, past_month_distance, Duty_factor_12, Impact_peak_12, Q_angle, average_run_hours, navicular_drop, Q_angle_asymmetry, past_month_ratio, knee_flexion_peak_torque, SC_past_season, rs591058, rs2252070, BMD_spine, class1_SNP_risk_score, lower_limb_days_total, hip_abduction_peak_torque, rs1144393, rs1800012, EDEQ_total, rs9340799, navicular_drop_asymmetry, rs1800795, rs970547, Age, knee_extension_peak_torque, rs13946

**MLP**

rs12722, rs4986938, rs11225395, rs1144393, rs2252070, rs591058, rs4789932, class1_SNP_risk_score, rs13946, navicular_drop, total_ad_ab_ratio, Q_angle_asymmetry, rs9340799, Q_angle, rs970547, tracking_period_injury, VALR_12, BMI, average_run_hours, Impact_peak_12, hip_abduction_peak_torque, Duty_factor_12, knee_extension_peak_torque, navicular_drop_asymmetry, fat_intake_avg, average_interval_training_frequency, Age, knee_flexion_peak_torque, BMD_spine, rs1800012, EDEQ_total, sex, past_month_distance, rs1800795, SC_past_season, non_running_past_season

**Bayesian Network**

tracking_period_injury, rs4986938, rs591058, knee_flexion_peak_torque, average_run_hours, Q_angle, Q_angle_asymmetry, knee_extension_peak_torque, lower_limb_days_total, navicular_drop, total_ad_ab_ratio, hip_abduction_peak_torque, rs650108, rs11225395, rs13946, past_month_distance, rs9340799, BMD_spine, Impact_peak_12

**Logistic Regression**

tracking_period_injury, class1_SNP_risk_score, past_month_distance, Duty_factor_12, Impact_peak_12, Q_angle, average_run_hours, navicular_drop, Q_angle_asymmetry, past_month_ratio, knee_flexion_peak_torque, SC_past_season, rs1144393, rs9340799, BMD_spine, lower_limb_days_total, rs1800795, rs4789932, rs591058, hip_abduction_peak_torque, rs4986938

**TSNN**

'rs2252070', 'rs4986938', 'rs12722', 'rs1144393', 'rs591058', 'rs4789932', 'rs11225395', 'rs13946', 'class1_SNP_risk_score', 'rs9340799', 'rs970547', 'rs1800012', 'sex', 'average_run_hours', 'Age', 'average_interval_training_frequency', 'BMD_spine', 'Q_angle_asymmetry', 'Q_angle', 'Impact_peak_12', 'navicular_drop', 'Duty_factor_12', 'VALR_12', 'knee_flexion_peak_torque', 'navicular_drop_asymmetry', 'total_ad_ab_ratio', 'hip_abduction_peak_torque', 'fat_intake_avg', 'past_month_distance', 'SC_past_season'

**TSGNN**

'rs12722', 'rs4986938', 'rs11225395', 'rs1144393', 'rs2252070', 'rs591058', 'rs4789932', 'class1_SNP_risk_score', 'rs13946', 'rs9340799', 'rs970547', 'rs1800012', 'tracking_period_injury', 'average_run_hours', 'average_interval_training_frequency', 'Age', 'EDEQ_total', 'navicular_drop', 'total_ad_ab_ratio', 'Q_angle_asymmetry', 'Q_angle', 'VALR_12', 'BMI', 'Impact_peak_12', 'hip_abduction_peak_torque', 'Duty_factor_12', 'knee_extension_peak_torque', 'navicular_drop_asymmetry', 'fat_intake_avg', 'past_month_distance', 'SC_past_season', 'non_running_past_season'

### Class 1-3

#### Decision Tree

rs591058, rs1800797, rs13946, rs25487, rs2228570, rs12722, rs2104772, rs1144393, rs3196378, rs1544410, rs2237352, rs1137101, rs10263021, rs4454832, rs7528684, rs2234693, rs10132091, rs7035322, rs1330363, rs1011814, rs1249269, rs4725069, rs820218, rs62051384, rs11232681, rs4701616, rs1800629, rs11177, rs143383, rs6617, rs6481512, rs17756404, rs4986938, rs13317, rs4730153, rs911263, rs970547, rs11225395, rs3018362, rs11629171, rs10992075, rs4789932, rs2252070, rs3789870, rs1590, rs42531, rs11154027, rs10759753, rs2761884, rs4362400, Q_angle, rs3219008, hip_abduction_peak_torque_asymmetry, rs3218791, total_ad_ab_ratio, rs17576, rs2289360, Age, rs1134170, rs2525504

#### Random Forest

rs591058, rs1800797, rs13946, rs25487, rs2228570, rs12722, rs2104772, rs1144393, rs3196378, rs1544410, rs2237352, rs1137101, rs10263021, rs4454832, rs7528684, rs2234693, rs10132091, rs7035322, rs1330363, rs1011814, rs1249269, rs4725069, rs820218, rs62051384, rs11232681, rs4701616, rs1800629, rs11177, rs143383, rs6617, rs6481512, rs17756404, rs4986938, rs13317, rs4730153, rs911263, rs970547, rs11225395, rs3018362, rs11629171, rs10992075, rs4789932, rs2252070, rs3789870, rs1590, rs42531, rs11154027, rs10759753, rs2761884, rs4362400, Q_angle, rs3219008, hip_abduction_peak_torque_asymmetry, rs3218791, total_ad_ab_ratio, rs17576, rs2289360, Age, rs1134170, rs2525504, fat_intake_BW, rs4919510, tracking_period_injury, rs1937810, Step_frequency_10, Athlete_Score, rs1718119, rs10484958, rs2285053, thigh_ffmi, class12_SNP_risk_score, fat_percentage_avg, hip_adduction_peak_torque_asymmetry, rs1800972, rs4328262, BMD_hip, calf_size, Flight_time_12, total_fl_ex_ratio, rs1800469, fat_intake_avg, navicular_drop, Step_frequency_12, knee_extension_peak_angle_asymmetry, glycine_intake_BW, rs2306033, rs9340799, rs17583842, rs4244032, rs1643821, rs1800470, rs3045, arginine_intake_BW, VALR_10, average_run_frequency, VALR_12, Q_angle_asymmetry, VILR_12, VILR_10, hip_adduction_peak_torque, rs4903399, thigh_lean_mass, rs3753841, calcium_intake_BW, total_lean_mass, leg_ffmi, rs2277698, BMD_spine, rs1800012, knee_flexion_peak_torque, total_ffmi, Flight_time_10, knee_extension_peak_torque, knee_flexion_peak_torque_asymmetry, average_run_hours, leg_lean_mass, average_interval_training_frequency, hip_abduction_peak_torque, rs12656106, average_energy_availability, VALR_asymmetry_12, knee_flexion_peak_angle_asymmetry, Impact_peak_12,

VILR_asymmetry_12, BMD_body, BMI, knee_flexion_peak_angle, hip_adduction_peak_angle, Duty_factor_10, knee_extension_peak_torque_asymmetry, rs187483, lower_leg_lean_mass, class1_SNP_risk_score, Duty_factor_asymmetry_12, Duty_factor_asymmetry_10, Impact_peak_10, Duty_factor_12, height, rs2281518, protein_intake_BW, navicular_drop_asymmetry, Cadence_asymmetry_10, past_month_injury, Mass, Impact_peak_asymmetry_10, sex, lower_leg_ffmi, EDEQ_total, class123_SNP_risk_score, rs1045485, rs1554606, knee_extension_peak_angle, fl_ex_ratio_asymmetry, rs78391032, past_stress_injury, Cadence_asymmetry_12, rs2305948, rs4654760, Contact_time_10, VALR_asymmetry_10, rs1887632, rs1800795, rs1021188, rs72758637, VILR_asymmetry_10, rs12154667, Contact_time_12, SC_past_season, rs1138545, rs3751143, rs1676303, rs7021589, SC_past_month, rs12574452, LEAF-Q, ad_ab_ratio_asymmetry, Impact_peak_asymmetry_12, vitaminD_intake_BW, rs13107325, rs1548456, rs2277268, resistance_training_past_season, past_month_volume_low, copper_intake_BW, past_month_distance, rs912336, hip_adduction_peak_angle_asymmetry, hip_abduction_peak_angle_asymmetry, hip_abduction_peak_angle, drills_past_season, non_running_past_season, past_month_min, rs35360670, rs420257, bodyweight_exercises_past_season, lower_limb_days_total, rs74544784, non_running_past_month, rs60713544, rs12429486, resistance_training_past_month, rs42517, rs2010963, rs71404070, core_stability_past_season, vitaminC_intake_BW, rs117544024, bodyweight_exercises_past_month

**SVM**

BMD_body, rs7035322, rs145648292, BMD_hip, omega3_intake_BW, Duty_factor_asymmetry_10, rs1676303, rs144414988, average_run_hours, ad_ab_ratio_asymmetry, hip_adduction_peak_torque, tracking_period_injury, Q_angle_asymmetry, past_month_ratio, rs1330363, rs9340799, knee_extension_peak_angle_asymmetry, rs2277268, copper_intake_BW, vitaminD_intake_BW, arginine_intake_BW, rs2858056, calcium_intake_BW, rs1011814, rs4903399, EDEQ_total, rs1590, rs4986938, Contact_time_12, rs3045, knee_flexion_peak_angle_asymmetry, rs4919510, rs1800469, rs1249269, Athlete_Score, rs11225395, rs10759753, rs2289360, SC_past_season, rs4701616, resistance_training_past_season, fat_percentage_avg

**KNN**

BMD_body, rs7035322, rs145648292, BMD_hip, omega3_intake_BW, Duty_factor_asymmetry_10, rs144414988, rs1676303, hip_adduction_peak_torque

**Naïve Bayes**

rs591058, rs1800797, rs13946, rs25487, rs2228570, rs12722, rs2104772, rs1144393, rs3196378, rs1544410, rs2237352, rs1137101, rs10263021, rs4454832, rs7528684, rs2234693, rs10132091, rs7035322, rs1330363, rs1011814, rs1249269, rs4725069, rs820218, rs62051384, rs11232681, rs4701616, rs1800629, rs11177, rs143383, rs6617, rs6481512, rs17756404, rs4986938, rs13317, rs4730153, rs911263, rs970547, rs11225395, rs3018362, rs11629171, rs10992075, rs4789932, rs2252070, rs3789870, rs1590, rs42531, rs11154027, rs10759753, rs2761884, rs4362400, Q_angle, rs3219008, hip_abduction_peak_torque_asymmetry, rs3218791, total_ad_ab_ratio, rs17576, rs2289360, Age, rs1134170, rs2525504, fat_intake_BW, rs4919510, tracking_period_injury, rs1937810, Step_frequency_10, Athlete_Score, rs1718119, rs10484958, rs2285053, thigh_ffmi, class12_SNP_risk_score, fat_percentage_avg, hip_adduction_peak_torque_asymmetry, rs1800972, rs4328262, BMD_hip, calf_size, Flight_time_12, total_fl_ex_ratio, rs1800469, fat_intake_avg, navicular_drop, Step_frequency_12, knee_extension_peak_angle_asymmetry, glycine_intake_BW, rs2306033, rs9340799, rs17583842, rs4244032, rs1643821, rs1800470, rs3045, arginine_intake_BW, VALR_10, average_run_frequency, VALR_12, Q_angle_asymmetry, VILR_12, VILR_10, hip_adduction_peak_torque, rs4903399, thigh_lean_mass, rs3753841, calcium_intake_BW, total_lean_mass, leg_ffmi, rs2277698, BMD_spine, rs1800012, knee_flexion_peak_torque, total_ffmi, Flight_time_10, knee_extension_peak_torque, knee_flexion_peak_torque_asymmetry, average_run_hours, leg_lean_mass, average_interval_training_frequency, hip_abduction_peak_torque, rs12656106, average_energy_availability, VALR_asymmetry_12, knee_flexion_peak_angle_asymmetry, Impact_peak_12, VILR_asymmetry_12, BMD_body, BMI, knee_flexion_peak_angle, hip_adduction_peak_angle, Duty_factor_10, knee_extension_peak_torque_asymmetry, rs187483, lower_leg_lean_mass, class1_SNP_risk_score, Duty_factor_asymmetry_12, Duty_factor_asymmetry_10, Impact_peak_10, Duty_factor_12, height, rs2281518, protein_intake_BW, navicular_drop_asymmetry, Cadence_asymmetry_10, past_month_injury, Mass, Impact_peak_asymmetry_10, sex, lower_leg_ffmi, EDEQ_total, class123_SNP_risk_score, rs1045485, rs1554606, knee_extension_peak_angle, fl_ex_ratio_asymmetry, rs78391032, past_stress_injury

**Adaboost**

rs591058, rs1800797, rs13946, rs25487, rs2228570, rs12722, rs2104772, rs1144393, rs3196378, rs1544410, rs2237352, rs1137101, rs10263021, rs4454832, rs7528684, rs2234693, rs10132091, rs7035322, rs1330363, rs1011814, rs1249269, rs4725069, rs820218, rs62051384, rs11232681, rs4701616, rs1800629, rs11177, rs143383, rs6617, rs6481512, rs17756404, rs4986938, rs13317, rs4730153, rs911263, rs970547, rs11225395, rs3018362, rs11629171, rs10992075, rs4789932, rs2252070, rs3789870, rs1590, rs42531, rs11154027, rs10759753, rs2761884, rs4362400, Q_angle, rs3219008, hip_abduction_peak_torque_asymmetry, rs3218791, total_ad_ab_ratio, rs17576, rs2289360, Age, rs1134170, rs2525504, fat_intake_BW, rs4919510, tracking_period_injury, rs1937810, Step_frequency_10, Athlete_Score, rs1718119, rs10484958, rs2285053, thigh_ffmi, class12_SNP_risk_score, fat_percentage_avg, hip_adduction_peak_torque_asymmetry, rs1800972, rs4328262, BMD_hip, calf_size, Flight_time_12, total_fl_ex_ratio, rs1800469, fat_intake_avg, navicular_drop, Step_frequency_12, knee_extension_peak_angle_asymmetry, glycine_intake_BW, rs2306033, rs9340799, rs17583842, rs4244032, rs1643821, rs1800470, rs3045, arginine_intake_BW, VALR_10, average_run_frequency, VALR_12,

Q_angle_asymmetry, VILR_12, VILR_10, hip_adduction_peak_torque, rs4903399, thigh_lean_mass, rs3753841, calcium_intake_BW, total_lean_mass, leg_ffmi, rs2277698, BMD_spine, rs1800012, knee_flexion_peak_torque, total_ffmi, Flight_time_10, knee_extension_peak_torque, knee_flexion_peak_torque_asymmetry, average_run_hours, leg_lean_mass, average_interval_training_frequency, hip_abduction_peak_torque, rs12656106, average_energy_availability, VALR_asymmetry_12, knee_flexion_peak_angle_asymmetry, Impact_peak_12, VILR_asymmetry_12, BMD_body, BMI, knee_flexion_peak_angle, hip_adduction_peak_angle, Duty_factor_10, knee_extension_peak_torque_asymmetry, rs187483, lower_leg_lean_mass, class1_SNP_risk_score, Duty_factor_asymmetry_12, Duty_factor_asymmetry_10, Impact_peak_10, Duty_factor_12, height, rs2281518, protein_intake_BW, navicular_drop_asymmetry, Cadence_asymmetry_10, past_month_injury, Mass, Impact_peak_asymmetry_10, sex, lower_leg_ffmi, EDEQ_total, class123_SNP_risk_score, rs1045485, rs1554606, knee_extension_peak_angle, fl_ex_ratio_asymmetry, rs78391032, past_stress_injury, Cadence_asymmetry_12, rs2305948, rs4654760, Contact_time_10

**Gradient Boosting**

BMD_body, rs7035322, rs145648292, BMD_hip, omega3_intake_BW, Duty_factor_asymmetry_10, rs144414988, rs1676303, hip_adduction_peak_torque, Q_angle_asymmetry, ad_ab_ratio_asymmetry, average_run_hours

**MLP**

rs591058, rs1800797, rs13946, rs25487, rs2228570, rs12722, rs2104772, rs1144393, rs3196378, rs1544410, rs2237352, rs1137101, rs10263021, rs4454832, rs7528684, rs2234693, rs10132091, rs7035322, rs1330363, rs1011814, rs1249269, rs4725069, rs820218, rs62051384, rs11232681, rs4701616, rs1800629, rs11177, rs143383, rs6617, rs6481512, rs17756404, rs4986938, rs13317, rs4730153, rs911263, rs970547, rs11225395, rs3018362, rs11629171, rs10992075, rs4789932, rs2252070, rs3789870, rs1590, rs42531, rs11154027, rs10759753, rs2761884, rs4362400, Q_angle, rs3219008, hip_abduction_peak_torque_asymmetry, rs3218791, total_ad_ab_ratio, rs17576, rs2289360, Age, rs1134170, rs2525504, fat_intake_BW, rs4919510, tracking_period_injury, rs1937810, Step_frequency_10, Athlete_Score, rs1718119, rs10484958, rs2285053, thigh_ffmi, class12_SNP_risk_score, fat_percentage_avg, hip_adduction_peak_torque_asymmetry, rs1800972, rs4328262, BMD_hip, calf_size, Flight_time_12, total_fl_ex_ratio, rs1800469, fat_intake_avg, navicular_drop, Step_frequency_12, knee_extension_peak_angle_asymmetry, glycine_intake_BW, rs2306033, rs9340799, rs17583842, rs4244032, rs1643821, rs1800470, rs3045, arginine_intake_BW, VALR_10, average_run_frequency, VALR_12, Q_angle_asymmetry, VILR_12, VILR_10, hip_adduction_peak_torque, rs4903399, thigh_lean_mass, rs3753841, calcium_intake_BW, total_lean_mass, leg_ffmi, rs2277698, BMD_spine, rs1800012, knee_flexion_peak_torque, total_ffmi, Flight_time_10, knee_extension_peak_torque, knee_flexion_peak_torque_asymmetry, average_run_hours, leg_lean_mass, average_interval_training_frequency, hip_abduction_peak_torque, rs12656106, average_energy_availability,

VALR_asymmetry_12, knee_flexion_peak_angle_asymmetry, Impact_peak_12, VILR_asymmetry_12, BMD_body, BMI, knee_flexion_peak_angle, hip_adduction_peak_angle, Duty_factor_10, knee_extension_peak_torque_asymmetry, rs187483, lower_leg_lean_mass, class1_SNP_risk_score, Duty_factor_asymmetry_12, Duty_factor_asymmetry_10, Impact_peak_10, Duty_factor_12, height, rs2281518, protein_intake_BW, navicular_drop_asymmetry, Cadence_asymmetry_10, past_month_injury, Mass

**Bayesian Network**

rs2252070, Impact_peak_12, navicular_drop, EDEQ_total, navicular_drop_asymmetry, rs13946, Q_angle, average_run_hours, class1_SNP_risk_score, rs9340799, lower_limb_days_total, Duty_factor_12, VALR_12, Q_angle_asymmetry, average_interval_training_frequency, total_ad_ab_ratio, hip_abduction_peak_torque, tracking_period_injury

**Logistic Regression**

rs145648292, average_run_hours, rs1676303, BMD_body, knee_extension_peak_angle_asymmetry, past_month_ratio, rs2277268, SC_past_season, rs3045, past_week_ratio_low, tracking_period_injury, rs7035322, Q_angle_asymmetry, Duty_factor_asymmetry_10, vitaminD_intake_BW, rs4988321, ad_ab_ratio_asymmetry, rs1144393, class1_SNP_risk_score, past_month_ratio_calculated_volume, rs4701616, rs13107325, calcium_intake_BW, rs1590, arginine_intake_BW, SC_past_month, rs11225395, hip_abduction_peak_angle, rs2104772, BMD_hip, resistance_training_past_season, rs2306033, hip_adduction_peak_torque, rs25489, rs4919510, rs10484958, fat_percentage_avg, leg_ffmi, rs912336, rs1800629, rs2289360, knee_flexion_peak_angle_asymmetry, rs2858056, average_energy_availability, rs10132091, rs2586488, resistance_training_past_month, Contact_time_12, rs1011814, past_month_distance, rs1249269, rs4903399, Alt_strike, rs1643821, rs1800972, rs1800469, rs2281518, rs11232681, rs62051384, rs3218791, rs1330363, rs2305948, Age, rs3219008, rs2010963, Athlete_Score, fl_ex_ratio_asymmetry, navicular_drop_asymmetry, past_month_injury, rs2252070, rs1800795, copper_intake_BW, protein_intake_BW, rs9340799, rs2234693, past_month_volume_high, rs25487, rs1045485, past_week_ratio_calculated_volume, rs2285053, rs6481512, rs10992075, rs143383, rs4328262, rs1887632, rs4986938, iron_intake_BW, drills_past_season, past_week_ratio_high, rs591058, rs17583842, circuit_training_past_month, rs12574452, past_month_ratio_moderate, EDEQ_total

**TSNN**

'rs591058', 'rs1800797', 'rs13946', 'rs25487', 'rs2228570', 'rs12722', 'rs2104772', 'rs1144393', 'rs3196378', 'rs1544410', 'rs2237352', 'rs1137101', 'rs10263021', 'rs4454832', 'rs7528684', 'rs2234693', 'rs10132091', 'rs7035322', 'rs1330363', 'rs1011814', 'rs1249269', 'rs4725069', 'rs820218', 'rs62051384', 'rs11232681', 'rs4701616', 'rs1800629', 'rs11177', 'rs143383', 'rs6617', 'rs6481512', 'rs17756404', 'rs4986938', 'rs13317', 'rs4730153', 'rs911263', 'rs970547', 'rs11225395', 'rs3018362', 'rs11629171', 'rs10992075', 'rs4789932', 'rs2252070', 'rs3789870', 'rs1590', 'rs42531', 'rs11154027', 'rs10759753', 'rs2761884', 'rs4362400', 'rs3219008', 'rs3218791', 'rs17576', 'rs2289360', 'rs1134170', 'rs2525504', 'rs4919510', 'rs1937810', 'rs1718119', 'rs10484958', 'rs2285053', 'class12_SNP_risk_score', 'rs1800972', 'rs4328262', 'rs1800469', 'rs2306033', 'rs9340799', 'rs17583842', 'Age', 'Q_angle', 'hip_abduction_peak_torque_asymmetry', 'total_ad_ab_ratio', 'Step_frequency_10', 'thigh_ffmi', 'hip_adduction_peak_torque_asymmetry', 'BMD_hip', 'calf_size', 'Flight_time_12', 'total_fl_ex_ratio', 'navicular_drop', 'Step_frequency_12', 'knee_extension_peak_angle_asymmetry', 'VALR_10', 'VALR_12', 'Q_angle_asymmetry', 'VILR_12', 'VILR_10', 'hip_adduction_peak_torque', 'thigh_lean_mass', 'total_lean_mass', 'leg_ffmi', 'BMD_spine', 'knee_flexion_peak_torque', 'total_ffmi', 'Flight_time_10', 'knee_extension_peak_torque', 'knee_flexion_peak_torque_asymmetry', 'leg_lean_mass', 'hip_abduction_peak_torque', 'VALR_asymmetry_12', 'knee_flexion_peak_angle_asymmetry', 'Impact_peak_12', 'VILR_asymmetry_12', 'BMD_body', 'BMI', 'knee_flexion_peak_angle', 'hip_adduction_peak_angle', 'Duty_factor_10', 'knee_extension_peak_torque_asymmetry', 'lower_leg_lean_mass', 'Duty_factor_asymmetry_12', 'Duty_factor_asymmetry_10', 'Impact_peak_10', 'Duty_factor_12', 'height', 'navicular_drop_asymmetry', 'Cadence_asymmetry_10', 'Mass', 'Impact_peak_asymmetry_10', 'lower_leg_ffmi', 'knee_extension_peak_angle', 'fl_ex_ratio_asymmetry', 'Cadence_asymmetry_12', 'Contact_time_10', 'VALR_asymmetry_10', 'VILR_asymmetry_10', 'Contact_time_12', 'ad_ab_ratio_asymmetry', 'fat_intake_BW', 'fat_percentage_avg', 'fat_intake_avg', 'glycine_intake_BW', 'arginine_intake_BW', 'calcium_intake_BW', 'average_energy_availability'

**TSGNN**

'rs591058', 'rs1800797', 'rs13946', 'rs25487', 'rs2228570', 'rs12722', 'rs2104772', 'rs1144393', 'rs3196378', 'rs1544410', 'rs2237352', 'rs1137101', 'rs10263021', 'rs4454832', 'rs7528684', 'rs2234693', 'rs10132091', 'rs7035322', 'rs1330363', 'rs1011814', 'rs1249269', 'rs4725069', 'rs820218', 'rs62051384', 'rs11232681', 'rs4701616', 'rs1800629', 'rs11177', 'rs143383', 'rs6617', 'rs6481512', 'rs17756404', 'rs4986938', 'rs13317', 'rs4730153', 'rs911263', 'rs970547', 'rs11225395', 'rs3018362', 'rs11629171', 'rs10992075', 'rs4789932', 'rs2252070', 'rs3789870', 'rs1590', 'rs42531', 'rs11154027', 'rs10759753', 'rs2761884', 'rs4362400', 'rs3219008', 'rs3218791', 'rs17576', 'rs2289360', 'rs1134170', 'rs2525504', 'rs4919510', 'rs1937810', 'rs1718119', 'rs10484958', 'rs2285053', 'class12_SNP_risk_score', 'rs1800972', 'rs4328262', 'rs1800469', 'rs2306033', 'rs9340799', 'rs17583842', 'rs4244032', 'rs1643821', 'rs1800470', 'rs3045', 'rs4903399', 'rs3753841', 'rs2277698', 'rs1800012', 'rs12656106', 'rs187483', 'class1_SNP_risk_score', 'rs2281518', 'sex', 'class123_SNP_risk_score', 'rs1045485', 'rs1554606', 'rs78391032', 'rs2305948', 'rs4654760', 'rs1887632', 'rs1800795', 'rs1021188', 'rs72758637', 'rs12154667', 'rs1138545', 'rs3751143', 'rs1676303', 'rs7021589', 'rs12574452', 'rs13107325', 'rs1548456', 'rs2277268', 'rs912336', 'rs35360670', 'rs420257', 'rs74544784', 'rs60713544', 'rs12429486', 'rs42517', 'rs2010963', 'rs71404070', 'rs117544024', 'rs2586488', 'rs25489', 'rs413826', 'rs650108', 'rs57104447', 'rs42522', 'rs710079', 'rs761804508', 'rs145648292', 'rs77569527', 'rs144371252', 'rs4988321', 'rs149047058', 'rs144414988', 'rs2858056', 'Age', 'tracking_period_injury', 'Athlete_Score', 'Q_angle', 'hip_abduction_peak_torque_asymmetry', 'total_ad_ab_ratio', 'Step_frequency_10', 'thigh_ffmi', 'hip_adduction_peak_torque_asymmetry', 'BMD_hip', 'calf_size', 'Flight_time_12', 'total_fl_ex_ratio', 'navicular_drop', 'Step_frequency_12', 'knee_extension_peak_angle_asymmetry', 'VALR_10', 'VALR_12', 'Q_angle_asymmetry', 'VILR_12', 'VILR_10', 'hip_adduction_peak_torque', 'thigh_lean_mass', 'total_lean_mass', 'leg_ffmi', 'BMD_spine', 'knee_flexion_peak_torque', 'total_ffmi', 'Flight_time_10', 'knee_extension_peak_torque', 'knee_flexion_peak_torque_asymmetry', 'leg_lean_mass', 'hip_abduction_peak_torque', 'VALR_asymmetry_12', 'knee_flexion_peak_angle_asymmetry', 'Impact_peak_12', 'VILR_asymmetry_12', 'BMD_body', 'BMI', 'knee_flexion_peak_angle', 'hip_adduction_peak_angle', 'Duty_factor_10', 'knee_extension_peak_torque_asymmetry', 'lower_leg_lean_mass', 'Duty_factor_asymmetry_12', 'Duty_factor_asymmetry_10', 'Impact_peak_10', 'fat_intake_BW', 'fat_percentage_avg', 'fat_intake_avg', 'glycine_intake_BW', 'arginine_intake_BW', 'calcium_intake_BW', 'average_energy_availability', 'protein_intake_BW', 'SC_past_season', 'SC_past_month', 'vitaminD_intake_BW', 'resistance_training_past_season', 'past_month_volume_low', 'copper_intake_BW', 'past_month_distance', 'drills_past_season', 'non_running_past_season', 'past_month_min', 'bodyweight_exercises_past_season', 'non_running_past_month'

### Model Interpretation

#### TSNN Interpretation

For each time-sequenced route, the effect of biases was peeled away into separate bias routes, and all that's left within the route becomes a multiplication sequence of (genotype feature * history feature * phenotype feature * behaviour feature * all weights * all attention weights * batch normalisation factors) along the route.

The peeled away bias terms were calculated individually as a separate value for the node it belongs. For instance, a bias term from a history node would generate routes of (bias * phenotype feature * behaviour feature * weights after the bias term was added * attention weights after the bias term was added * batch normalisation factors after the bias was added). When calculating total contributions, all routes from time-sequenced pathways and from bias pathways were added together, and the result matches the model's original prediction in logits.

When calculating an individual feature's contribution, all routes that pass through that feature, including time-sequenced routes and bias routes, were added together as the feature's total influence. When averaging across 20 runs, each feature's influence is divided by the sum of all absolute feature influence values during that run to perform a normalisation step. It is worth noting that the sum of all individual features' contributions does not equal to the model's original prediction, as each route is counted multiple times for different features.

### TSGNN Interpretation Output

**TSNN class 1**

**Consistent signs 17/20 (p=0.3244)**

Weight Matrix: W1_total

Number of elements with >= 17 consistent signs across all trials: 0/52 (0.00%)

Weight Matrix: W1_total

Number of elements with >= 17 consistent signs across all trials: 0/52 (0.00%)

Consistent elements in W1_total with >= 17 consistent signs:

Weight Matrix: W2_total

Number of elements with >= 17 consistent signs across all trials: 0/48 (0.00%)

Weight Matrix: W2_total

Number of elements with >= 17 consistent signs across all trials: 0/48 (0.00%)

Consistent elements in W2_total with >= 17 consistent signs:

Weight Matrix: W3_total

Number of elements with >= 17 consistent signs across all trials: 1/48 (2.08%)

Weight Matrix: W3_total

Number of elements with >= 17 consistent signs across all trials: 1/48 (2.08%)

Consistent elements in W3_total with >= 17 consistent signs:

Row 'fat_intake_avg' -> Column 'total_ad_ab_ratio': Sign Negative

Weight Matrix: W4

Number of elements with >= 17 consistent signs across all trials: 0/4 (0.00%)

Weight Matrix: W4

Number of elements with >= 17 consistent signs across all trials: 0/4 (0.00%)

Consistent elements in W4 with >= 17 consistent signs:

Routes with consistent signs on all of W1-W4 for >= 17/20 trials:

Total consistent routes: 0

**Consistent signs 16/20 (p=0.2681)**

Weight Matrix: W1_total

Number of elements with >= 16 consistent signs across all trials: 0/52 (0.00%)

Weight Matrix: W1_total

Number of elements with >= 16 consistent signs across all trials: 0/52 (0.00%)

Consistent elements in W1_total with >= 16 consistent signs:

Weight Matrix: W2_total

Number of elements with >= 16 consistent signs across all trials: 2/48 (4.17%)

Weight Matrix: W2_total

Number of elements with >= 16 consistent signs across all trials: 2/48 (4.17%)

Consistent elements in W2_total with >= 16 consistent signs:

Row 'knee_flexion_peak_torque' -> Column 'average_interval_training_frequency': Sign Negative

Row 'hip_abduction_peak_torque' -> Column 'History_extra': Sign Positive

Weight Matrix: W3_total

Number of elements with >= 16 consistent signs across all trials: 1/48 (2.08%)

Weight Matrix: W3_total

Number of elements with >= 16 consistent signs across all trials: 1/48 (2.08%)

Consistent elements in W3_total with >= 16 consistent signs:

Row 'fat_intake_avg' -> Column 'total_ad_ab_ratio': Sign Negative

Weight Matrix: W4

Number of elements with >= 16 consistent signs across all trials: 0/4 (0.00%)

Weight Matrix: W4

Number of elements with >= 16 consistent signs across all trials: 0/4 (0.00%)

Consistent elements in W4 with >= 16 consistent signs:

Routes with consistent signs on all of W1-W4 for >= 16/20 trials:

Total consistent routes: 0

**Consistent signs 15/20 (0.1009)**

Weight Matrix: W1_total

Number of elements with >= 15 consistent signs across all trials: 3/52 (5.77%)

Weight Matrix: W1_total

Number of elements with >= 15 consistent signs across all trials: 3/52 (5.77%)

Consistent elements in W1_total with >= 15 consistent signs:

Row 'Age' -> Column 'sex': Sign Positive

Row 'average_interval_training_frequency' -> Column 'rs1144393': Sign Positive

Row 'average_interval_training_frequency' -> Column 'rs970547': Sign Negative

Weight Matrix: W2_total

Number of elements with >= 15 consistent signs across all trials: 4/48 (8.33%)

Weight Matrix: W2_total

Number of elements with >= 15 consistent signs across all trials: 4/48 (8.33%)

Consistent elements in W2_total with >= 15 consistent signs:

Row 'Q_angle_asymmetry' -> Column 'Age': Sign Positive

Row 'knee_flexion_peak_torque' -> Column 'average_interval_training_frequency': Sign Negative

Row 'total_ad_ab_ratio' -> Column 'average_interval_training_frequency': Sign Positive

Row 'hip_abduction_peak_torque' -> Column 'History_extra': Sign Positive

Weight Matrix: W3_total

Number of elements with >= 15 consistent signs across all trials: 3/48 (6.25%)

Weight Matrix: W3_total

Number of elements with >= 15 consistent signs across all trials: 3/48 (6.25%)

Consistent elements in W3_total with >= 15 consistent signs:

Row 'fat_intake_avg' -> Column 'total_ad_ab_ratio': Sign Negative

Row 'past_month_distance' -> Column 'VALR_12': Sign Negative

Row 'Behaviour_extra' -> Column 'hip_abduction_peak_torque': Sign Positive

Weight Matrix: W4

Number of elements with >= 15 consistent signs across all trials: 0/4 (0.00%)

Weight Matrix: W4

Number of elements with >= 15 consistent signs across all trials: 0/4 (0.00%)

Consistent elements in W4 with >= 15 consistent signs:

Routes with consistent signs on all of W1-W4 for >= 15/20 trials:

Total consistent routes: 0

**Consistent signs 14/20 (0.0692)**

Weight Matrix: W1_total

Number of elements with >= 14 consistent signs across all trials: 4/52 (7.69%)

Weight Matrix: W1_total

Number of elements with >= 14 consistent signs across all trials: 4/52 (7.69%)

Consistent elements in W1_total with >= 14 consistent signs:

Row 'average_run_hours' -> Column 'rs970547': Sign Positive

Row 'Age' -> Column 'sex': Sign Positive

Row 'average_interval_training_frequency' -> Column 'rs1144393': Sign Positive

Row 'average_interval_training_frequency' -> Column 'rs970547': Sign Negative

Weight Matrix: W2_total

Number of elements with >= 14 consistent signs across all trials: 7/48 (14.58%)

Weight Matrix: W2_total

Number of elements with >= 14 consistent signs across all trials: 7/48 (14.58%)

Consistent elements in W2_total with >= 14 consistent signs:

Row 'Q_angle_asymmetry' -> Column 'Age': Sign Positive

Row 'Q_angle_asymmetry' -> Column 'average_interval_training_frequency': Sign Positive

Row 'knee_flexion_peak_torque' -> Column 'Age': Sign Negative

Row 'knee_flexion_peak_torque' -> Column 'average_interval_training_frequency': Sign Negative

Row 'navicular_drop_asymmetry' -> Column 'average_run_hours': Sign Positive

Row 'total_ad_ab_ratio' -> Column 'average_interval_training_frequency': Sign Positive

Row 'hip_abduction_peak_torque' -> Column 'History_extra': Sign Positive

Weight Matrix: W3_total

Number of elements with >= 14 consistent signs across all trials: 10/48 (20.83%)

Weight Matrix: W3_total

Number of elements with >= 14 consistent signs across all trials: 10/48 (20.83%)

Consistent elements in W3_total with >= 14 consistent signs:

Row 'fat_intake_avg' -> Column 'BMD_spine': Sign Negative

Row 'fat_intake_avg' -> Column 'total_ad_ab_ratio': Sign Negative

Row 'fat_intake_avg' -> Column 'Phenotype_extra': Sign Positive

Row 'past_month_distance' -> Column 'Q_angle_asymmetry': Sign Negative

Row 'past_month_distance' -> Column 'Impact_peak_12': Sign Negative

Row 'past_month_distance' -> Column 'VALR_12': Sign Negative

Row 'SC_past_season' -> Column 'Impact_peak_12': Sign Negative

Row 'Behaviour_extra' -> Column 'BMD_spine': Sign Positive

Row 'Behaviour_extra' -> Column 'VALR_12': Sign Positive

Row 'Behaviour_extra' -> Column 'hip_abduction_peak_torque': Sign Positive

Weight Matrix: W4

Number of elements with >= 14 consistent signs across all trials: 3/4 (75.00%)

Weight Matrix: W4

Number of elements with >= 14 consistent signs across all trials: 3/4 (75.00%)

Consistent elements in W4 with >= 14 consistent signs:

Row 'Output' -> Column 'past_month_distance': Sign Negative

Row 'Output' -> Column 'SC_past_season': Sign Negative

Row 'Output' -> Column 'Behaviour_extra': Sign Negative

Routes with consistent signs on all of W1-W4 for >= 14/20 trials:

Genotype: rs1144393 (+) -> History: average_interval_training_frequency (+) -> Phenotype: Q_angle_asymmetry (-) -> Behaviour: past_month_distance (-)

Genotype: sex (+) -> History: Age (+) -> Phenotype: Q_angle_asymmetry (-) -> Behaviour: past_month_distance (-)

Genotype: rs970547 (-) -> History: average_interval_training_frequency (+) -> Phenotype: Q_angle_asymmetry (-) -> Behaviour: past_month_distance (-)

Total consistent routes: 3

**SHAP compared to node contribution**

Enhanced Summary Statistics:

feature_set metric_type mean std min 25% median 75% max

All Features Contribution Correlation 0.172488 0.270565 -0.378825 0.008470 0.158817 0.395710 0.824268

All Features Ratio Correlation 0.103420 0.216951 -0.364808 -0.041671 0.091229 0.279336 0.637462

All Features Rank Correlation 0.124371 0.193438 -0.359644 -0.008516 0.112020 0.276289 0.496550

Top 5 Node Pos/Neg Contribution Correlation 0.206436 0.399965 -0.806639 -0.040488 0.178851 0.558161 0.919400

Top 5 Node Pos/Neg Ratio Correlation 0.216826 0.362704 -0.516250 -0.066835 0.227511 0.443749 0.888621

Top 5 Node Pos/Neg Rank Correlation 0.219320 0.359825 -0.595238 -0.026190 0.264286 0.482143 0.857143

Top 5 SHAP Pos/Neg Contribution Correlation 0.245100 0.380140 -0.524982 -0.039711 0.259875 0.538351 0.875567

Top 5 SHAP Pos/Neg Ratio Correlation 0.189911 0.318642 -0.484355 -0.037481 0.174875 0.429559 0.845410

Top 5 SHAP Pos/Neg Rank Correlation 0.246627 0.345107 -0.581534 -0.018759 0.237823 0.527674 0.782624

Combined 3 Node + 3 SHAP Contribution Correlation 0.266041 0.403678 -0.548258 -0.092766 0.324018 0.615985 0.921461

Combined 3 Node + 3 SHAP Ratio Correlation 0.242286 0.317293 -0.483461 0.025070 0.204124 0.467659 0.895888

Combined 3 Node + 3 SHAP Rank Correlation 0.252908 0.352153 -0.516419 -0.006061 0.254846 0.566667 0.833333

Top Node Ratios Contribution Correlation 0.256447 0.398349 -0.656974 -0.000695 0.291807 0.608927 0.944449

Top Node Ratios Ratio Correlation 0.161245 0.376793 -0.683323 -0.115320 0.170469 0.435529 0.877657

Top Node Ratios Rank Correlation 0.218357 0.384153 -0.745455 -0.060606 0.193939 0.518182 0.951515

Top SHAP Ratios Contribution Correlation 0.251656 0.361520 -0.482867 -0.029335 0.257112 0.539377 0.894494

Top SHAP Ratios Ratio Correlation 0.172361 0.317310 -0.430887 -0.070722 0.188097 0.443410 0.847735

Top SHAP Ratios Rank Correlation 0.248054 0.327485 -0.454545 0.004545 0.248485 0.493939 0.842424

Combined Ratios Contribution Correlation 0.205232 0.385780 -0.860023 -0.075428 0.243937 0.536525 0.867887

Combined Ratios Ratio Correlation 0.149532 0.310112 -0.597681 -0.063439 0.146359 0.431764 0.756764

Combined Ratios Rank Correlation 0.169025 0.345928 -0.716667 -0.066667 0.227972 0.411364 0.833333

**TSNN class 123**

**Consistent signs 18/20 (p=0.2588)**

Weight Matrix: W1_total

Number of elements with >= 18 consistent signs across all trials: 0/136 (0.00%)

Weight Matrix: W1_total

Number of elements with >= 18 consistent signs across all trials: 0/136 (0.00%)

Consistent elements in W1_total with >= 18 consistent signs:

Weight Matrix: W2_total

Number of elements with >= 18 consistent signs across all trials: 1/120 (0.83%)

Weight Matrix: W2_total

Number of elements with >= 18 consistent signs across all trials: 1/120 (0.83%)

Consistent elements in W2_total with >= 18 consistent signs:

Row 'Duty_factor_10' -> Column 'History_extra': Sign Positive

Weight Matrix: W3_total

Number of elements with >= 18 consistent signs across all trials: 0/480 (0.00%)

Weight Matrix: W3_total

Number of elements with >= 18 consistent signs across all trials: 0/480 (0.00%)

Consistent elements in W3_total with >= 18 consistent signs:

Weight Matrix: W4

Number of elements with >= 18 consistent signs across all trials: 0/8 (0.00%)

Weight Matrix: W4

Number of elements with >= 18 consistent signs across all trials: 0/8 (0.00%)

Consistent elements in W4 with >= 18 consistent signs:

Routes with consistent signs on all of W1-W4 for >= 18/20 trials:

Total consistent routes: 0

**Consistent signs 17/20 (p=0.0036)**

Weight Matrix: W1_total

Number of elements with >= 17 consistent signs across all trials: 3/136 (2.21%)

Weight Matrix: W1_total

Number of elements with >= 17 consistent signs across all trials: 3/136 (2.21%)

Consistent elements in W1_total with >= 17 consistent signs:

Row 'History_extra' -> Column 'rs1330363': Sign Positive

Row 'History_extra' -> Column 'rs3018362': Sign Positive

Row 'History_extra' -> Column 'rs1718119': Sign Positive

Weight Matrix: W2_total

Number of elements with >= 17 consistent signs across all trials: 1/120 (0.83%)

Weight Matrix: W2_total

Number of elements with >= 17 consistent signs across all trials: 1/120 (0.83%)

Consistent elements in W2_total with >= 17 consistent signs:

Row 'Duty_factor_10' -> Column 'History_extra': Sign Positive

Weight Matrix: W3_total

Number of elements with >= 17 consistent signs across all trials: 3/480 (0.62%)

Weight Matrix: W3_total

Number of elements with >= 17 consistent signs across all trials: 3/480 (0.62%)

Consistent elements in W3_total with >= 17 consistent signs:

Row 'fat_intake_BW' -> Column 'fl_ex_ratio_asymmetry': Sign Negative

Row 'calcium_intake_BW' -> Column 'Duty_factor_asymmetry_10': Sign Positive

Row 'average_energy_availability' -> Column 'VILR_asymmetry_12': Sign Positive

Weight Matrix: W4

Number of elements with >= 17 consistent signs across all trials: 0/8 (0.00%)

Weight Matrix: W4

Number of elements with >= 17 consistent signs across all trials: 0/8 (0.00%)

Consistent elements in W4 with >= 17 consistent signs:

Routes with consistent signs on all of W1-W4 for >= 17/20 trials:

Total consistent routes: 0

**Consistent signs 16/20 (p=0.0007)**

Weight Matrix: W1_total

Number of elements with >= 16 consistent signs across all trials: 12/136 (8.82%)

Weight Matrix: W1_total

Number of elements with >= 16 consistent signs across all trials: 12/136 (8.82%)

Consistent elements in W1_total with >= 16 consistent signs:

Row 'Age' -> Column 'rs13946': Sign Negative

Row 'Age' -> Column 'rs970547': Sign Negative

Row 'Age' -> Column 'rs2285053': Sign Positive

Row 'History_extra' -> Column 'rs1544410': Sign Positive

Row 'History_extra' -> Column 'rs1330363': Sign Positive

Row 'History_extra' -> Column 'rs4725069': Sign Positive

Row 'History_extra' -> Column 'rs62051384': Sign Positive

Row 'History_extra' -> Column 'rs13317': Sign Positive

Row 'History_extra' -> Column 'rs970547': Sign Positive

Row 'History_extra' -> Column 'rs3018362': Sign Positive

Row 'History_extra' -> Column 'rs1718119': Sign Positive

Row 'History_extra' -> Column 'rs1800469': Sign Positive

Weight Matrix: W2_total

Number of elements with >= 16 consistent signs across all trials: 2/120 (1.67%)

Weight Matrix: W2_total

Number of elements with >= 16 consistent signs across all trials: 2/120 (1.67%)

Consistent elements in W2_total with >= 16 consistent signs:

Row 'BMD_hip' -> Column 'Age': Sign Positive

Row 'Duty_factor_10' -> Column 'History_extra': Sign Positive

Weight Matrix: W3_total

Number of elements with >= 16 consistent signs across all trials: 6/480 (1.25%)

Weight Matrix: W3_total

Number of elements with >= 16 consistent signs across all trials: 6/480 (1.25%)

Consistent elements in W3_total with >= 16 consistent signs:

Row 'fat_intake_BW' -> Column 'fl_ex_ratio_asymmetry': Sign Negative

Row 'fat_percentage_avg' -> Column 'Q_angle': Sign Positive

Row 'fat_percentage_avg' -> Column 'Flight_time_12': Sign Positive

Row 'calcium_intake_BW' -> Column 'VALR_10': Sign Positive

Row 'calcium_intake_BW' -> Column 'Duty_factor_asymmetry_10': Sign Positive

Row 'average_energy_availability' -> Column 'VILR_asymmetry_12': Sign Positive

Weight Matrix: W4

Number of elements with >= 16 consistent signs across all trials: 0/8 (0.00%)

Weight Matrix: W4

Number of elements with >= 16 consistent signs across all trials: 0/8 (0.00%)

Consistent elements in W4 with >= 16 consistent signs:

Routes with consistent signs on all of W1-W4 for >= 16/20 trials:

Total consistent routes: 0

**Consistent signs 15/20 (p<0.0001)**

Weight Matrix: W1_total

Number of elements with >= 15 consistent signs across all trials: 24/136 (17.65%)

Weight Matrix: W1_total

Number of elements with >= 15 consistent signs across all trials: 24/136 (17.65%)

Consistent elements in W1_total with >= 15 consistent signs:

Row 'Age' -> Column 'rs13946': Sign Negative

Row 'Age' -> Column 'rs970547': Sign Negative

Row 'Age' -> Column 'rs2285053': Sign Positive

Row 'History_extra' -> Column 'rs13946': Sign Positive

Row 'History_extra' -> Column 'rs1544410': Sign Positive

Row 'History_extra' -> Column 'rs1330363': Sign Positive

Row 'History_extra' -> Column 'rs4725069': Sign Positive

Row 'History_extra' -> Column 'rs62051384': Sign Positive

Row 'History_extra' -> Column 'rs4701616': Sign Positive

Row 'History_extra' -> Column 'rs17756404': Sign Positive

Row 'History_extra' -> Column 'rs13317': Sign Positive

Row 'History_extra' -> Column 'rs911263': Sign Positive

Row 'History_extra' -> Column 'rs970547': Sign Positive

Row 'History_extra' -> Column 'rs11225395': Sign Positive

Row 'History_extra' -> Column 'rs3018362': Sign Positive

Row 'History_extra' -> Column 'rs2252070': Sign Positive

Row 'History_extra' -> Column 'rs2525504': Sign Positive

Row 'History_extra' -> Column 'rs4919510': Sign Positive

Row 'History_extra' -> Column 'rs1718119': Sign Positive

Row 'History_extra' -> Column 'rs10484958': Sign Negative

Row 'History_extra' -> Column 'rs4328262': Sign Positive

Row 'History_extra' -> Column 'rs1800469': Sign Positive

Row 'History_extra' -> Column 'rs2306033': Sign Positive

Row 'History_extra' -> Column 'rs9340799': Sign Positive

Weight Matrix: W2_total

Number of elements with >= 15 consistent signs across all trials: 10/120 (8.33%)

Weight Matrix: W2_total

Number of elements with >= 15 consistent signs across all trials: 10/120 (8.33%)

Consistent elements in W2_total with >= 15 consistent signs:

Row 'BMD_hip' -> Column 'Age': Sign Positive

Row 'leg_ffmi' -> Column 'Age': Sign Positive

Row 'Impact_peak_12' -> Column 'Age': Sign Positive

Row 'VILR_asymmetry_12' -> Column 'Age': Sign Positive

Row 'knee_flexion_peak_angle' -> Column 'History_extra': Sign Positive

Row 'Duty_factor_10' -> Column 'History_extra': Sign Positive

Row 'lower_leg_lean_mass' -> Column 'Age': Sign Positive

Row 'Duty_factor_asymmetry_12' -> Column 'History_extra': Sign Negative

Row 'height' -> Column 'History_extra': Sign Positive

Row 'Contact_time_12' -> Column 'Age': Sign Positive

Weight Matrix: W3_total

Number of elements with >= 15 consistent signs across all trials: 22/480 (4.58%)

Weight Matrix: W3_total

Number of elements with >= 15 consistent signs across all trials: 22/480 (4.58%)

Consistent elements in W3_total with >= 15 consistent signs:

Row 'fat_intake_BW' -> Column 'navicular_drop': Sign Negative

Row 'fat_intake_BW' -> Column 'Flight_time_10': Sign Negative

Row 'fat_intake_BW' -> Column 'knee_extension_peak_torque': Sign Positive

Row 'fat_intake_BW' -> Column 'fl_ex_ratio_asymmetry': Sign Negative

Row 'fat_percentage_avg' -> Column 'Q_angle': Sign Positive

Row 'fat_percentage_avg' -> Column 'Flight_time_12': Sign Positive

Row 'fat_percentage_avg' -> Column 'knee_flexion_peak_torque_asymmetry': Sign Positive

Row 'fat_percentage_avg' -> Column 'fl_ex_ratio_asymmetry': Sign Positive

Row 'fat_intake_avg' -> Column 'Step_frequency_10': Sign Negative

Row 'fat_intake_avg' -> Column 'lower_leg_ffmi': Sign Positive

Row 'fat_intake_avg' -> Column 'Phenotype_extra': Sign Positive

Row 'glycine_intake_BW' -> Column 'BMD_hip': Sign Positive

Row 'glycine_intake_BW' -> Column 'total_lean_mass': Sign Positive

Row 'glycine_intake_BW' -> Column 'knee_flexion_peak_angle': Sign Positive

Row 'calcium_intake_BW' -> Column 'VALR_10': Sign Positive

Row 'calcium_intake_BW' -> Column 'leg_lean_mass': Sign Negative

Row 'calcium_intake_BW' -> Column 'Duty_factor_asymmetry_10': Sign Positive

Row 'calcium_intake_BW' -> Column 'Impact_peak_10': Sign Positive

Row 'calcium_intake_BW' -> Column 'navicular_drop_asymmetry': Sign Positive

Row 'average_energy_availability' -> Column 'leg_ffmi': Sign Positive

Row 'average_energy_availability' -> Column 'VILR_asymmetry_12': Sign Positive

Row 'Behaviour_extra' -> Column 'Step_frequency_10': Sign Negative

Weight Matrix: W4

Number of elements with >= 15 consistent signs across all trials: 0/8 (0.00%)

Weight Matrix: W4

Number of elements with >= 15 consistent signs across all trials: 0/8 (0.00%)

Consistent elements in W4 with >= 15 consistent signs:

Routes with consistent signs on all of W1-W4 for >= 15/20 trials:

Total consistent routes: 0

**Consistent signs 14/20 (p=0.0005)**

Weight Matrix: W1_total

Number of elements with >= 14 consistent signs across all trials: 40/136 (29.41%)

Weight Matrix: W1_total

Number of elements with >= 14 consistent signs across all trials: 40/136 (29.41%)

Consistent elements in W1_total with >= 14 consistent signs:

Row 'Age' -> Column 'rs13946': Sign Negative

Row 'Age' -> Column 'rs10263021': Sign Negative

Row 'Age' -> Column 'rs4454832': Sign Negative

Row 'Age' -> Column 'rs1011814': Sign Negative

Row 'Age' -> Column 'rs970547': Sign Negative

Row 'Age' -> Column 'rs1590': Sign Negative

Row 'Age' -> Column 'rs4362400': Sign Negative

Row 'Age' -> Column 'rs2285053': Sign Positive

Row 'Age' -> Column 'rs9340799': Sign Negative

Row 'Age' -> Column 'rs17583842': Sign Negative

Row 'History_extra' -> Column 'rs591058': Sign Positive

Row 'History_extra' -> Column 'rs13946': Sign Positive

Row 'History_extra' -> Column 'rs1544410': Sign Positive

Row 'History_extra' -> Column 'rs10263021': Sign Positive

Row 'History_extra' -> Column 'rs4454832': Sign Negative

Row 'History_extra' -> Column 'rs7528684': Sign Positive

Row 'History_extra' -> Column 'rs1330363': Sign Positive

Row 'History_extra' -> Column 'rs4725069': Sign Positive

Row 'History_extra' -> Column 'rs820218': Sign Positive

Row 'History_extra' -> Column 'rs62051384': Sign Positive

Row 'History_extra' -> Column 'rs4701616': Sign Positive

Row 'History_extra' -> Column 'rs17756404': Sign Positive

Row 'History_extra' -> Column 'rs4986938': Sign Positive

Row 'History_extra' -> Column 'rs13317': Sign Positive

Row 'History_extra' -> Column 'rs911263': Sign Positive

Row 'History_extra' -> Column 'rs970547': Sign Positive

Row 'History_extra' -> Column 'rs11225395': Sign Positive

Row 'History_extra' -> Column 'rs3018362': Sign Positive

Row 'History_extra' -> Column 'rs2252070': Sign Positive

Row 'History_extra' -> Column 'rs11154027': Sign Positive

Row 'History_extra' -> Column 'rs3219008': Sign Positive

Row 'History_extra' -> Column 'rs2289360': Sign Positive

Row 'History_extra' -> Column 'rs2525504': Sign Positive

Row 'History_extra' -> Column 'rs4919510': Sign Positive

Row 'History_extra' -> Column 'rs1718119': Sign Positive

Row 'History_extra' -> Column 'rs10484958': Sign Negative

Row 'History_extra' -> Column 'rs4328262': Sign Positive

Row 'History_extra' -> Column 'rs1800469': Sign Positive

Row 'History_extra' -> Column 'rs2306033': Sign Positive

Row 'History_extra' -> Column 'rs9340799': Sign Positive

Weight Matrix: W2_total

Number of elements with >= 14 consistent signs across all trials: 17/120 (14.17%)

Weight Matrix: W2_total

Number of elements with >= 14 consistent signs across all trials: 17/120 (14.17%)

Consistent elements in W2_total with >= 14 consistent signs:

Row 'Step_frequency_10' -> Column 'History_extra': Sign Negative

Row 'hip_adduction_peak_torque_asymmetry' -> Column 'Age': Sign Positive

Row 'BMD_hip' -> Column 'Age': Sign Positive

Row 'calf_size' -> Column 'Age': Sign Negative

Row 'knee_extension_peak_angle_asymmetry' -> Column 'Age': Sign Negative

Row 'VALR_12' -> Column 'Age': Sign Negative

Row 'leg_ffmi' -> Column 'Age': Sign Positive

Row 'Impact_peak_12' -> Column 'Age': Sign Positive

Row 'VILR_asymmetry_12' -> Column 'Age': Sign Positive

Row 'BMI' -> Column 'Age': Sign Negative

Row 'knee_flexion_peak_angle' -> Column 'History_extra': Sign Positive

Row 'Duty_factor_10' -> Column 'History_extra': Sign Positive

Row 'lower_leg_lean_mass' -> Column 'Age': Sign Positive

Row 'Duty_factor_asymmetry_12' -> Column 'History_extra': Sign Negative

Row 'Duty_factor_12' -> Column 'History_extra': Sign Positive

Row 'height' -> Column 'History_extra': Sign Positive

Row 'Contact_time_12' -> Column 'Age': Sign Positive

Weight Matrix: W3_total

Number of elements with >= 14 consistent signs across all trials: 58/480 (12.08%)

Weight Matrix: W3_total

Number of elements with >= 14 consistent signs across all trials: 58/480 (12.08%)

Consistent elements in W3_total with >= 14 consistent signs:

Row 'fat_intake_BW' -> Column 'hip_adduction_peak_torque_asymmetry': Sign Positive

Row 'fat_intake_BW' -> Column 'Flight_time_12': Sign Positive

Row 'fat_intake_BW' -> Column 'navicular_drop': Sign Negative

Row 'fat_intake_BW' -> Column 'Flight_time_10': Sign Negative

Row 'fat_intake_BW' -> Column 'knee_extension_peak_torque': Sign Positive

Row 'fat_intake_BW' -> Column 'knee_extension_peak_torque_asymmetry': Sign Negative

Row 'fat_intake_BW' -> Column 'Duty_factor_12': Sign Positive

Row 'fat_intake_BW' -> Column 'fl_ex_ratio_asymmetry': Sign Negative

Row 'fat_percentage_avg' -> Column 'Q_angle': Sign Positive

Row 'fat_percentage_avg' -> Column 'hip_adduction_peak_torque_asymmetry': Sign Positive

Row 'fat_percentage_avg' -> Column 'Flight_time_12': Sign Positive

Row 'fat_percentage_avg' -> Column 'knee_flexion_peak_torque_asymmetry': Sign Positive

Row 'fat_percentage_avg' -> Column 'lower_leg_lean_mass': Sign Negative

Row 'fat_percentage_avg' -> Column 'Duty_factor_asymmetry_12': Sign Positive

Row 'fat_percentage_avg' -> Column 'fl_ex_ratio_asymmetry': Sign Positive

Row 'fat_intake_avg' -> Column 'Step_frequency_10': Sign Negative

Row 'fat_intake_avg' -> Column 'VILR_10': Sign Positive

Row 'fat_intake_avg' -> Column 'Duty_factor_asymmetry_12': Sign Positive

Row 'fat_intake_avg' -> Column 'lower_leg_ffmi': Sign Positive

Row 'fat_intake_avg' -> Column 'fl_ex_ratio_asymmetry': Sign Positive

Row 'fat_intake_avg' -> Column 'Cadence_asymmetry_12': Sign Positive

Row 'fat_intake_avg' -> Column 'Phenotype_extra': Sign Positive

Row 'glycine_intake_BW' -> Column 'BMD_hip': Sign Positive

Row 'glycine_intake_BW' -> Column 'Q_angle_asymmetry': Sign Negative

Row 'glycine_intake_BW' -> Column 'total_lean_mass': Sign Positive

Row 'glycine_intake_BW' -> Column 'knee_flexion_peak_torque': Sign Positive

Row 'glycine_intake_BW' -> Column 'VALR_asymmetry_12': Sign Negative

Row 'glycine_intake_BW' -> Column 'BMI': Sign Positive

Row 'glycine_intake_BW' -> Column 'knee_flexion_peak_angle': Sign Positive

Row 'glycine_intake_BW' -> Column 'lower_leg_ffmi': Sign Negative

Row 'arginine_intake_BW' -> Column 'BMD_hip': Sign Positive

Row 'arginine_intake_BW' -> Column 'calf_size': Sign Negative

Row 'arginine_intake_BW' -> Column 'Duty_factor_12': Sign Positive

Row 'arginine_intake_BW' -> Column 'Contact_time_12': Sign Negative

Row 'calcium_intake_BW' -> Column 'Step_frequency_10': Sign Positive

Row 'calcium_intake_BW' -> Column 'Flight_time_12': Sign Positive

Row 'calcium_intake_BW' -> Column 'VALR_10': Sign Positive

Row 'calcium_intake_BW' -> Column 'VILR_12': Sign Positive

Row 'calcium_intake_BW' -> Column 'total_ffmi': Sign Negative

Row 'calcium_intake_BW' -> Column 'leg_lean_mass': Sign Negative

Row 'calcium_intake_BW' -> Column 'Duty_factor_asymmetry_10': Sign Positive

Row 'calcium_intake_BW' -> Column 'Impact_peak_10': Sign Positive

Row 'calcium_intake_BW' -> Column 'Duty_factor_12': Sign Positive

Row 'calcium_intake_BW' -> Column 'navicular_drop_asymmetry': Sign Positive

Row 'calcium_intake_BW' -> Column 'ad_ab_ratio_asymmetry': Sign Positive

Row 'average_energy_availability' -> Column 'Step_frequency_10': Sign Negative

Row 'average_energy_availability' -> Column 'VILR_10': Sign Positive

Row 'average_energy_availability' -> Column 'leg_ffmi': Sign Positive

Row 'average_energy_availability' -> Column 'VALR_asymmetry_12': Sign Positive

Row 'average_energy_availability' -> Column 'VILR_asymmetry_12': Sign Positive

Row 'average_energy_availability' -> Column 'BMD_body': Sign Negative

Row 'average_energy_availability' -> Column 'Contact_time_12': Sign Positive

Row 'Behaviour_extra' -> Column 'Q_angle': Sign Positive

Row 'Behaviour_extra' -> Column 'Step_frequency_10': Sign Negative

Row 'Behaviour_extra' -> Column 'BMD_hip': Sign Positive

Row 'Behaviour_extra' -> Column 'Q_angle_asymmetry': Sign Negative

Row 'Behaviour_extra' -> Column 'thigh_lean_mass': Sign Positive

Row 'Behaviour_extra' -> Column 'Duty_factor_asymmetry_12': Sign Positive

Weight Matrix: W4

Number of elements with >= 14 consistent signs across all trials: 1/8 (12.50%)

Weight Matrix: W4

Number of elements with >= 14 consistent signs across all trials: 1/8 (12.50%)

Consistent elements in W4 with >= 14 consistent signs:

Row 'Output' -> Column 'average_energy_availability': Sign Negative

Routes with consistent signs on all of W1-W4 for >= 14/20 trials:

Genotype: rs9340799 (-) -> History: Age (+) -> Phenotype: VILR_asymmetry_12 (+) -> Behaviour: average_energy_availability (-)

Genotype: rs9340799 (-) -> History: Age (+) -> Phenotype: Contact_time_12 (+) -> Behaviour: average_energy_availability (-)

Genotype: rs820218 (+) -> History: History_extra (-) -> Phenotype: Step_frequency_10 (-) -> Behaviour: average_energy_availability (-)

Genotype: rs1330363 (+) -> History: History_extra (-) -> Phenotype: Step_frequency_10 (-) -> Behaviour: average_energy_availability (-)

Genotype: rs17756404 (+) -> History: History_extra (-) -> Phenotype: Step_frequency_10 (-) -> Behaviour: average_energy_availability (-)

Genotype: rs1590 (-) -> History: Age (+) -> Phenotype: VILR_asymmetry_12 (+) -> Behaviour: average_energy_availability (-)

Genotype: rs4454832 (-) -> History: Age (+) -> Phenotype: leg_ffmi (+) -> Behaviour: average_energy_availability (-)

Genotype: rs1590 (-) -> History: Age (+) -> Phenotype: Contact_time_12 (+) -> Behaviour: average_energy_availability (-)

Genotype: rs4919510 (+) -> History: History_extra (-) -> Phenotype: Step_frequency_10 (-) -> Behaviour: average_energy_availability (-)

Genotype: rs7528684 (+) -> History: History_extra (-) -> Phenotype: Step_frequency_10 (-) -> Behaviour: average_energy_availability (-)

Genotype: rs2285053 (+) -> History: Age (+) -> Phenotype: leg_ffmi (+) -> Behaviour: average_energy_availability (-)

Genotype: rs591058 (+) -> History: History_extra (-) -> Phenotype: Step_frequency_10 (-) -> Behaviour: average_energy_availability (-)

Genotype: rs11225395 (+) -> History: History_extra (-) -> Phenotype: Step_frequency_10 (-) -> Behaviour: average_energy_availability (-)

Genotype: rs13317 (+) -> History: History_extra (-) -> Phenotype: Step_frequency_10 (-) -> Behaviour: average_energy_availability (-)

Genotype: rs4362400 (-) -> History: Age (+) -> Phenotype: leg_ffmi (+) -> Behaviour: average_energy_availability (-)

Genotype: rs4986938 (+) -> History: History_extra (-) -> Phenotype: Step_frequency_10 (-) -> Behaviour: average_energy_availability (-)

Genotype: rs2306033 (+) -> History: History_extra (-) -> Phenotype: Step_frequency_10 (-) -> Behaviour: average_energy_availability (-)

Genotype: rs1011814 (-) -> History: Age (+) -> Phenotype: VILR_asymmetry_12 (+) -> Behaviour: average_energy_availability (-)

Genotype: rs1011814 (-) -> History: Age (+) -> Phenotype: Contact_time_12 (+) -> Behaviour: average_energy_availability (-)

Genotype: rs10263021 (-) -> History: Age (+) -> Phenotype: VILR_asymmetry_12 (+) -> Behaviour: average_energy_availability (-)

Genotype: rs970547 (-) -> History: Age (+) -> Phenotype: leg_ffmi (+) -> Behaviour: average_energy_availability (-)

Genotype: rs2289360 (+) -> History: History_extra (-) -> Phenotype: Step_frequency_10 (-) -> Behaviour: average_energy_availability (-)

Genotype: rs10263021 (-) -> History: Age (+) -> Phenotype: Contact_time_12 (+) -> Behaviour: average_energy_availability (-)

Genotype: rs10484958 (-) -> History: History_extra (-) -> Phenotype: Step_frequency_10 (-) -> Behaviour: average_energy_availability (-)

Genotype: rs4454832 (-) -> History: Age (+) -> Phenotype: VILR_asymmetry_12 (+) -> Behaviour: average_energy_availability (-)

Genotype: rs2252070 (+) -> History: History_extra (-) -> Phenotype: Step_frequency_10 (-) -> Behaviour: average_energy_availability (-)

Genotype: rs4454832 (-) -> History: Age (+) -> Phenotype: Contact_time_12 (+) -> Behaviour: average_energy_availability (-)

Genotype: rs2285053 (+) -> History: Age (+) -> Phenotype: VILR_asymmetry_12 (+) -> Behaviour: average_energy_availability (-)

Genotype: rs1800469 (+) -> History: History_extra (-) -> Phenotype: Step_frequency_10 (-) -> Behaviour: average_energy_availability (-)

Genotype: rs2285053 (+) -> History: Age (+) -> Phenotype: Contact_time_12 (+) -> Behaviour: average_energy_availability (-)

Genotype: rs970547 (-) -> History: Age (+) -> Phenotype: VILR_asymmetry_12 (+) -> Behaviour: average_energy_availability (-)

Genotype: rs13946 (-) -> History: Age (+) -> Phenotype: leg_ffmi (+) -> Behaviour: average_energy_availability (-)

Genotype: rs970547 (-) -> History: Age (+) -> Phenotype: Contact_time_12 (+) -> Behaviour: average_energy_availability (-)

Genotype: rs9340799 (+) -> History: History_extra (-) -> Phenotype: Step_frequency_10 (-) -> Behaviour: average_energy_availability (-)

Genotype: rs17583842 (-) -> History: Age (+) -> Phenotype: leg_ffmi (+) -> Behaviour: average_energy_availability (-)

Genotype: rs4362400 (-) -> History: Age (+) -> Phenotype: Contact_time_12 (+) -> Behaviour: average_energy_availability (-)

Genotype: rs3219008 (+) -> History: History_extra (-) -> Phenotype: Step_frequency_10 (-) -> Behaviour: average_energy_availability (-)

Genotype: rs9340799 (-) -> History: Age (+) -> Phenotype: leg_ffmi (+) -> Behaviour: average_energy_availability (-)

Genotype: rs4454832 (-) -> History: History_extra (-) -> Phenotype: Step_frequency_10 (-) -> Behaviour: average_energy_availability (-)

Genotype: rs11154027 (+) -> History: History_extra (-) -> Phenotype: Step_frequency_10 (-) -> Behaviour: average_energy_availability (-)

Genotype: rs1590 (-) -> History: Age (+) -> Phenotype: leg_ffmi (+) -> Behaviour: average_energy_availability (-)

Genotype: rs970547 (+) -> History: History_extra (-) -> Phenotype: Step_frequency_10 (-) -> Behaviour: average_energy_availability (-)

Genotype: rs1718119 (+) -> History: History_extra (-) -> Phenotype: Step_frequency_10 (-) -> Behaviour: average_energy_availability (-)

Genotype: rs1544410 (+) -> History: History_extra (-) -> Phenotype: Step_frequency_10 (-) -> Behaviour: average_energy_availability (-)

Genotype: rs62051384 (+) -> History: History_extra (-) -> Phenotype: Step_frequency_10 (-) -> Behaviour: average_energy_availability (-)

Genotype: rs4362400 (-) -> History: Age (+) -> Phenotype: VILR_asymmetry_12 (+) -> Behaviour: average_energy_availability (-)

Genotype: rs3018362 (+) -> History: History_extra (-) -> Phenotype: Step_frequency_10 (-) -> Behaviour: average_energy_availability (-)

Genotype: rs17583842 (-) -> History: Age (+) -> Phenotype: VILR_asymmetry_12 (+) -> Behaviour: average_energy_availability (-)

Genotype: rs10263021 (+) -> History: History_extra (-) -> Phenotype: Step_frequency_10 (-) -> Behaviour: average_energy_availability (-)

Genotype: rs911263 (+) -> History: History_extra (-) -> Phenotype: Step_frequency_10 (-) -> Behaviour: average_energy_availability (-)

Genotype: rs2525504 (+) -> History: History_extra (-) -> Phenotype: Step_frequency_10 (-) -> Behaviour: average_energy_availability (-)

Genotype: rs4725069 (+) -> History: History_extra (-) -> Phenotype: Step_frequency_10 (-) -> Behaviour: average_energy_availability (-)

Genotype: rs13946 (-) -> History: Age (+) -> Phenotype: VILR_asymmetry_12 (+) -> Behaviour: average_energy_availability (-)

Genotype: rs4701616 (+) -> History: History_extra (-) -> Phenotype: Step_frequency_10 (-) -> Behaviour: average_energy_availability (-)

Genotype: rs13946 (-) -> History: Age (+) -> Phenotype: Contact_time_12 (+) -> Behaviour: average_energy_availability (-)

Genotype: rs1011814 (-) -> History: Age (+) -> Phenotype: leg_ffmi (+) -> Behaviour: average_energy_availability (-)

Genotype: rs4328262 (+) -> History: History_extra (-) -> Phenotype: Step_frequency_10 (-) -> Behaviour: average_energy_availability (-)

Genotype: rs17583842 (-) -> History: Age (+) -> Phenotype: Contact_time_12 (+) -> Behaviour: average_energy_availability (-)

Genotype: rs13946 (+) -> History: History_extra (-) -> Phenotype: Step_frequency_10 (-) -> Behaviour: average_energy_availability (-)

Genotype: rs10263021 (-) -> History: Age (+) -> Phenotype: leg_ffmi (+) -> Behaviour: average_energy_availability (-)

Total consistent routes: 60

**SHAP compared to node contribution**

Enhanced Summary Statistics:

feature_set metric_type mean std min 25% median 75% max

All Features Contribution Correlation 0.358121 0.210931 -0.261885 0.229668 0.387996 0.520535 0.776114

All Features Ratio Correlation 0.196938 0.095426 -0.058380 0.136683 0.195792 0.254061 0.405980

All Features Rank Correlation 0.253139 0.122563 -0.031078 0.171795 0.250422 0.337906 0.509332

Top 5 Node Pos/Neg Contribution Correlation 0.366362 0.464436 -0.895712 0.114464 0.405654 0.774365 0.988631

Top 5 Node Pos/Neg Ratio Correlation 0.222548 0.344175 -0.587074 -0.012870 0.261057 0.524630 0.743778

Top 5 Node Pos/Neg Rank Correlation 0.331061 0.349878 -0.527273 0.124242 0.368182 0.603030 0.939394

Top 5 SHAP Pos/Neg Contribution Correlation 0.485621 0.338812 -0.690572 0.362222 0.601982 0.711466 0.901081

Top 5 SHAP Pos/Neg Ratio Correlation 0.509165 0.300516 -0.361161 0.368907 0.552924 0.766713 0.954433

Top 5 SHAP Pos/Neg Rank Correlation 0.463636 0.363536 -0.660606 0.248485 0.515152 0.757576 0.939394

Combined 3 Node + 3 SHAP Contribution Correlation 0.434325 0.336103 -0.682960 0.270531 0.478296 0.684801 0.924373

Combined 3 Node + 3 SHAP Ratio Correlation 0.417566 0.292045 -0.576483 0.272162 0.427113 0.623573 0.921212

Combined 3 Node + 3 SHAP Rank Correlation 0.442441 0.317606 -0.398601 0.270455 0.496503 0.671329 0.902098

Top Node Ratios Contribution Correlation 0.594956 0.315653 -0.542398 0.428886 0.662728 0.853022 0.987363

Top Node Ratios Ratio Correlation 0.433243 0.243924 -0.430249 0.280080 0.444455 0.624136 0.854309

Top Node Ratios Rank Correlation 0.504485 0.291359 -0.212121 0.296970 0.503030 0.745455 0.963636

Top SHAP Ratios Contribution Correlation 0.525983 0.385246 -0.626045 0.380277 0.624234 0.809395 0.986369

Top SHAP Ratios Ratio Correlation 0.460877 0.261455 -0.262129 0.281398 0.513690 0.667517 0.843582

Top SHAP Ratios Rank Correlation 0.496436 0.321667 -0.660606 0.330303 0.503030 0.772727 0.987879

Combined Ratios Contribution Correlation 0.496665 0.353278 -0.490260 0.314769 0.596718 0.724088 0.964221

Combined Ratios Ratio Correlation 0.332312 0.218443 -0.257447 0.169372 0.335593 0.489663 0.770289

Combined Ratios Rank Correlation 0.471319 0.284637 -0.314685 0.277972 0.486014 0.686713 0.939394

### TSGNN Interpretation

TSGNN interpretation uses a different route contribution calculation method in which biases are taken into account. To achieve this, a ratio was calculated by dividing each incoming message to a node by the sum of all incoming messages to that node. The final output from that node was divided and attributed to each incoming node based on this ratio. This method bypasses batch normalisation, bias addition, and activation function application within each node and directly attributes output based on proportion of input. The route contribution would thus be: proportion (calculated as: feature 1 * weight 1-2 / sum of all inputs to feature 2) * feature 2 output * weight 2-output.

During calculation, it is possible that the sum of all proportions is 0 but feature 2's node is still activated solely via bias term (TSGNN uses leaky relu instead of regular relu, so there would be a small output for every node as long as input + bias is not exactly 0). For these rare cases, individual bias routes were created. The sum of all routes above matches the model's original prediction in logits.

The calculation of feature contribution is divided into two parts. Direct contribution (X-target feature-output) is calculated by taking target feature node's output * weight. Indirect contribution (target feature-X-output) is calculated by summing all routes that originate from the target feature. Summing contributions from the two parts results in the final feature contribution value. When averaging across different runs, each feature contribution is divided by the sum of all absolute feature contribution values within that run as a normalisation step.

### TSGNN Interpretation Output

#### TSGNN Class 1

#### Consistent weights across 20 runs

Consistent Weights: 9/633 connections (p<0.0001)

================================================

| Source | Target | Sign |
|---|---|---|
| rs591058 | rs970547 | Negative |
| rs591058 | Q_angle_asymmetry | Negative |
| rs591058 | SC_past_season | Positive |
| rs4789932 | rs970547 | Positive |
| rs13946 | Q_angle | Negative |
| rs9340799 | non_running_past_season | Positive |
| rs1800012 | rs970547 | Negative |

| | Age | past_month_distance | Positive |
|---|---|---|---|
| hip_abduction_peak_torque | knee_extension_peak_torque | Positive | |

**Correlation metrics (normalised by node contributions)**

| | mean | std | median | 25% | 75% |
|---|---|---|---|---|---|
| all_features_contrib | 0.259341 | 0.211894 | 0.266009 | 0.130653 | 0.412649 |
| all_features_ratio | -0.022317 | 0.174446 | -0.037698 | -0.143505 | 0.116092 |
| all_features_rank | 0.185220 | 0.181770 | 0.217192 | 0.097691 | 0.321206 |
| top_node_5pos5neg_contrib | 0.449827 | 0.296040 | 0.509368 | 0.280848 | 0.655987 |
| top_node_5pos5neg_ratio | 0.379570 | 0.318533 | 0.412253 | 0.199588 | 0.626923 |
| top_node_5pos5neg_rank | 0.443152 | 0.279746 | 0.503030 | 0.269697 | 0.639394 |
| top_shap_5pos5neg_contrib | 0.334816 | 0.300396 | 0.397062 | 0.141382 | 0.547240 |
| top_shap_5pos5neg_ratio | 0.029910 | 0.318175 | 0.016327 | -0.214292 | 0.236457 |
| top_shap_5pos5neg_rank | 0.263879 | 0.316560 | 0.254545 | 0.036364 | 0.503030 |
| combined_3n3s_contrib | 0.320142 | 0.266580 | 0.320665 | 0.115200 | 0.522805 |
| combined_3n3s_ratio | 0.211423 | 0.332421 | 0.217701 | -0.016301 | 0.446065 |
| combined_3n3s_rank | 0.294826 | 0.240884 | 0.319930 | 0.094156 | 0.484091 |
| top_node_ratio5_contrib | 0.240083 | 0.352988 | 0.226041 | -0.012198 | 0.547198 |
| top_node_ratio5_ratio | -0.056517 | 0.330357 | -0.077003 | -0.288769 | 0.187833 |
| top_node_ratio5_rank | 0.190909 | 0.329022 | 0.151515 | -0.081818 | 0.421212 |
| top_shap_ratio5_contrib | 0.340527 | 0.333700 | 0.376544 | 0.117570 | 0.623312 |
| top_shap_ratio5_ratio | -0.042673 | 0.320122 | -0.059551 | -0.269025 | 0.160393 |
| top_shap_ratio5_rank | 0.293212 | 0.315037 | 0.315152 | 0.100000 | 0.539394 |
| combined_ratio3_contrib | 0.253299 | 0.328529 | 0.262850 | 0.041422 | 0.527752 |
| combined_ratio3_ratio | -0.078705 | 0.285332 | -0.071101 | -0.288901 | 0.139477 |

combined_ratio3_rank 0.231174 0.318738 0.254371 0.031818 0.442424

**TSGNN Class 123**

**Consistent weights across 20 runs**

Consistent Weights: 164/27225 connections (p<0.0001)

==================================================

| Source | Target | Sign |
|---|---|---|
| rs591058 | rs1800972 | Negative |
| rs2104772 | rs12722 | Negative |
| rs1144393 | rs4654760 | Positive |
| rs1144393 | rs1800795 | Positive |
| rs1144393 | Flight_time_12 | Positive |
| rs1144393 | hip_adduction_peak_angle | Positive |
| rs1144393 | resistance_training_past_season | Positive |
| rs1544410 | rs911263 | Negative |
| rs1544410 | leg_lean_mass | Negative |
| rs2237352 | rs10992075 | Negative |
| rs2237352 | rs12656106 | Negative |
| rs1137101 | calcium_intake_BW | Negative |
| rs10263021 | past_month_volume_low | Positive |
| rs10132091 | rs10759753 | Positive |
| rs10132091 | resistance_training_past_season | Negative |
| rs7035322 | rs2761884 | Negative |
| rs1330363 | resistance_training_past_season | Positive |
| rs1011814 | rs11154027 | Positive |

| | | |
|---|---|---|
| rs1249269 | drills_past_season | Positive |
| rs820218 | rs11154027 | Positive |
| rs1800629 | rs1554606 | Positive |
| rs11177 | rs2237352 | Positive |
| rs6481512 | rs1718119 | Positive |
| rs6481512 | rs420257 | Positive |
| rs6481512 | rs117544024 | Positive |
| rs6481512 | hip_adduction_peak_angle | Negative |
| rs4730153 | average_energy_availability | Negative |
| rs970547 | rs1887632 | Positive |
| rs3018362 | BMD_body | Negative |
| rs3018362 | average_energy_availability | Negative |
| rs11629171 | protein_intake_BW | Positive |
| rs10992075 | rs1554606 | Positive |
| rs4789932 | past_month_volume_low | Negative |
| rs1590 | past_month_volume_low | Positive |
| rs1590 | past_month_distance | Positive |
| rs4362400 | tracking_period_injury | Negative |
| rs4362400 | protein_intake_BW | Positive |
| rs3219008 | hip_adduction_peak_torque | Positive |
| rs3218791 | rs4919510 | Negative |
| rs2289360 | rs3219008 | Positive |
| rs2289360 | rs1643821 | Positive |
| rs2289360 | rs1548456 | Positive |
| rs2289360 | knee_extension_peak_angle_asymmetry | Positive |

| | |
|---|---|
| rs1134170 | rs1554606 Positive |
| rs4919510 | rs1011814 Negative |
| rs10484958 | rs4362400 Negative |
| rs10484958 | Flight_time_12 Negative |
| rs10484958 | fat_percentage_avg Negative |
| rs2285053 | rs2586488 Positive |
| rs1800972 | drills_past_season Positive |
| rs4328262 | total_ffmi Positive |
| rs2306033 | knee_extension_peak_torque Positive |
| rs9340799 | copper_intake_BW Positive |
| rs17583842 | rs4654760 Positive |
| rs1643821 | rs3219008 Positive |
| rs1643821 | rs2281518 Positive |
| rs3045 | Duty_factor_asymmetry_12 Negative |
| rs4903399 | rs4730153 Negative |
| rs187483 | rs1590 Positive |
| sex | rs13317 Negative |
| sex | rs3219008 Negative |
| sex | rs4919510 Negative |
| sex | tracking_period_injury Negative |
| rs1554606 | rs3219008 Positive |
| rs2305948 | rs1138545 Positive |
| rs2305948 | calcium_intake_BW Positive |
| rs1887632 | drills_past_season Positive |
| rs1800795 | rs1548456 Positive |

| | | |
|---|---|---|
| rs12154667 | rs1718119 | Positive |
| rs1676303 | knee_flexion_peak_torque_asymmetry | Negative |
| rs1676303 | non_running_past_month | Positive |
| rs12574452 | rs1800795 | Positive |
| rs2277268 | tracking_period_injury | Positive |
| rs2277268 | arginine_intake_BW | Negative |
| rs912336 | rs10484958 | Positive |
| rs74544784 | rs117544024 | Negative |
| rs60713544 | rs13317 | Positive |
| rs60713544 | rs1643821 | Positive |
| rs12429486 | rs187483 | Positive |
| rs12429486 | rs77569527 | Negative |
| rs12429486 | non_running_past_month | Positive |
| rs2010963 | rs25489 | Positive |
| rs2010963 | drills_past_season | Positive |
| rs71404070 | rs2228570 | Positive |
| rs71404070 | rs1718119 | Positive |
| rs71404070 | drills_past_season | Positive |
| rs117544024 | rs74544784 | Negative |
| rs117544024 | non_running_past_month | Positive |
| rs2586488 | drills_past_season | Positive |
| rs25489 | rs1718119 | Positive |
| rs25489 | rs1548456 | Positive |
| rs25489 | knee_flexion_peak_torque_asymmetry | Positive |
| rs413826 | rs11154027 | Positive |

| | | |
|---|---|---|
| rs650108 | rs42531 | Positive |
| rs650108 | past_month_distance | Positive |
| rs650108 | drills_past_season | Positive |
| rs57104447 | rs1590 | Positive |
| rs57104447 | rs1718119 | Positive |
| rs57104447 | rs187483 | Positive |
| rs145648292 | rs11154027 | Negative |
| rs145648292 | rs1718119 | Positive |
| rs145648292 | knee_extension_peak_angle_asymmetry | Negative |
| rs145648292 | knee_flexion_peak_torque_asymmetry | Negative |
| rs77569527 | rs187483 | Positive |
| rs144371252 | knee_extension_peak_angle_asymmetry | Positive |
| rs4988321 | rs1249269 | Positive |
| rs4988321 | rs1718119 | Positive |
| rs4988321 | rs2285053 | Positive |
| rs4988321 | rs1800972 | Positive |
| rs4988321 | rs3045 | Positive |
| rs149047058 | drills_past_season | Positive |
| rs144414988 | rs1330363 | Negative |
| rs144414988 | rs11154027 | Negative |
| rs144414988 | rs17583842 | Negative |
| rs144414988 | Flight_time_12 | Negative |
| rs144414988 | knee_flexion_peak_torque_asymmetry | Negative |
| rs144414988 | hip_abduction_peak_torque | Negative |
| rs144414988 | Impact_peak_12 | Negative |

| | |
|---|---|
| rs144414988 | BMD_body Negative |
| rs144414988 | glycine_intake_BW Positive |
| rs144414988 | vitaminD_intake_BW Positive |
| rs144414988 | past_month_min Negative |
| rs2858056 | rs1249269 Positive |
| rs2858056 | rs1643821 Positive |
| rs2858056 | drills_past_season Positive |
| tracking_period_injury | VILR_10 Positive |
| tracking_period_injury | knee_flexion_peak_torque_asymmetry Positive |
| tracking_period_injury | Duty_factor_asymmetry_12 Positive |
| tracking_period_injury | past_month_distance Positive |
| tracking_period_injury | drills_past_season Positive |
| tracking_period_injury | past_month_min Positive |
| Athlete_Score | thigh_ffmi Positive |
| Athlete_Score | BMD_body Positive |
| Athlete_Score | past_month_volume_low Positive |
| Athlete_Score | past_month_min Positive |
| Step_frequency_10 | knee_extension_peak_angle_asymmetry Positive |
| knee_extension_peak_angle_asymmetry | Q_angle Positive |
| knee_extension_peak_angle_asymmetry | total_ad_ab_ratio Positive |
| knee_extension_peak_angle_asymmetry | hip_adduction_peak_torque_asymmetry Positive |
| knee_extension_peak_angle_asymmetry | Flight_time_12 Positive |
| knee_extension_peak_angle_asymmetry | knee_extension_peak_torque Positive |
| knee_extension_peak_angle_asymmetry | knee_flexion_peak_torque_asymmetry Positive |

knee_extension_peak_angle_asymmetry hip_abduction_peak_torque Positive

knee_extension_peak_angle_asymmetry knee_flexion_peak_angle Positive

knee_extension_peak_angle_asymmetry hip_adduction_peak_angle Positive

knee_extension_peak_angle_asymmetry Duty_factor_10 Positive

knee_extension_peak_angle_asymmetry Duty_factor_asymmetry_12 Positive

Q_angle_asymmetry knee_flexion_peak_torque_asymmetry Negative

Q_angle_asymmetry knee_flexion_peak_angle Negative

Q_angle_asymmetry knee_extension_peak_torque_asymmetry Negative

knee_flexion_peak_angle_asymmetry copper_intake_BW Negative

knee_flexion_peak_angle Duty_factor_asymmetry_12 Positive

knee_extension_peak_torque_asymmetry resistance_training_past_season Negative

Duty_factor_asymmetry_10 knee_flexion_peak_angle Negative

Duty_factor_asymmetry_10 resistance_training_past_season Positive

fat_intake_BW drills_past_season Negative

fat_intake_avg drills_past_season Negative

glycine_intake_BW protein_intake_BW Negative

glycine_intake_BW past_month_volume_low Negative

arginine_intake_BW protein_intake_BW Negative

SC_past_season calcium_intake_BW Positive

vitaminD_intake_BW past_month_volume_low Negative

bodyweight_exercises_past_season past_month_distance Negative

bodyweight_exercises_past_season past_month_min Negative

**Correlation metrics (normalised by node contributions)**

| | mean | std | median | 25% | 75% |
|---|---|---|---|---|---|
| all_features_contrib | 0.432595 | 0.171193 | 0.446508 | 0.326830 | 0.532401 |
| all_features_ratio | 0.216271 | 0.093869 | 0.223943 | 0.176756 | 0.283707 |
| all_features_rank | 0.239616 | 0.110369 | 0.239495 | 0.169102 | 0.327044 |
| top_node_5pos5neg_contrib | 0.720708 | 0.256692 | 0.796896 | 0.666446 | 0.883926 |
| top_node_5pos5neg_ratio | 0.726897 | 0.258784 | 0.790268 | 0.580535 | 0.924397 |
| top_node_5pos5neg_rank | 0.712970 | 0.266977 | 0.781818 | 0.648485 | 0.878788 |
| top_shap_5pos5neg_contrib | 0.682784 | 0.264769 | 0.762191 | 0.588971 | 0.866429 |
| top_shap_5pos5neg_ratio | 0.605745 | 0.286005 | 0.695992 | 0.395083 | 0.795978 |
| top_shap_5pos5neg_rank | 0.628242 | 0.261616 | 0.721212 | 0.466667 | 0.821212 |
| combined_3n3s_contrib | 0.581678 | 0.234065 | 0.636798 | 0.466039 | 0.739642 |
| combined_3n3s_ratio | 0.687773 | 0.249279 | 0.746682 | 0.597434 | 0.852730 |
| combined_3n3s_rank | 0.525199 | 0.196714 | 0.548485 | 0.440559 | 0.651136 |
| top_node_ratio5_contrib | 0.540781 | 0.354823 | 0.656075 | 0.431964 | 0.777692 |
| top_node_ratio5_ratio | 0.346344 | 0.334464 | 0.382467 | 0.165050 | 0.574089 |
| top_node_ratio5_rank | 0.397576 | 0.316994 | 0.472727 | 0.245455 | 0.603030 |
| top_shap_ratio5_contrib | 0.407020 | 0.352406 | 0.506354 | 0.166462 | 0.668978 |
| top_shap_ratio5_ratio | 0.312529 | 0.310136 | 0.294759 | 0.084348 | 0.526956 |
| top_shap_ratio5_rank | 0.273576 | 0.356790 | 0.351515 | 0.078788 | 0.551515 |
| combined_ratio3_contrib | 0.509912 | 0.298496 | 0.542094 | 0.369058 | 0.715631 |
| combined_ratio3_ratio | 0.332913 | 0.278466 | 0.347539 | 0.178647 | 0.549010 |
| combined_ratio3_rank | 0.367387 | 0.309427 | 0.356643 | 0.152098 | 0.610140 |

### Logistic Regression Coefficients

Class 1:

=== Feature Coefficients (Descending Order) ===

| Feature | Coefficient |
|---|---|
| tracking_period_injury | 2.618926 |
| class1_SNP_risk_score | 1.360031 |
| past_month_distance | 1.204346 |
| Q_angle_asymmetry | 0.839222 |
| navicular_drop | 0.707992 |
| knee_flexion_peak_torque | 0.658180 |
| past_month_ratio | 0.480944 |
| rs591058 | 0.334506 |
| rs4986938 | -0.176714 |
| rs1800795 | -0.274394 |
| rs4789932 | -0.277614 |
| BMD_spine | -0.347275 |
| hip_abduction_peak_torque | -0.456419 |
| rs9340799 | -0.477965 |
| rs1144393 | -0.549924 |
| lower_limb_days_total | -0.581701 |
| SC_past_season | -0.620537 |
| Q_angle | -0.729596 |

Impact_peak_12 -0.757071

average_run_hours -0.959838

Duty_factor_12 -1.280184

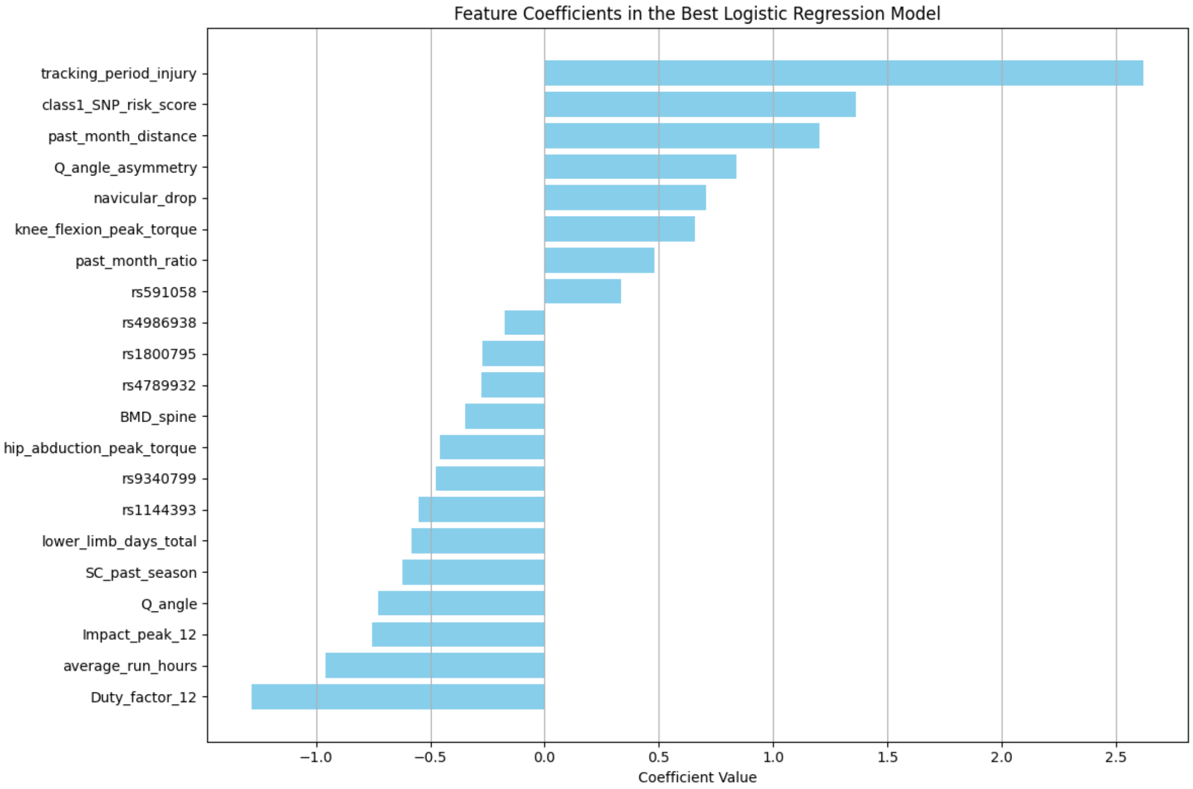

All features:

=== Feature Coefficients (Descending Order) ===

| Feature | Coefficient |
|---|---|
| rs145648292 | 3.373070 |
| rs1676303 | 3.299420 |
| class1_SNP_risk_score | 2.525343 |
| rs3045 | 2.059349 |
| past_month_ratio | 1.818201 |
| past_week_ratio_low | 1.694178 |

| | |
|---|---|
| Q_angle_asymmetry | 1.558657 |
| vitaminD_intake_BW | 1.539176 |
| rs912336 | 1.418766 |
| rs2104772 | 1.408350 |
| rs2277268 | 1.361082 |
| iron_intake_BW | 1.304049 |
| rs7035322 | 1.266843 |
| Duty_factor_asymmetry_10 | 1.266532 |
| resistance_training_past_month | 1.265438 |
| arginine_intake_BW | 1.225935 |
| rs13107325 | 1.120972 |
| rs4919510 | 1.107007 |
| past_month_distance | 1.057822 |
| BMD_hip | 1.045272 |
| rs1800629 | 1.008072 |
| copper_intake_BW | 0.997871 |
| rs3218791 | 0.979804 |
| rs2285053 | 0.947730 |
| SC_past_month | 0.856245 |
| rs10484958 | 0.852573 |
| knee_flexion_peak_angle_asymmetry | 0.849551 |
| leg_ffmi | 0.794903 |
| rs11225395 | 0.763624 |
| hip_adduction_peak_torque | 0.726183 |
| rs2252070 | 0.654123 |
| fat_percentage_avg | 0.502869 |

| | |
|---|---|
| rs4328262 | 0.498168 |
| rs2281518 | 0.480000 |
| past_month_injury | 0.475731 |
| past_week_ratio_calculated_volume | 0.437190 |
| navicular_drop_asymmetry | 0.412264 |
| rs1800972 | 0.405746 |
| rs591058 | 0.329773 |
| rs143383 | 0.313561 |
| rs2586488 | 0.307136 |
| rs4986938 | 0.271181 |
| rs2858056 | 0.262386 |
| rs9340799 | 0.196693 |
| Age | 0.185960 |
| rs6481512 | 0.119788 |
| rs1887632 | 0.064215 |
| rs1045485 | 0.036617 |
| rs17583842 | 0.000000 |
| EDEQ_total | -0.028527 |
| drills_past_season | -0.054817 |
| rs62051384 | -0.145910 |
| rs12574452 | -0.147123 |
| rs2010963 | -0.273623 |
| past_month_volume_high | -0.306241 |
| protein_intake_BW | -0.337953 |
| rs10992075 | -0.353891 |
| past_month_ratio_moderate | -0.357693 |

rs1800795 -0.387039

circuit_training_past_month -0.400959

rs1330363 -0.418570

rs1643821 -0.421583

rs4903399 -0.472789

rs1011814 -0.551650

rs25487 -0.555279

rs3219008 -0.555582

past_week_ratio_high -0.643633

calcium_intake_BW -0.650396

rs1249269 -0.660208

rs1800469 -0.660498

rs10132091 -0.704059

rs11232681 -0.706245

rs2305948 -0.743006

past_month_ratio_calculated_volume -0.795976

Athlete_Score -0.811113

Contact_time_12 -0.852111

rs2289360 -0.908303

fl_ex_ratio_asymmetry -1.001542

rs25489 -1.022105

resistance_training_past_season -1.031873

rs2234693 -1.144350

rs1590 -1.193586

average_energy_availability -1.223881

rs2306033 -1.324446

Alt_strike -1.384407

tracking_period_injury -1.430391

rs4701616 -1.650593

SC_past_season -1.670293

rs1144393 -1.751950

rs4988321 -1.836886

hip_abduction_peak_angle -1.882983

knee_extension_peak_angle_asymmetry -1.960411

ad_ab_ratio_asymmetry -2.040596

BMD_body -2.156149

average_run_hours -3.157189

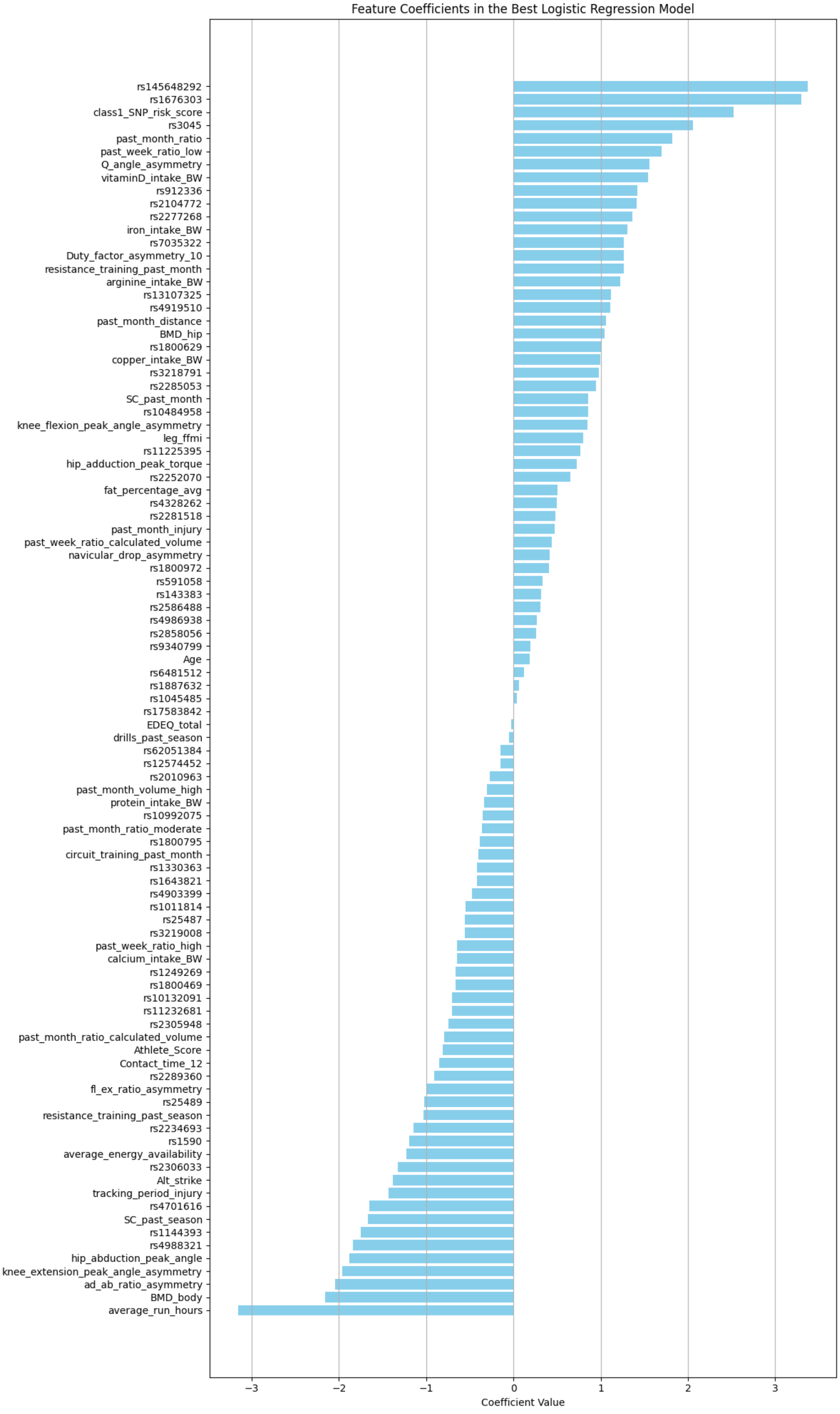
Feature Coefficients in the Best Logistic Regression Model
rs145648292
rs1676303
class1_SNP_risk_score
rs3045
past_month_ratio
past_week_ratio_low
Q_angle_asymmetry
vitaminD_intake_BW
rs912336
rs2104772
rs2277268
iron_intake_BW
rs7035322
Duty_factor_asymmetry_10
resistance_training_past_month
arginine_intake_BW
rs13107325
rs4919510
past_month_distance
BMD_hip
rs1800629
copper_intake_BW
rs3218791
rs2285053
SC_past_month
rs10484958
knee_flexion_peak_angle_asymmetry
leg_ffmi
rs11225395
hip_adduction_peak_torque
rs2252070
fat_percentage_avg
rs4328262
rs2281518
past_month_injury
past_week_ratio_calculated_volume
navicular_drop_asymmetry
rs1800972
rs591058
rs143383
rs2586488
rs4986938
rs2858056
rs9340799
Age
rs6481512
rs1887632
rs1045485
rs17583842
EDEQ_total
drills_past_season
rs62051384
rs12574452
rs2010963
past_month_volume_high
protein_intake_BW
rs10992075
past_month_ratio_moderate
rs1800795
circuit_training_past_month
rs1330363
rs1643821
rs4903399
rs1011814
rs25487
rs3219008
past_week_ratio_high
calcium_intake_BW
rs1249269
rs1800469
rs10132091
rs11232681
rs2305948
past_month_ratio_calculated_volume
Athlete_Score
Contact_time_12
rs2289360
fl_ex_ratio_asymmetry
rs25489
resistance_training_past_season
rs2234693
rs1590
average_energy_availability
rs2306033
Alt_strike
tracking_period_injury
rs4701616
SC_past_season
rs1144393
rs4988321
hip_abduction_peak_angle
knee_extension_peak_angle_asymmetry
ad_ab_ratio_asymmetry
BMD_body
average_run_hours
−3
−2
−1
0
1
2
3
Coefficient Value

### Model Interpretation Results

S11.1: Number of consistent weight values for TSNN. *p < 0.05; **p < 0.001

| Class 1 Features | | | | |
|---|---|---|---|---|
| Number of consistent runs (total n=20) | 17 | 16 | 15 | 14 |
| Consistent weights (total n=152) | 1 | 3 | 10 | 24 |
| p | 0.324 | 0.268 | 0.101 | 0.069 |
| **All Features** | | | | |
| Number of consistent runs (total n=20) | 18 | 17 | 16 | 15 |
| Consistent weights (total n=744) | 1 | 7 | 20 | 56 |
| p | 0.259 | 0.004* | 0.001* | <0.001** |

S11.2: Correlation analysis for calculated feature contributions versus SHAP values (averaged across 20 runs for 100 samples).

| | All Features (SD) | | Top 5 Positive and Negative (SD) | |
|---|---|---|---|---|
| | Pearson's r | Spearman's ρ | Pearson's r | Spearman's ρ |
| TSNN Class 1 | 0.17 (0.27) | 0.12 (0.19) | 0.21 (0.40) | 0.22 (0.36) |
| TSNN All | 0.36 (0.21) | 0.25 (0.12) | 0.37 (0.46) | 0.33 (0.35) |
| TSGNN Class 1 | 0.26 (0.21) | 0.19 (0.18) | 0.45 (0.30) | 0.44 (0.28) |
| TSGNN All | 0.43 (0.17) | 0.24 (0.11) | 0.72 (0.26) | 0.71 (0.27) |
